# Prognostication for prelabor rupture of membranes and the time of delivery in nationwide insured women: development, validation, and deployment

**DOI:** 10.1101/2021.06.16.21258884

**Authors:** Herdiantri Sufriyana, Yu-Wei Wu, Emily Chia-Yu Su

**Affiliations:** Graduate Institute of Biomedical Informatics, College of Medical Science and Technology, Taipei Medical University, 250 Wu-Xing Street, Taipei 11031, Taiwan; Department of Medical Physiology, Faculty of Medicine, Universitas Nahdlatul Ulama Surabaya, 57 Raya Jemursari Road, Surabaya 60237, Indonesia; Clinical Big Data Research Center, Taipei Medical University Hospital, 250 Wu-Xing Street, Taipei 11031, Taiwan; Research Center for Artificial Intelligence in Medicine, Taipei Medical University, 250 Wu-Xing Street, Taipei 11031, Taiwan

**Keywords:** prelabor rupture of membranes, preterm delivery, risk prediction, machine learning

## Abstract

**Importance:** Prognostic predictions of prelabor rupture of membranes lack proper sample sizes and external validation.

**Objective:** To develop, validate, and deploy statistical and/or machine learning prediction models using medical histories for prelabor rupture of membranes and the time of delivery.

**Design:** A retrospective cohort design within 2-year period (2015 to 2016) of a single-payer, government-owned health insurance database covering 75.8% individuals in a country

**Setting:** Nationwide healthcare providers (*n*=22,024) at primary, secondary, and tertiary levels

**Participants:** 12-to-55-year-old women that visit healthcare providers using the insurance from ∼1% random sample of insurance holders stratified by healthcare provider and category of family: (1) never visit; (2) visit only primary care; and (3) visit all levels of care

**Predictors:** Medical histories of diagnosis and procedure (International Classification of Disease version 10) before the latest visit of outcome within the database period

**Main Outcomes and Measures:** Prelabor rupture of membranes prognostication (area under curve, with sensitivity, specificity, and likelihood ratio), the time of delivery estimation (root mean square error), and inference time (minutes), with 95% confidence interval

**Results:** We selected 219,272 women aged 33 ± 12 years. The best prognostication achieved area under curve 0.73 (0.72 to 0.75), sensitivity 0.494 (0.489 to 0.500), specificity 0.816 (0.814 to 0.818), and likelihood ratio being positive 2.68 (2.63 to 2.75) and negative 0.62 (0.61 to 0.63). This outperformed models from previous studies according to area under curve of an external validation set, including one using a biomarker (area under curve 0.641; sensitivity 0.419; sensitivity 0.863; positive likelihood ratio 3.06; negative likelihood ratio 0.67; *n*=1177). Meanwhile, the best estimation achieved ± 2.2 and 2.6 weeks respectively for predicted events and non-events. Our web application only took 5.14 minutes (5.11 to 5.18) per prediction.

**Conclusions and Relevance:** Prelabor rupture of membranes and the time of delivery were predicted by medical histories; but, an impact study is required before clinical application.

**Key Points:** *Question:* Can we use medical histories of diagnosis and procedure in electronic health records to predict prelabor rupture of membranes and the time of delivery before the day in nationwide insured women?

*Findings:* In this prognostic study applying retrospective cohort paradigm, a significant predictive performance was achieved and validated. The area under receiver operating characteristics curve was 0.73 with the estimation errors of ± 2.2 and 2.6 weeks for the time of delivery.

*Meaning:* Preliminary prediction can be conducted in a wide population of insured women to predict prelabor rupture of membranes and estimate the time of delivery.

## Introduction

Preterm prelabor rupture of membranes (PROM) is widely used as an inclusion criterion for predictions of other conditions.^1-8^ The disease precedes 40%∼50% of all preterm deliveries and arises from multiple disease pathways.^9^ Yet, the antecedent remains unclear, and prognostic predictions lack proper sample sizes and external validation.

Preterm delivery occurs in ∼10% births in the United States, among which 2%∼3% are contributed by preterm PROM, while the term PROM occurs in ∼8% of pregnancies.^10^ Premature babies require a neonatal intensive care unit (NICU),^11,12^ which is scarce in several countries worldwide.^13-17^ Meanwhile, use of NICUs accounts for a majority of healthcare costs in pediatrics and the single largest item in healthcare spending.^18^ Predicting this disease, estimating the time of delivery, and tracing possible root causes would enable development of preventive strategies and improve efficiency of conducting a prospective cohort study of associated complications.

Prediction of PROM is mostly diagnostic.^19-22^ For prognostications, a model of preterm PROM was recently developed using maternal factors during the first trimester.^23^ Based on a training set (*n*=10,280), the area under receiver operating characteristic (ROC) curve (AUROC) was 0.667.Predictions of all-cause spontaneous preterm deliveries are also poor (AUROCs 0.54 to 0.70; <37 weeks’ gestation; *n*=118/2540), and they are exposed to high risks of bias based on a systematic review.^24^ However, there has been no development or external validation of a prognostic prediction model for PROM. In addition, because it is reasonably challenging, no studies have developed a model to estimate the time of delivery before the day.

For both classification and estimation tasks, machine learning algorithms have demonstrated promising performances for pregnancy outcomes^25^ and other conditions.^26-28^ Despite some hype, most of the greatest successes are diagnostic, especially deep-learning models that surpass human-level performance.^29-31^ Machine learning, however, is yet unable to infer causality; thus, human learning is needed to estimate what will likely happen if conditions differ from those existing in the dataset from which a machine learns.^32^ Nonetheless, solving PROM problems requires predictions and causal modeling to develop better preventive strategies at both the population and individual levels. To address this issue, we applied both human and machine learning tools.^33^ Using only medical histories, the model deployment could be accessible worldwide via a web application. This study aimed to develop, validate, and deploy a prognostic prediction model for PROM and an estimator of the time of delivery using a nationwide health insurance database.

## Methods

We reported this study according to transparent reporting of a multivariable prediction model for individual prognosis or diagnosis (TRIPOD).^34^ This study is a part of a DI-VNN project that applied our algorithm to various predicted outcomes. To develop the prediction models, we followed a protocol using the same hardware and software.^33^ The checklists for all of the guidelines, including those in the protocol, and comparable models are available in eTables 1 to 5 in the Supplement. Ethical clearance was waived by the Taipei Medical University Joint Institutional Review Board (TMU-JIRB number: N202106025).

**Table 1.**
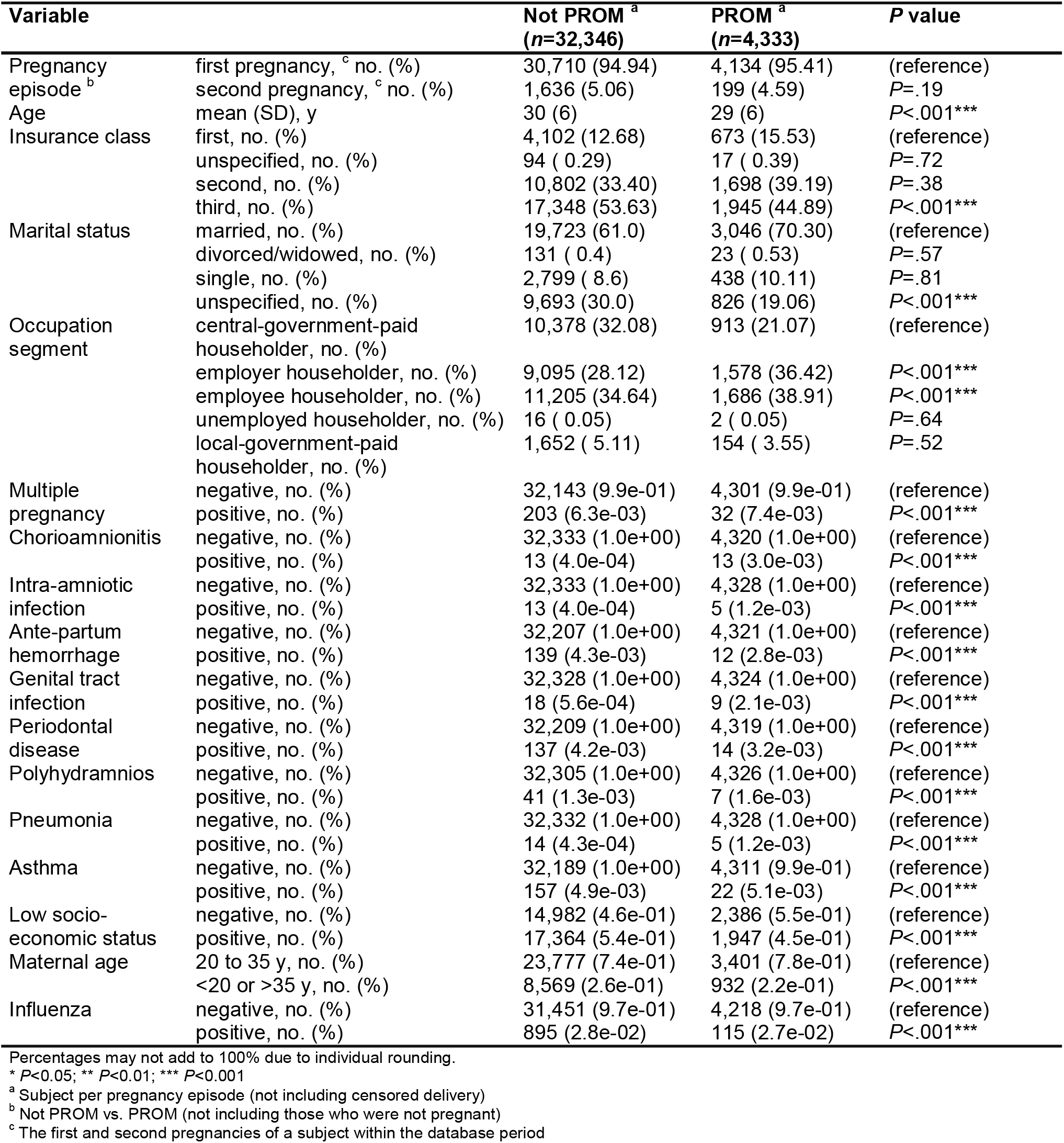
Baseline characteristics of subjects for association tests and internal validation set.

### Study design

We applied a retrospective design to select subjects from a nationwide health insurance dataset provided by a government-owned health insurance company in Indonesia. The health insurance covered 200,259,147 (75.8%) individuals in that country,^35^ including races of Asian and Austronesian. This dataset was the second version published on August 2019 (access approval no.: 5064/I.2/0421) covering ∼1% (*n*=1,697,452) of insurance holders in 2015 and 2016 from nationwide, affiliated healthcare providers (*n*=22,024; primary, secondary, and tertiary care). Sampling for the data source was stratified by healthcare provider and category of family: (1) never visit; (2) visit only primary care; and (3) visit all levels of care. Details of these sampling procedures are described in the Supplement.

We included health insurance holders of 12∼55-year-old females who had visited affiliated healthcare providers. We excluded visits after delivery. For a person who was pregnant twice within the period, we labeled the same person as a different subject for each pregnancy period. A complete list of codes for determining delivery or immediately after delivery care is available in eTable 7 of the Supplement.

We developed prediction models to classify if a visit was made by a subject for which the pregnancy period ended with PROM, and to estimate the time of delivery. Under-prognosis of PROM may cause pregnancy monitoring to be off-guard and was considered more serious than over-prognosis. A well-calibrated model with higher sensitivity should be given priority. Meanwhile, the error for estimating the time of delivery is acceptable around 2 to 4 weeks, since this is a common interval between antenatal visits closer to the time of delivery.

The outcome for the classification task was an event for a subject that had encountered the O42 code, which is the International Classification of Disease version 10 (ICD-10) code for PROM. Otherwise, a subject was assigned as a nonevent if the pregnancy ended within the dataset period using the same codes for pregnancy termination. If neither having pregnancy nor delivery, we assigned censoring labels. Based on the protocol,^33^ we used these labels to preserve outcome distribution in the target population and resolve class imbalance by inversely weighting the uncensored outcomes considering both the censored and uncensored ones. Meanwhile, the outcome for the estimation task was the number of days from the latest visit encountering of the outcome code to a visit when the prediction model was used.

Candidate predictors consisted of medical histories defined by one or more codes of diagnosis and procedure. The multiple codes composed latent candidate predictors determined by model-based statistical tests to find potential candidate predictors based on systematic human learning.^36^ Steps to determine candidate predictors were described in the protocol,^33^ to avoid zero variance, perfect separation problem, outcome leakage, and redundancy, and to mimic real-world settings.^37,38^ These resulted in 372 candidate predictors. Details of the candidate predictors and selection are described in eTable 8 of the Supplement.

### Model development, validation, and deployment

Five models were developed. These are described in the protocol,^33^ including hyperparameter tuning. Briefly, the first model was a statistical, i.e., ridge regression (RR), on 9 of 12 latent candidate predictors selected by the systematic human learning.^36^ One of them was not associated with PROM (see Results), and we did not select low SES and maternal age for reducing use of private data and preventing social and economic discrimination. Opposite to systematic human learning, we applied unsupervised machine learning to transform candidate predictors into principal components (PCs). These were used for supervised machine learning algorithms of an elastic net regression (PC-ENR), random forest (PC-RF), and gradient boosting machine (PC-GBM). The latter two algorithms outperformed other algorithms for pregnancy outcomes.^25^ Based on the PC-ENR model, the PCs were reduced into 60 PCs (see the protocol^33^) to pursue 200 events per variable (EPV) for the PC-RF and PC-GBM, as recommended by the prediction model risk of bias assessment tools (PROBAST).^38^ The fifth model was the DI-VNN. It was developed to achieve moderate predictive performance but interpretable results based on recent studies.^39,40^ The DI-VNN allows deep exploration of how this algorithm works.^41^ For this model, there were 144 candidate predictors after differential analyses with multiple testing corrections. For classification, we calibrated each model by a general additive model using locally weighted scatterplot smoothing (GAM-LOESS).

We split the dataset for internal and external validation. As recommended, we split out a dataset for external validation by geographical, temporal, and geotemporal splitting, approximately covering ∼20% of visits. This reflected the situation in some real-world settings but not that in nationwide, which represented by ∼20% random split of the remaining set; thus leaving ∼64% of the original sample size for internal validation. We split out ∼20% of the internal validation set to calibrate each model. The final predictive performance of internal validation came from this calibration subset by bootstrapping 30 times. Details on validation were described in the protocol.^33^

We deployed the best models as a web application. It needed a deidentified, two-column comma-separated value (CSV) file of admission dates and ICD-10 codes of the medical history of a subject. A user can set a threshold for the expected population-level sensitivity, specificity, positive predictive value, and negative predictive value. We showed an example in the Supplement. We also measured inference time for a prediction with 10 iterations.

### Statistical analysis

All evaluation metrics are expressed as estimates with the 95% confidence interval (CI). Based 12 association diagrams (eFigures 1∼12 in the Supplement) of PROM, we conducted inverse probability weighting (IPW) to verify associations of the latent candidate factors and PROM (eTables 11∼12 in the Supplement). Outcome regression was also conducted for comparison (eTables 13 in the Supplement).

**Figure 1.**
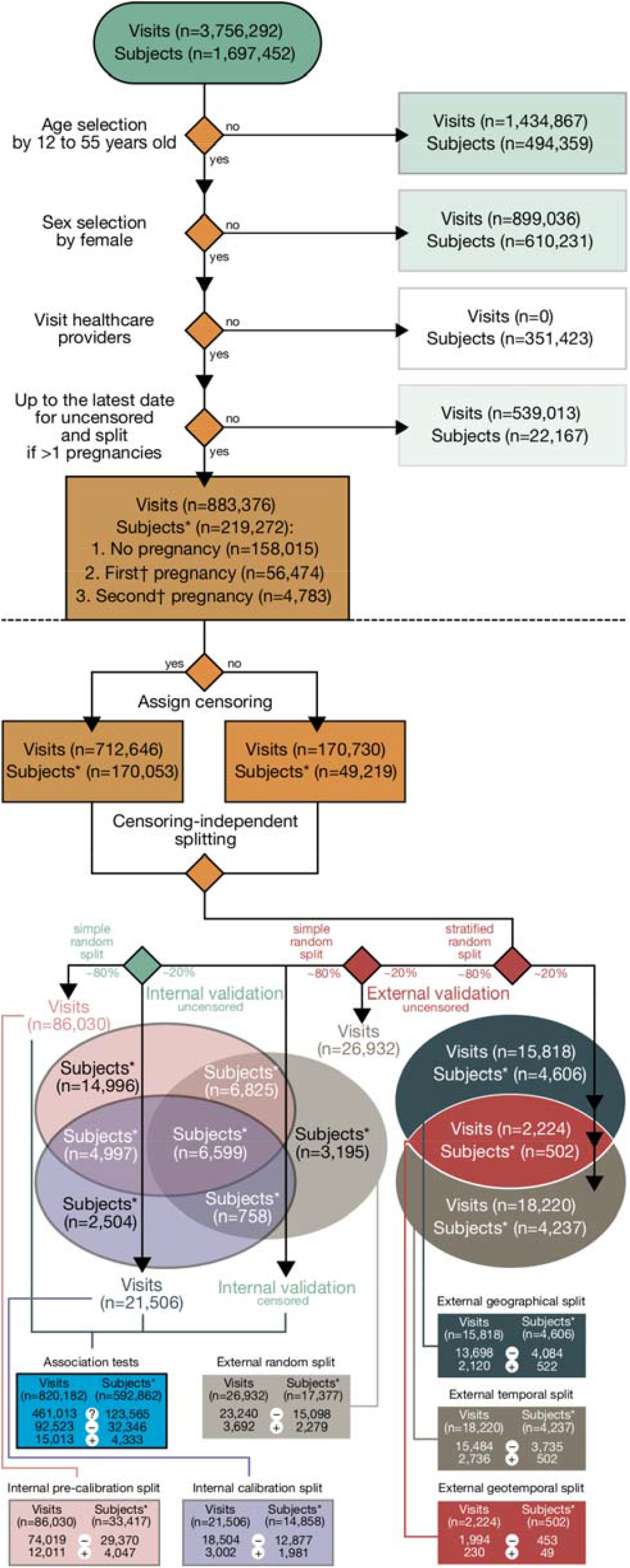
Subject selection by applying a retrospective design and data partitioning for internal and external validations. The set for association tests included censored outcomes. n, sample size; *, subject per pregnancy episode; **, the first and second pregnancies of a subject within the database period, not a parity; (?) censoring; (–) nonevents; and (+) events.

Calibration (plot, intercept, and slope) and discrimination (i.e. AUROC) were computed for classification tasks. We also computed sensitivity, specificity, positive likelihood ratio (LR+), and negative likelihood ratio (LR-). For estimation, we computed a proportion of weeks in which each predicted time, in weeks, was included within an interval estimate of the true one. The interval had to be the maximum ± *x* weeks when predicting > *x* weeks, e.g., for any women predicted to deliver in 6 weeks, this number should fall into the true time of delivery within ± 6 weeks. We determined the minimum and maximum predicted times of delivery with acceptable precision for each predicted outcome of PROM based on a visual assessment using internal validation. Root mean square error (RMSE) was computed within this acceptable range. We also evaluated the best time window using the best model for PROM prognostic predictions. The internal validation set was grouped by binning the days to the end of pregnancy every 4 weeks. An AUROC was computed for each bin. The best time window should be mostly greater than an AUROC of 0.5, which represents prediction by simple guessing.

Success criteria of the modeling were an AUROC greater than those of recent models (last 5 years) of PROM prognostic predictions using simple predictors (e.g., maternal factors), or greater or equal to those using high-resource predictors (e.g., biophysical or biochemical markers). To prevent a common cause of overfitting, we applied the PROBAST criterion, which is, the number of events in the training set should be ≥20 after being divided by the number of candidate predictors. We applied the preferred reporting items for systematic reviews and meta-analyses (PRISMA) 2020 expanded checklist (eTable 4 in the Supplement) to find the comparable models.^42^ Details on this procedure are described in the Supplement.

## Results

We selected all visits (*n*=883,376) by 12∼55-year-old women (*n*=219,272) (Figure 1 and Table 1). Of 12 latent candidate predictors, eleven had significant association with PROM, estimated by IPW (Table 2).

**Table 2.** Association between each latent candidate predictor and PROM by inverse probability weighting.

| Variable of interest | Unadjusted OR (95% CI; P value) | Adjusted OR (95% CI; P value) | Adjustment |
| --- | --- | --- | --- |
| Multiple pregnancy | 1.038 (1.034 to 1.043; P<.001***) | 1.062 (1.055 to 1.068; P<.001***) | Maternal age |
| Chorioamnionitis | 1.394 (1.373 to 1.416; P<.001***) | 1.351 (1.33 to 1.372; P<.001***) | Asthma + Influenza + Intra-amniotic infection |
| Intra-amniotic infection | 1.119 (1.085 to 1.155; P<.001***) | 1.118 (1.083 to 1.153; P<.001***) | Genital tract infection + Periodontal disease + Pneumonia + Multiple pregnancy |
| Ante-partum hemorrhage | 0.924 (0.921 to 0.928; P<.001***) | 0.929 (0.924 to 0.933; P<.001***) | Low socio-economic status + Maternal age |
| Genital tract infection | 1.116 (1.101 to 1.132; P<.001***) | 1.116 (1.101 to 1.132; P<.001***) | (no adjustment) |
| Periodontal disease | 0.92 (0.917 to 0.922; P<.001***) | 0.967 (0.96 to 0.973; P<.001***) | Asthma + Maternal age |
| Polyhydramnios | 1.039 (1.029 to 1.049; P<.001***) | 0.998 (0.989 to 1.006; P>.99) | Multiple pregnancy |
| Pneumonia | 0.979 (0.97 to 0.987; P<.001***) | 1.037 (1.025 to 1.049; P<.001***) | Asthma + Influenza |
| Asthma | 0.958 (0.953 to 0.962; P<.001***) | 0.971 (0.966 to 0.977; P<.001***) | Influenza |
| Low socio-economic status | 0.979 (0.978 to 0.98; P<.001***) | 0.979 (0.978 to 0.98; P<.001***) | (no adjustment) |
| Maternal age | 0.969 (0.969 to 0.97; P<.001***) | 0.969 (0.969 to 0.97; P<.001***) | (no adjustment) |
| Influenza | 0.995 (0.993 to 0.997; P<.001***) | 0.995 (0.993 to 0.997; P<.001***) | (no adjustment) |

\* P<0.05; \*\* P<0.01; \*\*\* P<0.001; CI, confidence interval

We depicted the final association diagram (eFigure 13 in the Supplement). This diagram and the association findings are further described in the Supplement.

### Prognostic prediction of prelabor rupture of membranes (PROM)

After calibration (Figure 2a), the two well-calibrated models were the PC-ENR and DI-VNN; however, distributions of predicted probabilities from 0 to 1 were mostly covered by the latter model. For clinical application, this will help to widely adjust threshold, depending on the local data distribution. The optimal threshold for the DI-VNN was 0.14. Both models had visually differentiated distributions of predicted probabilities between events and nonevents. Weights, variable importance values, and intermediate outputs, which indicated the extent a predictor contributes to a prediction, are respectively shown for (1) the RR and PC-ENR (eTables 14 to 16 in the Supplement); (2) the PC-RF and PC-GBM (eTables 15, 17, and 18 in the Supplement); and (3) the DI-VNN (eTables 19 and 20 in the Supplement).

**Figure 2.**
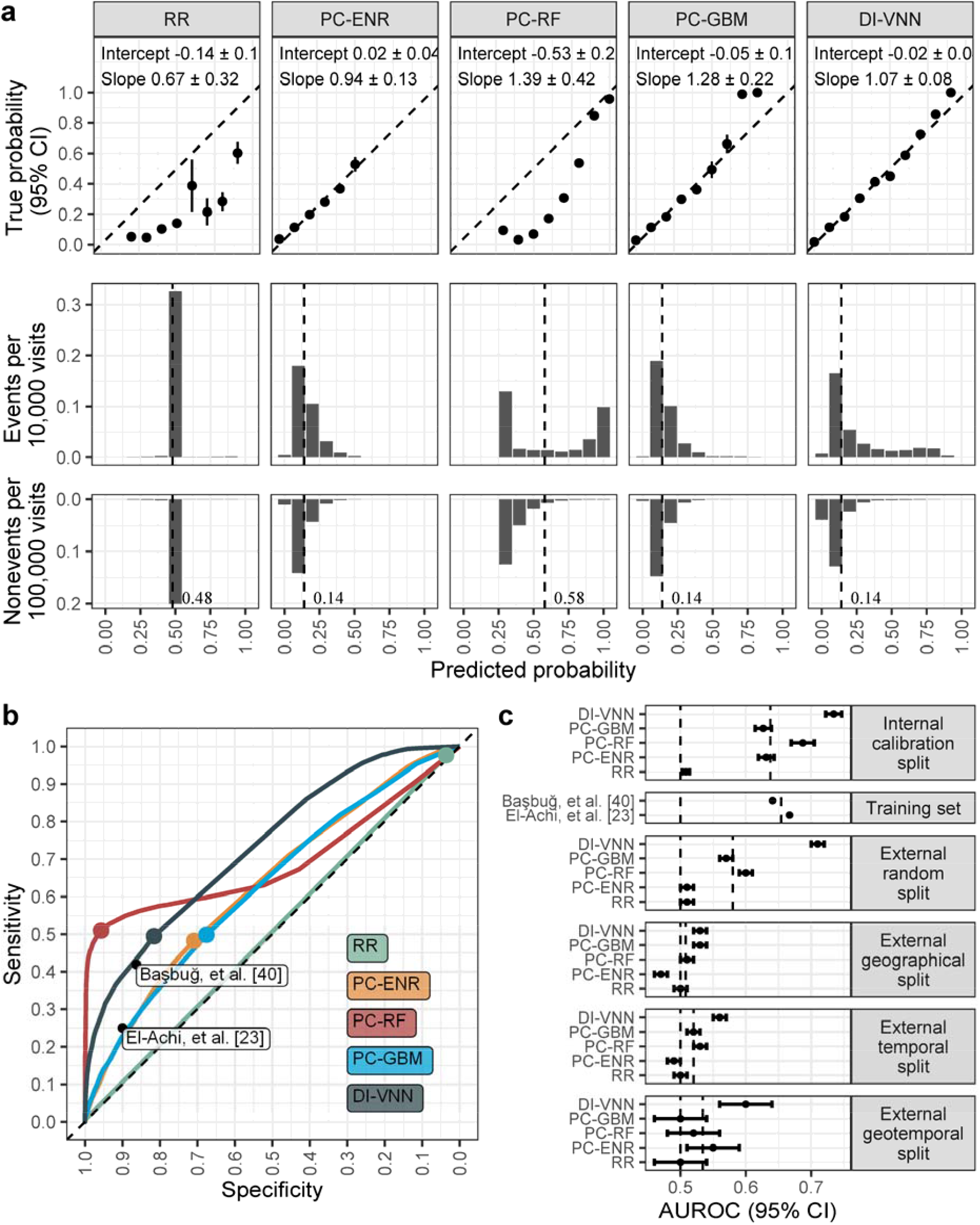
Model evaluation. (a) calibration; (b) receiver operating characteristic (ROC) curve; (c) area under ROC curve (AUROC). Thresholds (a, b) and average AUROCs per set (c). DI-VNN, deep-insight visible neural network; ENR, elastic net regression; GBM, gradient boosting machine; PC, principal component; RF, random forest; RR, ridge regression.

At 95% specificity, the PC-RF (0.513, 95% CI 0.509 to 0.517) was the most sensitive model (Figure 2b); unfortunately, this model was not well-calibrated. At the same specificity, the DI-VNN followed PC-RF with a sensitivity of 0.297 (95% CI 0.293 to 0.301; threshold at 0.29). With the optimal threshold at 0.14, the DI-VNN achieved a sensitivity of 0.494 (95% CI 0.489 to 0.5) and a specificity of 0.816 (95% CI 0.814 to 0.818). Potential utility of DI-VNN was also confirmed by LR+ (2.68, 95% CI 2.63 to 2.75) and LR- (0.62, 95% CI 0.61 to 0.63). By external validation (Figure 2c), the DI-VNN was the most robust.

For the random split that reflected common situations nationwide, the DI-VNN achieved an AUROC of 0.71 (95% CI 0.70 to 0.72). It was reasonably lower than that of a training set (0.73, 95% CI 0.72 to 0.75). For other external validations, the AUROCs of the DI-VNN were lower than that for the random split but higher than both the average AUROCs of all models and an AUROC of 0.5.

From PubMed, Scopus, and Web of Science, we identified 209 non-duplicated records. These were screened, retrieved, and assessed to find two prediction models^23,43^ for preterm PROM (eFigure 14 and eTables 4 and 5 in the Supplement). Prognostic predictions of PROM by the DI-VNN and PC-RF achieved AUROCs that were higher than those of previous models (Figure 2b and 2c): (1) a logistic regression using maternal factors with an AUROC of 0.667 (sensitivity 0.25; specificity 0.90; LR+ 2.50; and LR-0.83) but without internal validation (144 preterm PROM; 10,136 not preterm PROM);^23^ and (2) a prediction rule using serum alpha-fetoprotein with an AUROC of 0.641 (sensitivity 0.419; sensitivity 0.863; LR+ 3.06; and LR-0.67) but without internal validation (31 preterm PROM; 1146 not preterm PROM).^43^

For an exploratory data analysis, most of the AUROC intervals were greater than 0.5 from 44 ± 2 weeks before the end of the pregnancy (eFigure 15a in the Supplement). Population-level data exploration is also extensively described in the Supplement. An interactive interface of the DI-VNN are provided in our web application (https://predme.app/promtime), allowing users to explore this model at both the population and individual levels.

### Estimation of the time of delivery

Although term PROM may also happen, it is mostly related to preterm delivery. Since codes for preterm delivery or premature newborn might not be consistently assigned to all cases, we decided to estimate how many days from the current visit a mother would deliver. This estimation task is providing a benefit if the model could estimate the time of delivery within an interval estimate of a maximum ± *x* weeks when predicting > *x* weeks. We found the highest 78.57% of the 42 weeks were precisely predicted by PC-RF according to those criteria (Figure 3a). The time estimated by the DI-VNN was unfortunately within only the 6^th^ week from the prediction dates of any sample. This was due to the differential analysis (see Methods), which filtered predictors based on categorical outcomes only. Nonetheless, we used the DI-VNN, as the best classification model, to stratify the estimated time of delivery, including that estimated by the PC-RF. We confirmed this model consistently outperformed the other models for the time estimation task using external validation sets. A comparison of estimation performances, including those by external validation sets, is further described in the Supplement.

**Figure 3.**
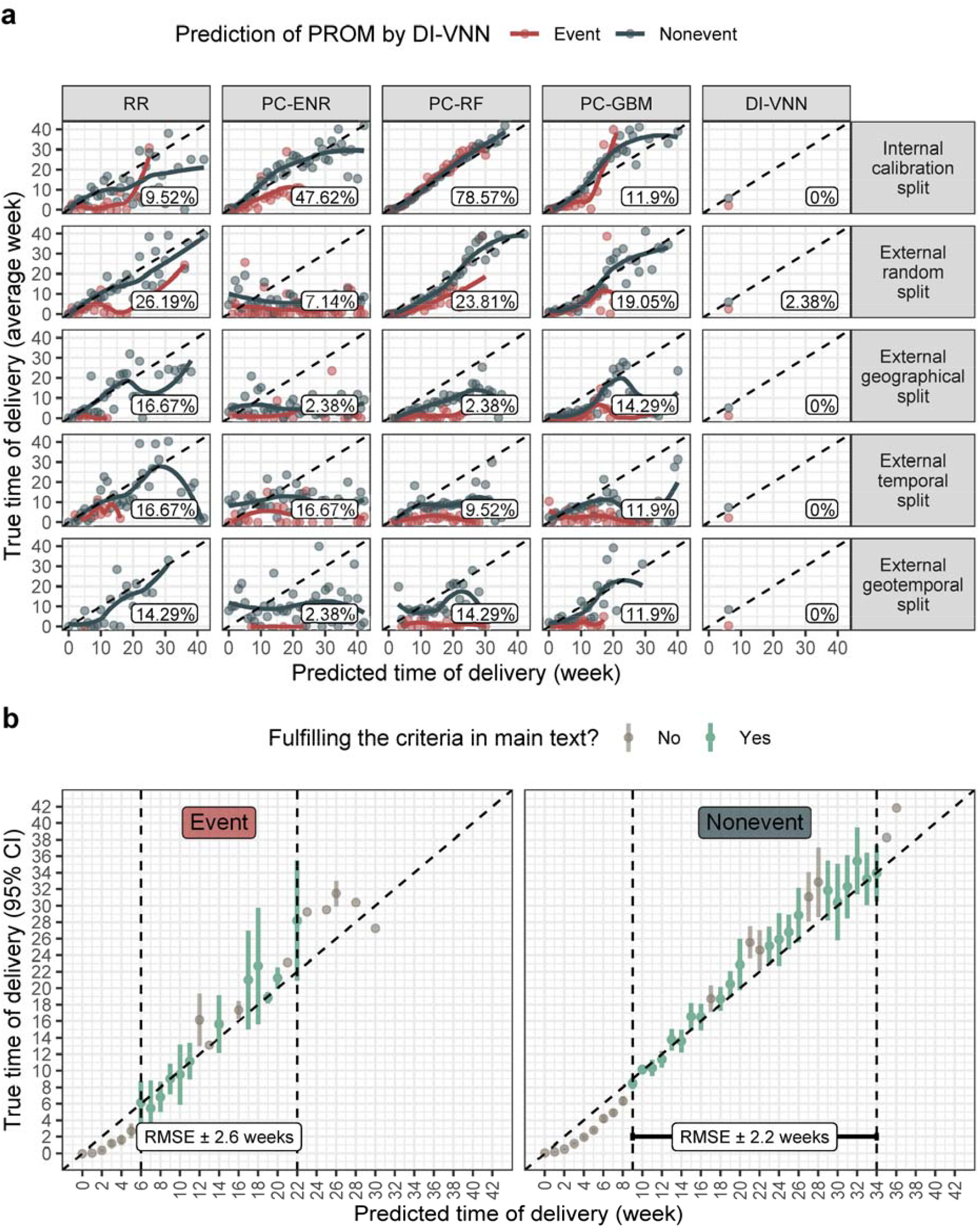
Estimation plots. (a) model comparison; (b) principal component-random forest (PC-RF) estimation window. Percent % criteria fulfilled (a) and precision (95% confidence interval) per week (b). DI-VNN, deep-insight visible neural network; ENR, elastic net regression; GBM, gradient boosting machine; RMSE, root mean squared error; RR, ridge regression.

With nonevents, the maximum estimated time of delivery by PC-RF was 36 weeks from a visit when the prediction was conducted, while the corresponding true time of delivery was 42 weeks (Figure 3b). This coincides with the maximum duration of the pregnancy. With events, the maximum predicted time was 6 to 10 weeks earlier.

### Web application

A web application was provided using the best models. We hosted this application on a public repository for clinical prediction models (https://predme.app/promtime). This allows a user, e.g., a doctor, to make a decision after critically appraising an individual prediction by several approaches. We describe a case example in the Supplement, reflecting a real-world situation.

## Discussion

The DI-VNN model in this study outperformed previous models^23,43^; it used a large training set and external validation sets (8778 visits and 3352 subjects for events only) and did not require biomarker testing. External validation by stratified random splitting was also applied to predict gestational diabetes using nationwide health insurance database.^44^ The PC-RF model also estimated the time of delivery in weeks to predict preterm delivery, while previous models only predicted whether a preterm delivery would happen without estimating the date interval.^24^ Our models also used a cohort paradigm to prevent temporal bias in the delivery prediction.^45^ We did not only rely on the DI-VNN and PC-RF models but also datasets to estimate the model performances at the individual level based on subpopulations similar to that individual,^46^ in a collaboration of machine learning and evidence-based medicine.^47^ This framework also answers key challenges to evaluate model weaknesses,^48^ verify if the predicted outcome is reasonable,^49^ and make sensible clinical predictions by utilizing electronic medical records.^50^

As inferred from the PC-RF model, the time of delivery of PROM events was estimated earlier than that of nonevents. This disease may be considered an infection-related preterm delivery. Clustering analysis of placental gene signatures assigned this type of preterm delivery as a subclass of preeclampsia.^51^ The DI-VNN model in this study also included medical histories of all subtypes of preeclampsia (eTable 20 in the Supplement). The DI-VNN model likely used preeclampsia-related codes as competing risks to predict PROM. Similarly, competing risk models were also previously developed for preeclampsia.^52^ Exploratory data analysis in this study also demonstrated optimal predictive performances that cover a full pregnancy period and a month before the pregnancy. This is also similar to findings based on predictive modeling for preeclampsia.^25^

However, we noted several limitations in our study. Although the DI-VNN outperformed those from previous studies,^23,43^ we could only apply it as a preliminary model for PROM because clinical acceptance requires an AUROC of ≥0.8.^53^ More-sensitive models are still needed for use as second-line models. As recommended in a clinician checklist^53^ for assessing the suitability of machine learning applications in healthcare (eTable 3 in the Supplement), external validation and determining an optimal threshold using local data are needed.

In conclusion, our DI-VNN model was found to be robust for PROM prognostic predictions. The PC-RF model was reasonably precise within a specific time window based on the predicted outcome by the DI-VNN. An impact study (i.e. clinical trial) are warranted to further assess these models.

## Data Availability

The data that support the findings of this study are available from the social security administrator for health or badan penyelenggara jaminan sosial (BPJS) kesehatan in Indonesia, but restrictions apply to the availability of these data, which were used under license for the current study (dataset request approval number: 5064/I.2/0421), and so are not publicly available. Data are however available from the authors upon reasonable request and with permission of the BPJS Kesehatan. To get this permission, one need to request an access from the BPJS Kesehatan for their sample dataset published in August 2019. Up to this date, there are three sample datasets they published in February 2019, August 2019, and December 2020. For the first and second versions, a request is applied via https://e-ppid.bpjs-kesehatan.go.id/, while the third is applied via https://data.bpjs-kesehatan.go.id. The R Markdown, R Script, and others are available in https://github.com/herdiantrisufriyana/prom. To pre-process the raw data into the input dataset of this study, follow the codes of the R Markdown in https://github.com/herdiantrisufriyana/medhist/tree/main/preprocessing.

https://e-ppid.bpjs-kesehatan.go.id/

https://data.bpjs-kesehatan.go.id

https://github.com/herdiantrisufriyana/medhist/preprocessing

## Acknowledgments

The BPJS kesehatan in Indonesia gave permission to access the sample dataset in this study. This study was funded by the Ministry of Science and Technology (MOST) in Taiwan (grant number MOST109-2221-E-038-018 and MOST110-2628-E-038-001) and the Higher Education Sprout Project from the Ministry of Education (MOE) in Taiwan (grant number DP2-110-21121-01-A-13) to Emily Chia-Yu Su. These funding bodies had no role in the design of the study and collection, analysis, and interpretation of data and in writing the manuscript.

## Author contributions

HS, YWW, and ECYS developed the concept and design of this study (DBPR). Dataset access was requested by HS. This author extracted and processed the data, performed the training and validation of the machine learning algorithms, conducted the literature search, and wrote a draft of the manuscript. This author and YWW independently assessed the eligibility criteria of ambiguous, reviewed studies which were previously determined by HS. YWW and ECYS critically revised the draft manuscript. All authors approved the submitted manuscript and agreed to be personally accountable for their own contributions and to ensure the accuracy and integrity of any part of the work, including ones in which the author was not personally involved.

## Competing interests

HS, YWW, and ECYS declare no competing interests.

## Data availability

The data that support the findings of this study are available from the social security administrator for health or *badan penyelenggara jaminan sosial (BPJS) kesehatan* in Indonesia, but restrictions apply to the availability of these data, which were used under license for the current study (dataset request approval number: 5064/I.2/0421), and so are not publicly available. Data are however available from the authors upon reasonable request and with permission of the BPJS Kesehatan. To get this permission, one need to request an access from the BPJS Kesehatan for their sample dataset published in August 2019. Up to this date, there are three sample datasets they published in February 2019, August 2019, and December 2020. For the first and second versions, a request is applied via https://e-ppid.bpjs-kesehatan.go.id/, while the third is applied via https://data.bpjs-kesehatan.go.id.

## Code availability

The R Markdown, R Script, and others are available in https://github.com/herdiantrisufriyana/prom. To pre-process the raw data into the input dataset of this study, follow the codes of the R Markdown in https://github.com/herdiantrisufriyana/medhist/tree/main/preprocessing.

## Notes

### Competing Interest Statement

The authors have declared no competing interest.

### Author Declarations

The dataset was opened publicly by request. The BPJS Kesehatan had already approved our request (dataset request approval no.: 5064/I.2/0421). The dataset had been already deidentified before going public; thus, the ethical clearance to Institutional Review Board of Taipei Medical University was waived.

### Summary of Updates

This version focuses on clinical audience for applying the prediction model. All computational/methodological aspects are out of scope of this paper and described elsewhere as protocol papers. These are intended for more general implementation of the proposed protocols, fully or partially implemented in our projects and collaborations. Yet, methods in this paper are sufficiently detailed for replicating this study while also referring to the protocol papers.

