## Supplement for "Prognostication for prelabor rupture of membranes and the time of delivery in nationwide insured women: development, validation, and deployment"

Please kindly see List of Figures and List of eTables in Appendix.

### Introduction

In this Supplement, we describe details on this study following chronological order of our analysis pipeline on association test or predictive modelling for prelabor rupture of membranes (PROM). There are four sections corresponding to the same sections in the main text, which are respectively Introduction, Methods, Results, and Discussion. Along with this PDF document, we also provide R Markdown (.Rmd) containing the same texts with this document but including the programming codes for the data analysis in-between of these texts. An exception is subsection of "Comparison to previous studies". The codes for core steps in the analysis pipeline are also provided exclusively in an R Script (.R). The codes beyond the core steps were used for analytic decision or creating tables or figures. These are shown to provide details on how data are processed to construct all tables and figures in both the main text and this Supplement, including those in Appendix of this Supplement (eTables 1 to 5, and 13) and those shown only in the R Markdown due to the complexity (eTables 6 to 12 and 14 to 21). A 5-minute video was provided in the protocol^1^ to briefly explain technical details on deep-insight visible neural network (DI-VNN) pipeline.

### Methods

#### Research guidelines

To ensure that we conducted rigorous research, we carried out and reported this study by applying three sets of standard guidelines, specifically designed for a multivariable prediction model applying a machine learning algorithm that is suitable for healthcare.^2-4^ The checklists for all the guidelines are shown (eTables 1 to 3 in Appendix). For a fair comparison to previous models, we also followed methods in the preferred reporting items for systematic reviews and meta-analyses (PRISMA) 2020 expanded checklist.^5^ The checklist and the comparable models are also described (eTables 4 and 5 in Appendix). We also applied four approaches based on previous methods^6-13^ to develop the pipeline with several modifications to provide a framework for improving interpretability of a deep learning model.^1^ All of the aforementioned procedures were comprehensively described in a human and machine learning pipelines we developed for responsible clinical prediction.^14^

#### Programming environment

We set up a programming environment for this study. Bioconductor was utilized as described in the main text. There were 198 R packages which are 9 base packages, 53 other packages, and 136 dependencies (eTable 6 in R Markdown).

#### Sampling procedures of the data source

The data source was a sample dataset of the whole health insurance database during 2015 and 2016 by cross-sectional design. Stratified random sampling was applied. The strata variable was constructed from 66,072 combinations of all the healthcare facilities (*n*=22,024) and category of family, which were: (1) a family of which members never visit the healthcare facilities; (2) a family of which members have visited only primary care; and (3) a family of which members have visited all levels of care. For each stratum, one to ten families were randomly included. This means only ten families were randomly included if more than that number, resulting 586,969 families with 1,697,452 subjects.

#### The sampling procedures of the dataset in this study

We conducted non-essential data cleaning, e.g. revising the inconsistent name of states, estimating the healthcare identifiers, *et cetera*. These procedures were parts of our R package of medhist 0.1.0. No sampling was conducted.

After the non-essential data cleaning, we applied retrospective cohort design, as described in the main text. For pregnant women, we use several codes for determining delivery or immediately after delivery care. The 220 codes are described (eTable 7 in R Markdown).

#### Data pre-processing

We conducted data pre-processing after defining the target population and sampling it retrospectively. Demographics were included as categorical variables for association tests. Then, we applied systematic human learning, as described in the main text, to determine what were latent candidate predictors that can be inferred from our dataset. We also computed a number of days for a code in the latest encounter before the time of prediction, including those by codes as a latent candidate predictor.

#### Data partition

To ensure all inference or derivation using training set only, we need to conduct data partition before continuing the downstream analysis. We described data partition for model validation in the main text (see Methods).

#### Association tests

We conducted association tests as described in the main text. This will help us to include only the confirmed associated predictors as latent candidate predictors before conducting pre-selection of those candidates to fulfill quality control of predictors in the main text. We included latent candidate predictors of which the data were available in training set. Details on this information and International Classification of Disease version 10 (ICD-10) codes or demographical variables for each candidate of latent candidate predictors are shown in the next section. A protocol of systematic human learning was followed for these association tests.^15^

#### Quality control of candidate predictors

For determining candidate predictors, we utilized nationwide medical histories, excluding those for validation. But, for the downstream analysis, medical histories were provider-wise by estimation. This means our prediction models only used medical histories recorded by a healthcare provider, which was blinded to those recorded by others. This reflects most real-world situations in which a healthcare provider does not have access to medical records of other providers.

All candidate predictors, including non-demographical associated factors, have non-zero variances (eTable 8 in R Markdown). There were 460 candidate predictors fulfilling this criterion. We also showed in the same eTable that there are 426 candidate predictors without perfect separation.

To prevent outcome leakage, we removed the maternal or baby diagnosis/procedure codes that demonstrated the delivery or after delivery care (typically up to 6 weeks following childbirth; eTable 9 in R Markdown). We only used the existing codes in the training set to determine outcome-leaker codes based on the previous codes for determining delivery or immediately after delivery care (eTable 7 in R Markdown). There were 54 codes that may leak the outcome.

To avoid redundancy, we computed pair-wise Pearson correlation coefficients (eTable 10 in R Markdown). None of the estimates showed a perfect correlation (*r*=1). High correlations (i.e., >~0.70) were reasonably identified between latent candidate predictors and the code components. We did not remove those pairs of predictors.

By systematic human learning and association tests using available data, we also determined latent candidate predictors (eTable 11 in R Markdown). There were 27 first- and 10 second-level factors that are considerably associated with PROM. Only data for 12 out of 27 factors were available in training set. Either the diagnosis/procedure codes, or demographical variables (not included as candidate predictors), for latent candidate predictors are also described (eTable 12 in R Markdown).

#### Feature extraction as historical rates

To deal with a problem in which a healthcare provider does not have access to medical records of other providers, medical histories were quantified as Kaplan-Meier (KM) estimators for each candidate predictors, so called historical rates. We inferred these rates given the day number from a code encounter to current visit for each candidate predictor, derived only from the same nationwide medical histories, as previously described.^16^ This used irredundant candidate predictors with non-zero variances and no perfect separation in training set only. The candidate predictors were transformed into the historical rates in all data partitions.

#### Feature representation as principal components by 10-fold cross validation

The historical rates of all candidate predictors were fitted to a principal component (PC) model. Only nationwide medical histories in training set were used for the model fitting. We applied 10-fold cross validation to estimate weights for all candidate predictors in each PC. A protocol for resampled dimensional reduction was followed for feature representation in this study.^17^

#### Set up tuning-training-calibrating configuration and internal validation

Previous data partition had not held out instances for calibration yet. This took 80% of training set. We also gave different weights for event and non-event by including censored outcome, as described in the main text. For hyperparameter tuning, we applied 5-fold cross validation, instead of 10-fold as applied for PC modeling. Meanwhile, the final training and calibration for each model were conducted by bootstrapping for 30 times. The same resampling methods were applied for both classification and estimation tasks. Parallel computing by multiple central processing units (CPUs) were applied for training all models.

#### Hyperparameter tuning, final training, and calibrating

We applied the tuning grids and the training configurations for all models, except DI-VNN which required several modifications. This is already described in a protocol we followed for these procedures, as previously described.^14^ More details for DI-VNN will be described in the next section.

#### Deep-insight visible neural network (DI-VNN)

We applied different pre-processing pipelines for feature selection and representation in DI-VNN. Instead of PCs, we used the historical rates of the candidate predictors. A 5-minute video explaining DI-VNN pipeline is available in the protocol.^1^ Only pre-calibration training set was used for the downstream pipeline. We followed a protocol for this model.^1,14^ Briefly, we applied differential analysis for feature selection. Then, 1-bit stochastic gradient descent transformation was applied using the post-normalization, feature-wise average based on nationwide training set to determine if a value is lower, equal to, or higher than the average respectively as -1, 0, and 1. Using a feature-to-feature Pearson correlation matrix, we created a feature map *t*-stochastic neighbor embedding (*t*-SNE) with Barnes-Hut approximation. We also subset this map hierarchically by clique-extracted ontology (CliXO) algorithm based on the same correlation matrix. We constructed convolutional neural network (CNN) model using the hierarchy with input of feature map onto which transformed, candidate predictors were projected. We trained the CNN model to get the learning representation.

#### Evaluating the best model for classification and estimation

Model evaluation is already clearly described in the main text. Practical costs of prediction errors were considered when evaluating the models. Under-prognosis may cause pregnancy monitoring to be off-guard. A pregnant woman with a preterm delivery may not reside in an area with a readily available NICU, particularly in low-resource settings. Over-prognosis may lead to unnecessary enrollment of patients into a cohort study in an early intervention for preventing PROM or complications, e.g., an antibiotic administration. This may also lead to unnecessary tests for more-specific prognostications.

In addition, unlike other time-varying outcomes, e.g., cancer, we did not predict a survival rate for the estimation task because the time interval for a pregnancy period is definitely known. It is also more intuitive for clinicians if the given information is the estimated days from the current visit to the day a pregnant woman will deliver a baby, as such normal delivery estimations are based on the last menstrual period and ultrasound examination.

We started from evaluating the calibration measures of all models for classification task. Then, we chose the best model for classification by AUROC using only internal validation (calibration split) bootstrapped for 30 times. Using the same subset, the best estimation model was also chosen.

#### External validation

We also confirmed the robustness of the best models using external validation sets. By stratified random splitting, we provided three non-overlapping splits comprising ~20% for external validation: (1) geographical split; (2) temporal split; and (3) geotemporal split. We set these subsets challenging to predict by our models, in which the geotemporal split was the most difficult. This way we can estimate robustness of our model generalizability. In addition, uncertainty intervals were also computed by bootstrapping for 30 times. But, these do not reflect common situations nationwide; thus, the ~20% of the remaining was held out by simple random splitting for external validation, which is called external random split. For association tests and predictive modeling, including calibration, we only used the remaining ~64% of all the selected visits. A clear description on model validation is given in a protocol we followed for these procedures, as previously described.^14^

#### Exploring the best model

Model exploration method is also described in the main text (see Model evaluation in Methods). In this Supplement (see R Markdown and R Script), we directly explored the best model for classification task. None of the remaining models were explored.

#### Comparison to previous studies

Previous prognostic models were developed for preterm PROM. Meanwhile, we also did not have data on gestational ages. Therefore, we had to develop both PROM prognostic prediction and the time of delivery estimation in days from the time of prediction. For simplicity, we did not compare our estimation model to those for estimating the time of delivery. Instead, we compared our PROM prediction model to previous models which were developed for preterm PROM. To find comparable models, we applied PRISMA 2020 guidelines to evaluate success criteria.^5^ To achieve that purpose, following the PICOTS framework, eligibility criteria included: (1) population, either pregnant or non-pregnant women without specifying their medical conditions; (2) index, the best prediction model in this study; (3) comparator, prediction models or rules; (4) outcome, either preterm or term PROM as a binary outcome (event or nonevent) with ≥20 EPVs; (5) time, prognostic prediction from days to weeks; and (6) setting, either primary care or hospital patients. We excluded any article types beyond original articles, including conference abstracts but not full papers. The studies were not grouped since no synthesis was conducted.

We searched for studies up to April 3, 2021. Unlike human learning that only used PubMed, we also included Scopus and Web of Science as literature/bibliographic databases in which we searched for previous models, because prediction studies were also extensively reported beyond life-science journals (i.e., computer-science journals). The keyword was “prelabor rupture of membranes prediction”. We limited the records to within the last 5 years and the English language. We applied a similar, but not exactly the same, search strategy to the three databases considering their different interfaces (eTable 4 in the Supplement). One researcher, HS, searched the literature and loosely filtered these by title and abstract. Then, HS manually assessed the eligibility by examining the full texts and identified ambiguous ones. HS and YWW independently assessed ambiguous studies. If no consensus was reached between HS and YWW, a final decision was made by ECYS. HS collected data from each full text. We only extracted the sensitivity and specificity of the best model with the most similar outcome definition from each study to that of our study. The AUROCs were also included. We followed the previous protocol for the remaining procedures to find these comparable models.^14^

#### Preparing web application

For web application, we prepared an example dataset, user interface, and processing script at the side of server computer. No line of codes for the web is included in the R Markdown or R Script. Description for this web application is already clearly described in the main text.

### Results

In the R Markdown, we show data for Figure 1 in the main text. Up to the latest date for uncensored outcome and after splitting if >1 pregnancies, the total visit was the sum of totals from all subsets, while the total subject was the sum of totals from all subsets with attention for the overlaps (d to j in footnote of data for Figure 1 in the R Markdown). For association tests, visits and subjects of the censored outcome were those after external random splitting (k and j in footnote of data for Figure 1 in the R Markdown). We also explicitly show PROM prevalence for each subset.

#### Association diagram

For latent candidate predictors whose data were available, we created the association diagrams (eFigures 1 to 12 in Appendix). We excluded all common effects of PROM that we found during the systematic human learning. These are not needed for association tests. Instead, inclusion of these variables will cause collider-stratification bias. In the association diagrams, we apply different colors based on the types of nodes representing several factors: (1) type A is a first-level factor (with variable prefixed by A) that has a role as a confounder; (2) type I is a first-level factor (with variable also prefixed by A) that has a role as a candidate factor of interest; (3) type U is an unmeasured variable that can affect measure variable of type-A/I/Y variable; and (4) type Y is a target or dependent factor which is PROM. Node, of which variable denoted by asterisk, represents a type-A/I/Y variable that can be represented by several diagnosis or procedure codes. In each of the diagrams, we also show adjustment formulas which were used for association tests. Conceivably, all variables of type A/I/Y with asterisk may be included in each formula, but, some of these variables cannot be included because these were not available in our data, particularly in the training set (likely because of low prevalence). All measurement errors that may be affected by Type-U variables in this study were assumed as independent non-differential errors, because all data were measured from electronic medical records. As the main text, results of the association tests were also described (eTable 13 in Appendix), either by outcome regression or inverse probability weighting (IPW).

We would describe each association diagram. Fifty-six studies were found from PubMed (eTable 11 in R Markdown).^18-73^ Since only some factors are possible to include in the association tests, only some of these studies were explained below. After verifying the assumption using our data, we constructed a final association diagram consisting all the confirmed latent candidate predictors.

##### Multiple pregnancy

Association model of multiple pregnancy on PROM^18^ included only maternal age as a confounder.^19,20^ However, several confounders were not blocked yet: (1) Assisted reproduction;^21,22^ and (2) Race.^20,23^ These are shown in the association diagram (eFigure 1 in Appendix). Assisted reproduction can be represented by diagnosis/procedure codes but unavailable in our training set. For race, it is conceivably not able being represented by those codes.

##### Chorioamnionitis

There were two assumptions regarding relationship between chorioamnionitis and PROM. Chorioamnionitis may affect or be affected by PROM. We applied chorioamnionitis as the former one (eFigure 2 in Appendix), as previously demonstrated.^24^ Similar assumptions were also regarded between chorioamnionitis and intra-amniotic infection (IAI), but, we only treated chorioamnionitis as being affected by IAI.^25^ Except cigarette smoking,^18,26^ data for all confounders were available in the training set: (1) influenza;^26,27^ (2) asthma;^28^ and (3) IAI.^18,25^

##### Intra-amniotic infection (IAI)

Similar to chorioamnionitis, IAI may affect or be affected by PROM. Consistently, we treated IAI as the former one (eFigure 3 in Appendix).^18,25^ We did not have data for these confounders, especially in training set: (1) cervical shortening;^18,29^ and (2) race.^23,30^ Therefore, we used these factors as the confounders: (1) GTI;^25,31-33^ (2) periodontal disease;^34,35^ (3) pneumonia;^35,36^ and (4) multiple pregnancy.^18,37^

##### Ante-partum hemorrhage (APH)

Most data for confounders of ante-partum hemorrhage (APH) and PROM^18^ were not available in the training set (eFigure 4 in Appendix). Only two confounders were adjusted: (1) low socioeconomic status (SES);^18,38^ and (2) maternal age.^19,39^ The other confounders were: (1) cigarette smoking;^18,38^ (2) illicit drug use;^18,38^ (3) race;^23,40^ (4) assisted reproduction;^21,41^ and (5) placenta on anterior wall.^39,42^

##### Genital tract infection (GTI)

Association between GTI and PROM^31,32^ was only confounded by tuberculosis.^43,44^ We did not have data for tuberculosis in the training set. Thus, GTI is the only variable in the association model (eFigure 5 in Appendix).

##### Periodontal disease

We also found association between periodontal disease and PROM.^34^ An association model was constructed by adding these confounders: (1) asthma;^28,45^ and (2) maternal age.^19,46^ Because of data availability, we could not include these common confounders into the model: (1) stress;^46,47^ (2) low education;^46,47^ and cigarette smoking (eFigure 6 in Appendix).^18,46^

##### Polyhydramnios

Polyhydramnios was also an associated factor of PROM.^48^ The confounders were: (1) assisted reproduction;^21,22^ and (2) multiple pregnancy.^18,49^ Only multiple pregnancy data were available in our training set; thus, a PROM association model was constructed using polyhydramnios and multiple pregnancy (eFigure 7 in Appendix).

##### Pneumonia

Association model of pneumonia on PROM^36^ was confounded by two factors. The first confounder was influenza,^27,50^ while the second one was asthma.^28,50^ Both were included in the association model (eFigure 8 in Appendix).

##### Asthma

As shown in the association model of pneumonia and PROM, asthma was also an associated factor of PROM.^28^ Influenza was the only confounder.^27,51^ Therefore, we included this confounder in the association model of asthma on PROM (eFigure 9 in Appendix).

##### Low socioeconomic status (SES)

Low SES was also indicated as an associated factor of PROM^18^. We could not find the confounder. The association model only included low SES. We represented several demographical factors as low SES (eFigure 10 in Appendix).

##### Maternal age

We could find maternal age as a confounder of PROM and multiple pregnancy/APH/ periodontal disease. Obviously, there is no confounder of PROM in an association model with maternal age as the variable of interest.^19^ We included this variable exclusively in the association model (eFigure 11 in Appendix).

##### Influenza

Similar to maternal age, obviously there is no confounder of PROM in an association model with influenza as the variable of interest ^27^. This disease has a specific agent. Therefore, the PROM association model only included this disease (eFigure 12 in Appendix).

##### Final association diagram

Chorioamnionitis (odds ratio [OR] 1.351, 95% confidence interval [CI] 1.33 to 1.372), intra-amniotic infection (OR 1.118, 95% CI 1.083 to 1.153), and genital tract infection (OR 1.116, 95% CI 1.101 to 1.132) were the top three highest effects estimated by IPW. Polyhydramnios was not verified as being associated with PROM by either IPW (OR 0.998, 95% CI 0.989 to 1.006) or the outcome regression (OR 1.238, 95% CI 0.851 to 1.801) using our data. Outcome regression showed the same ranks for chorioamnionitis and genital tract infection (GTI), while the effect estimate of intra-amniotic infection on PROM was not statistically significant by this method (OR 2.134, 95% CI 1 to 4.555). Three of 11 factors were assigned as associated factors by IPW but not by outcome regression. These included intra-amniotic infection and two variables: (1) pneumonia (OR 0.91, 95% CI 0.538 to 1.539); and (2) influenza (OR 0.957, 95% CI 0.863 to 1.061). Effects by outcome regression were mostly larger than those by IPW (eTable 13 in Appendix).

In the final association diagram (eFigure 13 in Appendix), infection- and immune-related conditions were seen: (1) influenza; (2) asthma; and (3) pneumonia. Maternal factors were also observed: (1) maternal age; (2) low socio-economic status; (3) multiple pregnancy; and (4) ante-partum hemorrhage. Both groups are shown colliding on periodontal disease in the final association diagram (eFigure 13 in Appendix), then collided with genital tract infection on intra-amniotic infection continuing to either chorioamnionitis or PROM.

#### Prognostic prediction of premature rupture of membranes

Predictive performances for classification task were already clearly described in the main text. In this Supplement, parameter estimates in each model is described. However, these were not always straightforward because of the complexity of several models.

For ridge regression (RR), the estimates were weights or beta values, as commonly described in a regression model (eTable 14 in R Markdown). For classification task, top three highest weights were assigned for chorioamnionitis, IAI, and GTI. This is similar to the effect ranks in association tests by IPW.

Three models used principal components (PCs), including PC elastic net regression (PC-ENR). To transform predictors into a PC, each predictor has a weight to multiply with. These weights were also shown (eTable 16 in R Markdown). For the PCs themselves, the parameter estimates in PC-ENR were similar to those in RR. The weights in PC-ENR were also shown (eTable 16 in R Markdown).

For PC random forest (PC-RF) and PC gradient boosting machine (PC-GBM), the parameter estimates may be represented by variable importance. It is a proportion of learners (trees) that include a predictor. These numbers were also shown (eTables 17 and 18 in R Markdown).

For DI-VNN, we filtered predictors by differential analysis which consisted of multiple univariable linear regressions. The parameter estimates were expressed as log of fold changes. These were equivalent to log of odds ratios. This number and others, including false discovery rate (FDR), were also shown for the selected predictors by FDR <0.05 (eTable 19 in R Markdown).

Parameter estimates of the DI-VNN were extremely enormous. To get similar sense with those of the other models, we used an intermediate output at a layer after being fed to Inception v4 for each ontology. This showed a learning representation by DI-VNN on a predictor. How these were computed had been described in a protocol we followed for these procedures.^1^ The intermediate outputs were shown (eTable 20 in R Markdown). We would point out several meanings of these outputs after the next section. In addition, connections between ontologies under the root are also shown (eTable 21 in R Markdown).

For comparison of our models with those of previous studies, we applied methods in the preferred reporting items for systematic reviews and meta-analyses (PRISMA) 2020 expanded checklist (eTable 4 in Appendix). From three literature databases and several steps, we found two prediction models as described in the main text (eTable 5 in Appendix). The steps are shown (eFigure 14 in Appendix).

#### Estimation of the time of delivery

Estimation performances were already clearly described in the main text. Parameter estimates of the estimation models were also shown in the same eTables with those of the classification models (eTable 14 in R Markdown). For estimation task, which implicitly related to preterm delivery, the lowest (negative) weights in the RR were also assigned to chorioamnionitis, IAI, and GTI, but, the second lowest rank was shifted up by multiple pregnancy. Lower-rank weight means earlier time of delivery, which is, likely having more chance to be a preterm delivery. It turns out our decision to choose IAI and chorioamnionitis as the associated factors, instead of those being affected of PROM, is consistent with these findings.

Although the RR mostly had achieved highest proportion of criteria fulfilled among the models by external validation sets, we noticed this only applies for the predicted non-events based on visual assessment. The associated factors may be well-generalized to estimate the time of delivery under non-events predicted (modeled) by DI-VNN. Nevertheless, we selected PC-RF as the best model for estimation task of the time of delivery.

Challenge on estimation task is reasonable considering different distributions among internal and external validation sets. In internal validation set, of which we only utilized the calibration set for model evaluation, predicted events by DI-VNN happened 15 weeks (95% CI 11 to 18; *n*=760) on average from the time of prediction. This was similar to those in external random (18, 95% CI 14 to 21; *n*=973) and temporal splits (15, 95% CI 12 to 19; *n*=687), but later than those in external geographical (9, 95% CI 6 to 13; *n*=500) and geotemporal splits (3, 95% CI 1 to 5; *n*=157). Meanwhile, the pregnancies predicted as non-events by DI-VNN ended at 40 weeks (95% CI 39 to 41; *n*=20,746) on average from time of prediction. This was earlier than those in external random (42, 95% CI 40 to 43; *n*=25,959), temporal (52, 95% CI 50 to 53; *n*=17,533), and geotemporal splits (60, 95% CI 54 to 65; *n*=2,067), but later than that in external geographical split (37, 95% CI 35 to 38; *n*=15,318).

#### Exploring deep-insight visible neural network

Population-level data exploration is described in this Supplement. For interactive figure and table of DI-VNN, we provide these in our web application (<https://predme.app/promtime>). Technical details for exploring DI-VNN were already described in a protocol we followed for these procedures.^1^

The network architecture of the DI-VNN is data-driven (eFigure 15b in Appendix). Using the clique-extracted ontology (CliXO)^74^ algorithm, we constructed an architecture for the convolutional neural network (CNN) using only a training set (see Methods). We can consider this algorithm as an agglomerative hierarchical clustering algorithm, but there could be more than one parent-child connection. A child ontology array has a similar value distribution with the parent one is because a neural network applies a backpropagation algorithm that updates the model parameters consecutively from the surface to the deeper layer following the path, which is the ontology network of DI-VNN. But, unlike other neural network models, DI-VNN isolates the backpropagation effect; therefore, we can trace the array values to interpret the possible meaning.

We explored each node of the DI-VNN at the population level. The diagnosis/procedure codes constructing the feature members were ICD-10 codes. We began from the most visually distinguished array (eFigure 15b in Appendix) which was ONT:171. These were N760 (acute vaginitis) and B379 (unspecified candidiasis). A positive output does not necessarily refer to an event. Nevertheless, positive and negative (color-coded) outputs tend to contribute to opposite outcomes, which are interpreted based on external contextual knowledge. Another distinguished array, ONT:144, was connected to the same node as that of the previous array. The feature member (9059, other microscopic examination of blood) tended to contribute to the same outcome as that of “unspecified candidiasis”. Unlike “acute vaginitis”, which is a local infection, both B379 and 9059 may be related to systemic infections. To this point, we gained insights into describing coincidences between systemic vs. local infection and PROM. Another array, ONT:154, was also visually distinguished. Although the value of the feature member assoc_A03 (chorioamnionitis) was zero, it was next to higher values that supported the same outcome as that of “acute vaginitis”. We traced the node on the upper layer to ONT:171. This array had a similar value distribution on the same channel ($z$=2) to that of ONT:154.

The feature position within any ontology array was determined using *t*-SNE with the Barnes-Hut approximation (see Methods).^7^ It mapped features on high dimensional to lower dimensional space, as multiple localities. This algorithm spreads small clusters instead of making a large bubble of clusters; thus, we expected our CNN algorithm can be better extracting predictive features from these localities. If a feature nearer to one than another, this means there is a closer relationship between both features. The localities clustered by *t*-SNE are grouped together at the root node on the most superficial layer (eFigure 15b in Appendix). Deeper layers have different subsets of features separated by the ontology grouping. By understanding how this algorithm works, we assumed assoc_A03 (ONT:154), i.e., “chorioamnionitis”, was closer to “acute vaginitis” than was “unspecified candidiasis” since both were in the same channel, the positions of the first and second dimensions were adjacent, and *t*-SNE preserved the neighborhood identity. Acute vaginitis is semantically a genital tract infection (GTI). By external contextual knowledge inferred from systematic human learning and confirmed by IPW using our data, GTI and chorioamnionitis were determined to be associated factors of PROM.

A node on more superficial layer, which is ONT:155, consisted a feature that prefers the same outcome that N760 (acute vaginitis) and assoc_A03 (chorioamnionitis) prefer, which is 598 (urethral catheterization). This feature is semantically related to acute vaginitis because the anatomical sites are adjacent. But, since ONT:155 is on a more superficial layer, this node will connect to the same node with many features from other ontology terms. This means more factors may need to interact with urethral catheterization (598) to be predictive for PROM. In addition, within the same ontology term, there is also 8602 (injection or tattooing of skin lesion or defect). Apparently, the CliXO algorithm have clustered these features together semantically, which are similarly invasive procedures. In addition, local infection from acute vaginitis may be related to chorioamnionitis, such that prefers the same outcome, as opposed to the possible systemic infection by the unspecified candidiasis.

Exploring this DI-VNN should be done with caution, since we developed the model using medical histories, which are diagnosis or procedure codes provided by medical doctors. We may or may not be modeling a human pathophysiology, but, we are definitely modeling the doctors’ behaviors of coding a diagnosis or procedure.^75^ By providing an interface to the internal properties of this model, a human user can assess each prediction case-by-case. From ONT:167 and ONT:149 on the deeper layer, we can find unusual features in the context of PROM, which are H527 (unspecified disorder of refraction) preferring non-events while 734 (flat foot), H521 (myopia), and H522 (astigmatism) preferring events. These codes might be responses to the subject symptoms of edema in the feet and blurry vision. Both symptoms in a pregnant woman may be typically associated with severe preeclampsia. But, a doctor may avoid this association if the context does not support the symptoms, e.g. symptoms by a non-pregnant subject. This may lead a doctor to assign these codes responding to those symptoms. For each prediction, a human user may need to explore the model to avoid misclassification by ignoring the prediction if it counters the clinical reasoning. More pragmatically at individual level, the predictive value may not be sufficient or the estimation may not be quite precise based on the corresponding subpopulation data with respectively the same predicted outcome or estimated time of delivery. In addition, albeit all of the population-level patterns from this exploration, every subject may reveal a different pattern using the same DI-VNN model, as described in the next section.

#### Web application

Briefly, we uploaded a record of 20 visits by a 19-year-old female subject from December 2, 2015 to July 30, 2016, consisting of 28 code entries. After determining the prediction date, which was set to July 30, 2016, we ran the application in 5.14 minutes (95% CI 5.11 to 5.18 minutes when we repeated it 10 times). We downloaded the report after the application was completed (eFigure 16 in Appendix). The predicted outcome was PROM, and the estimated time of delivery was 11 weeks after the time of prediction. The predicted probability was 0.867.

Furthermore, after the application was done (eFigure 16 in Appendix), we tried to adjust the threshold to the maximum value, such that the data for population-level performances are still available and the predictive value is also maximized depending on the prediction result. The population-level data was the same with internal validation (calibration split; *n*=21,506) which was used to plot the ROC curve (Figure 2b in the main text). The threshold was 0.67 with positive predictive value of 0.809 (95% CI 0.798 to 0.82). The sensitivity was reasonably low (0.107, 95% CI 0.104 to 0.11) if using this threshold. But, from a standpoint of prediction at individual level, a precise estimation is important to determine whether a decision corresponding to this prediction can be made with a good confidence. By default, the threshold is set at an optimum value of 0.14 (sensitivity 0.494, 95% CI 0.489 to 0.5). Based on the predicted probability case-by-case, a user can decide the threshold to adjust at.

From the reported timeline (eFigure 6 in Appendix), this subject is shown predicted to deliver on October 18^th^ 2016, approximately. If the predicted outcome is not PROM, the shaded area would be red; otherwise, turquoise color is applied to the area as shown. It depicted population-level estimation of true time of delivery for subjects that were also estimated to deliver within 11 weeks and predicted as PROM by the same threshold. The population-level data was the same with internal validation (*n*=107,536) which was used to plot the PC-RF estimation window (Figure 3b in the main text). By population-level estimation, the time of delivery might be at the beginning up to the end of October. Using threshold at 0.67, this population-level estimation was shifted earlier for a week compared to that at 0.14. In addition, for illustration purpose, just like a real-world setting, say we know the gestational age was 22-23 weeks’ gestation based on last menstrual period and ultrasound examination. If this estimation model is precise for this case, the subject would deliver at 33-34 weeks’ gestation, which is a preterm PROM.

We also saw the medical history of this subject from the reported timeline (eFigure 16 in Appendix). Up to the date of prediction, the prediction model used these features: (1) A09 (diarrhea and gastroenteritis of presumed infectious origin); (2) J069 (unspecified acute upper respiratory infection); (3) K30 (dyspepsia); and (4) 8878 (diagnostic ultrasound of gravid uterus). On the timeline, these were ordered from the most positive (top) to the most negative (bottom) based on each output in the ontology array.

In the prediction model, any of these features were classified in the ontology arrays of ONT:158, ONT:169, ONT:176, and root, as depicted by the ontology network (eFigure 16 in Appendix). We also identified the deepest ontology that includes all features in the timeline, which is ONT:169, but the predicted outcome based on this ontology is not PROM using the same threshold with that of the root. A user also can see a predictive performance of any ontologies, just like AUROC of the root (0.714, 95% CI 0.712 to 0.716). It is computed for the pre-calibrated DI-VNN only, since the calibrated one only used the predicted probability that was the output convoluted from the root ontology array.

Negative values at population level tend to event, as described in the previous section. From the ontology array (eFigure 16 in Appendix), J069 (unspecified acute upper respiratory infection) tends to event in that array, as shown as mostly negative outputs in the timeline. This feature was also surrounded by more negative outputs. A09 (diarrhea and gastroenteritis of presumed infectious origin) also had a negative value, but, this feature and K30 (dyspepsia) were surrounded by more positive outputs in the ontology array. If we apply the same illustrative gestational age, these infectious diseases (J069 and A09) were diagnosed respectively ten and four weeks the start of pregnancy, as shown on the timeline. Root is the only ontology that predicted PROM and included all features in this subject. This is implied all of these features should be taken together for the prediction. In addition, one may question why J069 (unspecified acute upper respiratory infection) tends to event while influenza have the opposite effect (OR 0.995,95% CI 0.993 to 0.997; eTable 13 in Appendix). We found that influenza, which is assoc_A28, did not include J069 (eTable 12 in R Markdown). This implied specific acute upper respiratory infection, such influenza, may not have the same effect with that by the unspecified one on PROM.

Beyond the root, the array of ontology ONT:169 is also shown (eFigure 17 in Appendix). A09 and J069 had negative values. As described in the main text, these features tend to an event. Respectively, the surrounding positive and negative values were subtler in this ontology array. Since this filtered array is fed forward to convolutional layers to be reduced each time passing a layer (see video in the protocol^1^), the value of A09 and J069 were averaged along with the adjacent values toward zero, opposite to event. In turn, this coincides with lower AUROC in ONT:169 compared to that in root.

Eventually, a user may want to know if the PROM prediction and estimated time of delivery are similar to true values. One can save this model online and return later to the web application to enter the true outcome and time of delivery. In this way, a user can collect data for external validation purposes, specifically describing the model performances based on local data distributions. In our case, the true outcome was also PROM and the time of delivery was 12 weeks after the time of prediction, a week later than the predicted time of delivery.

### Discussion

From association tests and feature extraction to model selection and exploration, we only used an internal validation set. But, the model is evaluated using four external validation sets with a large sample size. This has found the DI-VNN prediction was robust and the PC-RF estimation was precise within a reasonable time window. All of these models used only a medical history of a patient, which is easily extracted from the electronic medical record systems of most healthcare providers worldwide. Neither a biomarker testing nor even a laboratory test is needed. Eventually, the best models are ready to use for any healthcare providers using an open-access web application without changing their electronic health record systems and revealing private data.

To gain insights into PROM antecedents, we showed how this was achieved by exploring the model at both the population (eFigure 15b in Appendix) and individual levels (eFigure 16 in Appendix). If we assume that PROM is a subclass of preeclampsia, this may explain why population-level exploration implied an insight of systemic vs. local infection by competing risks. A hematogenous infection may be associated with reproductive-tract microbial dysbiosis and can affect several pregnancy outcomes, including PROM, preeclampsia, and fetal growth restriction.^76^ Both hematogenous and ascending infections from reproductive tract are found in PROM.^33,77^ Hematogenous infections included those from digestive and respiratory organs. Similar to the population-level exploration (eFigure 15 in Appendix), we also found a possibility of hematogenous infection at the individual level (eFigure 16 in Appendix), and these were from infectious gastroenteritis and unspecified acute upper respiratory infection. Both population- and individual-level explorations also implied a period surrounding the beginning of pregnancy as the onset of the optimal time to predict PROM. Several features in the DI-VNN were counterintuitive, e.g., H527 (unspecified disorder of refraction), 734 (flat foot), H521 (myopia), and H522 (astigmatism). Yet, these describe blurry vision and swelling in the feet, which are also symptoms of preeclampsia. Similar eye-related codes, i.e., myopia and astigmatism, were also found to be important in predicting preeclampsia in an RF model.^78^

### Appendix

List of eFigures

List of eTables

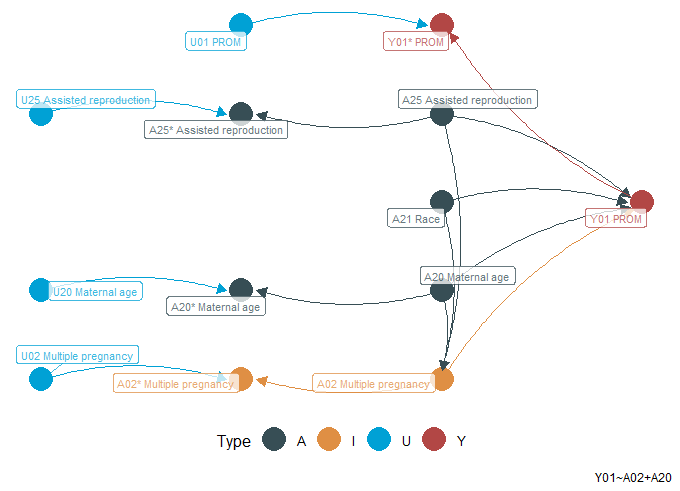

eFigure 1. Multiple pregnancy

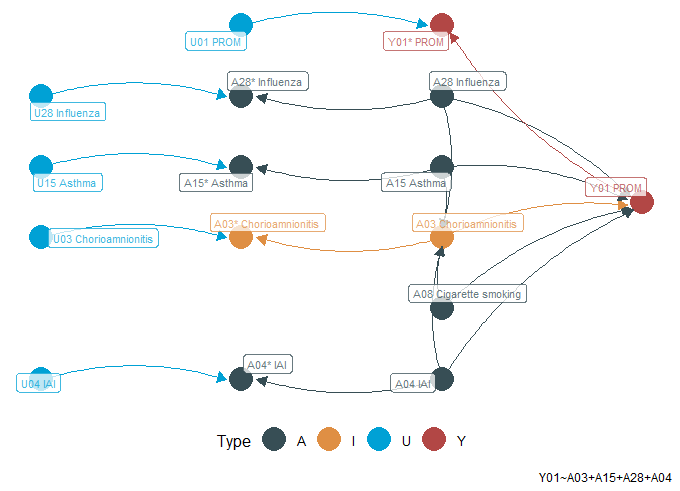

eFigure 2. Chorioamnionitis

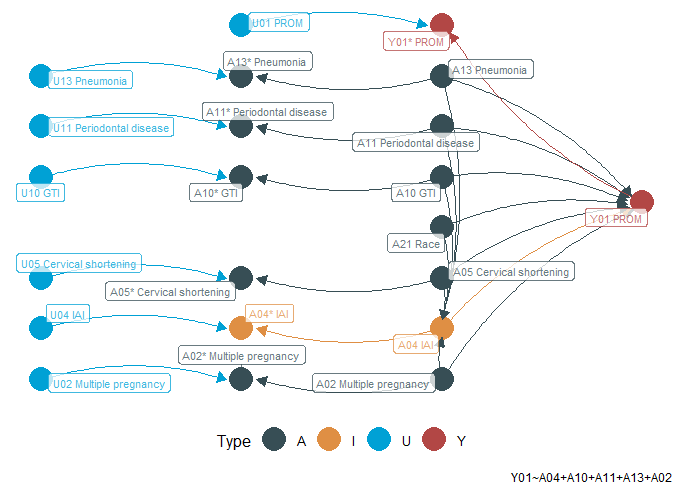

eFigure 3. Intra-amniotic infection (IAI)

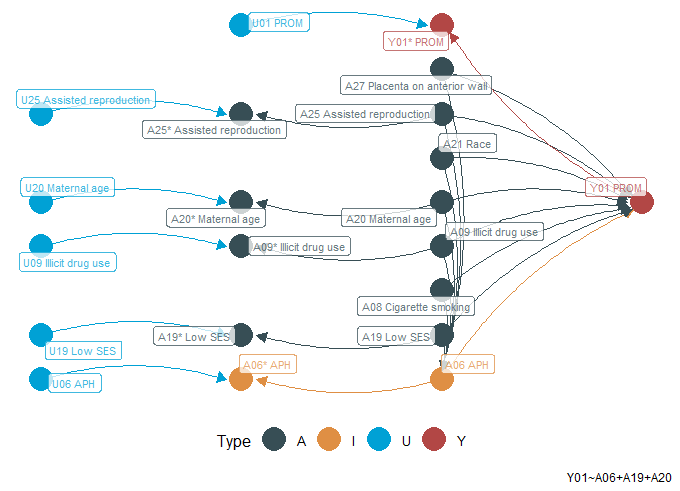

eFigure 4. Ante-partum hemorrhage (APH)

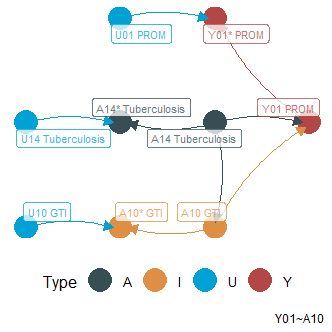

eFigure 5. Genital tract infection (GTI)

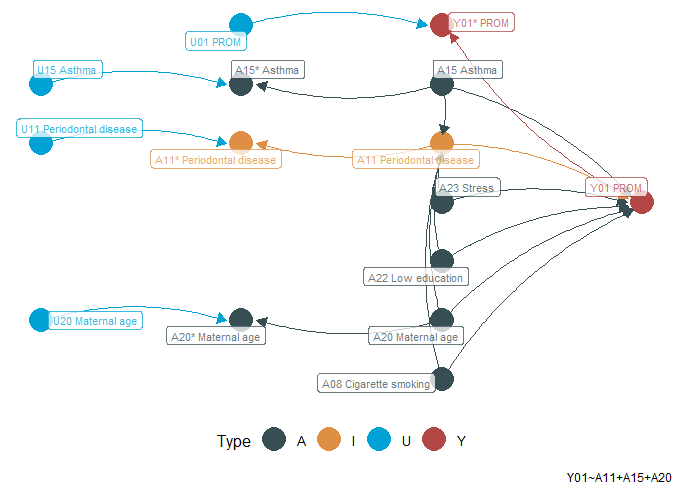

eFigure 6. Periodontal disease

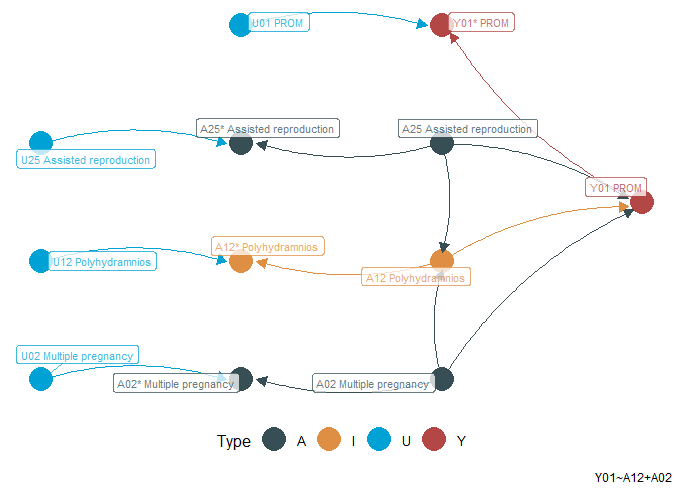

eFigure 7. Polyhydramnios

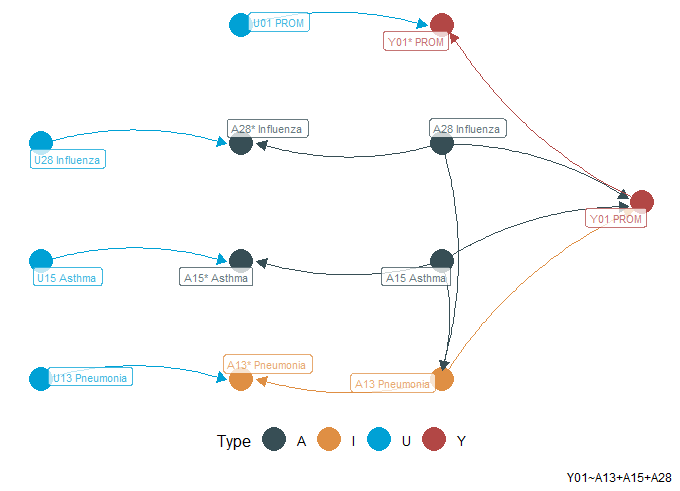

eFigure 8. Pneumonia

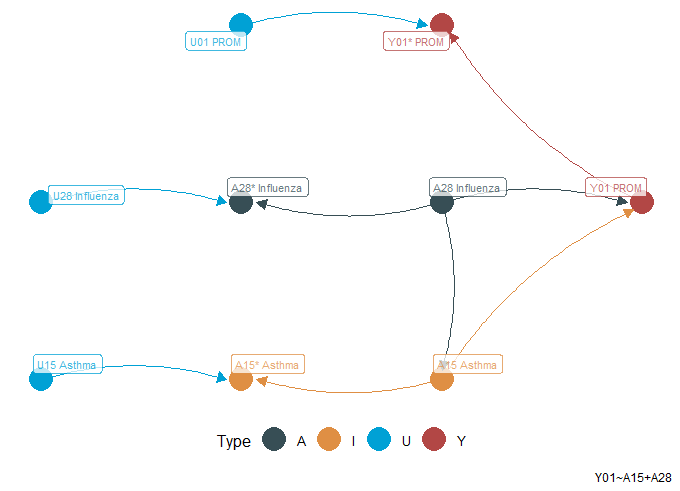

eFigure 9. Asthma

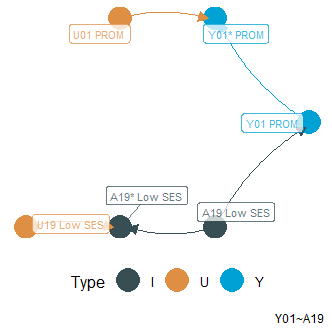

eFigure 10. Low socio-economic status (SES)

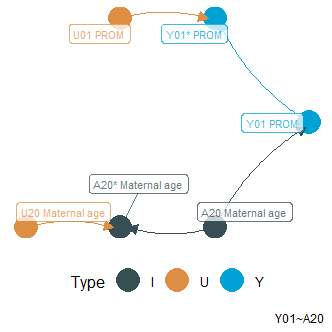

eFigure 11. Maternal age

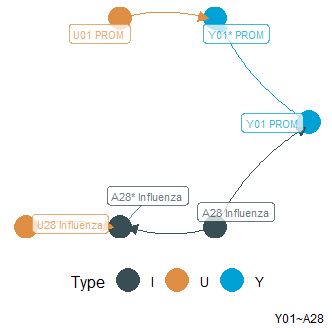

eFigure 12. Influenza

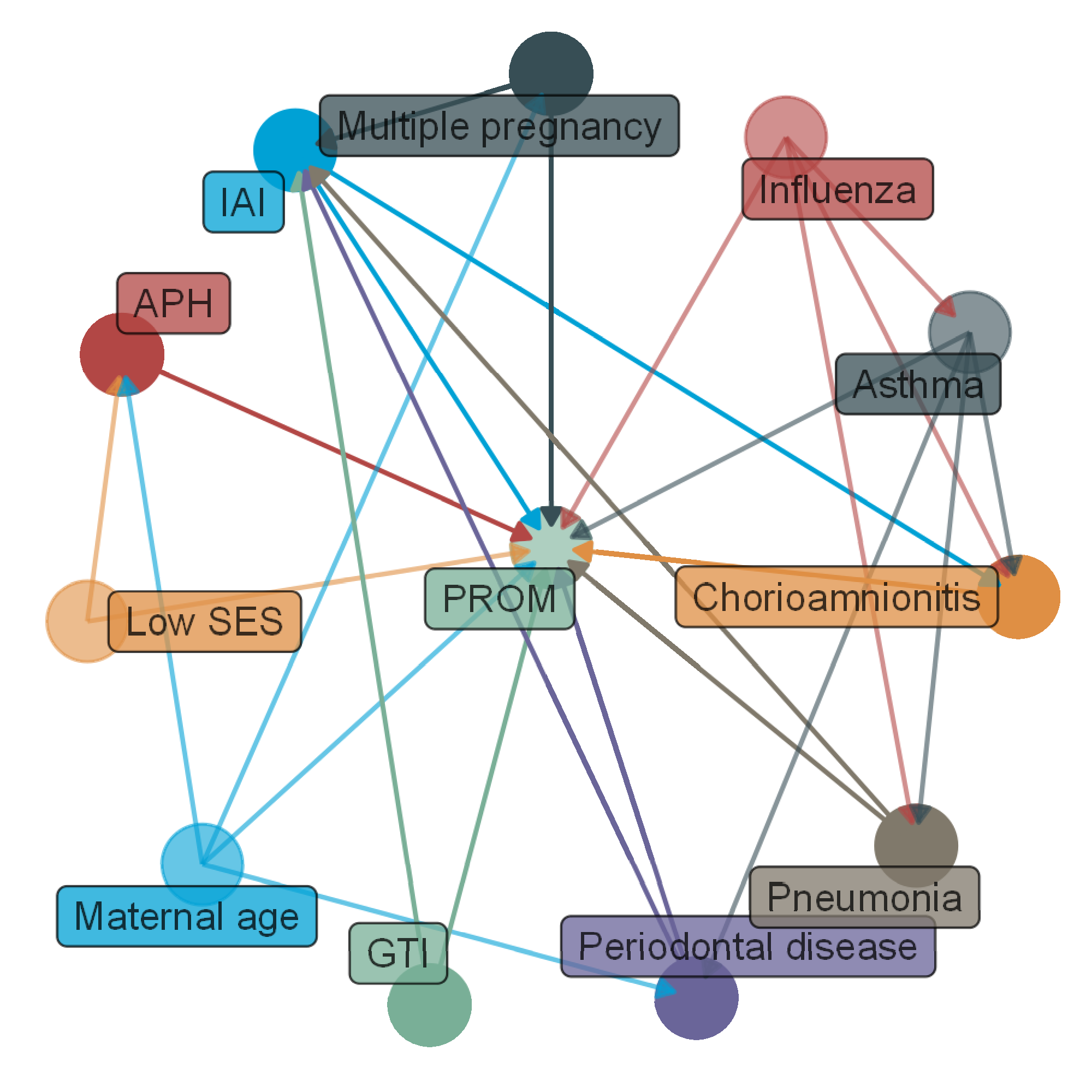

eFigure 13. Final association diagram. All associated factors were verified by inverse probability weighting (IPW) using our data. Association tests were only conducted between PROM and each of the associated factors. Inter-associated factor relationships were not verified, but this demonstrates how an associated factor is included in the associated model of another associated factor in this figure. APH, ante-partum hemorrhage; GTI, genital tract infection; IAI, intra-amniotic infection; PROM, prelabor rupture of membranes; SES, socio-economic status.

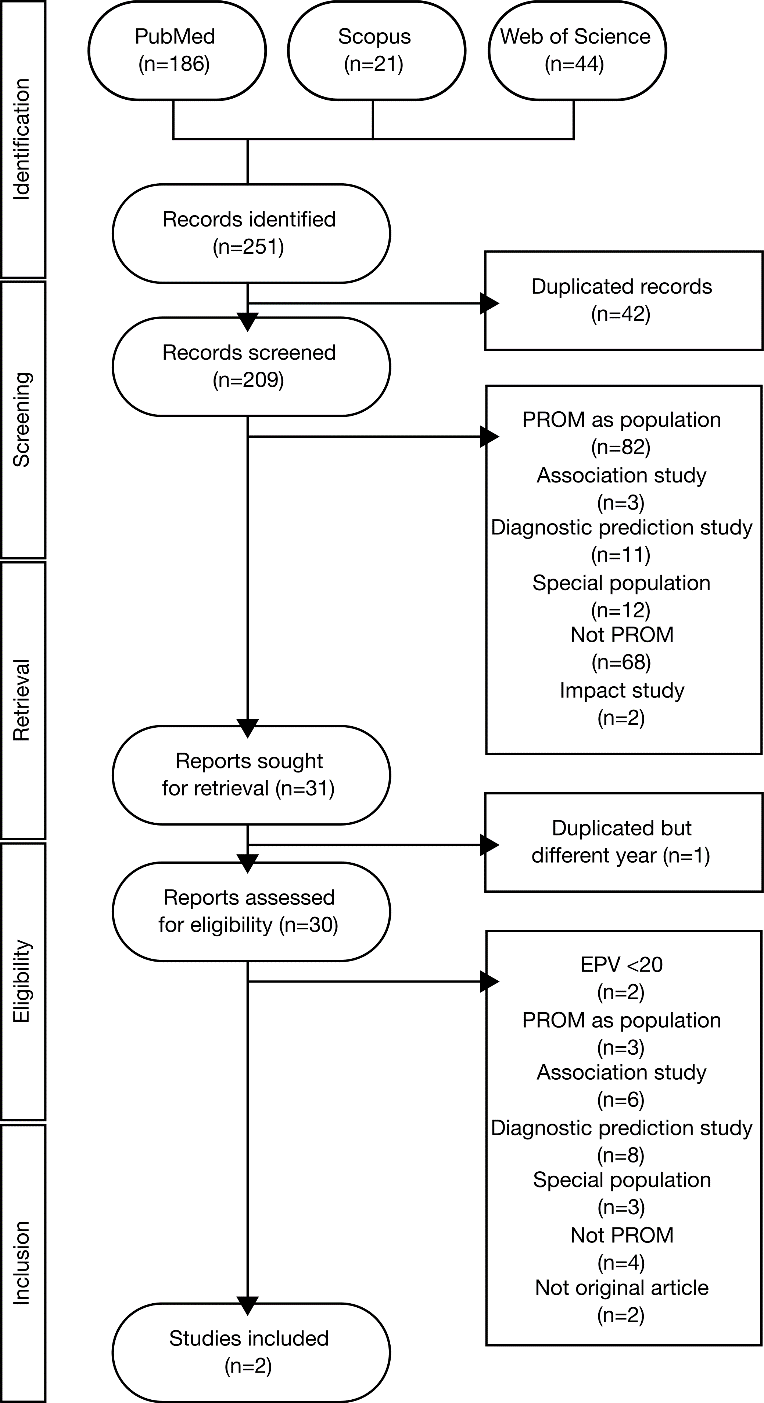

eFigure 14. Flow diagram to find comparable models from previous studies

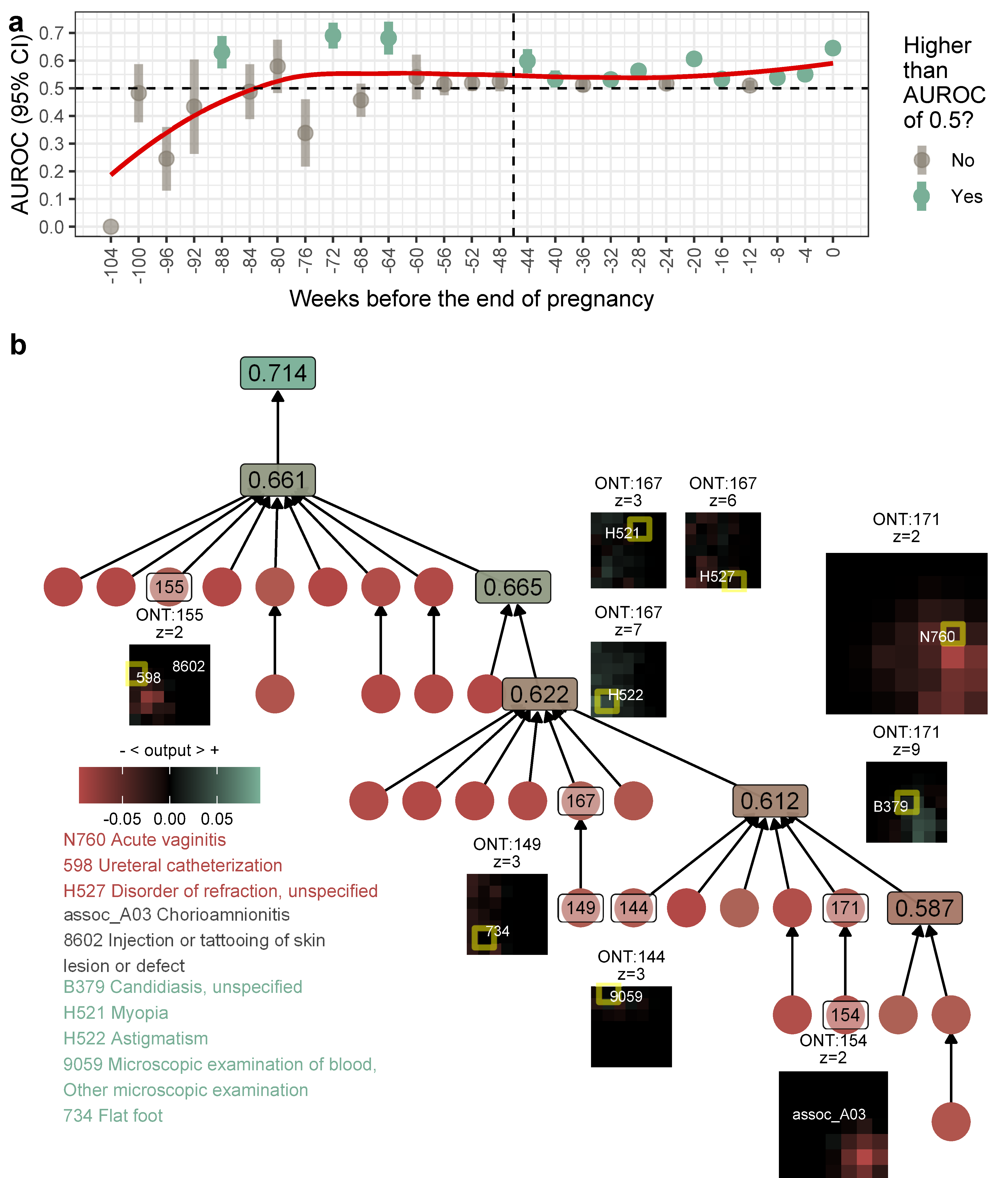

eFigure 15. Exploratory data analysis. (a) area under the receiver operating characteristic curve (AUROC) of DI-VNN every 4 weeks; (b) ontology (ONT) network and arrays of the DI-VNN. Showing the best time window for the prediction by the DI-VNN (a) and AUROCs of >0.55 for prediction using parts of the network architecture up to each layer on which a node resides (b). Each node is a CliXO term. We only show distinguished output arrays for a particular channel denoted by ‘z’. A feature in the array may tend to positive or negative output, color-coded based on the gradient as shown, including the feature description. Yellow squares in an array refer to a feature if only its output is non-zero. A feature may not have this square, e.g. assoc_A03 and 8602. ONT:154 is an example of a backpropagation effect from ONT:171. CliXO, clique-extracted ontology; DI-VNN, deep-insight visible neural network.

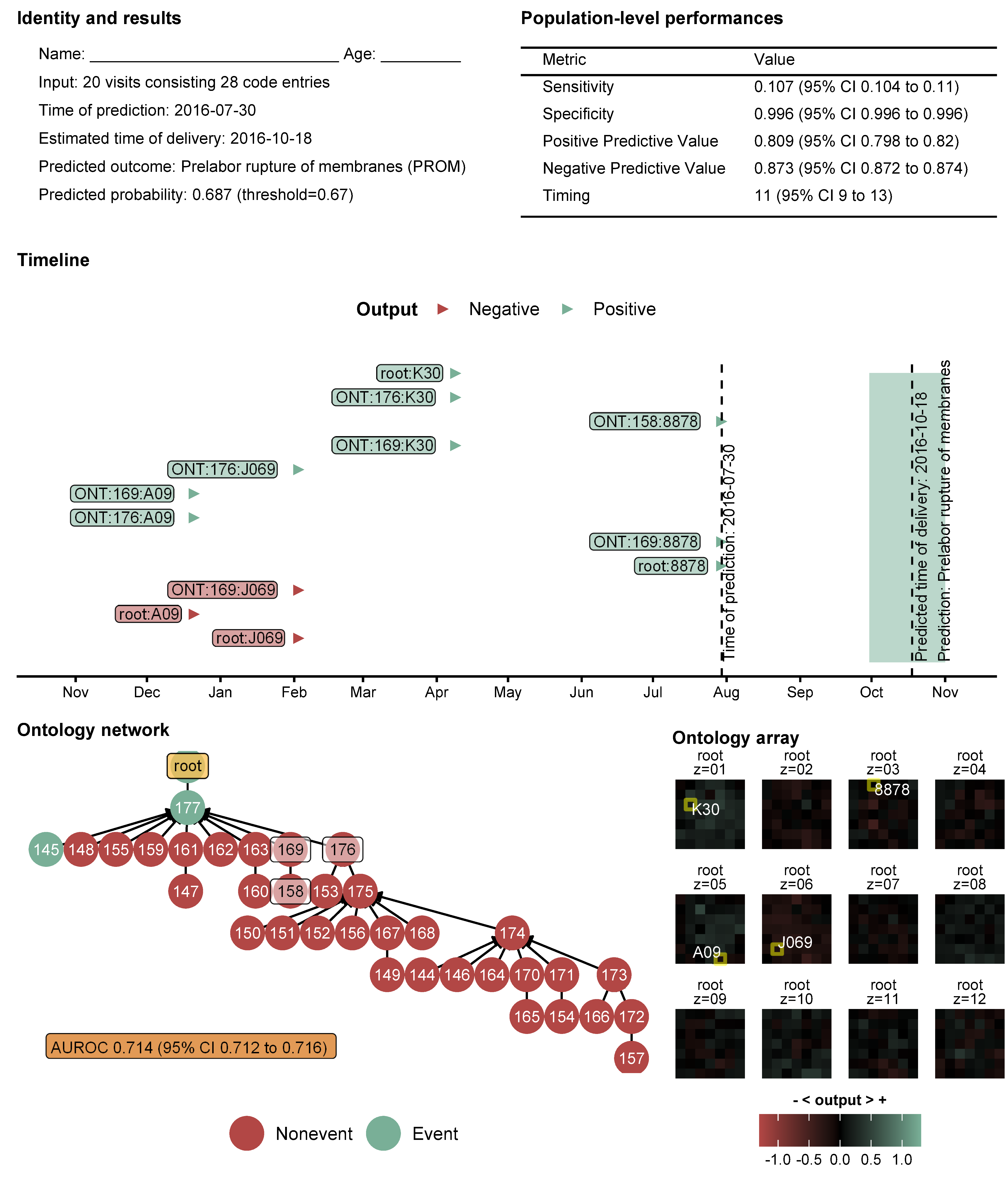

eFigure 16. A case example. This is an example for predicting prelabor rupture of membranes (PROM) by DI-VNN and estimating of the time of delivery by PC-RF. An ontology term in the timeline is prefixed by ONT, followed by the number and one of the feature members. 8878, diagnostic ultrasound of gravid uterus; A09, diarrhea and gastroenteritis of presumed infectious origin; AUROC, area under receiver operating characteristics curve; DI-VNN, deep-insight visible neural network; J069, unspecified acute upper respiratory infection; K30, dyspepsia; PC, principal component; RF, random forest.

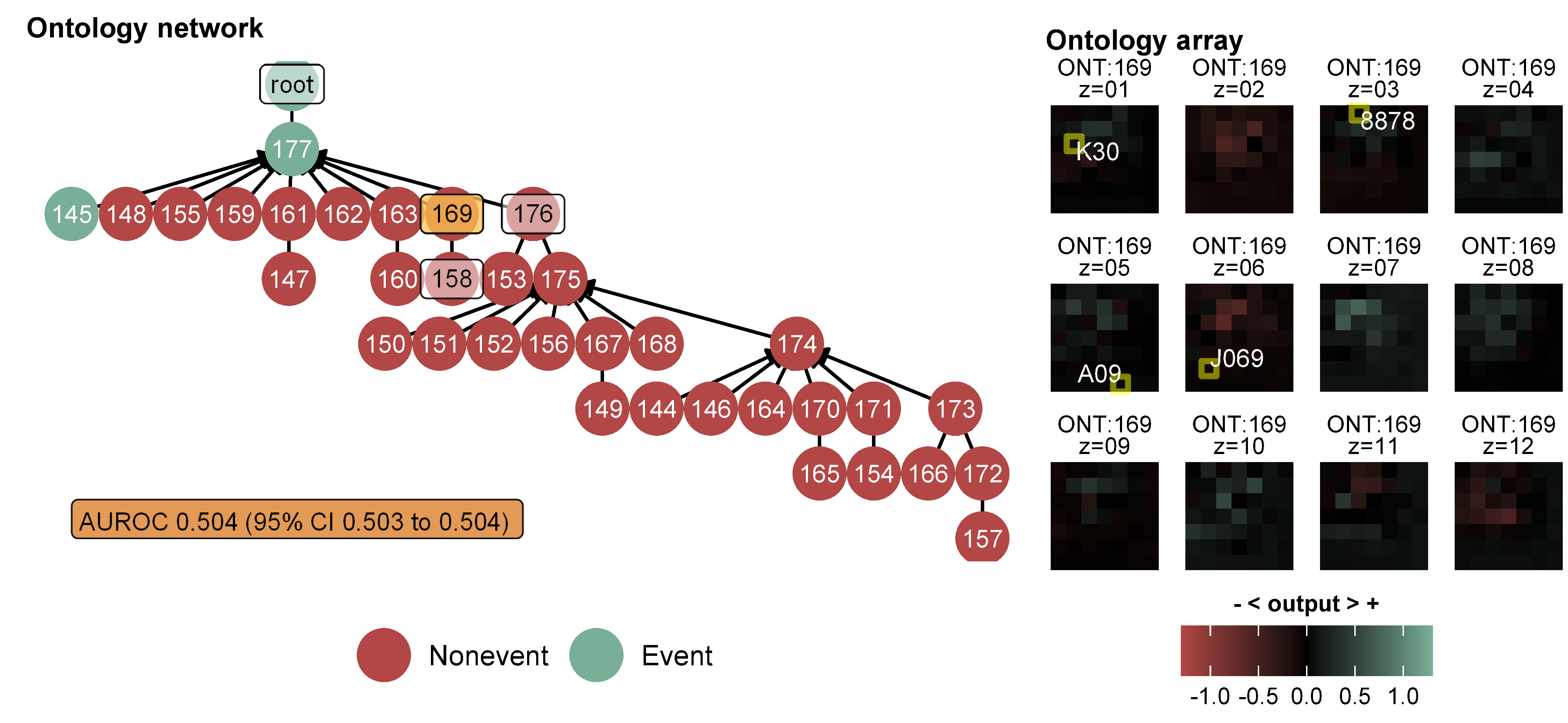

eFigure 17. Ontology ONT:169

eTable 1. TRIPOD checklist for prediction model development and validation

| **Section/ topic** | **Item** | | **Checklist item** | **Page** |
| --- | --- | --- | --- | --- |
| Title and abstract | | | | |
| Title | 1 | D;V | Identify the study as developing and/or validating a multivariable prediction model, the target population, and the outcome to be predicted. | 1 |
| Abstract | 2 | D;V | Provide a summary of objectives, study design, setting, participants, sample size, predictors, outcome, statistical analysis, results, and conclusions. | 3 |
| Introduction | | | | |
| Background and objectives | 3a | D;V | Explain the medical context (including whether diagnostic or prognostic) and rationale for developing or validating the multivariable prediction model, including references to existing models. | 4 |
|  | 3b | D;V | Specify the objectives, including whether the study describes the development or validation of the model or both. | 5 |
| Methods | | | | |
| Source of data | 4a | D;V | Describe the study design or source of data (e.g., randomized trial, cohort, or registry data), separately for the development and validation data sets, if applicable. | 5 |
|  | 4b | D;V | Specify the key study dates, including start of accrual; end of accrual; and, if applicable, end of follow-up. | 5 |
| Participants | 5a | D;V | Specify key elements of the study setting (e.g., primary care, secondary care, general population) including number and location of centers. | 5 |
|  | 5b | D;V | Describe eligibility criteria for participants. | 6 |
|  | 5c | D;V | Give details of treatments received, if relevant. | (not relevant) |
| Outcome | 6a | D;V | Clearly define the outcome that is predicted by the prediction model, including how and when assessed. | 6 |
|  | 6b | D;V | Report any actions to blind assessment of the outcome to be predicted. | 5 to 6 |
| Predictors | 7a | D;V | Clearly define all predictors used in developing or validating the multivariable prediction model, including how and when they were measured. | 6 |
|  | 7b | D;V | Report any actions to blind assessment of predictors for the outcome and other predictors. | 5 to 6 |
| Sample size | 8 | D;V | Explain how the study size was arrived at. | 5, 7 |
| Missing data | 9 | D;V | Describe how missing data were handled (e.g., complete-case analysis, single imputation, multiple imputation) with details of any imputation method. | 6 |
| Statistical analysis methods | 10a | D | Describe how predictors were handled in the analyses. | 7 |
|  | 10b | D | Specify type of model, all model-building procedures (including any predictor selection), and method for internal validation. | 7 |
|  | 10c | V | For validation, describe how the predictions were calculated. | 7 to 8 |
|  | 10d | D;V | Specify all measures used to assess model performance and, if relevant, to compare multiple models. | 8 |
|  | 10e | V | Describe any model updating (e.g., recalibration) arising from the validation, if done. | 7 |
| Risk groups | 11 | D;V | Provide details on how risk groups were created, if done. | 6 |
| Development vs. validation | 12 | V | For validation, identify any differences from the development data in setting, eligibility criteria, outcome, and predictors. | 7 |
| Results | | | | |
| Participants | 13a | D;V | Describe the flow of participants through the study, including the number of participants with and without the outcome and, if applicable, a summary of the follow-up time. A diagram may be helpful. | 9, 19 |
|  | 13b | D;V | Describe the characteristics of the participants (basic demographics, clinical features, available predictors), including the number of participants with missing data for predictors and outcome. | 9, 17 |
|  | 13c | V | For validation, show a comparison with the development data of the distribution of important variables (demographics, predictors and outcome). | 9, 17, 19 |
| Model development | 14a | D | Specify the number of participants and outcome events in each analysis. | 9, 19 |
|  | 14b | D | If done, report the unadjusted association between each candidate predictor and outcome. | 9, 17, 18 |
| Model specification | 15a | D | Present the full prediction model to allow predictions for individuals (i.e., all regression coefficients, and model intercept or baseline survival at a given time point). | 9 |
|  | 15b | D | Explain how to the use the prediction model. | 11 |
| Model performance | 16 | D;V | Report performance measures (with CIs) for the prediction model. | 9 to 11, 20 to 21 |
| Model-updating | 17 | V | If done, report the results from any model updating (i.e., model specification, model performance). | 9 to 10, 20 |
| Discussion | | | | |
| Limitations | 18 | D;V | Discuss any limitations of the study (such as nonrepresentative sample, few events per predictor, missing data). | 12 |
| Interpretation | 19a | V | For validation, discuss the results with reference to performance in the development data, and any other validation data. | 11 to 12 |
|  | 19b | D;V | Give an overall interpretation of the results, considering objectives, limitations, results from similar studies, and other relevant evidence. | 11 to 12 |
| Implications | 20 | D;V | Discuss the potential clinical use of the model and implications for future research. | 12 |
| Other information | | | | |
| Supplementary information | 21 | D;V | Provide information about the availability of supplementary resources, such as study protocol, Web calculator, and data sets. | 5, 13 to 14 |
| Funding | 22 | D;V | Give the source of funding and the role of the funders for the present study. | 13 |

eTable 2. Guidelines for developing and reporting machine learning predictive models in biomedical research and prediction model risk of bias assessment tools (PROBAST)

| Item number | Section | Topic | Checklist item with the description and/or the corresponding text | | |
| --- | --- | --- | --- | --- | --- |
| 1 | Title | Nature of study | ✓ |  | Identify the report as introducing a predictive model |
|  |  |  |  | • | Prognostication for prelabor rupture of membranes and the time of delivery in nationwide insured women: development, validation, and deployment |
| 2 | Abstract | Structured summary | ✓ |  | Background |
|  |  |  |  | • | Prognostic predictions of prelabor rupture of membranes (PROM) lack proper sample sizes and external validation. |
|  |  |  | ✓ |  | Objectives |
|  |  |  |  | • | To develop, validate, and deploy statistical and/or machine learning prediction models using medical histories for prelabor rupture of membranes and the time of delivery. |
|  |  |  | ✓ |  | Data sources |
|  |  |  |  | • | A retrospective cohort design within 2-year period (2015 to 2016) of a single-payer, government-owned health insurance database covering 75.8% individuals in a country |
|  |  |  |  | • | Nationwide healthcare providers (*n*=22,024) at primary, secondary, and tertiary levels |
|  |  |  |  | • | 12-to-55-year-old women that visit healthcare providers using the insurance from ~1% random sample of insurance holders stratified by healthcare provider and category of family: (1) never visit; (2) visit only primary care; and (3) visit all levels of care |
|  |  |  | ✓ |  | Performance metrics of the predictive model or models, in both point estimates and confidence intervals |
|  |  |  |  | • | The best prognostication achieved area under curve 0.73 (0.72 to 0.75), sensitivity 0.494 (0.489 to 0.500), specificity 0.816 (0.814 to 0.818), and likelihood ratio being positive 2.68 (2.63 to 2.75) and negative 0.62 (0.61 to 0.63). |
|  |  |  |  | • | Meanwhile, the best estimation achieved ± 2.2 and 2.6 weeks respectively for predicted events and non-events. Our web application only took 5.14 minutes (5.11 to 5.18) per prediction. |
|  |  |  | ✓ |  | Conclusion including the practical value of the developed predictive model or models |
|  |  |  |  | • | Prelabor rupture of membranes and the time of delivery were predicted by medical histories; but, an impact study is required before clinical application. |
| 3 | Introduction | Rationale | ✓ |  | Identify the clinical goal |
|  |  |  |  | • | Preterm prelabor rupture of membranes (PROM) is widely used as an inclusion criterion for predictions of other conditions. The disease precedes 40%~50% of all preterm deliveries and arises from multiple disease pathways. Yet, the antecedent remains unclear, and prognostic predictions lack proper sample sizes and external validation. |
|  |  |  |  | • | Predicting this disease, estimating the time of delivery, and tracing possible root causes would enable development of preventive strategies and improve efficiency of conducting a prospective cohort study of associated complications. |
|  |  |  | ✓ |  | Review the current practice and prediction accuracy of any existing models |
|  |  |  |  | • | Prediction of PROM is mostly diagnostic. For prognostications, a model of preterm PROM was recently developed using maternal factors during the first trimester. Based on a training set (*n*=10,280), the area under receiver operating characteristic (ROC) curve (AUROC) was 0.667. Predictions of all-cause spontaneous preterm deliveries are also poor (AUROCs 0.54 to 0.70; <37 weeks’ gestation; *n*=118/2540), and they are exposed to high risks of bias based on a systematic review. However, there has been no development or external validation of a prognostic prediction model for PROM. In addition, because it is reasonably challenging, no studies have developed a model to estimate the time of delivery before the day. |
|  |  | Objectives | ✓ |  | State the nature of study being predictive modelling, defining the target of prediction |
|  |  |  |  | • | This study aimed to develop, validate, and deploy a prognostic prediction model for PROM and an estimator of the time of delivery using a nationwide health insurance database. |
|  |  |  | ✓ |  | Identify how the prediction problem may benefit the clinical goal |
|  |  |  |  | • | Nonetheless, solving PROM problems requires predictions and causal modeling to develop better preventive strategies at both the population and individual levels. To address this issue, we applied both human and machine learning tools. Using only medical histories, the model deployment could be accessible worldwide via a web application. |
| 5 | Methods | Describe the setting | ✓ |  | Identify the clinical setting for the target predictive model |
|  |  |  |  | ❖ | Clinical setting: primary, secondary, and tertiary care |
|  |  |  |  | • | This dataset was the second version published on August 2019 (access approval no.: 5064/I.2/0421) covering ~1% (*n*=1,697,452) of insurance holders in 2015 and 2016 from nationwide, affiliated healthcare providers (*n*=22,024; primary, secondary, and tertiary care). |
|  |  |  | ✓ |  | Identify the modelling context in terms of facility type, size, volume, and duration of available data |
|  |  |  |  | ❖ | Facility type: primary care and hospital |
|  |  |  |  | ❖ | Size: 1,697,452 insurance holders |
|  |  |  |  | ❖ | Volume: 22,024 healthcare providers |
|  |  |  |  | ❖ | Duration: 2 years (2015 and 2016) |
|  |  |  |  | • | This dataset was the second version published on August 2019 (access approval no.: 5064/I.2/0421) covering ~1% (*n*=1,697,452) of insurance holders in 2015 and 2016 from nationwide, affiliated healthcare providers (*n*=22,024; primary, secondary, and tertiary care). |
| 6 | Methods | Define the prediction problem | ✓ |  | Define a measurement for the prediction goal (per patient or per hospitalization or per type of outcome) |
|  |  |  |  | ❖ | Prediction goal: pregnancy outcome (PROM or not) and the time of delivery per visit (or hospitalization) |
|  |  |  |  | • | We developed prediction models to classify if a visit was made by a subject for which the pregnancy period ended with PROM, and to estimate the time of delivery. |
|  |  |  | ✓ |  | Determine that the study is retrospective or prospective |
|  |  |  |  | ❖ | Study design: retrospective study |
|  |  |  |  | • | We applied a retrospective design to select subjects from a nationwide health insurance dataset provided by a government-owned health insurance company in Indonesia. |
|  |  |  | ✓ |  | Identify the problem to be prognostic or diagnostic |
|  |  |  |  | ❖ | Prediction problem: prognostic |
|  |  |  |  | • | The outcome for the classification task was an event for a subject that had encountered the O42 code, which is the International Classification of Disease version 10 (ICD-10) code for PROM. Otherwise, a subject was assigned as a nonevent if the pregnancy ended within the dataset period using the same codes for pregnancy termination. |
|  |  |  | ✓ |  | Determine the form of the prediction model: (1) classification if the target variable is categorical, (2) regression if the target variable is continuous, (3) survival prediction if the target variable is the time to an event. |
|  |  |  |  | ❖ | Prediction model form: classification (PROM or not) and regression (a number of days) |
|  |  |  |  | • | The outcome for the classification task was an event for a subject that had encountered the O42 code, which is the International Classification of Disease version 10 (ICD-10) code for PROM. Otherwise, a subject was assigned as a nonevent if the pregnancy ended within the dataset period using the same codes for pregnancy termination. |
|  |  |  |  | • | Meanwhile, the outcome for the estimation task was the number of days from the latest visit encountering of the outcome code to a visit when the prediction model was used. |
|  |  |  | – |  | Translate survival prediction into a regression problem, with the target measured over a temporal window following the time of prediction. |
|  |  |  |  | ❖ | Not applicable because we did not use survival rate for target variable. |
|  |  |  | ✓ |  | Explain practical costs of prediction errors (e.g., implications of under-diagnosis or over-diagnosis) |
|  |  |  |  | ❖ | Under-prognosis: unavailable resources for the premature baby |
|  |  |  |  | ❖ | Over-prognosis: unnecessary intervention and test |
|  |  |  |  | • | (Supplement) Practical costs of prediction errors were considered when evaluating the models. Under-prognosis may cause pregnancy monitoring to be off-guard. A pregnant woman with a preterm delivery may not reside in an area with a readily available NICU, particularly in low-resource settings. Over-prognosis may lead to unnecessary enrollment of patients into a cohort study in an early intervention for preventing PROM or complications, e.g., an antibiotic administration. This may also lead to unnecessary tests for more-specific prognostications. |
|  |  |  | ✓ |  | Defining quality metrics for prediction models |
|  |  |  |  | ❖ | Quality metrics: well-calibrated by internal validation for the classification model, maximum interval of ± *x* weeks when predicting > *x* weeks on each predicted outcome for the regression model |
|  |  |  |  | • | Calibration (plot, intercept, and slope) and discrimination (i.e. AUROC) were computed for classification tasks. We also computed sensitivity, specificity, positive likelihood ratio (LR+), and negative likelihood ratio (LR-). |
|  |  |  |  | • | For estimation, we computed a proportion of weeks in which each predicted time, in weeks, was included within an interval estimate of the true one. The interval had to be the maximum ± *x* weeks when predicting > *x* weeks, e.g., for any women predicted to deliver in 6 weeks, this number should fall into the true time of delivery within ± 6 weeks. |
|  |  |  | ✓ |  | Define the success criteria for prediction (e.g., based on metrics in internal validation or external validation in the context of the clinical problem) |
|  |  |  |  | ❖ | Success criteria: The AUROC is higher or not lower than recent models for preterm PROM, respectively if using predictors in low-resource or high-resource setting |
|  |  |  |  | • | Success criteria of the modeling were an AUROC greater than those of recent models (last 5 years) of PROM prognostic predictions using simple predictors (e.g., maternal factors), or greater or equal to those using high-resource predictors (e.g., biophysical or biochemical markers). |
| 7 | Methods | Prepare data for model building | ✓ |  | Identify relevant data sources and quote the ethics approval number for data access |
|  |  |  |  | ❖ | Data source: a nationwide health insurance dataset |
|  |  |  |  | ❖ | Ethic waiver number: N202106025 |
|  |  |  |  | ❖ | Data access approval number: 5064/I.2/0421 |
|  |  |  |  | • | Ethical clearance was waived by the Taipei Medical University Joint Institutional Review Board (TMU-JIRB number: N202106025). |
|  |  |  |  | • | We applied a retrospective design to select subjects from a nationwide health insurance dataset provided by a government-owned health insurance company in Indonesia. |
|  |  |  |  | • | This dataset was the second version published on August 2019 (access approval no.: 5064/I.2/0421) covering ~1% (*n*=1,697,452) of insurance holders in 2015 and 2016 from nationwide, affiliated healthcare providers (*n*=22,024; primary, secondary, and tertiary care). |
|  |  |  | ✓ |  | State the inclusion and exclusion criteria for data |
|  |  |  |  | ❖ | Inclusion criteria: the health insurance holders of 12-to-55-years-old females who had ever visit the affiliated healthcare providers within the dataset period |
|  |  |  |  | ❖ | Exclusion criteria: visits after delivery |
|  |  |  |  | • | We included health insurance holders of 12~55-year-old females who had visited affiliated healthcare providers. We excluded visits after delivery. For a person who was pregnant twice within the period, we labeled the same person as a different subject for each pregnancy period. A complete list of codes for determining delivery or immediately after delivery care is available in eTable 7 of the Supplement. |
|  |  |  | ✓ |  | Describe the time span of data and the sample or cohort size |
|  |  |  |  | ❖ | Time span of data: 2 years (2015 and 2016); from pre-pregnancy period to the end of pregnancy |
|  |  |  |  | ❖ | Sample size: internal pre-calibration split (*n*=86,030); internal calibration split (*n*=21,506); external random split (*n*=26,932); external geographical split (*n*=15,818); external temporal split (*n*=18,220); external geotemporal split (*n*=2,224) |
|  |  |  |  | • | Figure 1. Subject selection by applying a retrospective design and data partitioning for internal and external validations. |
|  |  |  |  | • | This dataset was the second version published on August 2019 (access approval no.: 5064/I.2/0421) covering ~1% (*n*=1,697,452) of insurance holders in 2015 and 2016 from nationwide, affiliated healthcare providers (*n*=22,024; primary, secondary, and tertiary care). |
|  |  |  | ✓ |  | Define the observational units on which the response variable and predictor variables are defined |
|  |  |  |  | ❖ | Response variable for classification task: event (PROM) and non-event (not PROM until the end of pregnancy) |
|  |  |  |  | ❖ | Response variable for estimation (regression) task: a number of days |
|  |  |  |  | ❖ | Predictors: historical rates (Kaplan-Meier estimates) |
|  |  |  |  | • | The outcome for the classification task was an event for a subject that had encountered the O42 code, which is the International Classification of Disease version 10 (ICD-10) code for PROM. Otherwise, a subject was assigned as a nonevent if the pregnancy ended within the dataset period using the same codes for pregnancy termination. |
|  |  |  |  | • | Meanwhile, the outcome for the estimation task was the number of days from the latest visit encountering of the outcome code to a visit when the prediction model was used. |
|  |  |  |  | • | Candidate predictors consisted of medical histories defined by one or more codes of diagnosis and procedure. |
|  |  |  | ✓ |  | Define the predictor variables. Extra caution is needed to prevent information leakage from the response variable to predictor variables. |
|  |  |  |  | ❖ | Medical histories: all of the ICD-10 codes for either diagnoses or procedures, except codes that demonstrate delivery or immediately after delivery care, including a code for the events |
|  |  |  |  | • | Candidate predictors consisted of medical histories defined by one or more codes of diagnosis and procedure. |
|  |  |  | ✓ |  | Describe the data pre-processing performed, including data cleaning and transformation. Remove outliers with impossible or extreme responses; state any criteria used for outlier removal. |
|  |  |  |  | ❖ | Data cleaning: assign censoring labels, remove codes that can leak the outcome, remove perfect-separation candidate predictors |
|  |  |  |  | ❖ | Data transformation: transform the candidate predictors into historical rates (KM estimates), transform the historical rates into PCs, transform the historical rates into -1, 0, or 1 by 1-bit stochastic gradient descent using the averages in differential analysis |
|  |  |  |  | ❖ | Outlier removal: not applicable because there was no continuous candidate predictor |
|  |  |  |  | • | If neither having pregnancy nor delivery, we assigned censoring labels. Based on the protocol, we used these labels to preserve outcome distribution in the target population and resolve class imbalance by inversely weighting the uncensored outcomes considering both the censored and uncensored ones. |
|  |  |  |  | • | Steps to determine candidate predictors were described in the protocol, to avoid zero variance, perfect separation problem, outcome leakage, and redundancy, and to mimic real-world setting. |
|  |  |  |  | • | (Supplement) All candidate predictors, including non-demographical associated factors, have non-zero variances (eTable 8 in R Markdown). There were 460 candidate predictors fulfilling this criterion. We also showed in the same eTable that there are 426 candidate predictors without perfect separation. |
|  |  |  |  | • | (Supplement) To prevent outcome leakage, we removed the maternal or baby diagnosis/procedure codes that demonstrated the delivery or after delivery care (typically up to 6 weeks following childbirth; eTable 9 in R Markdown). We only used the existing codes in the training set to determine outcome-leaker codes based on the previous codes for determining delivery or immediately after delivery care (eTable 7 in R Markdown). There were 54 codes that may leak the outcome. |
|  |  |  |  | • | (Supplement) To avoid redundancy, we computed pair-wise Pearson correlation coefficients (eTable 10 in R Markdown). None of the estimates showed a perfect correlation (*r*=1). High correlations (i.e., >~0.70) were reasonably identified between latent candidate predictors and the code components. We did not remove those pairs of predictors. |
|  |  |  |  | • | (Supplement) To deal with a problem in which a healthcare provider does not have access to medical records of other providers, medical histories were quantified as Kaplan-Meier (KM) estimators for each candidate predictors, so called historical rates. |
|  |  |  |  | • | (Supplement) The historical rates of all candidate predictors were fitted to a principal component (PC) model. |
|  |  |  |  | • | (Supplement) Then, 1-bit stochastic gradient descent transformation was applied using the post-normalization, feature-wise average based on nationwide training set to determine if a value is lower, equal to, or higher than the average respectively as -1, 0, and 1. |
|  |  |  | ✓ |  | State how missing values were handled |
|  |  |  |  | ❖ | Missing value handling: missing outcome was labelled as censoring and accounted for weighting; not applicable for medical histories since the positive value was defined as existence of a code entry, while otherwise determined the negative value |
|  |  |  |  | • | If neither having pregnancy nor delivery, we assigned censoring labels. Based on the protocol, we used these labels to preserve outcome distribution in the target population and resolve class imbalance by inversely weighting the uncensored outcomes considering both the censored and uncensored ones. |
|  |  |  |  | • | Candidate predictors consisted of medical histories defined by one or more codes of diagnosis and procedure. |
|  |  |  | ✓ |  | Describe the basic statistics of the dataset, particularly of the response variable. These include the ratio of positive to negative classes for a classification problem and the distribution of the response variable for regression problem. |
|  |  |  |  | ❖ | Classification outcome ratio: internal pre-calibration split (74,019 negatives; 12,011 positives); internal calibration split (18,504 negatives; 3,002 positives); external random split (23,240 negatives; 3,692 positives); external geographical split (13,698 negatives; 2,120 positives); external temporal split (15,484 negatives; 2,736 positives); external geotemporal split (1,994 negatives; 230 positives) |
|  |  |  |  | ❖ | Regression outcome distribution: if the outcome was predicted as event by DI-VNN, these were PROM times as average weeks from the time of prediction in: (1) internal validation set (calibration set) (15, 95% CI 11 to 18; *n*=760); (2) external random split (18, 95% CI 14 to 21; *n*=973); (3) external geographical split (9, 95% CI 6 to 13; *n*=500); (4) external temporal split (15, 95% CI 12 to 19; *n*=687); and (5) external geotemporal split (3, 95% CI 1 to 5; *n*=157). If the outcome was predicted as non-event by DI-VNN, these were the ends of pregnancies from the time of prediction in: (1) internal validation set (calibration set) (40, 95% CI 39 to 41; *n*=20,746); (2) external random split (42, 95% CI 40 to 43; *n*=25,959); (3) external geographical split (37, 95% CI 35 to 38; *n*=15,318); (4) external temporal split (52, 95% CI 50 to 53; *n*=17,533); and (5) external geotemporal split (60, 95% CI 54 to 65; *n*=2,067). |
|  |  |  |  | • | Figure 1. Subject selection by applying a retrospective design and data partitioning for internal and external validations. |
|  |  |  |  | • | (Supplement) Challenge on estimation task is reasonable considering different distributions among internal and external validation sets. In internal validation set, of which we only utilized the calibration set for model evaluation, predicted events by DI-VNN happened 15 weeks (95% CI 11 to 18; *n*=760) on average from the time of prediction. This was similar to those in external random (18, 95% CI 14 to 21; *n*=973) and temporal splits (15, 95% CI 12 to 19; *n*=687), but later than those in external geographical (9, 95% CI 6 to 13; *n*=500) and geotemporal splits (3, 95% CI 1 to 5; *n*=157). Meanwhile, the pregnancies predicted as non-events by DI-VNN ended at 40 weeks (95% CI 39 to 41; *n*=20,746) on average from time of prediction. This was earlier than those in external random (42, 95% CI 40 to 43; *n*=25,959), temporal (52, 95% CI 50 to 53; *n*=17,533), and geotemporal splits (60, 95% CI 54 to 65; *n*=2,067), but later than that in external geographical split (37, 95% CI 35 to 38; *n*=15,318). |
|  |  |  | ✓ |  | Define the model validation strategies. Internal validation is the minimum requirement; external validation should also be performed whenever possible. |
|  |  |  |  | ❖ | Internal validation: hyperparameter tuning by 10-fold cross-validation; final training, calibration, and external validation set by bootstrapping for 30 times |
|  |  |  |  | ❖ | External validation: simple random splitting; stratified random splitting by geographical, temporally, and geotemporal variables |
|  |  |  |  | • | We split the dataset for internal and external validation. As recommended, we split out a dataset for external validation by geographical, temporal, and geotemporal splitting, approximately covering ~20% of visits. This reflected the situation in some real-world settings but not that in nationwide, which represented by ~20% random split of the remaining set; thus leaving ~64% of the original sample size for internal validation. |
|  |  |  |  | • | (Supplement) For hyperparameter tuning, we applied 5-fold cross validation, instead of 10-fold as applied for PC modeling. Meanwhile, the final training and calibration for each model were conducted by bootstrapping for 30 times. |
|  |  |  | ✓ |  | Specify the internal validation strategy. Common methods include random split, time-based split, and patient-based split. |
|  |  |  |  | ❖ | Internal validation: hyperparameter tuning by 10-fold cross-validation; final training, calibration, and external validation set by bootstrapping for 30 times |
|  |  |  |  | • | (Supplement) For hyperparameter tuning, we applied 5-fold cross validation, instead of 10-fold as applied for PC modeling. Meanwhile, the final training and calibration for each model were conducted by bootstrapping for 30 times. |
|  |  |  |  | • | (Supplement) In addition, uncertainty intervals were also computed by bootstrapping for 30 times. |
|  |  |  | ✓ |  | Define the validation metrics. For regression problems, the normalized root-mean-square error should be used. For classification problems, the metrics should include sensitivity, specificity, positive predictive value, negative predictive value, area under the ROC curve, and calibration plot. |
|  |  |  |  | ❖ | Validation metrics: calibration plot, intercept and slope; AUROC; sensitivity, specificity, positive likelihood ratio (LR+), and negative likelihood ratio (LR-); proportion of predicted time that includes true one; RMSE within time window with acceptable precision per outcome |
|  |  |  |  | • | Calibration (plot, intercept, and slope) and discrimination (i.e. AUROC) were computed for classification tasks. We also computed sensitivity, specificity, positive likelihood ratio (LR+), and negative likelihood ratio (LR-). |
|  |  |  |  | • | For estimation, we computed a proportion of weeks in which each predicted time, in weeks, was included within an interval estimate of the true one. The interval had to be the maximum ± *x* weeks when predicting > *x* weeks, e.g., for any women predicted to deliver in 6 weeks, this number should fall into the true time of delivery within ± 6 weeks. |
|  |  |  |  | • | We determined the minimum and maximum predicted times of delivery with acceptable precision for each predicted outcome of PROM based on a visual assessment using internal validation. Root mean square error (RMSE) was computed within this acceptable range. We also evaluated the best time window using the best model for PROM prognostic predictions. |
|  |  |  | ✓ |  | For retrospective studies, split the data into a derivation set and a validation set. For prospective studies, define the starting time for validation data collection. |
|  |  |  |  | ❖ | Data partition: 64% derivation set and 36% validation set |
|  |  |  |  | • | We split the dataset for internal and external validation. As recommended, we split out a dataset for external validation by geographical, temporal, and geotemporal splitting, approximately covering ~20% of visits. This reflected the situation in some real-world settings but not that in nationwide, which represented by ~20% random split of the remaining set; thus leaving ~64% of the original sample size for internal validation. |
| 8 | Methods | Build the predictive model | ✓ |  | Identify independent variables that predominantly take a single value (e.g., being zero 99% of the time) |
|  |  |  |  | ❖ | Number of independent variables with zero variance: none; outcome-wise variances are shown in eTable 8 in the Supplement |
|  |  |  |  | • | Details of the candidate predictors and selection are described in eTable 8 of the Supplement. |
|  |  |  |  | • | (Supplement) All candidate predictors, including non-demographical associated factors, have non-zero variances (eTable 8 in R Markdown). There were 460 candidate predictors fulfilling this criterion. We also showed in the same eTable that there are 426 candidate predictors without perfect separation. |
|  |  |  | ✓ |  | Identify and remove redundant independent variables |
|  |  |  |  | ❖ | Number of redundant independent variables: none; Pearson correlation estimates are shown in eTable 8 in the Supplement |
|  |  |  |  | • | Details of the candidate predictors and selection are described in eTable 8 of the Supplement. |
|  |  |  |  | • | (Supplement) To avoid redundancy, we computed pair-wise Pearson correlation coefficients (eTable 10 in R Markdown). None of the estimates showed a perfect correlation (*r*=1). High correlations (i.e., >~0.70) were reasonably identified between latent candidate predictors and the code components. We did not remove those pairs of predictors. |
|  |  |  | ✓ |  | Identify the independent variables that may suffer from the perfect separation problem |
|  |  |  |  | ❖ | Perfect-separation variables: removed; shown in eTable 8 Supplemental Information |
|  |  |  |  | • | (Supplement) We also showed in the same eTable that there are 426 candidate predictors without perfect separation. |
|  |  |  | ✓ |  | Report the number of independent variables, the number of positive examples, and the number of negative examples |
|  |  |  |  | ❖ | Number of independent variables: shown in eTable 8 in Supplemental Information |
|  |  |  |  | • | Details of the candidate predictors and selection are described in eTable 8 “Supplementary Material”. |
|  |  |  |  | • | Table 1. Baseline characteristics of subjects for association tests and internal validation set. |
|  |  |  | ✓ |  | Assess whether sufficient data are available for a good fit of the model. In particular, for classification, there should be a sufficient number of observations in both positive and negative classes. |
|  |  |  |  | ❖ | Classification problem (bootstrapping for 30 times for final training): internal pre-calibration split (non DI-VNN: 12,011 events per 9/372/60 variables = 1,335/32/200 EPV; DI-VNN: 80% ´ 12,011 = 9,609 events per 144 variables = 67 EPV); internal calibration split (3,002 events per 1 variable = 3,002 EPV) |
|  |  |  |  | ❖ | Regression problem (bootstrapping for 30 times for final training): internal validation set (non DI-VNN: 107,536 visits per 9/372/60 variables = 11,948/289/1792 EPV; DI-VNN: 86,029 visits per 144 variables = 597 EPV) |
|  |  |  |  | • | Figure 1. Subject selection by applying a retrospective design and data partitioning for internal and external validations. |
|  |  |  |  | • | Briefly, the first model was a statistical, i.e., ridge regression (RR), on 9 of 12 latent candidate predictors selected by the systematic human learning. One of them was not associated with PROM (see Results), and we did not select low SES and maternal age for reducing use of private data and preventing social and economic discrimination. Opposite to systematic human learning, we applied unsupervised machine learning to transform candidate predictors into principal components (PCs). These were used for supervised machine learning algorithms of an elastic net regression (PC-ENR), random forest (PC-RF), and gradient boosting machine (PC-GBM). The latter two algorithms outperformed other algorithms for pregnancy outcomes. Based on the PC-ENR model, the PCs were reduced into 60 PCs (see the protocol) to pursue 200 events per variable (EPV) for the PC-RF and PC-GBM, as recommended by the prediction model risk of bias assessment tools (PROBAST). The fifth model was the DI-VNN. It was developed to achieve moderate predictive performance but interpretable results based on recent studies. The DI-VNN allows deep exploration of how this algorithm works. For this model, there were 144 candidate predictors after differential analyses with multiple testing corrections. |
|  |  |  |  | • | We split the dataset for internal and external validation. As recommended, we split out a dataset for external validation by geographical, temporal, and geotemporal splitting, approximately covering ~20% of visits. This reflected the situation in some real-world settings but not that in nationwide, which represented by ~20% random split of the remaining set; thus leaving ~64% of the original sample size for internal validation. |
|  |  |  | ✓ |  | Determine a set of candidate modelling techniques (e.g., logistic regression, random forest, or deep learning). If only one type of model was used, justify the decision for using that model. |
|  |  |  |  | ❖ | Candidate modelling techniques: logistic regression (ridge regression), logistic regression (elastic net regression), random forest, gradient boosting machine, and deep learning (deep-insight visible neural network, DI-VNN; which is a convolutional neural network, CNN) |
|  |  |  |  | • | Briefly, the first model was a statistical, i.e., ridge regression (RR), on 9 of 12 latent candidate predictors selected by the systematic human learning. One of them was not associated with PROM (see Results), and we did not select low SES and maternal age for reducing use of private data and preventing social and economic discrimination. Opposite to systematic human learning, we applied unsupervised machine learning to transform candidate predictors into principal components (PCs). These were used for supervised machine learning algorithms of an elastic net regression (PC-ENR), random forest (PC-RF), and gradient boosting machine (PC-GBM). The latter two algorithms outperformed other algorithms for pregnancy outcomes. Based on the PC-ENR model, the PCs were reduced into 60 PCs (see the protocol) to pursue 200 events per variable (EPV) for the PC-RF and PC-GBM, as recommended by the prediction model risk of bias assessment tools (PROBAST). The fifth model was the DI-VNN. It was developed to achieve moderate predictive performance but interpretable results based on recent studies. The DI-VNN allows deep exploration of how this algorithm works. For this model, there were 144 candidate predictors after differential analyses with multiple testing corrections. |
|  |  |  | ✓ |  | Define the performance metrics to select the best model |
|  |  |  |  | ❖ | Performance metrics for model selection: highest AUROC in most of external validation, highest proportion of predicted time that includes true one highest AUROC in most of external validation |
|  |  |  |  | • | Calibration (plot, intercept, and slope) and discrimination (i.e. AUROC) were computed for classification tasks. |
|  |  |  |  | • | For estimation, we computed a proportion of weeks in which each predicted time, in weeks, was included within an interval estimate of the true one. The interval had to be the maximum ± *x* weeks when predicting > *x* weeks, e.g., for any women predicted to deliver in 6 weeks, this number should fall into the true time of delivery within ± 6 weeks. |
|  |  |  | ✓ |  | Specify the model selection strategy. Common methods include K-fold validation or bootstrap to estimate the lost function on a grid of candidate parameter values. For K-fold validation, proper stratification by the response variable is needed. |
|  |  |  |  | ❖ | Model selection strategy: all models using grid search and including censored outcome to compute censored outcome weighting; hyperparameter tuning for non DI-VNN by *k*-fold cross validation (minimum 12,011 events must considerably exist in each fold); hyperparameter tuning of DI-VNN and final training by bootstrapping for 30 times |
|  |  |  |  | • | If neither having pregnancy nor delivery, we assigned censoring labels. Based on the protocol […], we used these labels to preserve outcome distribution in the target population and resolve class imbalance by inversely weighting the uncensored outcomes considering both the censored and uncensored ones. |
|  |  |  |  | • | (Supplement) For hyperparameter tuning, we applied 5-fold cross validation, instead of 10-fold as applied for PC modeling. Meanwhile, the final training and calibration for each model were conducted by bootstrapping for 30 times. |
|  |  |  |  | • | (Supplement) In addition, uncertainty intervals were also computed by bootstrapping for 30 times. |
|  |  |  | ✓ |  | (A desirable but not mandatory item) For model selection, include discussion on (1) balance between model accuracy and model simplicity or interpretability, and (2) the familiarity with the modelling techniques of the end user |
|  |  |  |  | ❖ | Modelling balance between accuracy and interpretability: DI-VNN was developed to balance these aspects |
|  |  |  |  | ❖ | Familiarity with the modelling techniques: RR was provided as a comparator and the most familiar modelling technique in this study |
|  |  |  |  | • | Briefly, the first model was a statistical, i.e., ridge regression (RR), on 9 of 12 latent candidate predictors selected by the systematic human learning. One of them was not associated with PROM (see Results), and we did not select low SES and maternal age for reducing use of private data and preventing social and economic discrimination. |
|  |  |  |  | • | The fifth model was the DI-VNN. It was developed to achieve moderate predictive performance but interpretable results based on recent studies. The DI-VNN allows deep exploration of how this algorithm works. |
| 9 | Results | Report the final model and performance | ✓ |  | Report the predictive performance of the final model in terms of the validation metrics specified in the methods section |
|  |  |  |  | ❖ | Predictive performance report: calibration plot, intercept and slope (Figure 2a); AUROC (Figure 2c); proportion of predicted time that includes true one (Figure 3a); RMSE within time window with acceptable precision per outcome (Figure 3b) |
|  |  |  |  | • | Figure 2. Model evaluation. |
|  |  |  |  | • | Figure 3. Estimation plots. |
|  |  |  | ✓ |  | If possible, report the parameter estimates in the model and their confidence intervals. When the direct calculation of confidence intervals is not possible, report nonparametric estimates from bootstrap samples. |
|  |  |  |  | ❖ | Parameter estimates: shown by eTable 14 to 20 in Supplemental Information |
|  |  |  |  | • | Weights, variable importance values, and intermediate outputs, which indicated the extent a predictor contributes to a prediction, are respectively shown for (1) the RR and PC-ENR (eTables 14 to 16 in the Supplement); (2) the PC-RF and PC-GBM (eTables 15, 17, and 18 in the Supplement); and (3) the DI-VNN (eTables 19 and 20 in the Supplement). |
|  |  |  | ✓ |  | Comparison with other models in the literature should be based on confidence intervals |
|  |  |  |  | ❖ | Other models in the literature: the models were systematically selected, and unfortunately, did not report the predictive performances by interval estimates; but, we used interval estimates of the predictive performances to compare those of our models with those of the other models from previous studies. |
|  |  |  |  | • | Figure 2. Model evaluation. |
|  |  |  | ✓ |  | Interpretation of the final model. If possible, report what variables were shown to be predictive of the response variable. State which subpopulation has the best prediction and which subpopulation is most difficult to predict. |
|  |  |  |  | ❖ | Predictive variables: N760 (acute vaginitis), B379 (unspecified candidiasis), and 9059 (other microscopic examination of blood) |
|  |  |  |  | ❖ | Subpopulation with the best prediction: visits from 46 weeks before the end of pregnancy |
|  |  |  |  | ❖ | Subpopulation that was most difficult to predict: visits more than 46 weeks before the end of pregnancy |
|  |  |  |  | • | For an exploratory data analysis, most of the AUROC intervals were greater than 0.5 from 44 ± 2 weeks before the end of the pregnancy (eFigure 15a in the Supplement). |
|  |  |  |  | • | (Supplement) We began from the most visually distinguished array (eFigure 15b in Appendix) which was ONT:171. These were N760 (acute vaginitis) and B379 (unspecified candidiasis). |
|  |  |  |  | • | (Supplement) Another distinguished array, ONT:144, was connected to the same node as that of the previous array. The feature member (9059, other microscopic examination of blood) tended to contribute to the same outcome as that of “unspecified candidiasis”. |
| 10 | Discussion | Clinical implications | ✓ |  | Report the clinical implications derived from the obtained predictive performance. For example, report the dollar amount that could be saved with better prediction. How many patients could benefit from a care model leveraging the model prediction? And to what extent? |
|  |  |  |  | ❖ | Potential cost efficiency: cost reduction for prognostic testing and future studies on PROM complications |
|  |  |  |  | ❖ | Potential healthcare impact: potential improvement on patient safety |
|  |  |  |  | • | The DI-VNN model in this study outperformed previous models; it used a large training set and external validation sets (8778 visits and 3352 subjects for events only) and did not require biomarker testing. |
| 11 | Discussion | Limitations of the model | ✓ |  | Discuss the following potential limitations: • Assumed input and output data format; • (Desirable but not mandatory items) Potential pitfalls in interpreting the model; • Potential bias of the data used in modelling; • Generalizability of the data |
|  |  |  |  | ❖ | Assumed input and output data format: consider diagnose conditions or determine procedures are similar to those whose codes are used in DI-VNN as the predictors |
|  |  |  |  | ❖ | Potential pitfalls in interpreting the model: a user needs specific competences in medicine, statistics, and machine learning |
|  |  |  |  | ❖ | Potential bias of the data used in modelling: population in the dataset covered Asian and Austronesian |
|  |  |  |  | ❖ | Generalizability of the data: limited as a preliminary model to select candidate for a more sensitive biomarker test that needs high-resource setting |
|  |  |  |  | • | However, we noted several limitations in our study. Although the DI-VNN outperformed those from previous studies, we could only apply it as a preliminary model for PROM because clinical acceptance requires an AUROC of ≥0.8.50 More-sensitive models are still needed for use as second-line models. As recommended in a clinician checklist for assessing the suitability of machine learning applications in healthcare (eTable 3 in the Supplement), external validation and determining an optimal threshold using local data are needed. |
| 12 | Discussion |  | ✓ |  | (Desirable but not mandatory items) Report unexpected signs of coefficients, indicating collinearity or complex interaction between predictor variables |
|  |  |  |  | ❖ | Unexpected signs of coefficients: H527 (unspecified disorder of refraction), 734 (flat foot), H521 (myopia), and H522 (astigmatism) |
|  |  |  |  | • | (Supplement) From ONT:167 and ONT:149 on the deeper layer, we can find unusual features in the context of PROM, which are H527 (unspecified disorder of refraction) preferring non-events while 734 (flat foot), H521 (myopia), and H522 (astigmatism) preferring events. These codes might be responses to the subject symptoms of edema in the feet and blurry vision. Both symptoms in a pregnant woman may be typically associated with severe preeclampsia. But, a doctor may avoid this association if the context does not support the symptoms, e.g. symptoms by a non-pregnant subject. This may lead a doctor to assign these codes responding to those symptoms. |

| DOMAIN 1: Participants | | | | | |
| --- | --- | --- | --- | --- | --- |
| Risk of Bias | | | | | |
| *Describe the sources of data and criteria for participant selection:* | | | | | |
|  | • | We applied a retrospective design to select subjects from a nationwide health insurance dataset provided by a government-owned health insurance company in Indonesia. | | | |
|  | • | We included health insurance holders of 12~55-year-old females who had visited affiliated healthcare providers. We excluded visits after delivery. For a person who was pregnant twice within the period, we labeled the same person as a different subject for each pregnancy period. A complete list of codes for determining delivery or immediately after delivery care is available in eTable 7 of the Supplement. | | | |
|  | • | The outcome for the classification task was an event for a subject that had encountered the O42 code, which is the International Classification of Disease version 10 (ICD-10) code for PROM. Otherwise, a subject was assigned as a nonevent if the pregnancy ended within the dataset period using the same codes for pregnancy termination. | | | |
|  | • | If neither having pregnancy nor delivery, we assigned censoring labels. Based on the protocol, we used these labels to preserve outcome distribution in the target population and resolve class imbalance by inversely weighting the uncensored outcomes considering both the censored and uncensored ones. | | | |
|  | • | Meanwhile, the outcome for the estimation task was the number of days from the latest visit encountering of the outcome code to a visit when the prediction model was used. | | | |
|  | • | Candidate predictors consisted of medical histories defined by one or more codes of diagnosis and procedure. | | | |
|  | • | We split the dataset for internal and external validation. As recommended, we split out a dataset for external validation by geographical, temporal, and geotemporal splitting, approximately covering ~20% of visits. This reflected the situation in some real-world settings but not that in nationwide, which represented by ~20% random split of the remaining set; thus leaving ~64% of the original sample size for internal validation. | | | |
|  | |  | | Dev | Val |
| 1.1  Were appropriate data sources used, e.g. cohort, RCT or nested case-control study data? | | | | Y | Y |
| 1.2  Were all inclusions and exclusions of participants appropriate? | | | | Y | Y |
| Risk of bias introduced by selection of participants | | | RISK: | Low | Low |
|  |  |  | *(low/ high/ unclear)* |  |  |
| *Rationale of bias rating:* | | | | | |
|  | ❖ | This study used retrospective cohort from health insurance dataset consisting medical histories. Medical history only used ICD-10, a standardized coding system, that is, do not have a wide range of possibility for an error in term of codifying what a doctor expects conditional on the gather information about a subject. The models were intended to predict what a doctor codes for a patient in the future, whether it will be included as an event or non-event as defined in this study, not what will be happened in a subject’s physiology. In addition, the classification outcome was weighted by taking censored outcome into account. This enabled correct estimation of baseline risk, which obtained corrected absolute values for the predicted probability and calibration measures of the models. | | | |
|  | ❖ | The intended population is represented by the selection criteria. The models will mostly use medical histories from the health insurance claim data in providers at any levels of care. This means only those who visits the providers will be applied using the models, as included in the inclusion criteria. The subject is not necessarily pregnant; thus, the model application will be not limited to pregnant women. The selection criteria did not increase or decrease risk of developing PROM, that may introduce selection bias. As a preliminary model, a wide range of population for the prediction is needed. | | | |
|  | ❖ | Validation used the same data source but different visits. The same design and selection criteria were applied. Therefore, the assessment results are the same with the development ones. | | | |

| DOMAIN 2: Predictors | | | | | |
| --- | --- | --- | --- | --- | --- |
| Risk of Bias | | | | | |
| *List and describe predictors included in the final model, e.g. definition and timing of assessment:* | | | | | |
|  | • | We included health insurance holders of 12~55-year-old females who had visited affiliated healthcare providers. We excluded visits after delivery. | | | |
|  | • | Candidate predictors consisted of medical histories defined by one or more codes of diagnosis and procedure. | | | |
|  | • | Steps to determine candidate predictors were described in the protocol, to avoid zero variance, perfect separation problem, outcome leakage, and redundancy, and to mimic real-world setting. | | | |
|  | • | Details of the candidate predictors and selection are described in eTable 8 of the Supplement. | | | |
|  | • | We split the dataset for internal and external validation. As recommended, we split out a dataset for external validation by geographical, temporal, and geotemporal splitting, approximately covering ~20% of visits. This reflected the situation in some real-world settings but not that in nationwide, which represented by ~20% random split of the remaining set; thus leaving ~64% of the original sample size for internal validation. | | | |
|  | |  | | Dev | Val |
| 2.1  Were predictors defined and assessed in a similar way for all participants? | | | | PY | PY |
| 2.2 Were predictor assessments made without knowledge of outcome data? | | | | Y | Y |
| 2.3  Are all predictors available at the time the model is intended to be used? | | | | Y | Y |
| Risk of bias introduced by predictors or their assessment | | | RISK: | Low | Low |
|  |  |  | *(low/ high/ unclear)* |  |  |
| *Rationale of bias rating:* | | | | | |
|  | ❖ | All predictors were derived from medical history. Albeit the same diagnosis or procedure was determined with different quality, this was determined in one country. There is only a single professional association for each medical specialty in this country, that has an authority for developing diagnosis and procedure guidelines; thus, each diagnosis or procedure was determined based on a single consensus. The codification was also standardized based on ICD-10. In addition, associated factors were derived from the medical histories. | | | |
|  | ❖ | In health insurance dataset, the future outcome is unknown for each visit when the diagnosis or procedure codes were entered. The codes, that may easily infer outcome, were already removed and transparently reported. | | | |
|  | ❖ | For medical history, the day intervals were computed from the code entries up to the time of prediction; thus, these are available at the time of prediction. | | | |
|  | ❖ | Validation used the same predictors. The same predictor/feature extraction was applied. Therefore, the assessment results are the same with the development ones. | | | |

| DOMAIN 3: Outcome | | | | | |
| --- | --- | --- | --- | --- | --- |
| Risk of Bias | | | | | |
| *Describe the outcome, how it was defined and determined, and the time interval between predictor assessment and outcome determination:* | | | | | |
|  | • | For a person who was pregnant twice within the period, we labeled the same person as a different subject for each pregnancy period. A complete list of codes for determining delivery or immediately after delivery care is available in eTable 7 of the Supplement. | | | |
|  | • | We developed prediction models to classify if a visit was made by a subject for which the pregnancy period ended with PROM, and to estimate the time of delivery. | | | |
|  | • | The outcome for the classification task was an event for a subject that had encountered the O42 code, which is the International Classification of Disease version 10 (ICD-10) code for PROM. Otherwise, a subject was assigned as a nonevent if the pregnancy ended within the dataset period using the same codes for pregnancy termination. | | | |
|  | • | If neither having pregnancy nor delivery, we assigned censoring labels. Based on the protocol, we used these labels to preserve outcome distribution in the target population and resolve class imbalance by inversely weighting the uncensored outcomes considering both the censored and uncensored ones. | | | |
|  | • | Meanwhile, the outcome for the estimation task was the number of days from the latest visit encountering of the outcome code to a visit when the prediction model was used. | | | |
|  | • | We split the dataset for internal and external validation. As recommended, we split out a dataset for external validation by geographical, temporal, and geotemporal splitting, approximately covering ~20% of visits. This reflected the situation in some real-world settings but not that in nationwide, which represented by ~20% random split of the remaining set; thus leaving ~64% of the original sample size for internal validation. | | | |
|  | |  | | Dev | Val |
| 3.1  Was the outcome determined appropriately? | | | | PY | PY |
| 3.2  Was a pre-specified or standard outcome definition used? | | | | Y | Y |
| 3.3  Were predictors excluded from the outcome definition? | | | | Y | Y |
| 3.4  Was the outcome defined and determined in a similar way for all participants? | | | | PY | PY |
| 3.5  Was the outcome determined without knowledge of predictor information? | | | | Y | Y |
| 3.6  Was the time interval between predictor assessment and outcome determination appropriate? | | | | Y | Y |
| Risk of bias introduced by the outcome or its determination | | | RISK: | Low | Low |
|  |  |  | *(low/ high/ unclear)* |  |  |
| *Rationale of bias rating:* | | | | | |
|  | ❖ | The models were intended to predict what a doctor codes for a patient in the future, whether it will be included as an event or non-event as defined in this study, not what will be happened in a subject’s physiology. PROM definition is well-established based on a point-of-care, amnion fluid leak test and the assessment of labor onset. This is considerably simple to determine. | | | |
|  | ❖ | The outcome was defined by an ICD-10 code for diagnosis of PROM. This code is straightforward. The definition was not favorably adjusted to achieve the best estimate of model performances. | | | |
|  | ❖ | Only a single ICD-10 code was used for events. This code was already removed; thus, the predictors from medical histories did not include this code. | | | |
|  | ❖ | Albeit PROM was determined with different quality, this was determined in one country. There is only a single professional association for obstetrics and gynecology in this country, that has an authority for developing diagnosis and procedure guidelines of PROM; thus, each diagnosis or procedure was determined based on a single consensus. The codification was also standardized based on ICD-10. In addition, PROM is simple to determine. | | | |
|  | ❖ | In health insurance dataset, the outcome may be determined based on the results of previous diagnostic tests but these results were not explicitly revealed by the previous diagnosis or procedure codes. Meanwhile, the codes, that may easily infer non-events (typically happen only during the delivery or post-delivery period), were already removed and transparently reported. | | | |
|  | ❖ | The models were developed for both prediction and predictor exploration; thus, a wide time interval between predictors and outcome was obtained up to the time of outcome to find the earliest time PROM can be predicted with acceptable performances. To help a user for interpreting the classification models regarding the timing of the outcome, the estimation models were also developed. | | | |
|  | ❖ | Validation used the same outcomes. The outcome was determined the same way. Therefore, the assessment results are the same with the development ones. | | | |

| DOMAIN 4: Analysis | | | | | |
| --- | --- | --- | --- | --- | --- |
| Risk of Bias | | | | | |
| *Describe numbers of participants, number of candidate predictors, outcome events and events per candidate predictor:* | | | | | |
|  | • | Figure 1. Subject selection by applying a retrospective design and data partitioning for internal and external validations. | | | |
|  | • | Briefly, the first model was a statistical, i.e., ridge regression (RR), on 9 of 12 latent candidate predictors selected by the systematic human learning. One of them was not associated with PROM (see Results), and we did not select low SES and maternal age for reducing use of private data and preventing social and economic discrimination. Opposite to systematic human learning, we applied unsupervised machine learning to transform candidate predictors into principal components (PCs). These were used for supervised machine learning algorithms of an elastic net regression (PC-ENR), random forest (PC-RF), and gradient boosting machine (PC-GBM). The latter two algorithms outperformed other algorithms for pregnancy outcomes. Based on the PC-ENR model, the PCs were reduced into 60 PCs (see the protocol) to pursue 200 events per variable (EPV) for the PC-RF and PC-GBM, as recommended by the prediction model risk of bias assessment tools (PROBAST). The fifth model was the DI-VNN. It was developed to achieve moderate predictive performance but interpretable results based on recent studies. The DI-VNN allows deep exploration of how this algorithm works. For this model, there were 144 candidate predictors after differential analyses with multiple testing corrections. | | | |
|  | • | Weights, variable importance values, and intermediate outputs, which indicated the extent a predictor contributes to a prediction, are respectively shown for (1) the RR and PC-ENR (eTables 14 to 16 in the Supplement); (2) the PC-RF and PC-GBM (eTables 15, 17, and 18 in the Supplement); and (3) the DI-VNN (eTables 19 and 20 in the Supplement). | | | |
|  | • | Figure 2. Model evaluation. | | | |
|  | • | Candidate predictors consisted of medical histories defined by one or more codes of diagnosis and procedure. | | | |
|  | • | Details of the candidate predictors and selection are described in eTable 8 of the Supplement. | | | |
| *Describe how the model was developed (for example in regards to modelling technique (e.g. survival or logistic modelling), predictor selection, and risk group definition):* | | | | | |
|  | • | If neither having pregnancy nor delivery, we assigned censoring labels. Based on the protocol, we used these labels to preserve outcome distribution in the target population and resolve class imbalance by inversely weighting the uncensored outcomes considering both the censored and uncensored ones. | | | |
|  | • | Briefly, the first model was a statistical, i.e., ridge regression (RR), on 9 of 12 latent candidate predictors selected by the systematic human learning. One of them was not associated with PROM (see Results), and we did not select low SES and maternal age for reducing use of private data and preventing social and economic discrimination. Opposite to systematic human learning, we applied unsupervised machine learning to transform candidate predictors into principal components (PCs). These were used for supervised machine learning algorithms of an elastic net regression (PC-ENR), random forest (PC-RF), and gradient boosting machine (PC-GBM). The latter two algorithms outperformed other algorithms for pregnancy outcomes. Based on the PC-ENR model, the PCs were reduced into 60 PCs (see the protocol) to pursue 200 events per variable (EPV) for the PC-RF and PC-GBM, as recommended by the prediction model risk of bias assessment tools (PROBAST). The fifth model was the DI-VNN. It was developed to achieve moderate predictive performance but interpretable results based on recent studies. The DI-VNN allows deep exploration of how this algorithm works. For this model, there were 144 candidate predictors after differential analyses with multiple testing corrections. | | | |
| *Describe whether and how the model was validated, either internally (e.g. bootstrapping, cross validation, random split sample) or externally (e.g. temporal validation, geographical validation, different setting, different type of participants):* | | | | | |
|  | • | Figure 1. Subject selection by applying a retrospective design and data partitioning for internal and external validations. | | | |
|  | • | We split the dataset for internal and external validation. As recommended, we split out a dataset for external validation by geographical, temporal, and geotemporal splitting, approximately covering ~20% of visits. This reflected the situation in some real-world settings but not that in nationwide, which represented by ~20% random split of the remaining set; thus leaving ~64% of the original sample size for internal validation. | | | |
|  | • | (Supplement) For hyperparameter tuning, we applied 5-fold cross validation, instead of 10-fold as applied for PC modeling. Meanwhile, the final training and calibration for each model were conducted by bootstrapping for 30 times. | | | |
|  | • | (Supplement) In addition, uncertainty intervals were also computed by bootstrapping for 30 times. | | | |
| *Describe the performance measures of the model, e.g. (re)calibration, discrimination, (re)classification, net benefit, and whether they were adjusted for optimism:* | | | | | |
|  | • | Calibration (plot, intercept, and slope) and discrimination (i.e. AUROC) were computed for classification tasks. We also computed sensitivity, specificity, positive likelihood ratio (LR+), and negative likelihood ratio (LR-). | | | |
|  | • | For estimation, we computed a proportion of weeks in which each predicted time, in weeks, was included within an interval estimate of the true one. The interval had to be the maximum ± *x* weeks when predicting > *x* weeks, e.g., for any women predicted to deliver in 6 weeks, this number should fall into the true time of delivery within ± 6 weeks. | | | |
|  | • | We determined the minimum and maximum predicted times of delivery with acceptable precision for each predicted outcome of PROM based on a visual assessment using internal validation. Root mean square error (RMSE) was computed within this acceptable range. We also evaluated the best time window using the best model for PROM prognostic predictions. | | | |
| *Describe any participants who were excluded from the analysis:* | | | | | |
|  | • | We included health insurance holders of 12~55-year-old females who had visited affiliated healthcare providers. We excluded visits after delivery. For a person who was pregnant twice within the period, we labeled the same person as a different subject for each pregnancy period. A complete list of codes for determining delivery or immediately after delivery care is available in eTable 7 of the Supplement. | | | |
|  | • | If neither having pregnancy nor delivery, we assigned censoring labels. Based on the protocol, we used these labels to preserve outcome distribution in the target population and resolve class imbalance by inversely weighting the uncensored outcomes considering both the censored and uncensored ones. | | | |
| *Describe missing data on predictors and outcomes as well as methods used for missing data:* | | | | | |
|  | • | Candidate predictors consisted of medical histories defined by one or more codes of diagnosis and procedure. | | | |
|  | • | If neither having pregnancy nor delivery, we assigned censoring labels. Based on the protocol, we used these labels to preserve outcome distribution in the target population and resolve class imbalance by inversely weighting the uncensored outcomes considering both the censored and uncensored ones. | | | |
|  | |  | | Dev | Val |
| 4.1  Were there a reasonable number of participants with the outcome? | | | | Y | Y |
| 4.2  Were continuous and categorical predictors handled appropriately? | | | | Y | Y |
| 4.3  Were all enrolled participants included in the analysis? | | | | Y | Y |
| 4.4  Were participants with missing data handled appropriately? | | | | Y | Y |
| 4.5 Was selection of predictors based on univariable analysis avoided? | | | | Y |  |
| 4.6  Were complexities in the data (e.g. censoring, competing risks, sampling of controls) accounted for appropriately? | | | | Y | Y |
| 4.7  Were relevant model performance measures evaluated appropriately? | | | | Y | Y |
| 4.8  Were model overfitting and optimism in model performance accounted for? | | | | Y |  |
| 4.9 Do predictors and their assigned weights in the final model correspond to the results from multivariable analysis? | | | | Y |  |
| Risk of bias introduced by the analysis | | | RISK: | Low | Low |
|  |  |  | *(low/ high/ unclear)* |  |  |
| *Rationale of bias rating:* | | | | | |
|  | ❖ | Classification problem (bootstrapping for 30 times for final training): internal pre-calibration split (non DI-VNN: 12,011 events per 9/372/60 variables = 1335/32/200 EPV; DI-VNN: 80% × 12,011 = 9609 events per 144 variables = 67 EPV); internal calibration split (3002 events per 1 variable = 3002 EPV). EPV of 200 or more were determined for support vector machine, random forest, and shallow, fully-connected neural network (a single hidden layer with ten nodes). DI-VNN is not a vanilla neural network. Instead, it is a convolutional neural network with inputs (features) selected by convolution. There is no consensus for EPV in deep learning. Since EPV is an approach to prevent overfitting, we can assess this by comparing internal and external validations among models. DI-VNN have better agreement between internal and external validations compared to random forest and gradient boosting machine that achieved 200 EPV or more. | | | |
|  | ❖ | Regression problem (bootstrapping for 30 times for final training): internal validation set (non DI-VNN: 107,536 visits per 9/372/60 variables = 11,948/289/1792 EPV; DI-VNN: 86,029 visits per 144 variables = 597 EPV) | | | |
|  | ❖ | Outcome ratio for external validation: external random split (23,240 negatives; 3692 positives); external geographical split (13,698 negatives; 2120 positives); external temporal split (15,484 negatives; 2736 positives); external geotemporal split (1994 negatives; 230 positives) | | | |
|  | ❖ | Age at admission was categorized using widely accepted cut points (by national consensus for high-risk pregnancy based on maternal age). Other categorical predictors were predefined. Medical histories were kept as continuous variables and optimized by mapping these to the corresponding KM estimates. These were determined using training set only. | | | |
|  | ❖ | Validation used the same format for predictors. The medical histories for validation were mapped to the corresponding KM estimates determined using the training set. Weights used for transforming features into PCs in validation sets were also ones that were determined using the training set. This was also applied when transforming features by 1-bit stochastic gradient descent in validation sets. The average values based on differential analysis were determined using the training set. | | | |
|  | ❖ | All visits and subjects were included in analysis after selection criteria. The predictive performances for model selection were determined using all of these visits. Therefore, no exclusion was conducted because of uninterpretable findings, outliers, or missing data in predictors. Although visits with censoring outcomes were removed, we took into account these for weighting the uncensored outcome when training the models. The removal was completely random, because the censoring outcomes happened because the subjects were not pregnant or delivered within the dataset period. It is independent to the event of pregnancy and delivery. | | | |
|  | ❖ | There is no missing data in medical histories because of the nature of the feature extraction. As described above, censored outcomes were independent and took into account for weighting the uncensored outcome when training the models. | | | |
|  | ❖ | Feature selection in RR was based on existing knowledge (association diagram) followed by association tests with confounder adjustment. For 2^nd^ to 4^th^ models, principal components were computed and selected from the highest proportion of variance explained to pursue EPV requirement. Random forest and gradient boosting machine models also used a multivariable predictor selection by the elastic net regression by bootstrapping for 30 times. Eventually, DI-VNN applied quantile-to-quantile normalization that involves all candidate predictors before applying differential analysis (fitting a univariable regression for each pair of a candidate predictor and outcome). In this method, predictors were selected in context with other predictors via quantile-to-quantile normalization. In addition, multiple testing corrections were also applied. | | | |
|  | ❖ | Although time-to-event analysis, such as a Cox regression, was recommended for prediction with censored participants, this restricts us to apply other machine learning algorithms. Thus, we took into account the censored outcome for weighting each uncensored outcome by the half of inverse probability when training the models. Potential competing risks, e.g. preeclampsia, were already accounted as the candidate predictors because of the nature of medical histories as the predictors. There is no need for sampling of controls because of retrospective design in this study. | | | |
|  | ❖ | There were calibration plot and AUROC to evaluate the predictive performances. These were also adjusted for optimism by resampling. All evaluation metrics were expressed as 95% confidence intervals determined using the resampling subsets. | | | |
|  | ❖ | Internal validation was applied. Shrinkage technique by elastic net regression was also applied. For DI-VNN, bootstrapping for 30 times was applied when hyperparameter tuning and the final training. | | | |
|  | ❖ | Assigned weights in the final models of RR and PC-ENR were already fitted using the final set of predictors since the iterative processes were conducted by the programming, instead manually by human. For PC-RF and PC-GBM, feature selection by PC-ENR was only used to determine candidate predictors for both models. The parameters were then fitted, and those in the final models were also fitted using the final set of predictors. For DI-VNN, this also applies but using differential analysis for feature selection and backpropagation algorithm to fit the weights. | | | |
|  | ❖ | Validation applied the same procedure regarding post-enrolled data handling, missing data, censoring, competing risks, and evaluation metrics, with those of development. This is because of the same dataset but split carefully for external validation. Therefore, the assessment results are the same with the development ones. | | | |

| Overall judgement about risk of bias of the prediction model evaluation | | |
| --- | --- | --- |
| Overall judgement of risk of bias | RISK: | Low |
|  | *(low/ high/ unclear)* |  |
| *Summary of sources of potential bias:* | | |
| – | | |

eTable 3. Clinical checklists for assessing suitability of machine learning applications in healthcare

| Item | | Response | |
| --- | --- | --- | --- |
| Q1. | What is the purpose and context of the algorithm? | ❖ | We developed a prognostic prediction algorithm (prognostication) of prelabor rupture of membranes (PROM) and an estimation algorithm for the time of delivery. |
|  |  | ❖ | The target population is 12-to-55-years-old females regardless whether they will be pregnant or not, but, precisely, these algorithms are applied for pregnant women only. |
|  |  | ❖ | These algorithms are intended to implement on each visit of a patient to a healthcare provider at any levels (primary, secondary, and tertiary care). |
| Q2. | How good were the data used to train the algorithm? |  |  |
| Q2a. | To what extent were the data accurate and free of bias? | ❖ | We utilized electronic medical records to develop our algorithms. The records were retrospectively selected from ~1% sampling of health insurance holders registered in all healthcare providers affiliated to a nationwide, government-owned health insurance company, the *badan penyelenggara jaminan sosial (BPJS) kesehatan*, in Indonesia. Population of this country covered races of Asian and Austronesian. |
|  |  | ❖ | The health insurance claim data may be inaccurate, and this, in turn, decreases algorithm performances. Nevertheless, given the similar quality of input, we demonstrated our algorithms were in agreement between internal validation set (calibration set) and an external validation set. Therefore, we can expect similar results on average for future application. |
|  |  | ❖ | The selection criteria were visits by the health insurance holders of 12-to-55-years-old females. We excluded visits after delivery. The pregnancy can be the first time or not. |
|  |  | ❖ | From 3,756,292 visits and 1,697,452 insurance holders, we selected 883,376 visits and 219,272 insurance holders. Since these females may be pregnant or not, finally we used data of 170,730 visits and 49,219 women with recorded pregnancy outcome for internal and external validation sets. |
|  |  | ❖ | However, we took into account of the subjects with unknown (missing) outcome for weighting the known outcome (PROM vs. not PROM) over both unknown and known outcomes when developing the algorithms. All predictor data were completed. |
| Q2b. | Were data labelled correctly? | ❖ | Our algorithms were intended to predict what a doctor codes for a patient in the future, whether it will be O42 (ICD-10 code for PROM) or other codes related to the end of pregnancy, and how many days after current visit that code will be encountered. The prediction is not what will be happened in a subject’s physiology and regardless of how accurate the diagnosis of the doctor. This is considerably similar to a consensus adjudication by panels of clinicians that are blind to these predictions, as likely happening in real-world practice. |
| Q2c. | Were the data standardized and interoperable? | ❖ | A clinician only uploads a deidentified, two-column spreadsheet of admission dates (yyyy-mm-dd) and either diagnosis or procedure ICD-10 codes from a patient. This coding system is widely used worldwide. |
| Q3. | Were there sufficient data to train the algorithm? | ❖ | We utilized 86,030 visits (12,011 PROM) to train and 21,506 visits (3002 PROM) to calibrate our algorithms. Using pre-calibrated set with 144 variables to predict PROM and both sets with 60 variables estimate the time of delivery, this is respectively 67 events per variable (EPV) and 1792 EPV. |
|  |  | ❖ | For predicting PROM, we applied a common method, originally used in genomics, to deal with high dimensionality problem. It means using many predictors in relative to the sample size, that may cause poor prediction using data beyond training set. By differential analysis, we reduced candidate predictors from 372 to 144 variables. This analysis and other techniques in deep-insight visible neural network (DI-VNN) algorithm were shown having the best agreement in predicting PROM between development and an external validation set, compared to other state-of-the-art algorithms with 200 EPV. Meanwhile, estimating the time of delivery with 1792 EPV is considerably sufficient. |
| Q4. | How well does the algorithm perform? | ❖ | For predicting PROM, our algorithm has achieved area under receiver operating characteristics curve (AUROC) of 0.73 (95% CI 0.72 to 0.75) using in-sample or internal validation (*n*=21,506) by bootstrapping for 30 times. By systematically searching previous models within the last five years, this is higher than AUROC of 0.67 achieved by the best existing model to predict preterm PROM using maternal factors during first trimester based on a training set only (*n*=10,280).^14^ This is even higher than AUROC of 0.641 achieved by a previous model that uses a biomarker testing (serum alpha-fetoprotein) based on a training set only. |
|  |  | ❖ | We also conducted out-of-sample or external validation. The AUROC is 0.71 (95% CI 0.70 to 0.72) by external random split (23,240 not PROM; 3692 PROM). This subset reflects common situations nationwide. To demonstrate the robustness of the algorithm performance, we also included external validation sets with geographically and/or temporally different populations. But, these may only reflect the worst possible scenario, but may not reflect the common situations. The AUROCs were 0.53 (95% CI 0.52 to 0.54) by external geographical split (13,698 not PROM; 2,120 PROM), 0.56 (95% CI 0.55 to 0.57) by external temporal split (15,484 not PROM; 2736 PROM); and 0.60 (95% CI 0.56 to 0.64) by external geotemporal split (1994 not PROM; 230 PROM). External validation is intended to show the robustness by giving a stress test, but, it cannot be used for generalization considering the smaller size. |
|  |  | ❖ | We recommend 0.14 for initial threshold while determining another threshold based on local data. By this threshold, our model achieved sensitivity of 0.494 (95% CI 0.489 to 0.5) and specificity of 0.816 (95% CI 0.814 to 0.818). Compared to the previous model with AUROC of 0.67, our model achieved sensitivity of ~0.39 which is higher than 0.25 by that model at 90% specificity. The sensitivity and specificity of our model at threshold 0.14 was also slightly higher than those of the previous model using serum alpha-fetoprotein testing. |
|  |  | ❖ | For estimating the time of delivery, if the prediction is PROM, then the time of delivery is precisely estimated (±2.6 weeks) when the result is 6 to 22 weeks’ gestation. If the prediction is not PROM, then the time of delivery is precisely estimated (±2.2 weeks) when the result is 9 to 34 weeks’ gestation. This is internally validated using a large dataset (*n*=107,536) by bootstrapping for 30 times. |
|  |  | ❖ | In a nationwide external validation set (*n*=26,932), the RMSEs are ±9.0 and ±6.3 weeks respectively for PROM and not PROM, as predicted by DI-VNN. In other external validation sets, the RMSEs for the respective predicted outcomes were ±13.4 and ±13.2 weeks in geographical split (*n*=15,818), ±12.6 and ±13.6 weeks in temporal split (*n*=18,220), and ±13.9 and ±16.5 weeks in geotemporal split (*n*=2224). |
|  |  | ❖ | An open-access web application is provided on a public repository (https://predme.app/promtime) to use our algorithms. We invite any investigators to independently validate our algorithms using their own data. |
| Q5. | Is the algorithm transferable to new clinical settings? | ❖ | By the web application, a clinician can recalibrate our algorithms using data with a specific population to choose a threshold that maximize the performances in local setting. While finding the best threshold, a clinician can estimate the algorithm performances at population level given a threshold, including one with the most precise number for positive/negative predictive values (the population-level proportion of the predicted PROM/not PROM that are true rather than false), depending on the prediction outcome and time of delivery of subject case-by-case. |
| Q6. | Are the outputs of the algorithm clinically intelligible? | ❖ | Our algorithm for PROM prediction allows a clinician to critically appraise how the algorithm ends up with the prediction. By the timeline in the report, a clinician can see a chronological visualization of a patient’s medical history used to infer the prediction, including the tendencies of the conditions in the medical history to the event (PROM) and non-event (not PROM). A clinician can also identify the nodes in the network, that include these conditions and see these on the saliency maps. Therefore, a clinician knows to what extend the algorithm taking each condition in the medical history in shifting its decision closer to event or non-event. |
| Q7. | How will this algorithm fit into and complement current workflows? | ❖ | Our algorithms can be applied in a routine antenatal visit when the work pace is hours to days (outpatient, not emergency setting). |
|  |  | ❖ | We recommend the implementation to be the same with ordering a routine laboratory test. If ordered by a clinician in a visit, a unit equipped with access to electronic medical records can execute our algorithms to predict PROM and estimate the time of delivery. The unit needs a staff that has a proficient knowledge in diagnostic or prognostic method evaluation (i.e. adjusting threshold to pursue a particular sensitivity, specificity, or predictive values). This may be a clinical informatician, an existing clinical pathologist, an existing biostatistician, or a trained physician. |
|  |  | ❖ | No significant change may happen on the existing clinical workflow. In addition, a diagnosis or procedure code entry at that visit with the previous ones is needed to execute these algorithms. In our simulation, an execution using a record (medical history) of 20 visits consisting 28 code entries from a patient needs 5.14 minutes (95% CI 5.11 to 5.18 minutes when we repeated for ten times). |
|  |  | ❖ | After executing the algorithms, a staff can use subpopulation-level estimation of the time of delivery given the same predicted time and outcome. If our algorithms are used as a preliminary test to decide if a specific laboratory test is needed, we recommend to set the threshold to get 95% sensitivity. Alternatively, a threshold can be set to pursue the highest positive or negative predictive value (depending on the prediction result), such that the data for population-level performances are still available. |
| Q8. | Has use of the algorithm been shown to improve patient care and outcomes? | ❖ | We call a pilot study, a prospective clinical trial, and a clinical impact study (effectiveness and cost-effectiveness) for these algorithms. Future investigators can simply use an open-access web application we provided to use our algorithms. We expect these algorithms to prevent PROM by early intervention for 50% better than simple guessing. But, such intervention may need further investigations. These algorithms can improve the efficiency of such investigation. If antibiotic is used for the prevention, we expect to reduce the unnecessary use by half than if a treat-all intervention is given. We also expect these algorithms can reduce a need for a specific laboratory test requiring high-resource setting by 95%. |
| Q9. | Could the algorithm cause patient harm? | ❖ | Unnecessary use of antibiotic may be induced by false positive of these algorithms. A clinician need to assess carefully the need of antibiotic based on conventional guidelines beyond consideration related to the result of these algorithms. A clinician should also advise a patient to watch the delivery within the population-level estimated time although the prediction gives a negative result for PROM. The negative result may cause pregnancy monitoring off-guard. To prevent the harms, repeating the prediction more frequent are needed toward the end of pregnancy. A clinician should also continuously questioning counterintuitive internal properties of a prediction by our algorithms. |
| Q10. | Does the algorithm raise ethical, legal or social concerns? | ❖ | We consider ethical, legal, and social issues when developing these algorithms and the recommended setting to apply the algorithms. |
|  |  | ❖ | No privacy issue will be raised since this algorithm only needs two-column medical history (admission date and ICD-10 code) without identity or even demographic information. |
|  |  | ❖ | Our algorithms are also equitable and inclusive because of no need of high resources, including those for the electronic medical record system. |
|  |  | ❖ | Apart of availability of an independent validation and a clinical trial for any algorithms, a local recalibration of our algorithms are warranted, because a perfect consistency of predictive performances is difficult if not impossible. A responsibility to ensure the recalibration is on the healthcare provider. A competent staff for adjusting the local or individual threshold (i.e. a clinical informatician, a clinical pathologist, an existing biostatistician, or a trained physician) is also needed. Failed to provide both efforts may raise legal issues using these algorithms. |
|  |  | ❖ | Non-commercial use is permitted. Local recalibration and a competent is preferable before these algorithms are commercialized in a healthcare provider. Any commercialization need to apply permission to the developer of these algorithms. |
|  |  | ❖ | Social issue may be raised if the commercialization is applied, especially in non-insured healthcare service. Worthiness of the prediction over the cost is needed to assess by a patient equipped with sufficient information. A patient education is needed when offering a prediction service by these algorithms. Sufficient information about the limitations of these algorithms should be given. |

eTable 4. The preferred reporting items for systematic reviews and meta-analyses (PRISMA) 2020 expanded checklist

| Section and topic | Item # | Elements recommended for reporting | |
| --- | --- | --- | --- |
| Eligibility criteria | 5 | ❖ | Following PICOTS framework, the eligibility criteria were: (1) Population, either pregnant or non-pregnant women without specializing the medical conditions; (2) Index, the best prediction model in this study; (3) Comparator, prediction models or rules; (4) Outcome, either preterm or term PROM as a binary outcome (event or non-event) with 20 EPV or more; (5) Time, prognostic prediction from days to weeks; and (6) Setting, either primary care or hospital patient. |
|  |  | ❖ | We excluded any article types beyond original article, including conference abstract but not the full paper. |
|  |  | ❖ | The studies were not grouped since no synthesis was conducted. |
| Information sources | 6 | ❖ | We searched the studies up to April 3^rd^ 2021. |
|  |  | ❖ | Bibliographic databases were searched, which were PubMed, Scopus, and Web of Science. |
| Search strategy | 7 | ❖ | The keyword was ‘prelabor rupture of membranes prediction’. |
|  |  | ❖ | In PubMed, we used the keyword and filtered by article type (i.e. journal article), publication date (i.e. five years), and language (i.e. English). |
|  |  | ❖ | In Scopus, we used the keyword within article title, abstract, and keywords, published from 2016 to present. Then the results were limited by document type (i.e. article) and language (i.e. English). The configuration resulted in this syntax: TITLE-ABS-KEY (prelabor AND rupture AND of AND membranes AND prediction) AND PUBYEAR > 2015 AND (LIMIT-TO (DOCTYPE, "ar")) AND (LIMIT-TO (LANGUAGE, "English")) |
|  |  | ❖ | In Web of Science, we used the keyword for topic with timespan of last five years. Then the results were refined by document types (i.e. article, early access, and proceedings paper) and language (i.e. English). |
|  |  | ❖ | For each database, we also applied dual filter, which is, filtering review and exclude the results from the existing ones. This is because the search platform still included review even though the documents were already filtered for only original articles. |
| Selection process | 8 | ❖ | HS searched for the literatures and loosely filtered these by title and abstract. Then, HS manually assessed the eligibility by the full texts and identified the ambiguous ones. Independently, HS and YWW assessed the ambiguous studies. If no consensus made between HS and YWW, final decision was made by ECYS. |
| Data collection process | 9 | ❖ | HS collected data from each full text. |
| Data items (outcomes) | 10a | ❖ | We only extracted a sensitivity and a specificity of the best model with the most similar outcome definition from each study to that of our study. A summary ROC curve was constructed using the sensitivities and specificities, overlaid by those of our models for PROM prediction. |
|  |  | ❖ | The AUROCs were also included to compare with the models in this study (with or without 95% CI). If not reported, the AUROC was inferred by trapezoidal rule utilizing the sensitivity (Sn.) and specificity (Sp.): (*Sn*×(1-*Sp*))/2+*Sn*×*Sp*+((1-*Sn*)×*Sp*)/2. |
| Data items (other variables) | 10b | ❖ | We also extracted the study design, population, setting, outcome definition, sample size, including details on events and non-events, number of candidate predictors, EPV, predictors in the best final model, and the most recommended validation techniques (external over internal validation; bootstrapping over cross validation, or cross validation over test split). |
| Effect measures | 12 | ❖ | The plots of sensitivities and specificities would overlay those of our models for PROM prediction. We also plotted the sensitivity and specificity for each of our models at optimum threshold, on each ROC curve. |
|  |  | ❖ | AUROCs were compared with the models in this study (with or without 95% CI). |

eTable 5. Comparable models to evaluate the success criteria

| Study/model | Metrics (95% CI) | Other variables | |
| --- | --- | --- | --- |
| El-Achi, et al. | Sensitivity: | ❖ | Study design: retrospective cohort |
| LR using maternal factors | 0.25 | ❖ | Population: pregnant women at 11–<14 weeks’ gestation, singleton, and excluded if loss to follow-up, termination of pregnancy, or carrying a multiple pregnancy. |
|  |  | ❖ | Setting: hospital |
|  | Specificity: | ❖ | Outcome definition: event (preterm PROM) and non-event (not preterm PROM) |
|  | 0.9 | ❖ | Sample size: event (*n*=144) and non-event (*n*=10,136) |
|  |  | ❖ | Predictors in the best final model: parity (nulliparous/parous), pre-existing diabetes mellitus (no/ type 1/type 2), maternal age (<25/25–≤30/30–≤35/>35; years), and body mass index (<20/20–≤25/25–≤30/>30; kg/m^2^) |
|  | AUROC: | ❖ | The most recommended validation technique: no internal validation |
|  | 0.667 |  |  |
| Başbuğ, et al. | Sensitivity: | ❖ | Study design: retrospective cohort |
| Prediction rule using a serum biomarker | 0.419 | ❖ | Population: pregnant women at 16–20 weeks’ gestation, singleton, underwent triple blood testing (serum alpha-fetoprotein, human chorionic gonadotropin, and unconjugated estriol), and excluded if congenital anomalies, hematoma, loss to follow-up, or carrying a multiple pregnancy. |
|  |  | ❖ | Setting: primary care |
|  | Specificity: | ❖ | Outcome definition: event (preterm PROM) and non-event (not preterm PROM) |
|  | 0.863 | ❖ | Sample size: event (*n*=31) and non-event (*n*=1146) |
|  |  | ❖ | Predictors in the best final model: serum alpha-fetoprotein |
|  | AUROC: * | ❖ | The most recommended validation technique: no internal validation |
|  | 0.641 |  |  |
| * Inferred from sensitivity and specificity | | |  |

| Review step | Total (*n*) | Record | |
| --- | --- | --- | --- |
| 1. a. PubMed | 186 | [1] | Good clinical practice advice: Prediction of preterm labor and preterm premature rupture of membranes, Int J Gynaecol Obstet 144 (2019), 340-346. |
|  |  | [2] | K. Abdel-Malek, M.A. El-Halwagi, B.E. Hammad, O. Azmy, O. Helal, M. Eid, and M. Abdel-Rasheed, Role of maternal serum ferritin in prediction of preterm labour, J Obstet Gynaecol 38 (2018), 222-225. |
|  |  | [3] | W.E.t. Ackerman, I.A. Buhimschi, D. Brubaker, S. Maxwell, K.M. Rood, M.R. Chance, H. Jing, S. Mesiano, and C.S. Buhimschi, Integrated microRNA and mRNA network analysis of the human myometrial transcriptome in the transition from quiescence to labor, Biol Reprod 98 (2018), 834-845. |
|  |  | [4] | S. Alla, A. Ramseyer, J.R. Whittington, S. Peeples, S.T. Ounpraseuth, and E.F. Magann, Maternal features at time of preterm prelabor rupture of membranes and short-term neonatal outcomes, J Matern Fetal Neonatal Med (2020), 1-7. |
|  |  | [5] | R.M. Alvarez, I. Biliatis, A. Rockall, E. Papadakou, S.A. Sohaib, N.M. deSouza, J. Butler, M. Nobbenhuis, D. Barton, J.H. Shepherd, and T. Ind, MRI measurement of residual cervical length after radical trachelectomy for cervical cancer and the risk of adverse pregnancy outcomes: a blinded imaging analysis, Bjog 125 (2018), 1726-1733. |
|  |  | [6] | N. Aracic, I. Stipic, I. Jakus Alujevic, P. Poljak, and M. Stipic, The value of ultrasound measurement of cervical length and parity in prediction of cesarean section risk in term premature rupture of membranes and unfavorable cervix, J Perinat Med 45 (2017), 99-104. |
|  |  | [7] | N. Asadi, A. Faraji, A. Keshavarzi, M. Akbarzadeh-Jahromi, and S. Yoosefi, Predictive value of procalcitonin, C-reactive protein, and white blood cells for chorioamnionitis among women with preterm premature rupture of membranes, Int J Gynaecol Obstet 147 (2019), 83-88. |
|  |  | [8] | M.T. Aung, Y. Yu, K.K. Ferguson, D.E. Cantonwine, L. Zeng, T.F. McElrath, S. Pennathur, B. Mukherjee, and J.D. Meeker, Prediction and associations of preterm birth and its subtypes with eicosanoid enzymatic pathways and inflammatory markers, Sci Rep 9 (2019), 17049. |
|  |  | [9] | A. Aviram, P. Quaglietta, C. Warshafsky, A. Zaltz, E. Weiner, N. Melamed, E. Ng, J. Barrett, and S. Ronzoni, Utility of ultrasound assessment in management of pregnancies with preterm prelabor rupture of membranes, Ultrasound Obstet Gynecol 55 (2020), 806-814. |
|  |  | [10] | U. Baboolall, Y. Zha, X. Gong, D.R. Deng, F. Qiao, and H. Liu, Variations of plasma D-dimer level at various points of normal pregnancy and its trends in complicated pregnancies: A retrospective observational cohort study, Medicine (Baltimore) 98 (2019), e15903. |
|  |  | [11] | F. Bahlmann and A. Al Naimi, Using the angiogenic factors sFlt-1 and PlGF with Doppler ultrasound of the uterine artery for confirming preeclampsia, Arch Gynecol Obstet 294 (2016), 1133-1139. |
|  |  | [12] | A.R. Ballard, L.H. Mallett, J.E. Pruszynski, and J.B. Cantey, Chorioamnionitis and subsequent bronchopulmonary dysplasia in very-low-birth weight infants: a 25-year cohort, J Perinatol 36 (2016), 1045-1048. |
|  |  | [13] | S.V. Barinov, G.C. Di Renzo, A.A. Belinina, O.V. Koliado, and O.V. Remneva, Clinical and biochemical markers of spontaneous preterm birth in singleton and multiple pregnancies, J Matern Fetal Neonatal Med (2021), 1-6. |
|  |  | [14] | D. Başbuğ, A. Başbuğ, and C. Gülerman, Is unexplained elevated maternal serum alpha-fetoprotein still important predictor for adverse pregnancy outcome?, Ginekol Pol 88 (2017), 325-330. |
|  |  | [15] | E. Baser, D. Aydogan Kirmizi, D. Ulubas Isik, S. Ozdemirci, T. Onat, E. Serdar Yalvac, N. Demirel, and O. Moraloglu Tekin, The effects of latency period in PPROM cases managed expectantly, J Matern Fetal Neonatal Med 33 (2020), 2274-2283. |
|  |  | [16] | J.B. Beavers, S. Bai, J. Perry, J. Simpson, and S. Peeples, Implementation and Evaluation of the Early-Onset Sepsis Risk Calculator in a High-Risk University Nursery, Clin Pediatr (Phila) 57 (2018), 1080-1085. |
|  |  | [17] | M.A. Belfort, Predicting premature preterm rupture of the membranes after fetal surgery, Bjog 125 (2018), 1293. |
|  |  | [18] | Z. Bouzari, R. Shahhosseini, M. Mohammadnetaj, S. Barat, S. Yazdani, and K. Hajian-Tilaki, Vaginal discharge concentrations of β-human chorionic gonadotropin, creatinine, and urea for the diagnosis of premature rupture of membranes, Int J Gynaecol Obstet 141 (2018), 97-101. |
|  |  | [19] | A.P. Burgess, J. Katz, J. Pessolano, J. Ponterio, M. Moretti, and N.A. Lakhi, Determination of antepartum and intrapartum risk factors associated with neonatal intensive care unit admission, J Perinat Med 44 (2016), 589-596. |
|  |  | [20] | J. Caloone, M. Rabilloud, F. Boutitie, A. Traverse-Glehen, F. Allias-Montmayeur, L. Denis, C. Boisson-Gaudin, I.J. Hot, P. Guerre, M. Cortet, and C. Huissoud, Accuracy of several maternal seric markers for predicting histological chorioamnionitis after preterm premature rupture of membranes: a prospective and multicentric study, Eur J Obstet Gynecol Reprod Biol 205 (2016), 133-140. |
|  |  | [21] | M.T. Canda, N. Demir, and O. Sezer, Fetal Nasal Bone Length as a Novel Marker for Prediction of Adverse Perinatal Outcomes in the First-Trimester of Pregnancy, Balkan Med J 34 (2017), 127-131. |
|  |  | [22] | W. Cao, C. Luo, M. Lei, M. Shen, W. Ding, M. Wang, M. Song, J. Ge, and Q. Zhang, Development and Validation of a Dynamic Nomogram to Predict the Risk of Neonatal White Matter Damage, Front Hum Neurosci 14 (2020), 584236. |
|  |  | [23] | D. Carola, M. Vasconcellos, A. Sloane, D. McElwee, C. Edwards, J. Greenspan, and Z.H. Aghai, Utility of Early-Onset Sepsis Risk Calculator for Neonates Born to Mothers with Chorioamnionitis, J Pediatr 195 (2018), 48-52.e41. |
|  |  | [24] | O. Cetin, Z.D. Aydın, F.F. Verit, A.G. Zebitay, E. Karaman, S. Elasan, M. Turfan, and O. Yucel, Is Maternal Blood Procalcitonin Level a Reliable Predictor for Early Onset Neonatal Sepsis in Preterm Premature Rupture of Membranes?, Gynecol Obstet Invest 82 (2017), 163-169. |
|  |  | [25] | H.H. Cha, J.M. Kim, H.M. Kim, M.J. Kim, G.O. Chong, and W.J. Seong, Association between gestational age at delivery and lymphocyte-monocyte ratio in the routine second trimester complete blood cell count, Yeungnam Univ J Med 38 (2021), 34-38. |
|  |  | [26] | P. Chaemsaithong, R. Romero, N. Docheva, N. Chaiyasit, G. Bhatti, P. Pacora, S.S. Hassan, L. Yeo, and O. Erez, Comparison of rapid MMP-8 and interleukin-6 point-of-care tests to identify intra-amniotic inflammation/infection and impending preterm delivery in patients with preterm labor and intact membranes(), J Matern Fetal Neonatal Med 31 (2018), 228-244. |
|  |  | [27] | N. Chaiyasit, R. Romero, P. Chaemsaithong, N. Docheva, G. Bhatti, J.P. Kusanovic, Z. Dong, L. Yeo, P. Pacora, S.S. Hassan, and O. Erez, Clinical chorioamnionitis at term VIII: a rapid MMP-8 test for the identification of intra-amniotic inflammation, J Perinat Med 45 (2017), 539-550. |
|  |  | [28] | I. Chandra and L. Sun, Third trimester preterm and term premature rupture of membranes: Is there any difference in maternal characteristics and pregnancy outcomes?, J Chin Med Assoc 80 (2017), 657-661. |
|  |  | [29] | S. Chaudhury, J. Saqibuddin, R. Birkett, K. Falcon-Girard, M. Kraus, L.M. Ernst, W. Grobman, and K.K. Mestan, Variations in Umbilical Cord Hematopoietic and Mesenchymal Stem Cells With Bronchopulmonary Dysplasia, Front Pediatr 7 (2019), 475. |
|  |  | [30] | A.L. Chen, I.T. Goldfarb, A.O. Scourtas, and D.J. Roberts, The histologic evolution of revealed, acute abruptions, Hum Pathol 67 (2017), 187-197. |
|  |  | [31] | K.W. Cheung, L.N. Tan, M.T.Y. Seto, S. Moholkar, G. Masson, and M.D. Kilby, Prenatal Diagnosis, Management, and Outcome of Fetal Subdural Haematoma: A Case Report and Systematic Review, Fetal Diagn Ther 46 (2019), 285-295. |
|  |  | [32] | H.Y. Cho, I. Jung, J.Y. Kwon, S.J. Kim, Y.W. Park, and Y.H. Kim, The Delta Neutrophil Index as a predictive marker of histological chorioamnionitis in patients with preterm premature rupture of membranes: A retrospective study, PLoS One 12 (2017), e0173382. |
|  |  | [33] | T. Cobo, V. Aldecoa, F. Figueras, A. Herranz, S. Ferrero, M. Izquierdo, C. Murillo, R. Amoedo, C. Rueda, J. Bosch, R.J. Martínez-Portilla, E. Gratacós, and M. Palacio, Development and validation of a multivariable prediction model of spontaneous preterm delivery and microbial invasion of the amniotic cavity in women with preterm labor, Am J Obstet Gynecol 223 (2020), 421.e421-421.e414. |
|  |  | [34] | T. Cobo, J. Munrós, J. Ríos, J. Ferreri, F. Migliorelli, N. Baños, E. Gratacós, and M. Palacio, Contribution of Amniotic Fluid along Gestation to the Prediction of Perinatal Mortality in Women with Early Preterm Premature Rupture of Membranes, Fetal Diagn Ther 43 (2018), 105-112. |
|  |  | [35] | Y. Dabi, S. Nedellec, C. Bonneau, B. Trouchard, R. Rouzier, and A. Benachi, Clinical validation of a model predicting the risk of preterm delivery, PLoS One 12 (2017), e0171801. |
|  |  | [36] | C.B. Dartibale, N.S. Uchimura, L. Nery, A.P. Schumeish, L.Y.T. Uchimura, R.G. Santana, and T.T. Uchimura, Qualitative Determination of Human Chorionic Gonadotropin in Vaginal Washings for the Early Diagnosis of Premature Rupture of Fetal Membranes, Rev Bras Ginecol Obstet 39 (2017), 317-321. |
|  |  | [37] | G. Daskalakis, P. Fotinopoulos, V. Pergialiotis, M. Theodora, P. Antsaklis, M. Sindos, N. Papantoniou, and D. Loutradis, Delayed interval delivery of the second twin in a woman with altered markers of inflammation, BMC Pregnancy Childbirth 18 (2018), 206. |
|  |  | [38] | C. Davies, G. Segre, A. Estradé, J. Radua, A. De Micheli, U. Provenzani, D. Oliver, G. Salazar de Pablo, V. Ramella-Cravaro, M. Besozzi, P. Dazzan, M. Miele, G. Caputo, C. Spallarossa, G. Crossland, A. Ilyas, G. Spada, P. Politi, R.M. Murray, P. McGuire, and P. Fusar-Poli, Prenatal and perinatal risk and protective factors for psychosis: a systematic review and meta-analysis, Lancet Psychiatry 7 (2020), 399-410. |
|  |  | [39] | D.A. De Silva, S. Lisonkova, P. von Dadelszen, A.R. Synnes, and L.A. Magee, Timing of delivery in a high-risk obstetric population: a clinical prediction model, BMC Pregnancy Childbirth 17 (2017), 202. |
|  |  | [40] | N. Dorfeuille, V. Morin, A. Tétu, S. Demers, G. Laforest, K. Gouin, B. Piedboeuf, and E. Bujold, Vaginal Fluid Inflammatory Biomarkers and the Risk of Adverse Neonatal Outcomes in Women with PPROM, Am J Perinatol 33 (2016), 1003-1007. |
|  |  | [41] | R.D. Ducci, P.J. Lorenzoni, C.S. Kay, L.C. Werneck, and R.H. Scola, Clinical follow-up of pregnancy in myasthenia gravis patients, Neuromuscul Disord 27 (2017), 352-357. |
|  |  | [42] | J.R. Duncan, K.M. Dorsett, M.M. Aziz, Z. Bursac, M.A. Cleves, A.J. Talati, M.H. Schenone, N.L. Meyer, and G. Mari, Estimated fetal weight and severe neonatal outcomes in preterm prelabor rupture of membranes, J Perinat Med 48 (2020), 687-693. |
|  |  | [43] | J.R. Duncan, K.M. Dorsett, G. Vilchez, M.H. Schenone, and G. Mari, Uterine artery pulsatility index for the prediction of obstetrical complications in preterm prelabor rupture of membranes, J Matern Fetal Neonatal Med (2019), 1-4. |
|  |  | [44] | J.R. Duncan, K. Leavitt, K. Duncan, and G. Vilchez, Detection of small for gestational age in preterm prelabor rupture of membranes by Hadlock versus the Fetal Medicine Foundation growth charts, Obstet Gynecol Sci (2021). |
|  |  | [45] | J.R. Duncan, P. Sawangkum, E.A. Hoover, M.M. Aziz, and G. Vilchez, Birthweight and Apgar at 5 minutes of life for the prediction of severe neonatal outcomes in preterm prelabor rupture of membranes, J Matern Fetal Neonatal Med (2020), 1-5. |
|  |  | [46] | J.R. Duncan, C. Schenone, K.M. Dorset, P.J. Goedecke, A.M. Tobiasz, N.L. Meyer, and M.H. Schenone, Estimated fetal weight accuracy in pregnancies with preterm prelabor rupture of membranes by the Hadlock method, J Matern Fetal Neonatal Med (2020), 1-5. |
|  |  | [47] | J.R. Duncan, A.M. Tobiasz, K.M. Dorsett, M.M. Aziz, R.E. Thompson, Z. Bursac, A.J. Talati, G. Mari, and M.H. Schenone, Fetal pulmonary artery acceleration/ejection time prognostic accuracy for respiratory complications in preterm prelabor rupture of membranes, J Matern Fetal Neonatal Med 33 (2020), 2054-2058. |
|  |  | [48] | B. Dundar, B. Dincgez Cakmak, G. Ozgen, F.N. Tasgoz, T. Guclu, and G. Ocakoglu, Platelet indices in preterm premature rupture of membranes and their relation with adverse neonatal outcomes, J Obstet Gynaecol Res 44 (2018), 67-73. |
|  |  | [49] | V. El-Achi, F. Park, C. O'Brien, J. Tooher, and J. Hyett, Does low dose aspirin prescribed for risk of early onset preeclampsia reduce the prevalence of preterm prelabor rupture of membranes?, J Matern Fetal Neonatal Med 34 (2021), 618-623. |
|  |  | [50] | E. Elçi, G. Güneş Elçi, and S. Sayan, Comparison of the accuracy and reliability of the AmniSure, AMNIOQUICK, and AL-SENSE tests for early diagnosis of premature rupture of membranes, Int J Gynaecol Obstet 149 (2020), 93-97. |
|  |  | [51] | G.U. Eleje, E.C. Ezugwu, A.C. Eke, J.I. Ikechebelu, C.O. Ezeama, I.U. Ezebialu, N.O. Ojiegbe, C.C. Obiora, C.I. Okafor, G.O. Udigwe, B.O. Nwosu, and F.O. Ezugwu, Accuracy and response time of dual biomarker model of insulin-like growth factor binding protein-1/ alpha fetoprotein (Amnioquick duo+) in comparison to placental alpha-microglobulin-1 test in diagnosis of premature rupture of membranes, J Obstet Gynaecol Res 43 (2017), 825-833. |
|  |  | [52] | G.U. Eleje, E.C. Ezugwu, A.C. Eke, J.I. Ikechebelu, C.C. Obiora, N.O. Ojiegbe, I.U. Ezebialu, C.O. Ezeama, B.O. Nwosu, G.O. Udigwe, C.I. Okafor, and F.O. Ezugwu, Comparison of the duo of insulin-like growth factor binding protein-1/alpha fetoprotein (Amnioquick duo+®) and traditional clinical assessment for diagnosing premature rupture of fetal membranes, J Perinat Med 45 (2017), 105-112. |
|  |  | [53] | G.U. Eleje, E.C. Ezugwu, I.U. Ezebialu, N.O. Ojiegbe, R.O. Egeonu, C.C. Obiora, C.G. Okafor, J.I. Ikechebelu, and A.C. Eke, Performance indices of AmnioQuick Duo+ versus placental α-microglobulin-1 tests for women with prolonged premature rupture of membranes, Int J Gynaecol Obstet 144 (2019), 180-186. |
|  |  | [54] | G.U. Eleje, C.O. Ukah, I.V. Onyiaorah, E.C. Ezugwu, E.O. Ugwu, S.R. Ohayi, L.I. Eleje, R.O. Egeonu, I.U. Ezebialu, C.C. Obiora, J.T. Enebe, L.O. Ajah, C.G. Okafor, C.C. Okoro, A.O. Asogwa, D.K. Ogbuokiri, J.I. Ikechebelu, and A.C. Eke, Diagnostic value of Chorioquick for detecting chorioamnionitis in women with premature rupture of membranes, Int J Gynaecol Obstet 149 (2020), 98-105. |
|  |  | [55] | H. Erfani, S. Haeri, S.A. Shainker, A.F. Saad, R. Ruano, T.N. Dunn, A. Rezaei, S. Aalipour, A.A. Nassr, A.A. Shamshirsaz, M. Vaughn, W. Lindsley, M.H. Spiel, S.A. Shazly, E.R. Ibirogba, S.L. Clark, G.R. Saade, M.A. Belfort, and A.A. Shamshirsaz, Vasa previa: a multicenter retrospective cohort study, Am J Obstet Gynecol 221 (2019), 644.e641-644.e645. |
|  |  | [56] | S. Esin, M. Hayran, Y.A. Tohma, M. Guden, I. Alay, D. Esinler, S. Yalvac, and O. Kandemir, Estimation of fetal weight by ultrasonography after preterm premature rupture of membranes: comparison of different formulas, J Perinat Med 45 (2017), 253-266. |
|  |  | [57] | F.B. Flosi, F.C. da Silva, G.R. de Jesús, L.G.C. Velarde, and R.A.M. de Sá, Assessment of Fetal Lung Maturity Using Quantitative Ultrasound Analysis in Patients with Prelabor Rupture of Membranes, Fetal Diagn Ther 47 (2020), 636-641. |
|  |  | [58] | K. Fuwa, M. Seki, Y. Hirata, I. Yanagihara, Y. Nakura, C. Takano, K. Kuroda, and S. Hayakawa, Rapid and simple detection of Ureaplasma species from vaginal swab samples using a loop-mediated isothermal amplification method, Am J Reprod Immunol 79 (2018). |
|  |  | [59] | A. Gámez-Varela, M. Martínez-Rodríguez, H. López-Briones, J. Luna-García, E. Chávez-González, R. Villalobos-Gómez, E. Hernandez-Andrade, and R. Cruz-Martínez, Preoperative Cervical Length Predicts the Risk of Delivery within One Week after Pleuroamniotic Shunt in Fetuses with Severe Hydrothorax, Fetal Diagn Ther (2021), 1-7. |
|  |  | [60] | C. Gezer, A. Ekin, C. Golbasi, C. Kocahakimoglu, U. Bozkurt, A. Dogan, U. Solmaz, H. Golbasi, and C.E. Taner, Use of urea and creatinine levels in vaginal fluid for the diagnosis of preterm premature rupture of membranes and delivery interval after membrane rupture, J Matern Fetal Neonatal Med 30 (2017), 772-778. |
|  |  | [61] | L.L. Gievers, J. Sedler, C.A. Phillipi, D. Dukhovny, J. Geddes, P. Graven, B. Chan, and S. Khaki, Implementation of the sepsis risk score for chorioamnionitis-exposed newborns, J Perinatol 38 (2018), 1581-1587. |
|  |  | [62] | M. Gioan, F. Fenollar, A. Loundou, J.P. Menard, J. Blanc, C. D'Ercole, and F. Bretelle, Development of a nomogram for individual preterm birth risk evaluation, J Gynecol Obstet Hum Reprod 47 (2018), 545-548. |
|  |  | [63] | P. Girón de Velasco-Sada, I. Falces-Romero, I. Quiles-Melero, A. García-Perea, and J. Mingorance, Evaluation of two real time PCR assays for the detection of bacterial DNA in amniotic fluid, J Microbiol Methods 144 (2018), 107-110. |
|  |  | [64] | L. Goodfellow, A. Care, A. Sharp, J. Ivandic, B. Poljak, D. Roberts, and Z. Alfirevic, Effect of QUiPP prediction algorithm on treatment decisions in women with a previous preterm birth: a prospective cohort study, Bjog 126 (2019), 1569-1575. |
|  |  | [65] | S. Gupta, H.S. Gaikwad, B. Nath, and A. Batra, Can vitamin C and interleukin 6 levels predict preterm premature rupture of membranes: evaluating possibilities in North Indian population, Obstet Gynecol Sci 63 (2020), 432-439. |
|  |  | [66] | R.B. Helmig and J.B. Gertsen, Diagnostic accuracy of polymerase chain reaction for intrapartum detection of group B streptococcus colonization, Acta Obstet Gynecol Scand 96 (2017), 1070-1074. |
|  |  | [67] | A. Hesson and E. Langen, Outcomes in oligohydramnios: the role of etiology in predicting pulmonary morbidity/mortality, J Perinat Med 46 (2018), 948-950. |
|  |  | [68] | L. Hiersch, E. Krispin, A. Aviram, M. Mor-Shacham, R. Gabbay-Benziv, Y. Yogev, and E. Ashwal, Predictors for prolonged interval from premature rupture of membranes to spontaneous onset of labor at term, J Matern Fetal Neonatal Med 30 (2017), 1465-1470. |
|  |  | [69] | Y. Horikoshi, C. Yaguchi, N. Furuta-Isomura, T. Itoh, K. Kawai, T. Oda, M. Matsumoto, Y. Kohmura-Kobayashi, N. Tamura, T. Uchida, N. Kanayama, and H. Itoh, Gross appearance of the fetal membrane on the placental surface is associated with histological chorioamnionitis and neonatal respiratory disorders, PLoS One 15 (2020), e0242579. |
|  |  | [70] | T. Horinouchi, T. Yoshizato, Y. Kozuma, T. Shinagawa, M. Muto, T. Yamasaki, D. Hori, and K. Ushijima, Prediction of histological chorioamnionitis and neonatal and infantile outcomes using procalcitonin in the umbilical cord blood and amniotic fluid at birth, J Obstet Gynaecol Res 44 (2018), 630-636. |
|  |  | [71] | K. Hughes, S. Sim, A. Roman, K. Michalak, S. Kane, and P. Sheehan, Outcomes and predictive tests from a dedicated specialist clinic for women at high risk of preterm labour: A ten year audit, Aust N Z J Obstet Gynaecol 57 (2017), 405-411. |
|  |  | [72] | K.M. Hughes, S.C. Kane, T.P. Haines, and P.M. Sheehan, Cervical length surveillance for predicting spontaneous preterm birth in women with uterine anomalies: A cohort study, Acta Obstet Gynecol Scand 99 (2020), 1519-1526. |
|  |  | [73] | A. Husain, A. Naseem, S. Anjum, S. Imran, M. Arifuzzaman, and S.O. Adil, Predictability of intrapartum cardiotocography with meconium stained liquor and its correlation with perinatal outcome, J Pak Med Assoc 68 (2018), 1014-1018. |
|  |  | [74] | I. Igbinosa, F.A. Moore, 3rd, C. Johnson, and J.E. Block, Comparison of rapid immunoassays for rupture of fetal membranes, BMC Pregnancy Childbirth 17 (2017), 128. |
|  |  | [75] | M. Jain, A. Bang, A. Tiwari, and S. Jain, Prediction of significant hyperbilirubinemia in term neonates by early non-invasive bilirubin measurement, World J Pediatr 13 (2017), 222-227. |
|  |  | [76] | P. Janku, M. Kacerovsky, B. Zednikova, C. Andrys, M. Kolackova, M. Drahosova, L. Pliskova, H. Zemlickova, R. Gerychova, O. Simetka, P. Matlak, B. Jacobsson, and I. Musilova, Pentraxin 3 in Noninvasively Obtained Cervical Fluid Samples from Pregnancies Complicated by Preterm Prelabor Rupture of Membranes, Fetal Diagn Ther 46 (2019), 402-410. |
|  |  | [77] | F. Jochum, C. Le Ray, P. Blanc-Petitjean, B. Langer, N. Meyer, F. Severac, and N. Sananes, Externally Validated Score to Predict Cesarean Delivery After Labor Induction With Cervi Ripening, Obstet Gynecol 134 (2019), 502-510. |
|  |  | [78] | M. Julien, L. N'Guema, A. Bouzerara, B. Toro, E. Lecarpentier, and J. Guibourdenche, Premature rupture of the membranes: analytical evaluation of diagnostic tests, Ann Biol Clin (Paris) 76 (2018), 300-306. |
|  |  | [79] | S. Jwala, T.L. Tran, C. Terenna, A. McGregor, J. Andrel, B.E. Leiby, J.K. Baxter, and V. Berghella, Evaluation of additive effect of quantitative fetal fibronectin to cervical length for prediction of spontaneous preterm birth among asymptomatic low-risk women, Acta Obstet Gynecol Scand 95 (2016), 948-955. |
|  |  | [80] | M. Kacerovsky, I. Musilova, T. Bestvina, M. Stepan, T. Cobo, and B. Jacobsson, Preterm Prelabor Rupture of Membranes between 34 and 37 Weeks: A Point-of-Care Test of Vaginal Fluid Interleukin-6 Concentrations for a Noninvasive Detection of Intra-Amniotic Inflammation, Fetal Diagn Ther 43 (2018), 175-183. |
|  |  | [81] | M. Kaneko, M. Sato, K. Ogasawara, T. Imamura, K. Hashimoto, N. Momoi, and M. Hosoya, Serum cytokine concentrations, chorioamnionitis and the onset of bronchopulmonary dysplasia in premature infants, J Neonatal Perinatal Med 10 (2017), 147-155. |
|  |  | [82] | H. Kansu-Celik, Y. Tasci, B.K. Karakaya, M. Cinar, T. Candar, and G.S. Caglar, Maternal serum advanced glycation end products level as an early marker for predicting preterm labor/PPROM: a prospective preliminary study, J Matern Fetal Neonatal Med 32 (2019), 2758-2762. |
|  |  | [83] | V. Kathir, D. Maurya, and A. Keepanasseril, Transvaginal sonographic assessment of cervix in prediction of admission to delivery interval in preterm premature rupture of membranes, J Matern Fetal Neonatal Med 31 (2018), 2717-2720. |
|  |  | [84] | T. Kawakita, U.M. Reddy, J.C. Huang, T.C. Auguste, D. Bauer, and R.T. Overcash, Externally Validated Prediction Model of Vaginal Delivery After Preterm Induction With Unfavorable Cervix, Obstet Gynecol 136 (2020), 716-724. |
|  |  | [85] | B. Kaya, U. Turhan, S. Sezer, S. Kaya, İ. Dağ, and A. Tayyar, Maternal serum galectin-1 and galectin-3 levels in pregnancies complicated with preterm prelabor rupture of membranes, J Matern Fetal Neonatal Med 33 (2020), 861-868. |
|  |  | [86] | G. Kayem, F. Batteux, N. Girard, T. Schmitz, M. Willaime, F. Maillard, P.H. Jarreau, and F. Goffinet, Predictive value of vaginal IL-6 and TNFα bedside tests repeated until delivery for the prediction of maternal-fetal infection in cases of premature rupture of membranes, Eur J Obstet Gynecol Reprod Biol 211 (2017), 8-14. |
|  |  | [87] | A. Kesrouani, E. Chalhoub, E. El Rassy, M. Germanos, A. Khazzaka, J. Rizkallah, E. Attieh, and N. Aouad, Prediction of preterm delivery by second trimester inflammatory biomarkers in the amniotic fluid, Cytokine 85 (2016), 67-70. |
|  |  | [88] | H.J. Kim, K.H. Park, Y.M. Kim, E. Joo, K. Ahn, and S. Shin, A protein microarray analysis of amniotic fluid proteins for the prediction of spontaneous preterm delivery in women with preterm premature rupture of membranes at 23 to 30 weeks of gestation, PLoS One 15 (2020), e0244720. |
|  |  | [89] | Y.M. Kim, J.Y. Park, J.H. Sung, S.J. Choi, S.Y. Oh, C.R. Roh, and J.H. Kim, Predicting factors for success of vaginal delivery in preterm induction with prostaglandin E(2), Obstet Gynecol Sci 60 (2017), 163-169. |
|  |  | [90] | V. Kiver, V. Boos, A. Thomas, W. Henrich, and A. Weichert, Perinatal outcomes after previable preterm premature rupture of membranes before 24 weeks of gestation, J Perinat Med 46 (2018), 555-565. |
|  |  | [91] | S.Y. Kook, K.H. Park, J.A. Jang, Y.M. Kim, H. Park, and S.J. Jeon, Vitamin D-binding protein in cervicovaginal fluid as a non-invasive predictor of intra-amniotic infection and impending preterm delivery in women with preterm labor or preterm premature rupture of membranes, PLoS One 13 (2018), e0198842. |
|  |  | [92] | C.L. Kothari, C. Romph, T. Bautista, and D. Lenz, Perinatal Periods of Risk Analysis: Disentangling Race and Socioeconomic Status to Inform a Black Infant Mortality Community Action Initiative, Matern Child Health J 21 (2017), 49-58. |
|  |  | [93] | M. Kunze, M. Klar, C.A. Morfeld, B. Thorns, R.L. Schild, F. Markfeld-Erol, R. Rasenack, H. Proempeler, R. Hentschel, and W.R. Schaefer, Cytokines in noninvasively obtained amniotic fluid as predictors of fetal inflammatory response syndrome, Am J Obstet Gynecol 215 (2016), 96.e91-98. |
|  |  | [94] | N.B. Kunzier, W.L. Kinzler, M.R. Chavez, T.M. Adams, D.A. Brand, and A.M. Vintzileos, The use of cervical sonography to differentiate true from false labor in term patients presenting for labor check, Am J Obstet Gynecol 215 (2016), 372.e371-375. |
|  |  | [95] | M. Kurakazu, F. Yotsumoto, H. Arima, D. Izuchi, D. Urushiyama, K. Miyata, C. Kiyoshima, S. Fukagawa, K. Yoshikawa, M. Kurakazu, T. Hirakawa, K. Shigekawa, R. Araki, A. Sanui, M. Murata, K. Nabeshima, and S. Miyamoto, The combination of maternal blood and amniotic fluid biomarkers improves the predictive accuracy of histologic chorioamnionitis, Placenta 80 (2019), 4-7. |
|  |  | [96] | M. Kurek Eken, A. Tüten, E. Özkaya, G. Karatekin, and A. Karateke, Major determinants of survival and length of stay in the neonatal intensive care unit of newborns from women with premature preterm rupture of membranes, J Matern Fetal Neonatal Med 30 (2017), 1972-1975. |
|  |  | [97] | C.A. Labarrere, H.L. DiCarlo, E. Bammerlin, J.W. Hardin, Y.M. Kim, P. Chaemsaithong, D.M. Haas, G.S. Kassab, and R. Romero, Failure of physiologic transformation of spiral arteries, endothelial and trophoblast cell activation, and acute atherosis in the basal plate of the placenta, Am J Obstet Gynecol 216 (2017), 287.e281-287.e216. |
|  |  | [98] | S.M. Lee, K.H. Park, E.Y. Jung, J.A. Jang, and H.N. Yoo, Frequency and clinical significance of short cervix in patients with preterm premature rupture of membranes, PLoS One 12 (2017), e0174657. |
|  |  | [99] | S.M. Lee, K.H. Park, E.Y. Jung, S.Y. Kook, H. Park, and S.J. Jeon, Inflammatory proteins in maternal plasma, cervicovaginal and amniotic fluids as predictors of intra-amniotic infection in preterm premature rupture of membranes, PLoS One 13 (2018), e0200311. |
|  |  | [100] | Y.J. Lee, S.C. Kim, J.K. Joo, D.H. Lee, K.H. Kim, and K.S. Lee, Amniotic fluid index, single deepest pocket and transvaginal cervical length: Parameter of predictive delivery latency in preterm premature rupture of membranes, Taiwan J Obstet Gynecol 57 (2018), 374-378. |
|  |  | [101] | W.F. Li, A.S. Chao, S.D. Chang, P.J. Cheng, L.Y. Yang, and Y.L. Chang, Effects and outcomes of septostomy in twin-to-twin transfusion syndrome after fetoscopic laser therapy, BMC Pregnancy Childbirth 19 (2019), 397. |
|  |  | [102] | H. Liang, Z. Xie, B. Liu, X. Song, and G. Zhao, A routine urine test has partial predictive value in premature rupture of the membranes, J Int Med Res 47 (2019), 2361-2370. |
|  |  | [103] | L. Liu, H. Wang, Y. Zhang, J. Niu, Z. Li, and R. Tang, Effect of pregravid obesity on perinatal outcomes in singleton pregnancies following in vitro fertilization and the weight-loss goals to reduce the risks of poor pregnancy outcomes: A retrospective cohort study, PLoS One 15 (2020), e0227766. |
|  |  | [104] | Y. Liu, Y. Liu, C. Du, R. Zhang, Z. Feng, and J. Zhang, Diagnostic value of amniotic fluid inflammatory biomarkers for subclinical chorioamnionitis, Int J Gynaecol Obstet 134 (2016), 160-164. |
|  |  | [105] | K. Lyubomirskaya, Y. Krut, L. Sergeyeva, S. Khmil, O. Lototska, N. Petrenko, and A. Kamyshnyi, Preterm premature rupture of membranes: prediction of risks in women of Zaporizhzhia region of Ukraine, Pol Merkur Lekarski 48 (2020), 399-405. |
|  |  | [106] | Y. Ma, Y. Xu, L. Jiang, and X. Shao, Application of a Prediction Model Based on the Laboratory Index Score in Prelabor Rupture of Membranes with Histologic Chorioamnionitis During Late Pregnancy, Med Sci Monit 26 (2020), e924756. |
|  |  | [107] | P. Mach, A. Köninger, L. Wicherek, R. Kimmig, S. Kasimir-Bauer, C. Birdir, B. Schmidt, and A. Gellhaus, Serum concentrations of soluble B7-H4 in early pregnancy are elevated in women with preterm premature rupture of fetal membranes, Am J Reprod Immunol 76 (2016), 149-154. |
|  |  | [108] | Y. Maki, S. Furukawa, T. Nakayama, M. Oohashi, N. Shiiba, K. Furuta, S. Tokunaga, and H. Sameshima, Clinical chorioamnionitis criteria are not sufficient for predicting intra-amniotic infection, J Matern Fetal Neonatal Med (2020), 1-6. |
|  |  | [109] | R.J. Martinez-Portilla, A. Hawkins-Villarreal, P. Alvarez-Ponce, Z.L. Chinolla-Arellano, A.L. Moreno-Espinosa, A.L. Sandoval-Mejia, and N. Moreno-Uribe, Maternal Serum Interleukin-6: A Non-Invasive Predictor of Histological Chorioamnionitis in Women with Preterm-Prelabor Rupture of Membranes, Fetal Diagn Ther 45 (2019), 168-175. |
|  |  | [110] | Y. Mei and Y. Lin, Clinical significance of primary symptoms in women with placental abruption, J Matern Fetal Neonatal Med 31 (2018), 2446-2449. |
|  |  | [111] | M. Merello, L. Lotte, S. Gonfrier, S. Eleni Dit Trolli, F. Casagrande, R. Ruimy, and A. Bongain, Enterobacteria vaginal colonization among patients with preterm premature rupture of membranes from 24 to 34 weeks of gestation and neonatal infection risk, J Gynecol Obstet Hum Reprod 48 (2019), 187-191. |
|  |  | [112] | M. Mikołajczyk, P. Wirstlein, M. Adamczyk, J. Skrzypczak, and E. Wender-Ożegowska, Value of cervicovaginal fluid cytokines in prediction of fetal inflammatory response syndrome in pregnancies complicated with preterm premature rupture of membranes (pPROM), J Perinat Med 48 (2020), 249-255. |
|  |  | [113] | H. Miremberg, N. Elia, C. Marelly, O. Gluck, G. Barda, J. Bar, and E. Weiner, Should the interval between doses of antenatal corticosteroids be shortened in certain cases? Factors predicting preterm delivery < 48 h from presentation, Arch Gynecol Obstet (2021). |
|  |  | [114] | S. Mishra, S. Jaiswar, S. Saad, S. Tripathi, N. Singh, S. Deo, M. Agarwal, and N. Mishra, Platelet indices as a predictive marker in neonatal sepsis and respiratory distress in preterm prelabor rupture of membranes, Int J Hematol 113 (2021), 199-206. |
|  |  | [115] | B.P. Modi, H.I. Parikh, M.E. Teves, R. Kulkarni, J. Liyu, R. Romero, T.P. York, and J.F. Strauss, 3rd, Discovery of rare ancestry-specific variants in the fetal genome that confer risk of preterm premature rupture of membranes (PPROM) and preterm birth, BMC Med Genet 19 (2018), 181. |
|  |  | [116] | A.F. Moron, A. Athanasiou, M. Barbosa, H.J. Milani, S.G. Sarmento, S. Cavalheiro, and S.S. Witkin, Amniotic fluid lactic acid and matrix metalloproteinase-8 levels at the time of fetal surgery for a spine defect: association with subsequent preterm prelabour rupture of membranes, Bjog 125 (2018), 1288-1292. |
|  |  | [117] | L.J. Moulton, J. Eric Jelovsek, M. Lachiewicz, K. Chagin, and O. Goje, A model to predict risk of postpartum infection after Caesarean delivery, J Matern Fetal Neonatal Med 31 (2018), 2409-2417. |
|  |  | [118] | A.S. Mousavi, N. Hashemi, M. Kashanian, N. Sheikhansari, A. Bordbar, and S. Parashi, Comparison between maternal and neonatal outcome of PPROM in the cases of amniotic fluid index (AFI) of more and less than 5 cm, J Obstet Gynaecol 38 (2018), 611-615. |
|  |  | [119] | I. Musilova, C. Andrys, M. Holeckova, V. Kolarova, L. Pliskova, M. Drahosova, R. Bolehovska, R. Pilka, K. Huml, T. Cobo, B. Jacobsson, and M. Kacerovsky, Interleukin-6 measured using the automated electrochemiluminescence immunoassay method for the identification of intra-amniotic inflammation in preterm prelabor rupture of membranes, J Matern Fetal Neonatal Med 33 (2020), 1919-1926. |
|  |  | [120] | I. Musilova, C. Andrys, H. Hornychova, L. Pliskova, M. Drahosova, B. Zednikova, R. Bolehovska, T. Faist, B. Jacobsson, and M. Kacerovsky, Gastric fluid used to assess changes during the latency period in preterm prelabor rupture of membranes, Pediatr Res 84 (2018), 240-247. |
|  |  | [121] | I. Musilova, T. Bestvina, M. Hudeckova, I. Michalec, T. Cobo, B. Jacobsson, and M. Kacerovsky, Vaginal fluid interleukin-6 concentrations as a point-of-care test is of value in women with preterm prelabor rupture of membranes, Am J Obstet Gynecol 215 (2016), 619.e611-619.e612. |
|  |  | [122] | I. Musilova, T. Bestvina, M. Hudeckova, I. Michalec, T. Cobo, B. Jacobsson, and M. Kacerovsky, Vaginal fluid IL-6 concentrations as a point-of-care test is of value in women with preterm PROM, Am J Obstet Gynecol (2016). |
|  |  | [123] | I. Musilova, M. Kacerovsky, M. Stepan, T. Bestvina, L. Pliskova, B. Zednikova, and B. Jacobsson, Maternal serum C-reactive protein concentration and intra-amniotic inflammation in women with preterm prelabor rupture of membranes, PLoS One 12 (2017), e0182731. |
|  |  | [124] | T. Myntti, L. Rahkonen, A. Pätäri-Sampo, M. Tikkanen, T. Sorsa, J. Juhila, O. Helve, S. Andersson, J. Paavonen, and V. Stefanovic, Comparison of amniotic fluid matrix metalloproteinase-8 and cathelicidin in the diagnosis of intra-amniotic infection, J Perinatol 36 (2016), 1049-1054. |
|  |  | [125] | M. Nakahara, S. Goto, E. Kato, S. Nojiri, A. Itakura, and S. Takeda, Maternal risk score for the prediction of fetal inflammatory response syndrome after preterm premature rupture of membranes, J Obstet Gynaecol Res 46 (2020), 2019-2026. |
|  |  | [126] | G. Natarajan, S. Shankaran, S. Saha, A. Laptook, A. Das, R. Higgins, B.J. Stoll, E.F. Bell, W.A. Carlo, C. D'Angio, S.B. DeMauro, P. Sanchez, K. Van Meurs, B. Vohr, N. Newman, E. Hale, and M. Walsh, Antecedents and Outcomes of Abnormal Cranial Imaging in Moderately Preterm Infants, J Pediatr 195 (2018), 66-72.e63. |
|  |  | [127] | D. Odd, A. Heep, K. Luyt, and T. Draycott, Hypoxic-ischemic brain injury: Planned delivery before intrapartum events, J Neonatal Perinatal Med 10 (2017), 347-353. |
|  |  | [128] | K.J. Oh, J. Lee, R. Romero, H.S. Park, J.S. Hong, and B.H. Yoon, A new rapid bedside test to diagnose and monitor intraamniotic inflammation in preterm PROM using transcervically collected fluid, Am J Obstet Gynecol 223 (2020), 423.e421-423.e415. |
|  |  | [129] | S. Ona, S.R. Easter, M. Prabhu, G. Wilkie, R.E. Tuomala, L.E. Riley, and K. Diouf, Diagnostic Validity of the Proposed Eunice Kennedy Shriver National Institute of Child Health and Human Development Criteria for Intrauterine Inflammation or Infection, Obstet Gynecol 133 (2019), 33-39. |
|  |  | [130] | A. Ozel, E. Alici Davutoglu, A. Yurtkal, and R. Madazli, How do platelet-to-lymphocyte ratio and neutrophil-to-lymphocyte ratio change in women with preterm premature rupture of membranes, and threaten preterm labour?, J Obstet Gynaecol 40 (2020), 195-199. |
|  |  | [131] | P. Pacora, R. Romero, O. Erez, E. Maymon, B. Panaitescu, J.P. Kusanovic, A.L. Tarca, C.D. Hsu, and S.S. Hassan, The diagnostic performance of the beta-glucan assay in the detection of intra-amniotic infection with Candida species, J Matern Fetal Neonatal Med 32 (2019), 1703-1720. |
|  |  | [132] | T. Paramel Jayaprakash, E.C. Wagner, J. van Schalkwyk, A.Y. Albert, J.E. Hill, and D.M. Money, High Diversity and Variability in the Vaginal Microbiome in Women following Preterm Premature Rupture of Membranes (PPROM): A Prospective Cohort Study, PLoS One 11 (2016), e0166794. |
|  |  | [133] | H. Park, K.H. Park, Y.M. Kim, S.Y. Kook, S.J. Jeon, and H.N. Yoo, Plasma inflammatory and immune proteins as predictors of intra-amniotic infection and spontaneous preterm delivery in women with preterm labor: a retrospective study, BMC Pregnancy Childbirth 18 (2018), 146. |
|  |  | [134] | J.W. Park, K.H. Park, S.Y. Kook, Y.M. Jung, and Y.M. Kim, Immune biomarkers in maternal plasma to identify histologic chorioamnionitis in women with preterm labor, Arch Gynecol Obstet 299 (2019), 725-732. |
|  |  | [135] | J.W. Park, K.H. Park, J.E. Lee, Y.M. Kim, S.J. Lee, and D.H. Cheon, Antibody Microarray Analysis of Plasma Proteins for the Prediction of Histologic Chorioamnionitis in Women With Preterm Premature Rupture of Membranes, Reprod Sci 26 (2019), 1476-1484. |
|  |  | [136] | S. Park, Y.A. You, H. Yun, S.J. Choi, H.S. Hwang, S.K. Choi, S.M. Lee, and Y.J. Kim, Cervicovaginal fluid cytokines as predictive markers of preterm birth in symptomatic women, Obstet Gynecol Sci 63 (2020), 455-463. |
|  |  | [137] | A.S. Patil, N.W. Gaikwad, C.A. Grotegut, S.D. Dowden, and D.M. Haas, Alterations in endogenous progesterone metabolism associated with spontaneous very preterm delivery, Hum Reprod Open 2020 (2020), hoaa007. |
|  |  | [138] | M.S. Payne, J.P. Newnham, D.A. Doherty, L.L. Furfaro, N.L. Pendal, D.E. Loh, and J.A. Keelan, A specific bacterial DNA signature in the vagina of Australian women in midpregnancy predicts high risk of spontaneous preterm birth (the Predict1000 study), Am J Obstet Gynecol 224 (2021), 206.e201-206.e223. |
|  |  | [139] | C. Petit, P. Deruelle, H. Behal, T. Rakza, S. Balagny, D. Subtil, E. Clouqueur, and C. Garabedian, Preterm premature rupture of membranes: Which criteria contraindicate home care management?, Acta Obstet Gynecol Scand 97 (2018), 1499-1507. |
|  |  | [140] | A. Pinton, F. Severac, N. Meyer, C.Y. Akladios, A. Gaudineau, R. Favre, B. Langer, and N. Sananes, A comparison of vaginal ultrasound and digital examination in predicting preterm delivery in women with threatened preterm labor: a cohort study, Acta Obstet Gynecol Scand 96 (2017), 447-453. |
|  |  | [141] | I.H. Polat, S. Marin, J. Ríos, M. Larroya, A.B. Sánchez-García, C. Murillo, C. Rueda, M. Cascante, E. Gratacós, and T. Cobo, Exploratory and confirmatory analysis to investigate the presence of vaginal metabolome expression of microbial invasion of the amniotic cavity in women with preterm labor using high-performance liquid chromatography, Am J Obstet Gynecol 224 (2021), 90.e91-90.e99. |
|  |  | [142] | N. Pruksanusak, P. Thongphanang, T. Suntharasaj, C. Suwanrath, and A. Geater, Combined maternal-associated risk factors with intrapartum fetal heart rate classification systems to predict peripartum asphyxia neonates, Eur J Obstet Gynecol Reprod Biol 218 (2017), 85-91. |
|  |  | [143] | G. Raba, M. Kacerovsky, and P. Laudański, Eotaxin-2 as a potential marker of preterm premature rupture of membranes: A prospective, cohort, multicenter study, Adv Clin Exp Med 30 (2021), 197-202. |
|  |  | [144] | A.M. Ramseyer, J.R. Whittington, E.F. Magann, J.A. Pates, and S.T. Ounpraseuth, Can maternal characteristics on admission for preterm prelabor rupture of membranes predict pregnancy latency?, Am J Obstet Gynecol MFM 2 (2020), 100194. |
|  |  | [145] | S. Rasmussen, C. Ebbing, and L.M. Irgens, Predicting preeclampsia from a history of preterm birth, PLoS One 12 (2017), e0181016. |
|  |  | [146] | R. Revello, M.J. Alcaide, D. Abehsera, M. Martin-Camean, E.F.G.M. Sousa, B. Alonso-Luque, and J.L. Bartha, Prediction of chorioamnionitis in cases of intraamniotic infection by ureaplasma urealyticum in women with very preterm premature rupture of membranes or preterm labour, J Matern Fetal Neonatal Med 31 (2018), 1839-1844. |
|  |  | [147] | M. Rewatkar, S. Jain, M. Jain, and K. Mohod, C-reactive protein and white blood cell count as predictors of maternal and neonatal infections in prelabour rupture of membranes between 34 and 41 weeks of gestation, J Obstet Gynaecol 38 (2018), 622-628. |
|  |  | [148] | M. Roethlisberger, B. Strizek, I. Gottschalk, M.R. Mallmann, A. Geipel, U. Gembruch, and C. Berg, First-trimester intervention in twin reversed arterial perfusion sequence: does size matter?, Ultrasound Obstet Gynecol 50 (2017), 40-44. |
|  |  | [149] | L.C. Rogers, L. Scott, and J.E. Block, Accurate Point-of-Care Detection of Ruptured Fetal Membranes: Improved Diagnostic Performance Characteristics with a Monoclonal/Polyclonal Immunoassay, Clin Med Insights Reprod Health 10 (2016), 15-18. |
|  |  | [150] | S. Ronzoni, R. D'Souza, O. Shynlova, S. Lye, and K.E. Murphy, Maternal blood endotoxin activity in pregnancies complicated by preterm premature rupture of membranes, J Matern Fetal Neonatal Med 32 (2019), 3473-3479. |
|  |  | [151] | S. Ronzoni, V. Steckle, R. D'Souza, K.E. Murphy, S. Lye, and O. Shynlova, Cytokine Changes in Maternal Peripheral Blood Correlate With Time-to-Delivery in Pregnancies Complicated by Premature Prelabor Rupture of the Membranes, Reprod Sci 26 (2019), 1266-1276. |
|  |  | [152] | M. Rouzaire, M. Corvaisier, V. Roumeau, A. Mulliez, F. Sendy, A. Delabaere, and D. Gallot, Predictors of Short Latency Period Exceeding 48 h after Preterm Premature Rupture of Membranes, J Clin Med 10 (2021). |
|  |  | [153] | H.K. Ryu, J.H. Moon, H.J. Heo, J.W. Kim, and Y.H. Kim, Maternal c-reactive protein and oxidative stress markers as predictors of delivery latency in patients experiencing preterm premature rupture of membranes, Int J Gynaecol Obstet 136 (2017), 145-150. |
|  |  | [154] | M. Sahni, M.E. Franco-Fuenmayor, and K. Shattuck, Management of Late Preterm and Term Neonates exposed to maternal Chorioamnionitis, BMC Pediatr 19 (2019), 282. |
|  |  | [155] | T. Samejima, T. Nagamatsu, D.J. Schust, N. Itaoka, T. Iriyama, T. Nakayama, A. Komatsu, K. Kawana, Y. Osuga, and T. Fujii, Elevated concentration of secretory leukocyte protease inhibitor in the cervical mucus before delivery, Am J Obstet Gynecol 214 (2016), 741.e741-747. |
|  |  | [156] | T. Samejima and K. Takechi, Elevated C-reactive protein levels in histological chorioamnionitis at term: impact of funisitis on term neonates, J Matern Fetal Neonatal Med 30 (2017), 1428-1433. |
|  |  | [157] | J.B. Sauberan, B. Choi, A.R. Paradyse, and J. Le, Quality and Clinical Outcomes Associated with a Gentamicin Use System Change for Managing Chorioamnionitis, J Med Syst 41 (2017), 202. |
|  |  | [158] | W.A. Sayed Ahmed, M.R. Ahmed, M.L. Mohamed, M.A. Hamdy, Z. Kamel, and K.M. Elnahas, Maternal serum interleukin-6 in the management of patients with preterm premature rupture of membranes, J Matern Fetal Neonatal Med 29 (2016), 3162-3166. |
|  |  | [159] | G. Seliger, M. Bergner, R. Haase, H. Stepan, E. Schleußner, J. Zöllkau, S. Seeger, F.B. Kraus, G.G.R. Hiller, A. Wienke, and M. Tchirikov, Daily monitoring of vaginal interleukin 6 as a predictor of intraamniotic inflammation after preterm premature rupture of membranes - a new method of sampling studied in a prospective multicenter trial, J Perinat Med (2021). |
|  |  | [160] | W.H. Sim, H. Ng, and P. Sheehan, Maternal and neonatal outcomes following expectant management of preterm prelabor rupture of membranes before viability, J Matern Fetal Neonatal Med 33 (2020), 533-541. |
|  |  | [161] | G. Sisti, S. Paccosi, A. Parenti, V. Seravalli, M. Di Tommaso, and S.S. Witkin, Insulin-like growth factor binding protein-1 predicts preterm premature rupture of membranes in twin pregnancies, Arch Gynecol Obstet 300 (2019), 583-587. |
|  |  | [162] | G.H. Son, Y. Kim, J.J. Lee, K.Y. Lee, H. Ham, J.E. Song, S.T. Park, and Y.H. Kim, MicroRNA-548 regulates high mobility group box 1 expression in patients with preterm birth and chorioamnionitis, Sci Rep 9 (2019), 19746. |
|  |  | [163] | J. Stranik, M. Kacerovsky, O. Soucek, M. Kolackova, I. Musilova, L. Pliskova, R. Bolehovska, P. Bostik, J. Matulova, B. Jacobsson, and C. Andrys, IgGFc-binding protein in pregnancies complicated by spontaneous preterm delivery: a retrospective cohort study, Sci Rep 11 (2021), 6107. |
|  |  | [164] | J.K. Straughen, D.P. Misra, L.M. Ernst, A.K. Charles, S. VanHorn, S. Ghosh, I. Buhimschi, C. Buhimschi, G. Divine, and C.M. Salafia, Methods to decrease variability in histological scoring in placentas from a cohort of preterm infants, BMJ Open 7 (2017), e013877. |
|  |  | [165] | J.H. Sung, S.J. Choi, S.Y. Oh, C.R. Roh, and J.H. Kim, Revisiting the diagnostic criteria of clinical chorioamnionitis in preterm birth, Bjog 124 (2017), 775-783. |
|  |  | [166] | J.H. Sung, J.Y. Kuk, H.H. Cha, S.J. Choi, S.Y. Oh, C.R. Roh, and J.H. Kim, Amniopatch treatment for preterm premature rupture of membranes before 23 weeks' gestation and factors associated with its success, Taiwan J Obstet Gynecol 56 (2017), 599-605. |
|  |  | [167] | S. Suzuki, Association between clinical chorioamnionitis and histological funisitis at term, J Neonatal Perinatal Med 12 (2019), 37-40. |
|  |  | [168] | J. Takeda, X. Fang, and D.M. Olson, Pregnant human peripheral leukocyte migration during several late pregnancy clinical conditions: a cross-sectional observational study, BMC Pregnancy Childbirth 17 (2017), 16. |
|  |  | [169] | J.L.S. Tebet, G.M. Kirsztajn, T.A. Facca, S.K. Nishida, A.R. Pereira, S.R. Moreira, J.O.P. Medina, and N. Sass, Pregnancy in renal transplant patients: Renal function markers and maternal-fetal outcomes, Pregnancy Hypertens 15 (2019), 108-113. |
|  |  | [170] | P.J. Teoh, A. Ridout, P. Seed, R.M. Tribe, and A.H. Shennan, Gender and preterm birth: Is male fetal gender a clinically important risk factor for preterm birth in high-risk women?, Eur J Obstet Gynecol Reprod Biol 225 (2018), 155-159. |
|  |  | [171] | M.E. Toukam, M. Luisin, J. Chevreau, S. Lanta-Delmas, J. Gondry, and P. Tourneux, A predictive neonatal mortality score for women with premature rupture of membranes after 22-27 weeks of gestation, J Matern Fetal Neonatal Med 32 (2019), 258-264. |
|  |  | [172] | L.A. Underhill, N. Avalos, R. Tucker, Z. Zhang, G. Messerlian, and B. Lechner, Serum Decorin and Biglycan as Potential Biomarkers to Predict PPROM in Early Gestation, Reprod Sci (2019), 1933719119831790. |
|  |  | [173] | L.A. Underhill, N. Avalos, R. Tucker, Z. Zhang, G. Messerlian, and B. Lechner, Serum Decorin and Biglycan as Potential Biomarkers to Predict PPROM in Early Gestation, Reprod Sci 27 (2020), 1620-1626. |
|  |  | [174] | D. Urushiyama, W. Suda, E. Ohnishi, R. Araki, C. Kiyoshima, M. Kurakazu, A. Sanui, F. Yotsumoto, M. Murata, K. Nabeshima, S. Yasunaga, S. Saito, M. Nomiyama, M. Hattori, S. Miyamoto, and K. Hata, Microbiome profile of the amniotic fluid as a predictive biomarker of perinatal outcome, Sci Rep 7 (2017), 12171. |
|  |  | [175] | L. van der Krogt, A.E. Ridout, P.T. Seed, and A.H. Shennan, Placental inflammation and its relationship to cervicovaginal fetal fibronectin in preterm birth, Eur J Obstet Gynecol Reprod Biol 214 (2017), 173-177. |
|  |  | [176] | B.I. Vandermolen, N.L. Hezelgrave, E.M. Smout, D.S. Abbott, P.T. Seed, and A.H. Shennan, Quantitative fetal fibronectin and cervical length to predict preterm birth in asymptomatic women with previous cervical surgery, Am J Obstet Gynecol 215 (2016), 480.e481-480.e410. |
|  |  | [177] | S.W. Verbruggen, M.L. Oyen, A.T. Phillips, and N.C. Nowlan, Function and failure of the fetal membrane: Modelling the mechanics of the chorion and amnion, PLoS One 12 (2017), e0171588. |
|  |  | [178] | F. Wang, Y. Wang, R. Wang, H. Qiu, and L. Chen, Predictive value of maternal serum NF-κB p65 and sTREM-1 for subclinical chorioamnionitis in premature rupture of membranes, Am J Reprod Immunol 76 (2016), 217-223. |
|  |  | [179] | E. Weiner, Y. Mizrachi, E. Grinstein, O. Feldstein, N. Rymer-Haskel, E. Juravel, L. Schreiber, J. Bar, and M. Kovo, The role of placental histopathological lesions in predicting recurrence of preeclampsia, Prenat Diagn 36 (2016), 953-960. |
|  |  | [180] | Y. Yagur, O. Weitzner, E. Ravid, and T. Biron-Shental, Can we predict preterm delivery in patients with premature rupture of membranes?, Arch Gynecol Obstet 300 (2019), 615-621. |
|  |  | [181] | Y. Yamamoto, A. Hirose, V. Jain, and L.K. Hornberger, Branch pulmonary artery Doppler parameters predict early survival-non-survival in premature rupture of membranes, J Perinatol 40 (2020), 1821-1827. |
|  |  | [182] | F. Zhan, S. Zhu, H. Liu, Q. Wang, and G. Zhao, Blood routine test is a good indicator for predicting premature rupture of membranes, J Clin Lab Anal 33 (2019), e22673. |
|  |  | [183] | L.X. Zhang, Y. Sun, H. Zhao, N. Zhu, X.D. Sun, X. Jin, A.M. Zou, Y. Mi, and J.R. Xu, A Bayesian Stepwise Discriminant Model for Predicting Risk Factors of Preterm Premature Rupture of Membranes: A Case-control Study, Chin Med J (Engl) 130 (2017), 2416-2422. |
|  |  | [184] | L. Zhao, Y. Lin, T. Jiang, L. Wang, M. Li, Y. Wang, G. Sun, and M. Xiao, Prediction of the induction to delivery time interval in vaginal dinoprostone-induced labor: a retrospective study in a Chinese tertiary maternity hospital, J Int Med Res 47 (2019), 2647-2654. |
|  |  | [185] | J. Zhu, C. Ma, L. Zhu, J. Li, F. Peng, L. Huang, and X. Luan, A role for the NLRC4 inflammasome in premature rupture of membrane, PLoS One 15 (2020), e0237847. |
|  |  | [186] | Y. Zipori, C. Ben-David, R. Lauterbach, A. Weissman, R. Beloosesky, Y. Ginsberg, Z. Weiner, and N. Khatib, Vaginal birth after cesarean in women with pre-labor rupture of membranes at term, J Matern Fetal Neonatal Med (2020), 1-6. |
| 1. b. Scopus | 21 | [1] | A. Aviram, P. Quaglietta, C. Warshafsky, A. Zaltz, E. Weiner, N. Melamed, E. Ng, J. Barrett, and S. Ronzoni, Utility of ultrasound assessment in management of pregnancies with preterm prelabor rupture of membranes, Ultrasound in Obstetrics and Gynecology 55 (2020), 806-814. |
|  |  | [2] | W. Cao, C. Luo, M. Lei, M. Shen, W. Ding, M. Wang, M. Song, J. Ge, and Q. Zhang, Development and Validation of a Dynamic Nomogram to Predict the Risk of Neonatal White Matter Damage, Front Hum Neurosci 14 (2021). |
|  |  | [3] | J.R. Duncan, K.M. Dorsett, M.M. Aziz, Z. Bursac, M.A. Cleves, A.J. Talati, M.H. Schenone, N.L. Meyer, and G. Mari, Estimated fetal weight and severe neonatal outcomes in preterm prelabor rupture of membranes, J Perinat Med 48 (2020), 687-693. |
|  |  | [4] | J.R. Duncan, K.M. Dorsett, G. Vilchez, M.H. Schenone, and G. Mari, Uterine artery pulsatility index for the prediction of obstetrical complications in preterm prelabor rupture of membranes, Journal of Maternal-Fetal and Neonatal Medicine (2019). |
|  |  | [5] | J.R. Duncan, P. Sawangkum, E.A. Hoover, M.M. Aziz, and G. Vilchez, Birthweight and Apgar at 5 minutes of life for the prediction of severe neonatal outcomes in preterm prelabor rupture of membranes, Journal of Maternal-Fetal and Neonatal Medicine (2020). |
|  |  | [6] | J.R. Duncan, A.M. Tobiasz, K.M. Dorsett, M.M. Aziz, R.E. Thompson, Z. Bursac, A.J. Talati, G. Mari, and M.H. Schenone, Fetal pulmonary artery acceleration/ejection time prognostic accuracy for respiratory complications in preterm prelabor rupture of membranes, Journal of Maternal-Fetal and Neonatal Medicine 33 (2020), 2054-2058. |
|  |  | [7] | T. Kawakita, U.M. Reddy, J.C. Huang, T.C. Auguste, D. Bauer, and R.T. Overcash, Externally Validated Prediction Model of Vaginal Delivery After Preterm Induction With Unfavorable Cervix, Obstet Gynecol 136 (2020), 716-724. |
|  |  | [8] | B. Kaya, U. Turhan, S. Sezer, S. Kaya, İ. Dağ, and A. Tayyar, Maternal serum galectin-1 and galectin-3 levels in pregnancies complicated with preterm prelabor rupture of membranes, Journal of Maternal-Fetal and Neonatal Medicine 33 (2020), 861-868. |
|  |  | [9] | K. Kuhrt, N. Hezelgrave, C. Foster, P.T. Seed, and A.H. Shennan, Development and validation of a tool incorporating quantitative fetal fibronectin to predict spontaneous preterm birth in symptomatic women, Ultrasound in Obstetrics and Gynecology 47 (2016), 210-216. |
|  |  | [10] | K. Kuhrt, E. Smout, N. Hezelgrave, P.T. Seed, J. Carter, and A.H. Shennan, Development and validation of a tool incorporating cervical length and quantitative fetal fibronectin to predict spontaneous preterm birth in asymptomatic high-risk women, Ultrasound in Obstetrics and Gynecology 47 (2016), 104-109. |
|  |  | [11] | C.A. Labarrere, H.L. DiCarlo, E. Bammerlin, J.W. Hardin, Y.M. Kim, P. Chaemsaithong, D.M. Haas, G.S. Kassab, and R. Romero, Failure of physiologic transformation of spiral arteries, endothelial and trophoblast cell activation, and acute atherosis in the basal plate of the placenta, Am J Obstet Gynecol 216 (2017), 287.e281-287.e216. |
|  |  | [12] | F. Lemoine, R.C. Moore, A. Chapple, F.A. Moore, and E. Sutton, Neonatal Survivability following Previable PPROM after Hospital Readmission for Intervention, AJP Reports 10 (2020), E395-E402. |
|  |  | [13] | Y. Ma, Y. Xu, L. Jiang, and X. Shao, Application of a prediction model based on the laboratory index score in prelabor rupture of membranes with histologic chorioamnionitis during late pregnancy, Medical Science Monitor 26 (2020). |
|  |  | [14] | Y. Maki, S. Furukawa, T. Nakayama, M. Oohashi, N. Shiiba, K. Furuta, S. Tokunaga, and H. Sameshima, Clinical chorioamnionitis criteria are not sufficient for predicting intra-amniotic infection, Journal of Maternal-Fetal and Neonatal Medicine (2020). |
|  |  | [15] | R.J. Martinez-Portilla, A. Hawkins-Villarreal, P. Alvarez-Ponce, Z.L. Chinolla-Arellano, A.L. Moreno-Espinosa, A.L. Sandoval-Mejia, and N. Moreno-Uribe, Maternal Serum Interleukin-6: A Non-Invasive Predictor of Histological Chorioamnionitis in Women with Preterm-Prelabor Rupture of Membranes, Fetal Diagn Ther 45 (2019), 168-175. |
|  |  | [16] | I. Musilova, C. Andrys, M. Drahosova, O. Soucek, R. Kutova, L. Pliskova, R. Spacek, P. Laudanski, B. Jacobsson, and M. Kacerovsky, Amniotic fluid calreticulin in pregnancies complicated by the preterm prelabor rupture of membranes, Journal of Maternal-Fetal and Neonatal Medicine 29 (2016), 3921-3929. |
|  |  | [17] | I. Musilova, C. Andrys, M. Drahosova, O. Soucek, L. Pliskova, M. Stepan, T. Bestvina, J. Maly, B. Jacobsson, and M. Kacerovsky, Amniotic fluid cathepsin-G in pregnancies complicated by the preterm prelabor rupture of membranes, Journal of Maternal-Fetal and Neonatal Medicine 30 (2017), 2097-2104. |
|  |  | [18] | T. Myntti, L. Rahkonen, A. Pätäri-Sampo, M. Tikkanen, T. Sorsa, J. Juhila, O. Helve, S. Andersson, J. Paavonen, and V. Stefanovic, Comparison of amniotic fluid matrix metalloproteinase-8 and cathelicidin in the diagnosis of intra-amniotic infection, Journal of Perinatology 36 (2016), 1049-1054. |
|  |  | [19] | D. Odd, A. Heep, K. Luyt, and T. Draycott, Hypoxic-ischemic brain injury: Planned delivery before intrapartum events, J Neonatal Perinatal Med 10 (2017), 347-353. |
|  |  | [20] | S. Ronzoni, V. Steckle, R. D’Souza, K.E. Murphy, S. Lye, and O. Shynlova, Cytokine Changes in Maternal Peripheral Blood Correlate With Time-to-Delivery in Pregnancies Complicated by Premature Prelabor Rupture of the Membranes, Reproductive Sciences 26 (2019), 1266-1276. |
|  |  | [21] | B.I. Vandermolen, N.L. Hezelgrave, E.M. Smout, D.S. Abbott, P.T. Seed, and A.H. Shennan, Quantitative fetal fibronectin and cervical length to predict preterm birth in asymptomatic women with previous cervical surgery, Am J Obstet Gynecol 215 (2016), 480.e481-480.e410. |
| 1. c. Web of Science | 44 | [1] | N. Aracic, I. Stipic, I.J. Alujevic, P. Poljak, and M. Stipic, The value of ultrasound measurement of cervical length and parity in prediction of cesarean section risk in term premature rupture of membranes and unfavorable cervix, J Perinat Med 45 (2017), 99-104. |
|  |  | [2] | I.M. Aris, S. Logan, C. Lim, M. Choolani, A. Biswas, and S. Bhattacharya, Preterm prelabour rupture of membranes: a retrospective cohort study of association with adverse outcome in subsequent pregnancy, Bjog-an International Journal of Obstetrics and Gynaecology 124 (2017), 1698-1707. |
|  |  | [3] | A. Aviram, P. Quaglietta, C. Warshafsky, A. Zaltz, E. Weiner, N. Melamed, E. Ng, J. Barrett, and S. Ronzoni, Utility of ultrasound assessment in management of pregnancies with preterm prelabor rupture of membranes, Ultrasound in Obstetrics & Gynecology 55 (2020), 806-814. |
|  |  | [4] | E. Baser, D.A. Kirmizi, D.U. Isik, S. Ozdemirci, T. Onat, E.S. Yalvac, N. Demirel, and O.M. Tekin, The effects of latency period in PPROM cases managed expectantly, Journal of Maternal-Fetal & Neonatal Medicine 33 (2020), 2274-2283. |
|  |  | [5] | T. Cobo, V. Aldecoa, F. Figueras, A. Herranz, S. Ferrero, M. Izquierdo, C. Murillo, R. Amoedo, C. Rueda, J. Bosch, R.J. Martinez-Portilla, E. Gratacos, and M. Palacio, Development and validation of a multivariable prediction model of spontaneous preterm delivery and microbial invasion of the amniotic cavity in women with preterm labor, Am J Obstet Gynecol 223 (2020). |
|  |  | [6] | T. Cobo, J. Munros, J. Rios, J. Ferreri, F. Migliorelli, N. Banos, E. Gratacos, and M. Palacio, Contribution of Amniotic Fluid along Gestation to the Prediction of Perinatal Mortality in Women with Early Preterm Premature Rupture of Membranes, Fetal Diagn Ther 43 (2018), 105-112. |
|  |  | [7] | G. Daskalakis, P. Fotinopoulos, V. Pergialiotis, M. Theodora, P. Antsaklis, M. Sindos, N. Papantoniou, and D. Loutradis, Delayed interval delivery of the second twin in a woman with altered markers of inflammation, BMC Pregnancy Childbirth 18 (2018). |
|  |  | [8] | J.R. Duncan, K.M. Dorsett, M.M. Aziz, Z. Bursac, M.A. Cleves, A.J. Talati, M.H. Schenone, N.L. Meyer, and G. Mari, Estimated fetal weight and severe neonatal outcomes in preterm prelabor rupture of membranes, J Perinat Med 48 (2020), 687-693. |
|  |  | [9] | J.R. Duncan, K.M. Dorsett, G. Vilchez, M.H. Schenone, and G. Mari, Uterine artery pulsatility index for the prediction of obstetrical complications in preterm prelabor rupture of membranes, Journal of Maternal-Fetal & Neonatal Medicine. |
|  |  | [10] | J.R. Duncan, P. Sawangkum, E.A. Hoover, M.M. Aziz, and G. Vilchez, Birthweight and Apgar at 5 minutes of life for the prediction of severe neonatal outcomes in preterm prelabor rupture of membranes, Journal of Maternal-Fetal & Neonatal Medicine. |
|  |  | [11] | J.R. Duncan, A.M. Tobiasz, K.M. Dorsett, M.M. Aziz, R.E. Thompson, Z. Bursac, A.J. Talati, G. Mari, and M.H. Schenone, Fetal pulmonary artery acceleration/ejection time prognostic accuracy for respiratory complications in preterm prelabor rupture of membranes, Journal of Maternal-Fetal & Neonatal Medicine 33 (2020), 2054-2058. |
|  |  | [12] | V. El-Achi, B. de Vries, C. O'Brien, F. Park, J. Tooher, and J. Hyett, First-Trimester Prediction of Preterm Prelabour Rupture of Membranes, Fetal Diagn Ther 47 (2020), 624-629. |
|  |  | [13] | G.U. Eleje, E.C. Ezugwu, A.C. Eke, J.I. Ikechebelu, C.C. Obiora, N.O. Ojiegbe, I.U. Ezebialu, C.O. Ezeama, B.O. Nwosu, G.O. Udigwe, C.I. Okafor, and F.O. Ezugwu, Comparison of the duo of insulin-like growth factor binding protein-1/alpha fetoprotein (Amnioquick duo plus (R)) and traditional clinical assessment for diagnosing premature rupture of fetal membranes, J Perinat Med 45 (2017), 105-112. |
|  |  | [14] | A. Frick, V. Kostiv, D. Vojtassakova, R. Akolekar, and K.H. Nicolaides, Comparison of different methods of measuring angle of progression in prediction of labor outcome, Ultrasound in Obstetrics & Gynecology 55 (2020), 391-400. |
|  |  | [15] | X.H. Fu, X.B. He, D.F. Sun, Q.H. Fan, B.L. Zhang, Q. Wang, Y.F. Wang, and H.L. Chen, Histological chorioamnionitis could be predicted with high accuracy in preterm labor with intact membranes before delivery, Clin Exp Obstet Gynecol 45 (2018), 720-724. |
|  |  | [16] | N. Garry, O. Keenan, S.W. Lindow, and T. Darcy, Pregnancy outcomes following elective abdominal cerclage following cervical excision surgery for neoplastic disease, European Journal of Obstetrics & Gynecology and Reproductive Biology 256 (2021), 225-229. |
|  |  | [17] | L. Hiersch, E. Krispin, A. Aviram, M. Mor-Shacham, R. Gabbay-Benziv, Y. Yogev, and E. Ashwal, Predictors for prolonged interval from premature rupture of membranes to spontaneous onset of labor at term, Journal of Maternal-Fetal & Neonatal Medicine 30 (2017), 1465-1470. |
|  |  | [18] | J.K. Iversen, B.H. Kahrs, E.A. Torkildsen, and T.M. Eggebo, Fetal molding examined with transperineal ultrasound and associations with position and delivery mode, Am J Obstet Gynecol 223 (2020). |
|  |  | [19] | E.Y. Jung, K.H. Park, B.R. Han, S.H. Cho, and A. Ryu, Measurement of Interleukin 8 in Cervicovaginal Fluid in Women With Preterm Premature Rupture of Membranes: A Comparison of Amniotic Fluid Samples, Reproductive Sciences 24 (2017), 142-147. |
|  |  | [20] | T. Kawakita, U.M. Reddy, J.C. Huang, T.C. Auguste, D. Bauer, and R.T. Overcash, Externally Validated Prediction Model of Vaginal Delivery After Preterm Induction With Unfavorable Cervix, Obstet Gynecol 136 (2020), 716-724. |
|  |  | [21] | B. Kaya, U. Turhan, S. Sezer, S. Kaya, I. Dag, and A. Tayyar, Maternal serum galectin-1 and galectin-3 levels in pregnancies complicated with preterm prelabor rupture of membranes, Journal of Maternal-Fetal & Neonatal Medicine 33 (2020), 861-868. |
|  |  | [22] | G. Kayem, F. Batteux, N. Girard, T. Schmitz, M. Willaime, F. Maillard, P.H. Jarreau, and F. Goffinet, Predictive value of vaginal IL-6 and TNF alpha bedside tests repeated until delivery for the prediction of maternal-fetal infection in cases of premature rupture of membranes, European Journal of Obstetrics & Gynecology and Reproductive Biology 211 (2017), 8-14. |
|  |  | [23] | H.J. Kim, K.H. Park, Y.M. Kim, E. Joo, K. Ahn, and S. Shin, A protein microarray analysis of amniotic fluid proteins for the prediction of spontaneous preterm delivery in women with preterm premature rupture of membranes at 23 to 30 weeks of gestation, PLoS One 15 (2020). |
|  |  | [24] | P. Kosinski and M. Wielgos, Foetoscopic endotracheal occlusion (FETO) for severe isolated left-sided congenital diaphragmatic hernia: single center Polish experience, Journal of Maternal-Fetal & Neonatal Medicine 31 (2018), 2521-2526. |
|  |  | [25] | C.A. Labarrere, H.L. DiCarlo, E. Bammerlin, J.W. Hardin, Y.M. Kim, P. Chaemsaithong, D.M. Haas, G.S. Kassab, and R. Romero, Failure of physiologic transformation of spiral arteries, endothelial and trophoblast cell activation, and acute atherosis in the basal plate of the placenta, Am J Obstet Gynecol 216 (2017). |
|  |  | [26] | J. Li, B. Yu, W. Wang, D. Luo, Q.L. Dai, and X.Q. Gan, Does intact umbilical cord milking increase infection rates in preterm infants with premature prolonged rupture of membranes?, Journal of Maternal-Fetal & Neonatal Medicine 33 (2020), 184-190. |
|  |  | [27] | G.C. Ma, Y.C. Chen, W.J. Wu, S.P. Chang, T.Y. Chang, W.H. Lin, and M. Chen, Prenatal Diagnosis of Autosomal Recessive Renal Tubular Dysgenesis with Anhydramnios Caused by a Mutation in the AGT Gene, Diagnostics 9 (2019). |
|  |  | [28] | Y. Ma, Y. Xu, L.J. Jiang, and X.N. Shao, Application of a Prediction Model Based on the Laboratory Index Score in Prelabor Rupture of Membranes with Histologic Chorioamnionitis During Late Pregnancy, Medical Science Monitor 26 (2020). |
|  |  | [29] | Y. Maki, S. Furukawa, T. Nakayama, M. Oohashi, N. Shiiba, K. Furuta, S. Tokunaga, and H. Sameshima, Clinical chorioamnionitis criteria are not sufficient for predicting intra-amniotic infection, Journal of Maternal-Fetal & Neonatal Medicine. |
|  |  | [30] | R.J. Martinez-Portilla, A. Hawkins-Villarreal, P. Alvarez-Ponce, Z.L. Chinolla-Arellano, A.L. Moreno-Espinosa, A.L. Sandoval-Mejia, and N. Moreno-Uribe, Maternal Serum Interleukin-6: A Non-Invasive Predictor of Histological Chorioamnionitis in Women with Preterm-Prelabor Rupture of Membranes, Fetal Diagn Ther 45 (2019), 168-175. |
|  |  | [31] | M. Mikolajczyk, P. Wirstlein, M. Adamczyk, J. Skrzypczak, and E. Wender-Ozegowska, Value of cervicovaginal fluid cytokines in prediction of fetal inflammatory response syndrome in pregnancies complicated with preterm premature rupture of membranes (pPROM), J Perinat Med 48 (2020), 249-255. |
|  |  | [32] | I. Musilova, C. Andrys, M. Drahosova, O. Soucek, L. Pliskova, B. Jacobsson, and M. Kacerovsky, Cervical fluid interleukin 6 and intra-amniotic complications of preterm prelabor rupture of membranes, Journal of Maternal-Fetal & Neonatal Medicine 31 (2018), 827-836. |
|  |  | [33] | I. Musilova, M. Kacerovsky, M. Stepan, T. Bestvina, L. Pliskova, B. Zednikova, and B. Jacobsson, Maternal serum C-reactive protein concentration and intra-amniotic inflammation in women with preterm prelabor rupture of membranes, PLoS One 12 (2017). |
|  |  | [34] | I. Musilova, L. Pliskova, R. Gerychova, P. Janku, O. Simetka, P. Matlak, B. Jacobsson, and M. Kacerovsky, Maternal white blood cell count cannot identify the presence of microbial invasion of the amniotic cavity or intra-amniotic inflammation in women with preterm prelabor rupture of membranes, PLoS One 12 (2017). |
|  |  | [35] | M. Nakahara, S. Goto, E. Kato, S. Nojiri, A. Itakura, and S. Takeda, Maternal risk score for the prediction of fetal inflammatory response syndrome after preterm premature rupture of membranes, Journal of Obstetrics and Gynaecology Research 46 (2020), 2019-2026. |
|  |  | [36] | M. Rewatkar, S. Jain, M. Jain, and K. Mohod, C-reactive protein and white blood cell count as predictors of maternal and neonatal infections in prelabour rupture of membranes between 34 and 41 weeks of gestation, Journal of Obstetrics and Gynaecology 38 (2018), 622-628. |
|  |  | [37] | S. Ronzoni, V. Steckle, R. D'Souza, K.E. Murphy, S. Lye, and O. Shynlova, Cytokine Changes in Maternal Peripheral Blood Correlate With Time-to-Delivery in Pregnancies Complicated by Premature Prelabor Rupture of the Membranes, Reproductive Sciences 26 (2019), 1266-1276. |
|  |  | [38] | S. Shinar, S. Agrawal, D. El-Chaar, N. Abbasi, R. Beecroft, J. Kachura, J. Keunen, G. Seaward, T. Van Mieghem, R. Windrim, and G. Ryan, Selective fetal reduction in complicated monochorionic twin pregnancies: A comparison of techniques, Prenat Diagn 41 (2021), 52-60. |
|  |  | [39] | J.H. Sung, S.J. Choi, S.Y. Oh, C.R. Roh, and J.H. Kim, Revisiting the diagnostic criteria of clinical chorioamnionitis in preterm birth, Bjog-an International Journal of Obstetrics and Gynaecology 124 (2017), 775-783. |
|  |  | [40] | K. Uno, M. Mayama, M. Yoshihara, T. Takeda, S. Tano, T. Suzuki, Y. Kishigami, and H. Oguchi, Reasons for previous Cesarean deliveries impact a woman's independent decision of delivery mode and the success of trial of labor after Cesarean, BMC Pregnancy Childbirth 20 (2020). |
|  |  | [41] | S. Usman, H. Barton, C. Wilhelm-Benartzi, and C.C. Lees, Ultrasound is better tolerated than vaginal examination in and before labour, Australian & New Zealand Journal of Obstetrics & Gynaecology 59 (2019), 362-366. |
|  |  | [42] | S. Usman, M. Wilkinson, H. Barton, and C.C. Lees, The feasibility and accuracy of ultrasound assessment in the labor room, Journal of Maternal-Fetal & Neonatal Medicine 32 (2019), 3442-3451. |
|  |  | [43] | N. Volpe, E. di Pasquo, A. Ferretti, A. Dall'Asta, S. Fieni, T. Frusca, and T. Ghi, Hyperechoic amniotic membranes in patients with preterm premature rupture of membranes (p-PROM) and pregnancy outcome, J Perinat Med 49 (2021), 311-318. |
|  |  | [44] | E. Weiner, J. Barrett, A. Zaltz, M. Ram, A. Aviram, M. Kibel, H. Lipworth, E. Asztalos, and N. Melamed, Amniotic fluid volume at presentation with early preterm prelabor rupture of membranes and association with severe neonatal respiratory morbidity, Ultrasound in Obstetrics & Gynecology 54 (2019), 767-773. |
| 2. Non-duplicated records | 209 | [1] | Good clinical practice advice: Prediction of preterm labor and preterm premature rupture of membranes, Int J Gynaecol Obstet 144 (2019), 340-346. |
|  |  | [2] | K. Abdel-Malek, M.A. El-Halwagi, B.E. Hammad, O. Azmy, O. Helal, M. Eid, and M. Abdel-Rasheed, Role of maternal serum ferritin in prediction of preterm labour, J Obstet Gynaecol 38 (2018), 222-225. |
|  |  | [3] | W.E.t. Ackerman, I.A. Buhimschi, D. Brubaker, S. Maxwell, K.M. Rood, M.R. Chance, H. Jing, S. Mesiano, and C.S. Buhimschi, Integrated microRNA and mRNA network analysis of the human myometrial transcriptome in the transition from quiescence to labor, Biol Reprod 98 (2018), 834-845. |
|  |  | [4] | S. Alla, A. Ramseyer, J.R. Whittington, S. Peeples, S.T. Ounpraseuth, and E.F. Magann, Maternal features at time of preterm prelabor rupture of membranes and short-term neonatal outcomes, J Matern Fetal Neonatal Med (2020), 1-7. |
|  |  | [5] | R.M. Alvarez, I. Biliatis, A. Rockall, E. Papadakou, S.A. Sohaib, N.M. deSouza, J. Butler, M. Nobbenhuis, D. Barton, J.H. Shepherd, and T. Ind, MRI measurement of residual cervical length after radical trachelectomy for cervical cancer and the risk of adverse pregnancy outcomes: a blinded imaging analysis, Bjog 125 (2018), 1726-1733. |
|  |  | [6] | N. Aracic, I. Stipic, I. Jakus Alujevic, P. Poljak, and M. Stipic, The value of ultrasound measurement of cervical length and parity in prediction of cesarean section risk in term premature rupture of membranes and unfavorable cervix, J Perinat Med 45 (2017), 99-104. |
|  |  | [7] | I.M. Aris, S. Logan, C. Lim, M. Choolani, A. Biswas, and S. Bhattacharya, Preterm prelabour rupture of membranes: a retrospective cohort study of association with adverse outcome in subsequent pregnancy, Bjog-an International Journal of Obstetrics and Gynaecology 124 (2017), 1698-1707. |
|  |  | [8] | N. Asadi, A. Faraji, A. Keshavarzi, M. Akbarzadeh-Jahromi, and S. Yoosefi, Predictive value of procalcitonin, C-reactive protein, and white blood cells for chorioamnionitis among women with preterm premature rupture of membranes, Int J Gynaecol Obstet 147 (2019), 83-88. |
|  |  | [9] | M.T. Aung, Y. Yu, K.K. Ferguson, D.E. Cantonwine, L. Zeng, T.F. McElrath, S. Pennathur, B. Mukherjee, and J.D. Meeker, Prediction and associations of preterm birth and its subtypes with eicosanoid enzymatic pathways and inflammatory markers, Sci Rep 9 (2019), 17049. |
|  |  | [10] | A. Aviram, P. Quaglietta, C. Warshafsky, A. Zaltz, E. Weiner, N. Melamed, E. Ng, J. Barrett, and S. Ronzoni, Utility of ultrasound assessment in management of pregnancies with preterm prelabor rupture of membranes, Ultrasound Obstet Gynecol 55 (2020), 806-814. |
|  |  | [11] | U. Baboolall, Y. Zha, X. Gong, D.R. Deng, F. Qiao, and H. Liu, Variations of plasma D-dimer level at various points of normal pregnancy and its trends in complicated pregnancies: A retrospective observational cohort study, Medicine (Baltimore) 98 (2019), e15903. |
|  |  | [12] | F. Bahlmann and A. Al Naimi, Using the angiogenic factors sFlt-1 and PlGF with Doppler ultrasound of the uterine artery for confirming preeclampsia, Arch Gynecol Obstet 294 (2016), 1133-1139. |
|  |  | [13] | A.R. Ballard, L.H. Mallett, J.E. Pruszynski, and J.B. Cantey, Chorioamnionitis and subsequent bronchopulmonary dysplasia in very-low-birth weight infants: a 25-year cohort, J Perinatol 36 (2016), 1045-1048. |
|  |  | [14] | S.V. Barinov, G.C. Di Renzo, A.A. Belinina, O.V. Koliado, and O.V. Remneva, Clinical and biochemical markers of spontaneous preterm birth in singleton and multiple pregnancies, J Matern Fetal Neonatal Med (2021), 1-6. |
|  |  | [15] | D. Başbuğ, A. Başbuğ, and C. Gülerman, Is unexplained elevated maternal serum alpha-fetoprotein still important predictor for adverse pregnancy outcome?, Ginekol Pol 88 (2017), 325-330. |
|  |  | [16] | E. Baser, D. Aydogan Kirmizi, D. Ulubas Isik, S. Ozdemirci, T. Onat, E. Serdar Yalvac, N. Demirel, and O. Moraloglu Tekin, The effects of latency period in PPROM cases managed expectantly, J Matern Fetal Neonatal Med 33 (2020), 2274-2283. |
|  |  | [17] | J.B. Beavers, S. Bai, J. Perry, J. Simpson, and S. Peeples, Implementation and Evaluation of the Early-Onset Sepsis Risk Calculator in a High-Risk University Nursery, Clin Pediatr (Phila) 57 (2018), 1080-1085. |
|  |  | [18] | M.A. Belfort, Predicting premature preterm rupture of the membranes after fetal surgery, Bjog 125 (2018), 1293. |
|  |  | [19] | Z. Bouzari, R. Shahhosseini, M. Mohammadnetaj, S. Barat, S. Yazdani, and K. Hajian-Tilaki, Vaginal discharge concentrations of β-human chorionic gonadotropin, creatinine, and urea for the diagnosis of premature rupture of membranes, Int J Gynaecol Obstet 141 (2018), 97-101. |
|  |  | [20] | A.P. Burgess, J. Katz, J. Pessolano, J. Ponterio, M. Moretti, and N.A. Lakhi, Determination of antepartum and intrapartum risk factors associated with neonatal intensive care unit admission, J Perinat Med 44 (2016), 589-596. |
|  |  | [21] | J. Caloone, M. Rabilloud, F. Boutitie, A. Traverse-Glehen, F. Allias-Montmayeur, L. Denis, C. Boisson-Gaudin, I.J. Hot, P. Guerre, M. Cortet, and C. Huissoud, Accuracy of several maternal seric markers for predicting histological chorioamnionitis after preterm premature rupture of membranes: a prospective and multicentric study, Eur J Obstet Gynecol Reprod Biol 205 (2016), 133-140. |
|  |  | [22] | M.T. Canda, N. Demir, and O. Sezer, Fetal Nasal Bone Length as a Novel Marker for Prediction of Adverse Perinatal Outcomes in the First-Trimester of Pregnancy, Balkan Med J 34 (2017), 127-131. |
|  |  | [23] | W. Cao, C. Luo, M. Lei, M. Shen, W. Ding, M. Wang, M. Song, J. Ge, and Q. Zhang, Development and Validation of a Dynamic Nomogram to Predict the Risk of Neonatal White Matter Damage, Front Hum Neurosci 14 (2020), 584236. |
|  |  | [24] | D. Carola, M. Vasconcellos, A. Sloane, D. McElwee, C. Edwards, J. Greenspan, and Z.H. Aghai, Utility of Early-Onset Sepsis Risk Calculator for Neonates Born to Mothers with Chorioamnionitis, J Pediatr 195 (2018), 48-52.e41. |
|  |  | [25] | O. Cetin, Z.D. Aydın, F.F. Verit, A.G. Zebitay, E. Karaman, S. Elasan, M. Turfan, and O. Yucel, Is Maternal Blood Procalcitonin Level a Reliable Predictor for Early Onset Neonatal Sepsis in Preterm Premature Rupture of Membranes?, Gynecol Obstet Invest 82 (2017), 163-169. |
|  |  | [26] | H.H. Cha, J.M. Kim, H.M. Kim, M.J. Kim, G.O. Chong, and W.J. Seong, Association between gestational age at delivery and lymphocyte-monocyte ratio in the routine second trimester complete blood cell count, Yeungnam Univ J Med 38 (2021), 34-38. |
|  |  | [27] | P. Chaemsaithong, R. Romero, N. Docheva, N. Chaiyasit, G. Bhatti, P. Pacora, S.S. Hassan, L. Yeo, and O. Erez, Comparison of rapid MMP-8 and interleukin-6 point-of-care tests to identify intra-amniotic inflammation/infection and impending preterm delivery in patients with preterm labor and intact membranes(), J Matern Fetal Neonatal Med 31 (2018), 228-244. |
|  |  | [28] | N. Chaiyasit, R. Romero, P. Chaemsaithong, N. Docheva, G. Bhatti, J.P. Kusanovic, Z. Dong, L. Yeo, P. Pacora, S.S. Hassan, and O. Erez, Clinical chorioamnionitis at term VIII: a rapid MMP-8 test for the identification of intra-amniotic inflammation, J Perinat Med 45 (2017), 539-550. |
|  |  | [29] | I. Chandra and L. Sun, Third trimester preterm and term premature rupture of membranes: Is there any difference in maternal characteristics and pregnancy outcomes?, J Chin Med Assoc 80 (2017), 657-661. |
|  |  | [30] | S. Chaudhury, J. Saqibuddin, R. Birkett, K. Falcon-Girard, M. Kraus, L.M. Ernst, W. Grobman, and K.K. Mestan, Variations in Umbilical Cord Hematopoietic and Mesenchymal Stem Cells With Bronchopulmonary Dysplasia, Front Pediatr 7 (2019), 475. |
|  |  | [31] | A.L. Chen, I.T. Goldfarb, A.O. Scourtas, and D.J. Roberts, The histologic evolution of revealed, acute abruptions, Hum Pathol 67 (2017), 187-197. |
|  |  | [32] | K.W. Cheung, L.N. Tan, M.T.Y. Seto, S. Moholkar, G. Masson, and M.D. Kilby, Prenatal Diagnosis, Management, and Outcome of Fetal Subdural Haematoma: A Case Report and Systematic Review, Fetal Diagn Ther 46 (2019), 285-295. |
|  |  | [33] | H.Y. Cho, I. Jung, J.Y. Kwon, S.J. Kim, Y.W. Park, and Y.H. Kim, The Delta Neutrophil Index as a predictive marker of histological chorioamnionitis in patients with preterm premature rupture of membranes: A retrospective study, PLoS One 12 (2017), e0173382. |
|  |  | [34] | T. Cobo, V. Aldecoa, F. Figueras, A. Herranz, S. Ferrero, M. Izquierdo, C. Murillo, R. Amoedo, C. Rueda, J. Bosch, R.J. Martínez-Portilla, E. Gratacós, and M. Palacio, Development and validation of a multivariable prediction model of spontaneous preterm delivery and microbial invasion of the amniotic cavity in women with preterm labor, Am J Obstet Gynecol 223 (2020), 421.e421-421.e414. |
|  |  | [35] | T. Cobo, J. Munrós, J. Ríos, J. Ferreri, F. Migliorelli, N. Baños, E. Gratacós, and M. Palacio, Contribution of Amniotic Fluid along Gestation to the Prediction of Perinatal Mortality in Women with Early Preterm Premature Rupture of Membranes, Fetal Diagn Ther 43 (2018), 105-112. |
|  |  | [36] | Y. Dabi, S. Nedellec, C. Bonneau, B. Trouchard, R. Rouzier, and A. Benachi, Clinical validation of a model predicting the risk of preterm delivery, PLoS One 12 (2017), e0171801. |
|  |  | [37] | C.B. Dartibale, N.S. Uchimura, L. Nery, A.P. Schumeish, L.Y.T. Uchimura, R.G. Santana, and T.T. Uchimura, Qualitative Determination of Human Chorionic Gonadotropin in Vaginal Washings for the Early Diagnosis of Premature Rupture of Fetal Membranes, Rev Bras Ginecol Obstet 39 (2017), 317-321. |
|  |  | [38] | G. Daskalakis, P. Fotinopoulos, V. Pergialiotis, M. Theodora, P. Antsaklis, M. Sindos, N. Papantoniou, and D. Loutradis, Delayed interval delivery of the second twin in a woman with altered markers of inflammation, BMC Pregnancy Childbirth 18 (2018), 206. |
|  |  | [39] | C. Davies, G. Segre, A. Estradé, J. Radua, A. De Micheli, U. Provenzani, D. Oliver, G. Salazar de Pablo, V. Ramella-Cravaro, M. Besozzi, P. Dazzan, M. Miele, G. Caputo, C. Spallarossa, G. Crossland, A. Ilyas, G. Spada, P. Politi, R.M. Murray, P. McGuire, and P. Fusar-Poli, Prenatal and perinatal risk and protective factors for psychosis: a systematic review and meta-analysis, Lancet Psychiatry 7 (2020), 399-410. |
|  |  | [40] | D.A. De Silva, S. Lisonkova, P. von Dadelszen, A.R. Synnes, and L.A. Magee, Timing of delivery in a high-risk obstetric population: a clinical prediction model, BMC Pregnancy Childbirth 17 (2017), 202. |
|  |  | [41] | N. Dorfeuille, V. Morin, A. Tétu, S. Demers, G. Laforest, K. Gouin, B. Piedboeuf, and E. Bujold, Vaginal Fluid Inflammatory Biomarkers and the Risk of Adverse Neonatal Outcomes in Women with PPROM, Am J Perinatol 33 (2016), 1003-1007. |
|  |  | [42] | R.D. Ducci, P.J. Lorenzoni, C.S. Kay, L.C. Werneck, and R.H. Scola, Clinical follow-up of pregnancy in myasthenia gravis patients, Neuromuscul Disord 27 (2017), 352-357. |
|  |  | [43] | J.R. Duncan, K.M. Dorsett, M.M. Aziz, Z. Bursac, M.A. Cleves, A.J. Talati, M.H. Schenone, N.L. Meyer, and G. Mari, Estimated fetal weight and severe neonatal outcomes in preterm prelabor rupture of membranes, J Perinat Med 48 (2020), 687-693. |
|  |  | [44] | J.R. Duncan, K.M. Dorsett, G. Vilchez, M.H. Schenone, and G. Mari, Uterine artery pulsatility index for the prediction of obstetrical complications in preterm prelabor rupture of membranes, J Matern Fetal Neonatal Med (2019), 1-4. |
|  |  | [45] | J.R. Duncan, K. Leavitt, K. Duncan, and G. Vilchez, Detection of small for gestational age in preterm prelabor rupture of membranes by Hadlock versus the Fetal Medicine Foundation growth charts, Obstet Gynecol Sci (2021). |
|  |  | [46] | J.R. Duncan, P. Sawangkum, E.A. Hoover, M.M. Aziz, and G. Vilchez, Birthweight and Apgar at 5 minutes of life for the prediction of severe neonatal outcomes in preterm prelabor rupture of membranes, J Matern Fetal Neonatal Med (2020), 1-5. |
|  |  | [47] | J.R. Duncan, C. Schenone, K.M. Dorset, P.J. Goedecke, A.M. Tobiasz, N.L. Meyer, and M.H. Schenone, Estimated fetal weight accuracy in pregnancies with preterm prelabor rupture of membranes by the Hadlock method, J Matern Fetal Neonatal Med (2020), 1-5. |
|  |  | [48] | J.R. Duncan, A.M. Tobiasz, K.M. Dorsett, M.M. Aziz, R.E. Thompson, Z. Bursac, A.J. Talati, G. Mari, and M.H. Schenone, Fetal pulmonary artery acceleration/ejection time prognostic accuracy for respiratory complications in preterm prelabor rupture of membranes, J Matern Fetal Neonatal Med 33 (2020), 2054-2058. |
|  |  | [49] | B. Dundar, B. Dincgez Cakmak, G. Ozgen, F.N. Tasgoz, T. Guclu, and G. Ocakoglu, Platelet indices in preterm premature rupture of membranes and their relation with adverse neonatal outcomes, J Obstet Gynaecol Res 44 (2018), 67-73. |
|  |  | [50] | V. El-Achi, B. de Vries, C. O'Brien, F. Park, J. Tooher, and J. Hyett, First-Trimester Prediction of Preterm Prelabour Rupture of Membranes, Fetal Diagn Ther 47 (2020), 624-629. |
|  |  | [51] | V. El-Achi, F. Park, C. O'Brien, J. Tooher, and J. Hyett, Does low dose aspirin prescribed for risk of early onset preeclampsia reduce the prevalence of preterm prelabor rupture of membranes?, J Matern Fetal Neonatal Med 34 (2021), 618-623. |
|  |  | [52] | E. Elçi, G. Güneş Elçi, and S. Sayan, Comparison of the accuracy and reliability of the AmniSure, AMNIOQUICK, and AL-SENSE tests for early diagnosis of premature rupture of membranes, Int J Gynaecol Obstet 149 (2020), 93-97. |
|  |  | [53] | G.U. Eleje, E.C. Ezugwu, A.C. Eke, J.I. Ikechebelu, C.O. Ezeama, I.U. Ezebialu, N.O. Ojiegbe, C.C. Obiora, C.I. Okafor, G.O. Udigwe, B.O. Nwosu, and F.O. Ezugwu, Accuracy and response time of dual biomarker model of insulin-like growth factor binding protein-1/ alpha fetoprotein (Amnioquick duo+) in comparison to placental alpha-microglobulin-1 test in diagnosis of premature rupture of membranes, J Obstet Gynaecol Res 43 (2017), 825-833. |
|  |  | [54] | G.U. Eleje, E.C. Ezugwu, A.C. Eke, J.I. Ikechebelu, C.C. Obiora, N.O. Ojiegbe, I.U. Ezebialu, C.O. Ezeama, B.O. Nwosu, G.O. Udigwe, C.I. Okafor, and F.O. Ezugwu, Comparison of the duo of insulin-like growth factor binding protein-1/alpha fetoprotein (Amnioquick duo+®) and traditional clinical assessment for diagnosing premature rupture of fetal membranes, J Perinat Med 45 (2017), 105-112. |
|  |  | [55] | G.U. Eleje, E.C. Ezugwu, I.U. Ezebialu, N.O. Ojiegbe, R.O. Egeonu, C.C. Obiora, C.G. Okafor, J.I. Ikechebelu, and A.C. Eke, Performance indices of AmnioQuick Duo+ versus placental α-microglobulin-1 tests for women with prolonged premature rupture of membranes, Int J Gynaecol Obstet 144 (2019), 180-186. |
|  |  | [56] | G.U. Eleje, C.O. Ukah, I.V. Onyiaorah, E.C. Ezugwu, E.O. Ugwu, S.R. Ohayi, L.I. Eleje, R.O. Egeonu, I.U. Ezebialu, C.C. Obiora, J.T. Enebe, L.O. Ajah, C.G. Okafor, C.C. Okoro, A.O. Asogwa, D.K. Ogbuokiri, J.I. Ikechebelu, and A.C. Eke, Diagnostic value of Chorioquick for detecting chorioamnionitis in women with premature rupture of membranes, Int J Gynaecol Obstet 149 (2020), 98-105. |
|  |  | [57] | H. Erfani, S. Haeri, S.A. Shainker, A.F. Saad, R. Ruano, T.N. Dunn, A. Rezaei, S. Aalipour, A.A. Nassr, A.A. Shamshirsaz, M. Vaughn, W. Lindsley, M.H. Spiel, S.A. Shazly, E.R. Ibirogba, S.L. Clark, G.R. Saade, M.A. Belfort, and A.A. Shamshirsaz, Vasa previa: a multicenter retrospective cohort study, Am J Obstet Gynecol 221 (2019), 644.e641-644.e645. |
|  |  | [58] | S. Esin, M. Hayran, Y.A. Tohma, M. Guden, I. Alay, D. Esinler, S. Yalvac, and O. Kandemir, Estimation of fetal weight by ultrasonography after preterm premature rupture of membranes: comparison of different formulas, J Perinat Med 45 (2017), 253-266. |
|  |  | [59] | F.B. Flosi, F.C. da Silva, G.R. de Jesús, L.G.C. Velarde, and R.A.M. de Sá, Assessment of Fetal Lung Maturity Using Quantitative Ultrasound Analysis in Patients with Prelabor Rupture of Membranes, Fetal Diagn Ther 47 (2020), 636-641. |
|  |  | [60] | A. Frick, V. Kostiv, D. Vojtassakova, R. Akolekar, and K.H. Nicolaides, Comparison of different methods of measuring angle of progression in prediction of labor outcome, Ultrasound in Obstetrics & Gynecology 55 (2020), 391-400. |
|  |  | [61] | X.H. Fu, X.B. He, D.F. Sun, Q.H. Fan, B.L. Zhang, Q. Wang, Y.F. Wang, and H.L. Chen, Histological chorioamnionitis could be predicted with high accuracy in preterm labor with intact membranes before delivery, Clin Exp Obstet Gynecol 45 (2018), 720-724. |
|  |  | [62] | K. Fuwa, M. Seki, Y. Hirata, I. Yanagihara, Y. Nakura, C. Takano, K. Kuroda, and S. Hayakawa, Rapid and simple detection of Ureaplasma species from vaginal swab samples using a loop-mediated isothermal amplification method, Am J Reprod Immunol 79 (2018). |
|  |  | [63] | A. Gámez-Varela, M. Martínez-Rodríguez, H. López-Briones, J. Luna-García, E. Chávez-González, R. Villalobos-Gómez, E. Hernandez-Andrade, and R. Cruz-Martínez, Preoperative Cervical Length Predicts the Risk of Delivery within One Week after Pleuroamniotic Shunt in Fetuses with Severe Hydrothorax, Fetal Diagn Ther (2021), 1-7. |
|  |  | [64] | N. Garry, O. Keenan, S.W. Lindow, and T. Darcy, Pregnancy outcomes following elective abdominal cerclage following cervical excision surgery for neoplastic disease, European Journal of Obstetrics & Gynecology and Reproductive Biology 256 (2021), 225-229. |
|  |  | [65] | C. Gezer, A. Ekin, C. Golbasi, C. Kocahakimoglu, U. Bozkurt, A. Dogan, U. Solmaz, H. Golbasi, and C.E. Taner, Use of urea and creatinine levels in vaginal fluid for the diagnosis of preterm premature rupture of membranes and delivery interval after membrane rupture, J Matern Fetal Neonatal Med 30 (2017), 772-778. |
|  |  | [66] | L.L. Gievers, J. Sedler, C.A. Phillipi, D. Dukhovny, J. Geddes, P. Graven, B. Chan, and S. Khaki, Implementation of the sepsis risk score for chorioamnionitis-exposed newborns, J Perinatol 38 (2018), 1581-1587. |
|  |  | [67] | M. Gioan, F. Fenollar, A. Loundou, J.P. Menard, J. Blanc, C. D'Ercole, and F. Bretelle, Development of a nomogram for individual preterm birth risk evaluation, J Gynecol Obstet Hum Reprod 47 (2018), 545-548. |
|  |  | [68] | P. Girón de Velasco-Sada, I. Falces-Romero, I. Quiles-Melero, A. García-Perea, and J. Mingorance, Evaluation of two real time PCR assays for the detection of bacterial DNA in amniotic fluid, J Microbiol Methods 144 (2018), 107-110. |
|  |  | [69] | L. Goodfellow, A. Care, A. Sharp, J. Ivandic, B. Poljak, D. Roberts, and Z. Alfirevic, Effect of QUiPP prediction algorithm on treatment decisions in women with a previous preterm birth: a prospective cohort study, Bjog 126 (2019), 1569-1575. |
|  |  | [70] | S. Gupta, H.S. Gaikwad, B. Nath, and A. Batra, Can vitamin C and interleukin 6 levels predict preterm premature rupture of membranes: evaluating possibilities in North Indian population, Obstet Gynecol Sci 63 (2020), 432-439. |
|  |  | [71] | R.B. Helmig and J.B. Gertsen, Diagnostic accuracy of polymerase chain reaction for intrapartum detection of group B streptococcus colonization, Acta Obstet Gynecol Scand 96 (2017), 1070-1074. |
|  |  | [72] | A. Hesson and E. Langen, Outcomes in oligohydramnios: the role of etiology in predicting pulmonary morbidity/mortality, J Perinat Med 46 (2018), 948-950. |
|  |  | [73] | L. Hiersch, E. Krispin, A. Aviram, M. Mor-Shacham, R. Gabbay-Benziv, Y. Yogev, and E. Ashwal, Predictors for prolonged interval from premature rupture of membranes to spontaneous onset of labor at term, J Matern Fetal Neonatal Med 30 (2017), 1465-1470. |
|  |  | [74] | Y. Horikoshi, C. Yaguchi, N. Furuta-Isomura, T. Itoh, K. Kawai, T. Oda, M. Matsumoto, Y. Kohmura-Kobayashi, N. Tamura, T. Uchida, N. Kanayama, and H. Itoh, Gross appearance of the fetal membrane on the placental surface is associated with histological chorioamnionitis and neonatal respiratory disorders, PLoS One 15 (2020), e0242579. |
|  |  | [75] | T. Horinouchi, T. Yoshizato, Y. Kozuma, T. Shinagawa, M. Muto, T. Yamasaki, D. Hori, and K. Ushijima, Prediction of histological chorioamnionitis and neonatal and infantile outcomes using procalcitonin in the umbilical cord blood and amniotic fluid at birth, J Obstet Gynaecol Res 44 (2018), 630-636. |
|  |  | [76] | K. Hughes, S. Sim, A. Roman, K. Michalak, S. Kane, and P. Sheehan, Outcomes and predictive tests from a dedicated specialist clinic for women at high risk of preterm labour: A ten year audit, Aust N Z J Obstet Gynaecol 57 (2017), 405-411. |
|  |  | [77] | K.M. Hughes, S.C. Kane, T.P. Haines, and P.M. Sheehan, Cervical length surveillance for predicting spontaneous preterm birth in women with uterine anomalies: A cohort study, Acta Obstet Gynecol Scand 99 (2020), 1519-1526. |
|  |  | [78] | A. Husain, A. Naseem, S. Anjum, S. Imran, M. Arifuzzaman, and S.O. Adil, Predictability of intrapartum cardiotocography with meconium stained liquor and its correlation with perinatal outcome, J Pak Med Assoc 68 (2018), 1014-1018. |
|  |  | [79] | I. Igbinosa, F.A. Moore, 3rd, C. Johnson, and J.E. Block, Comparison of rapid immunoassays for rupture of fetal membranes, BMC Pregnancy Childbirth 17 (2017), 128. |
|  |  | [80] | J.K. Iversen, B.H. Kahrs, E.A. Torkildsen, and T.M. Eggebo, Fetal molding examined with transperineal ultrasound and associations with position and delivery mode, Am J Obstet Gynecol 223 (2020). |
|  |  | [81] | M. Jain, A. Bang, A. Tiwari, and S. Jain, Prediction of significant hyperbilirubinemia in term neonates by early non-invasive bilirubin measurement, World J Pediatr 13 (2017), 222-227. |
|  |  | [82] | P. Janku, M. Kacerovsky, B. Zednikova, C. Andrys, M. Kolackova, M. Drahosova, L. Pliskova, H. Zemlickova, R. Gerychova, O. Simetka, P. Matlak, B. Jacobsson, and I. Musilova, Pentraxin 3 in Noninvasively Obtained Cervical Fluid Samples from Pregnancies Complicated by Preterm Prelabor Rupture of Membranes, Fetal Diagn Ther 46 (2019), 402-410. |
|  |  | [83] | F. Jochum, C. Le Ray, P. Blanc-Petitjean, B. Langer, N. Meyer, F. Severac, and N. Sananes, Externally Validated Score to Predict Cesarean Delivery After Labor Induction With Cervi Ripening, Obstet Gynecol 134 (2019), 502-510. |
|  |  | [84] | M. Julien, L. N'Guema, A. Bouzerara, B. Toro, E. Lecarpentier, and J. Guibourdenche, Premature rupture of the membranes: analytical evaluation of diagnostic tests, Ann Biol Clin (Paris) 76 (2018), 300-306. |
|  |  | [85] | E.Y. Jung, K.H. Park, B.R. Han, S.H. Cho, and A. Ryu, Measurement of Interleukin 8 in Cervicovaginal Fluid in Women With Preterm Premature Rupture of Membranes: A Comparison of Amniotic Fluid Samples, Reproductive Sciences 24 (2017), 142-147. |
|  |  | [86] | S. Jwala, T.L. Tran, C. Terenna, A. McGregor, J. Andrel, B.E. Leiby, J.K. Baxter, and V. Berghella, Evaluation of additive effect of quantitative fetal fibronectin to cervical length for prediction of spontaneous preterm birth among asymptomatic low-risk women, Acta Obstet Gynecol Scand 95 (2016), 948-955. |
|  |  | [87] | M. Kacerovsky, I. Musilova, T. Bestvina, M. Stepan, T. Cobo, and B. Jacobsson, Preterm Prelabor Rupture of Membranes between 34 and 37 Weeks: A Point-of-Care Test of Vaginal Fluid Interleukin-6 Concentrations for a Noninvasive Detection of Intra-Amniotic Inflammation, Fetal Diagn Ther 43 (2018), 175-183. |
|  |  | [88] | M. Kaneko, M. Sato, K. Ogasawara, T. Imamura, K. Hashimoto, N. Momoi, and M. Hosoya, Serum cytokine concentrations, chorioamnionitis and the onset of bronchopulmonary dysplasia in premature infants, J Neonatal Perinatal Med 10 (2017), 147-155. |
|  |  | [89] | H. Kansu-Celik, Y. Tasci, B.K. Karakaya, M. Cinar, T. Candar, and G.S. Caglar, Maternal serum advanced glycation end products level as an early marker for predicting preterm labor/PPROM: a prospective preliminary study, J Matern Fetal Neonatal Med 32 (2019), 2758-2762. |
|  |  | [90] | V. Kathir, D. Maurya, and A. Keepanasseril, Transvaginal sonographic assessment of cervix in prediction of admission to delivery interval in preterm premature rupture of membranes, J Matern Fetal Neonatal Med 31 (2018), 2717-2720. |
|  |  | [91] | T. Kawakita, U.M. Reddy, J.C. Huang, T.C. Auguste, D. Bauer, and R.T. Overcash, Externally Validated Prediction Model of Vaginal Delivery After Preterm Induction With Unfavorable Cervix, Obstet Gynecol 136 (2020), 716-724. |
|  |  | [92] | B. Kaya, U. Turhan, S. Sezer, S. Kaya, İ. Dağ, and A. Tayyar, Maternal serum galectin-1 and galectin-3 levels in pregnancies complicated with preterm prelabor rupture of membranes, J Matern Fetal Neonatal Med 33 (2020), 861-868. |
|  |  | [93] | G. Kayem, F. Batteux, N. Girard, T. Schmitz, M. Willaime, F. Maillard, P.H. Jarreau, and F. Goffinet, Predictive value of vaginal IL-6 and TNFα bedside tests repeated until delivery for the prediction of maternal-fetal infection in cases of premature rupture of membranes, Eur J Obstet Gynecol Reprod Biol 211 (2017), 8-14. |
|  |  | [94] | A. Kesrouani, E. Chalhoub, E. El Rassy, M. Germanos, A. Khazzaka, J. Rizkallah, E. Attieh, and N. Aouad, Prediction of preterm delivery by second trimester inflammatory biomarkers in the amniotic fluid, Cytokine 85 (2016), 67-70. |
|  |  | [95] | H.J. Kim, K.H. Park, Y.M. Kim, E. Joo, K. Ahn, and S. Shin, A protein microarray analysis of amniotic fluid proteins for the prediction of spontaneous preterm delivery in women with preterm premature rupture of membranes at 23 to 30 weeks of gestation, PLoS One 15 (2020), e0244720. |
|  |  | [96] | Y.M. Kim, J.Y. Park, J.H. Sung, S.J. Choi, S.Y. Oh, C.R. Roh, and J.H. Kim, Predicting factors for success of vaginal delivery in preterm induction with prostaglandin E(2), Obstet Gynecol Sci 60 (2017), 163-169. |
|  |  | [97] | V. Kiver, V. Boos, A. Thomas, W. Henrich, and A. Weichert, Perinatal outcomes after previable preterm premature rupture of membranes before 24 weeks of gestation, J Perinat Med 46 (2018), 555-565. |
|  |  | [98] | S.Y. Kook, K.H. Park, J.A. Jang, Y.M. Kim, H. Park, and S.J. Jeon, Vitamin D-binding protein in cervicovaginal fluid as a non-invasive predictor of intra-amniotic infection and impending preterm delivery in women with preterm labor or preterm premature rupture of membranes, PLoS One 13 (2018), e0198842. |
|  |  | [99] | P. Kosinski and M. Wielgos, Foetoscopic endotracheal occlusion (FETO) for severe isolated left-sided congenital diaphragmatic hernia: single center Polish experience, Journal of Maternal-Fetal & Neonatal Medicine 31 (2018), 2521-2526. |
|  |  | [100] | C.L. Kothari, C. Romph, T. Bautista, and D. Lenz, Perinatal Periods of Risk Analysis: Disentangling Race and Socioeconomic Status to Inform a Black Infant Mortality Community Action Initiative, Matern Child Health J 21 (2017), 49-58. |
|  |  | [101] | K. Kuhrt, N. Hezelgrave, C. Foster, P.T. Seed, and A.H. Shennan, Development and validation of a tool incorporating quantitative fetal fibronectin to predict spontaneous preterm birth in symptomatic women, Ultrasound in Obstetrics and Gynecology 47 (2016), 210-216. |
|  |  | [102] | K. Kuhrt, E. Smout, N. Hezelgrave, P.T. Seed, J. Carter, and A.H. Shennan, Development and validation of a tool incorporating cervical length and quantitative fetal fibronectin to predict spontaneous preterm birth in asymptomatic high-risk women, Ultrasound in Obstetrics and Gynecology 47 (2016), 104-109. |
|  |  | [103] | M. Kunze, M. Klar, C.A. Morfeld, B. Thorns, R.L. Schild, F. Markfeld-Erol, R. Rasenack, H. Proempeler, R. Hentschel, and W.R. Schaefer, Cytokines in noninvasively obtained amniotic fluid as predictors of fetal inflammatory response syndrome, Am J Obstet Gynecol 215 (2016), 96.e91-98. |
|  |  | [104] | N.B. Kunzier, W.L. Kinzler, M.R. Chavez, T.M. Adams, D.A. Brand, and A.M. Vintzileos, The use of cervical sonography to differentiate true from false labor in term patients presenting for labor check, Am J Obstet Gynecol 215 (2016), 372.e371-375. |
|  |  | [105] | M. Kurakazu, F. Yotsumoto, H. Arima, D. Izuchi, D. Urushiyama, K. Miyata, C. Kiyoshima, S. Fukagawa, K. Yoshikawa, M. Kurakazu, T. Hirakawa, K. Shigekawa, R. Araki, A. Sanui, M. Murata, K. Nabeshima, and S. Miyamoto, The combination of maternal blood and amniotic fluid biomarkers improves the predictive accuracy of histologic chorioamnionitis, Placenta 80 (2019), 4-7. |
|  |  | [106] | M. Kurek Eken, A. Tüten, E. Özkaya, G. Karatekin, and A. Karateke, Major determinants of survival and length of stay in the neonatal intensive care unit of newborns from women with premature preterm rupture of membranes, J Matern Fetal Neonatal Med 30 (2017), 1972-1975. |
|  |  | [107] | C.A. Labarrere, H.L. DiCarlo, E. Bammerlin, J.W. Hardin, Y.M. Kim, P. Chaemsaithong, D.M. Haas, G.S. Kassab, and R. Romero, Failure of physiologic transformation of spiral arteries, endothelial and trophoblast cell activation, and acute atherosis in the basal plate of the placenta, Am J Obstet Gynecol 216 (2017), 287.e281-287.e216. |
|  |  | [108] | S.M. Lee, K.H. Park, E.Y. Jung, J.A. Jang, and H.N. Yoo, Frequency and clinical significance of short cervix in patients with preterm premature rupture of membranes, PLoS One 12 (2017), e0174657. |
|  |  | [109] | S.M. Lee, K.H. Park, E.Y. Jung, S.Y. Kook, H. Park, and S.J. Jeon, Inflammatory proteins in maternal plasma, cervicovaginal and amniotic fluids as predictors of intra-amniotic infection in preterm premature rupture of membranes, PLoS One 13 (2018), e0200311. |
|  |  | [110] | Y.J. Lee, S.C. Kim, J.K. Joo, D.H. Lee, K.H. Kim, and K.S. Lee, Amniotic fluid index, single deepest pocket and transvaginal cervical length: Parameter of predictive delivery latency in preterm premature rupture of membranes, Taiwan J Obstet Gynecol 57 (2018), 374-378. |
|  |  | [111] | F. Lemoine, R.C. Moore, A. Chapple, F.A. Moore, and E. Sutton, Neonatal Survivability following Previable PPROM after Hospital Readmission for Intervention, AJP Reports 10 (2020), E395-E402. |
|  |  | [112] | J. Li, B. Yu, W. Wang, D. Luo, Q.L. Dai, and X.Q. Gan, Does intact umbilical cord milking increase infection rates in preterm infants with premature prolonged rupture of membranes?, Journal of Maternal-Fetal & Neonatal Medicine 33 (2020), 184-190. |
|  |  | [113] | W.F. Li, A.S. Chao, S.D. Chang, P.J. Cheng, L.Y. Yang, and Y.L. Chang, Effects and outcomes of septostomy in twin-to-twin transfusion syndrome after fetoscopic laser therapy, BMC Pregnancy Childbirth 19 (2019), 397. |
|  |  | [114] | H. Liang, Z. Xie, B. Liu, X. Song, and G. Zhao, A routine urine test has partial predictive value in premature rupture of the membranes, J Int Med Res 47 (2019), 2361-2370. |
|  |  | [115] | L. Liu, H. Wang, Y. Zhang, J. Niu, Z. Li, and R. Tang, Effect of pregravid obesity on perinatal outcomes in singleton pregnancies following in vitro fertilization and the weight-loss goals to reduce the risks of poor pregnancy outcomes: A retrospective cohort study, PLoS One 15 (2020), e0227766. |
|  |  | [116] | Y. Liu, Y. Liu, C. Du, R. Zhang, Z. Feng, and J. Zhang, Diagnostic value of amniotic fluid inflammatory biomarkers for subclinical chorioamnionitis, Int J Gynaecol Obstet 134 (2016), 160-164. |
|  |  | [117] | K. Lyubomirskaya, Y. Krut, L. Sergeyeva, S. Khmil, O. Lototska, N. Petrenko, and A. Kamyshnyi, Preterm premature rupture of membranes: prediction of risks in women of Zaporizhzhia region of Ukraine, Pol Merkur Lekarski 48 (2020), 399-405. |
|  |  | [118] | G.C. Ma, Y.C. Chen, W.J. Wu, S.P. Chang, T.Y. Chang, W.H. Lin, and M. Chen, Prenatal Diagnosis of Autosomal Recessive Renal Tubular Dysgenesis with Anhydramnios Caused by a Mutation in the AGT Gene, Diagnostics 9 (2019). |
|  |  | [119] | Y. Ma, Y. Xu, L. Jiang, and X. Shao, Application of a Prediction Model Based on the Laboratory Index Score in Prelabor Rupture of Membranes with Histologic Chorioamnionitis During Late Pregnancy, Med Sci Monit 26 (2020), e924756. |
|  |  | [120] | P. Mach, A. Köninger, L. Wicherek, R. Kimmig, S. Kasimir-Bauer, C. Birdir, B. Schmidt, and A. Gellhaus, Serum concentrations of soluble B7-H4 in early pregnancy are elevated in women with preterm premature rupture of fetal membranes, Am J Reprod Immunol 76 (2016), 149-154. |
|  |  | [121] | Y. Maki, S. Furukawa, T. Nakayama, M. Oohashi, N. Shiiba, K. Furuta, S. Tokunaga, and H. Sameshima, Clinical chorioamnionitis criteria are not sufficient for predicting intra-amniotic infection, J Matern Fetal Neonatal Med (2020), 1-6. |
|  |  | [122] | R.J. Martinez-Portilla, A. Hawkins-Villarreal, P. Alvarez-Ponce, Z.L. Chinolla-Arellano, A.L. Moreno-Espinosa, A.L. Sandoval-Mejia, and N. Moreno-Uribe, Maternal Serum Interleukin-6: A Non-Invasive Predictor of Histological Chorioamnionitis in Women with Preterm-Prelabor Rupture of Membranes, Fetal Diagn Ther 45 (2019), 168-175. |
|  |  | [123] | Y. Mei and Y. Lin, Clinical significance of primary symptoms in women with placental abruption, J Matern Fetal Neonatal Med 31 (2018), 2446-2449. |
|  |  | [124] | M. Merello, L. Lotte, S. Gonfrier, S. Eleni Dit Trolli, F. Casagrande, R. Ruimy, and A. Bongain, Enterobacteria vaginal colonization among patients with preterm premature rupture of membranes from 24 to 34 weeks of gestation and neonatal infection risk, J Gynecol Obstet Hum Reprod 48 (2019), 187-191. |
|  |  | [125] | M. Mikołajczyk, P. Wirstlein, M. Adamczyk, J. Skrzypczak, and E. Wender-Ożegowska, Value of cervicovaginal fluid cytokines in prediction of fetal inflammatory response syndrome in pregnancies complicated with preterm premature rupture of membranes (pPROM), J Perinat Med 48 (2020), 249-255. |
|  |  | [126] | H. Miremberg, N. Elia, C. Marelly, O. Gluck, G. Barda, J. Bar, and E. Weiner, Should the interval between doses of antenatal corticosteroids be shortened in certain cases? Factors predicting preterm delivery < 48 h from presentation, Arch Gynecol Obstet (2021). |
|  |  | [127] | S. Mishra, S. Jaiswar, S. Saad, S. Tripathi, N. Singh, S. Deo, M. Agarwal, and N. Mishra, Platelet indices as a predictive marker in neonatal sepsis and respiratory distress in preterm prelabor rupture of membranes, Int J Hematol 113 (2021), 199-206. |
|  |  | [128] | B.P. Modi, H.I. Parikh, M.E. Teves, R. Kulkarni, J. Liyu, R. Romero, T.P. York, and J.F. Strauss, 3rd, Discovery of rare ancestry-specific variants in the fetal genome that confer risk of preterm premature rupture of membranes (PPROM) and preterm birth, BMC Med Genet 19 (2018), 181. |
|  |  | [129] | A.F. Moron, A. Athanasiou, M. Barbosa, H.J. Milani, S.G. Sarmento, S. Cavalheiro, and S.S. Witkin, Amniotic fluid lactic acid and matrix metalloproteinase-8 levels at the time of fetal surgery for a spine defect: association with subsequent preterm prelabour rupture of membranes, Bjog 125 (2018), 1288-1292. |
|  |  | [130] | L.J. Moulton, J. Eric Jelovsek, M. Lachiewicz, K. Chagin, and O. Goje, A model to predict risk of postpartum infection after Caesarean delivery, J Matern Fetal Neonatal Med 31 (2018), 2409-2417. |
|  |  | [131] | A.S. Mousavi, N. Hashemi, M. Kashanian, N. Sheikhansari, A. Bordbar, and S. Parashi, Comparison between maternal and neonatal outcome of PPROM in the cases of amniotic fluid index (AFI) of more and less than 5 cm, J Obstet Gynaecol 38 (2018), 611-615. |
|  |  | [132] | I. Musilova, C. Andrys, M. Drahosova, O. Soucek, R. Kutova, L. Pliskova, R. Spacek, P. Laudanski, B. Jacobsson, and M. Kacerovsky, Amniotic fluid calreticulin in pregnancies complicated by the preterm prelabor rupture of membranes, Journal of Maternal-Fetal and Neonatal Medicine 29 (2016), 3921-3929. |
|  |  | [133] | I. Musilova, C. Andrys, M. Drahosova, O. Soucek, L. Pliskova, B. Jacobsson, and M. Kacerovsky, Cervical fluid interleukin 6 and intra-amniotic complications of preterm prelabor rupture of membranes, Journal of Maternal-Fetal & Neonatal Medicine 31 (2018), 827-836. |
|  |  | [134] | I. Musilova, C. Andrys, M. Drahosova, O. Soucek, L. Pliskova, M. Stepan, T. Bestvina, J. Maly, B. Jacobsson, and M. Kacerovsky, Amniotic fluid cathepsin-G in pregnancies complicated by the preterm prelabor rupture of membranes, Journal of Maternal-Fetal and Neonatal Medicine 30 (2017), 2097-2104. |
|  |  | [135] | I. Musilova, C. Andrys, M. Holeckova, V. Kolarova, L. Pliskova, M. Drahosova, R. Bolehovska, R. Pilka, K. Huml, T. Cobo, B. Jacobsson, and M. Kacerovsky, Interleukin-6 measured using the automated electrochemiluminescence immunoassay method for the identification of intra-amniotic inflammation in preterm prelabor rupture of membranes, J Matern Fetal Neonatal Med 33 (2020), 1919-1926. |
|  |  | [136] | I. Musilova, C. Andrys, H. Hornychova, L. Pliskova, M. Drahosova, B. Zednikova, R. Bolehovska, T. Faist, B. Jacobsson, and M. Kacerovsky, Gastric fluid used to assess changes during the latency period in preterm prelabor rupture of membranes, Pediatr Res 84 (2018), 240-247. |
|  |  | [137] | I. Musilova, T. Bestvina, M. Hudeckova, I. Michalec, T. Cobo, B. Jacobsson, and M. Kacerovsky, Vaginal fluid interleukin-6 concentrations as a point-of-care test is of value in women with preterm prelabor rupture of membranes, Am J Obstet Gynecol 215 (2016), 619.e611-619.e612. |
|  |  | [138] | I. Musilova, T. Bestvina, M. Hudeckova, I. Michalec, T. Cobo, B. Jacobsson, and M. Kacerovsky, Vaginal fluid IL-6 concentrations as a point-of-care test is of value in women with preterm PROM, Am J Obstet Gynecol (2016). |
|  |  | [139] | I. Musilova, M. Kacerovsky, M. Stepan, T. Bestvina, L. Pliskova, B. Zednikova, and B. Jacobsson, Maternal serum C-reactive protein concentration and intra-amniotic inflammation in women with preterm prelabor rupture of membranes, PLoS One 12 (2017), e0182731. |
|  |  | [140] | I. Musilova, L. Pliskova, R. Gerychova, P. Janku, O. Simetka, P. Matlak, B. Jacobsson, and M. Kacerovsky, Maternal white blood cell count cannot identify the presence of microbial invasion of the amniotic cavity or intra-amniotic inflammation in women with preterm prelabor rupture of membranes, PLoS One 12 (2017). |
|  |  | [141] | T. Myntti, L. Rahkonen, A. Pätäri-Sampo, M. Tikkanen, T. Sorsa, J. Juhila, O. Helve, S. Andersson, J. Paavonen, and V. Stefanovic, Comparison of amniotic fluid matrix metalloproteinase-8 and cathelicidin in the diagnosis of intra-amniotic infection, J Perinatol 36 (2016), 1049-1054. |
|  |  | [142] | M. Nakahara, S. Goto, E. Kato, S. Nojiri, A. Itakura, and S. Takeda, Maternal risk score for the prediction of fetal inflammatory response syndrome after preterm premature rupture of membranes, J Obstet Gynaecol Res 46 (2020), 2019-2026. |
|  |  | [143] | G. Natarajan, S. Shankaran, S. Saha, A. Laptook, A. Das, R. Higgins, B.J. Stoll, E.F. Bell, W.A. Carlo, C. D'Angio, S.B. DeMauro, P. Sanchez, K. Van Meurs, B. Vohr, N. Newman, E. Hale, and M. Walsh, Antecedents and Outcomes of Abnormal Cranial Imaging in Moderately Preterm Infants, J Pediatr 195 (2018), 66-72.e63. |
|  |  | [144] | D. Odd, A. Heep, K. Luyt, and T. Draycott, Hypoxic-ischemic brain injury: Planned delivery before intrapartum events, J Neonatal Perinatal Med 10 (2017), 347-353. |
|  |  | [145] | K.J. Oh, J. Lee, R. Romero, H.S. Park, J.S. Hong, and B.H. Yoon, A new rapid bedside test to diagnose and monitor intraamniotic inflammation in preterm PROM using transcervically collected fluid, Am J Obstet Gynecol 223 (2020), 423.e421-423.e415. |
|  |  | [146] | S. Ona, S.R. Easter, M. Prabhu, G. Wilkie, R.E. Tuomala, L.E. Riley, and K. Diouf, Diagnostic Validity of the Proposed Eunice Kennedy Shriver National Institute of Child Health and Human Development Criteria for Intrauterine Inflammation or Infection, Obstet Gynecol 133 (2019), 33-39. |
|  |  | [147] | A. Ozel, E. Alici Davutoglu, A. Yurtkal, and R. Madazli, How do platelet-to-lymphocyte ratio and neutrophil-to-lymphocyte ratio change in women with preterm premature rupture of membranes, and threaten preterm labour?, J Obstet Gynaecol 40 (2020), 195-199. |
|  |  | [148] | P. Pacora, R. Romero, O. Erez, E. Maymon, B. Panaitescu, J.P. Kusanovic, A.L. Tarca, C.D. Hsu, and S.S. Hassan, The diagnostic performance of the beta-glucan assay in the detection of intra-amniotic infection with Candida species, J Matern Fetal Neonatal Med 32 (2019), 1703-1720. |
|  |  | [149] | T. Paramel Jayaprakash, E.C. Wagner, J. van Schalkwyk, A.Y. Albert, J.E. Hill, and D.M. Money, High Diversity and Variability in the Vaginal Microbiome in Women following Preterm Premature Rupture of Membranes (PPROM): A Prospective Cohort Study, PLoS One 11 (2016), e0166794. |
|  |  | [150] | H. Park, K.H. Park, Y.M. Kim, S.Y. Kook, S.J. Jeon, and H.N. Yoo, Plasma inflammatory and immune proteins as predictors of intra-amniotic infection and spontaneous preterm delivery in women with preterm labor: a retrospective study, BMC Pregnancy Childbirth 18 (2018), 146. |
|  |  | [151] | J.W. Park, K.H. Park, S.Y. Kook, Y.M. Jung, and Y.M. Kim, Immune biomarkers in maternal plasma to identify histologic chorioamnionitis in women with preterm labor, Arch Gynecol Obstet 299 (2019), 725-732. |
|  |  | [152] | J.W. Park, K.H. Park, J.E. Lee, Y.M. Kim, S.J. Lee, and D.H. Cheon, Antibody Microarray Analysis of Plasma Proteins for the Prediction of Histologic Chorioamnionitis in Women With Preterm Premature Rupture of Membranes, Reprod Sci 26 (2019), 1476-1484. |
|  |  | [153] | S. Park, Y.A. You, H. Yun, S.J. Choi, H.S. Hwang, S.K. Choi, S.M. Lee, and Y.J. Kim, Cervicovaginal fluid cytokines as predictive markers of preterm birth in symptomatic women, Obstet Gynecol Sci 63 (2020), 455-463. |
|  |  | [154] | A.S. Patil, N.W. Gaikwad, C.A. Grotegut, S.D. Dowden, and D.M. Haas, Alterations in endogenous progesterone metabolism associated with spontaneous very preterm delivery, Hum Reprod Open 2020 (2020), hoaa007. |
|  |  | [155] | M.S. Payne, J.P. Newnham, D.A. Doherty, L.L. Furfaro, N.L. Pendal, D.E. Loh, and J.A. Keelan, A specific bacterial DNA signature in the vagina of Australian women in midpregnancy predicts high risk of spontaneous preterm birth (the Predict1000 study), Am J Obstet Gynecol 224 (2021), 206.e201-206.e223. |
|  |  | [156] | C. Petit, P. Deruelle, H. Behal, T. Rakza, S. Balagny, D. Subtil, E. Clouqueur, and C. Garabedian, Preterm premature rupture of membranes: Which criteria contraindicate home care management?, Acta Obstet Gynecol Scand 97 (2018), 1499-1507. |
|  |  | [157] | A. Pinton, F. Severac, N. Meyer, C.Y. Akladios, A. Gaudineau, R. Favre, B. Langer, and N. Sananes, A comparison of vaginal ultrasound and digital examination in predicting preterm delivery in women with threatened preterm labor: a cohort study, Acta Obstet Gynecol Scand 96 (2017), 447-453. |
|  |  | [158] | I.H. Polat, S. Marin, J. Ríos, M. Larroya, A.B. Sánchez-García, C. Murillo, C. Rueda, M. Cascante, E. Gratacós, and T. Cobo, Exploratory and confirmatory analysis to investigate the presence of vaginal metabolome expression of microbial invasion of the amniotic cavity in women with preterm labor using high-performance liquid chromatography, Am J Obstet Gynecol 224 (2021), 90.e91-90.e99. |
|  |  | [159] | N. Pruksanusak, P. Thongphanang, T. Suntharasaj, C. Suwanrath, and A. Geater, Combined maternal-associated risk factors with intrapartum fetal heart rate classification systems to predict peripartum asphyxia neonates, Eur J Obstet Gynecol Reprod Biol 218 (2017), 85-91. |
|  |  | [160] | G. Raba, M. Kacerovsky, and P. Laudański, Eotaxin-2 as a potential marker of preterm premature rupture of membranes: A prospective, cohort, multicenter study, Adv Clin Exp Med 30 (2021), 197-202. |
|  |  | [161] | A.M. Ramseyer, J.R. Whittington, E.F. Magann, J.A. Pates, and S.T. Ounpraseuth, Can maternal characteristics on admission for preterm prelabor rupture of membranes predict pregnancy latency?, Am J Obstet Gynecol MFM 2 (2020), 100194. |
|  |  | [162] | S. Rasmussen, C. Ebbing, and L.M. Irgens, Predicting preeclampsia from a history of preterm birth, PLoS One 12 (2017), e0181016. |
|  |  | [163] | R. Revello, M.J. Alcaide, D. Abehsera, M. Martin-Camean, E.F.G.M. Sousa, B. Alonso-Luque, and J.L. Bartha, Prediction of chorioamnionitis in cases of intraamniotic infection by ureaplasma urealyticum in women with very preterm premature rupture of membranes or preterm labour, J Matern Fetal Neonatal Med 31 (2018), 1839-1844. |
|  |  | [164] | M. Rewatkar, S. Jain, M. Jain, and K. Mohod, C-reactive protein and white blood cell count as predictors of maternal and neonatal infections in prelabour rupture of membranes between 34 and 41 weeks of gestation, J Obstet Gynaecol 38 (2018), 622-628. |
|  |  | [165] | M. Roethlisberger, B. Strizek, I. Gottschalk, M.R. Mallmann, A. Geipel, U. Gembruch, and C. Berg, First-trimester intervention in twin reversed arterial perfusion sequence: does size matter?, Ultrasound Obstet Gynecol 50 (2017), 40-44. |
|  |  | [166] | L.C. Rogers, L. Scott, and J.E. Block, Accurate Point-of-Care Detection of Ruptured Fetal Membranes: Improved Diagnostic Performance Characteristics with a Monoclonal/Polyclonal Immunoassay, Clin Med Insights Reprod Health 10 (2016), 15-18. |
|  |  | [167] | S. Ronzoni, R. D'Souza, O. Shynlova, S. Lye, and K.E. Murphy, Maternal blood endotoxin activity in pregnancies complicated by preterm premature rupture of membranes, J Matern Fetal Neonatal Med 32 (2019), 3473-3479. |
|  |  | [168] | S. Ronzoni, V. Steckle, R. D'Souza, K.E. Murphy, S. Lye, and O. Shynlova, Cytokine Changes in Maternal Peripheral Blood Correlate With Time-to-Delivery in Pregnancies Complicated by Premature Prelabor Rupture of the Membranes, Reprod Sci 26 (2019), 1266-1276. |
|  |  | [169] | M. Rouzaire, M. Corvaisier, V. Roumeau, A. Mulliez, F. Sendy, A. Delabaere, and D. Gallot, Predictors of Short Latency Period Exceeding 48 h after Preterm Premature Rupture of Membranes, J Clin Med 10 (2021). |
|  |  | [170] | H.K. Ryu, J.H. Moon, H.J. Heo, J.W. Kim, and Y.H. Kim, Maternal c-reactive protein and oxidative stress markers as predictors of delivery latency in patients experiencing preterm premature rupture of membranes, Int J Gynaecol Obstet 136 (2017), 145-150. |
|  |  | [171] | M. Sahni, M.E. Franco-Fuenmayor, and K. Shattuck, Management of Late Preterm and Term Neonates exposed to maternal Chorioamnionitis, BMC Pediatr 19 (2019), 282. |
|  |  | [172] | T. Samejima, T. Nagamatsu, D.J. Schust, N. Itaoka, T. Iriyama, T. Nakayama, A. Komatsu, K. Kawana, Y. Osuga, and T. Fujii, Elevated concentration of secretory leukocyte protease inhibitor in the cervical mucus before delivery, Am J Obstet Gynecol 214 (2016), 741.e741-747. |
|  |  | [173] | T. Samejima and K. Takechi, Elevated C-reactive protein levels in histological chorioamnionitis at term: impact of funisitis on term neonates, J Matern Fetal Neonatal Med 30 (2017), 1428-1433. |
|  |  | [174] | J.B. Sauberan, B. Choi, A.R. Paradyse, and J. Le, Quality and Clinical Outcomes Associated with a Gentamicin Use System Change for Managing Chorioamnionitis, J Med Syst 41 (2017), 202. |
|  |  | [175] | W.A. Sayed Ahmed, M.R. Ahmed, M.L. Mohamed, M.A. Hamdy, Z. Kamel, and K.M. Elnahas, Maternal serum interleukin-6 in the management of patients with preterm premature rupture of membranes, J Matern Fetal Neonatal Med 29 (2016), 3162-3166. |
|  |  | [176] | G. Seliger, M. Bergner, R. Haase, H. Stepan, E. Schleußner, J. Zöllkau, S. Seeger, F.B. Kraus, G.G.R. Hiller, A. Wienke, and M. Tchirikov, Daily monitoring of vaginal interleukin 6 as a predictor of intraamniotic inflammation after preterm premature rupture of membranes - a new method of sampling studied in a prospective multicenter trial, J Perinat Med (2021). |
|  |  | [177] | S. Shinar, S. Agrawal, D. El-Chaar, N. Abbasi, R. Beecroft, J. Kachura, J. Keunen, G. Seaward, T. Van Mieghem, R. Windrim, and G. Ryan, Selective fetal reduction in complicated monochorionic twin pregnancies: A comparison of techniques, Prenat Diagn 41 (2021), 52-60. |
|  |  | [178] | W.H. Sim, H. Ng, and P. Sheehan, Maternal and neonatal outcomes following expectant management of preterm prelabor rupture of membranes before viability, J Matern Fetal Neonatal Med 33 (2020), 533-541. |
|  |  | [179] | G. Sisti, S. Paccosi, A. Parenti, V. Seravalli, M. Di Tommaso, and S.S. Witkin, Insulin-like growth factor binding protein-1 predicts preterm premature rupture of membranes in twin pregnancies, Arch Gynecol Obstet 300 (2019), 583-587. |
|  |  | [180] | G.H. Son, Y. Kim, J.J. Lee, K.Y. Lee, H. Ham, J.E. Song, S.T. Park, and Y.H. Kim, MicroRNA-548 regulates high mobility group box 1 expression in patients with preterm birth and chorioamnionitis, Sci Rep 9 (2019), 19746. |
|  |  | [181] | J. Stranik, M. Kacerovsky, O. Soucek, M. Kolackova, I. Musilova, L. Pliskova, R. Bolehovska, P. Bostik, J. Matulova, B. Jacobsson, and C. Andrys, IgGFc-binding protein in pregnancies complicated by spontaneous preterm delivery: a retrospective cohort study, Sci Rep 11 (2021), 6107. |
|  |  | [182] | J.K. Straughen, D.P. Misra, L.M. Ernst, A.K. Charles, S. VanHorn, S. Ghosh, I. Buhimschi, C. Buhimschi, G. Divine, and C.M. Salafia, Methods to decrease variability in histological scoring in placentas from a cohort of preterm infants, BMJ Open 7 (2017), e013877. |
|  |  | [183] | J.H. Sung, S.J. Choi, S.Y. Oh, C.R. Roh, and J.H. Kim, Revisiting the diagnostic criteria of clinical chorioamnionitis in preterm birth, Bjog 124 (2017), 775-783. |
|  |  | [184] | J.H. Sung, J.Y. Kuk, H.H. Cha, S.J. Choi, S.Y. Oh, C.R. Roh, and J.H. Kim, Amniopatch treatment for preterm premature rupture of membranes before 23 weeks' gestation and factors associated with its success, Taiwan J Obstet Gynecol 56 (2017), 599-605. |
|  |  | [185] | S. Suzuki, Association between clinical chorioamnionitis and histological funisitis at term, J Neonatal Perinatal Med 12 (2019), 37-40. |
|  |  | [186] | J. Takeda, X. Fang, and D.M. Olson, Pregnant human peripheral leukocyte migration during several late pregnancy clinical conditions: a cross-sectional observational study, BMC Pregnancy Childbirth 17 (2017), 16. |
|  |  | [187] | J.L.S. Tebet, G.M. Kirsztajn, T.A. Facca, S.K. Nishida, A.R. Pereira, S.R. Moreira, J.O.P. Medina, and N. Sass, Pregnancy in renal transplant patients: Renal function markers and maternal-fetal outcomes, Pregnancy Hypertens 15 (2019), 108-113. |
|  |  | [188] | P.J. Teoh, A. Ridout, P. Seed, R.M. Tribe, and A.H. Shennan, Gender and preterm birth: Is male fetal gender a clinically important risk factor for preterm birth in high-risk women?, Eur J Obstet Gynecol Reprod Biol 225 (2018), 155-159. |
|  |  | [189] | M.E. Toukam, M. Luisin, J. Chevreau, S. Lanta-Delmas, J. Gondry, and P. Tourneux, A predictive neonatal mortality score for women with premature rupture of membranes after 22-27 weeks of gestation, J Matern Fetal Neonatal Med 32 (2019), 258-264. |
|  |  | [190] | L.A. Underhill, N. Avalos, R. Tucker, Z. Zhang, G. Messerlian, and B. Lechner, Serum Decorin and Biglycan as Potential Biomarkers to Predict PPROM in Early Gestation, Reprod Sci (2019), 1933719119831790. |
|  |  | [191] | L.A. Underhill, N. Avalos, R. Tucker, Z. Zhang, G. Messerlian, and B. Lechner, Serum Decorin and Biglycan as Potential Biomarkers to Predict PPROM in Early Gestation, Reprod Sci 27 (2020), 1620-1626. |
|  |  | [192] | K. Uno, M. Mayama, M. Yoshihara, T. Takeda, S. Tano, T. Suzuki, Y. Kishigami, and H. Oguchi, Reasons for previous Cesarean deliveries impact a woman's independent decision of delivery mode and the success of trial of labor after Cesarean, BMC Pregnancy Childbirth 20 (2020). |
|  |  | [193] | D. Urushiyama, W. Suda, E. Ohnishi, R. Araki, C. Kiyoshima, M. Kurakazu, A. Sanui, F. Yotsumoto, M. Murata, K. Nabeshima, S. Yasunaga, S. Saito, M. Nomiyama, M. Hattori, S. Miyamoto, and K. Hata, Microbiome profile of the amniotic fluid as a predictive biomarker of perinatal outcome, Sci Rep 7 (2017), 12171. |
|  |  | [194] | S. Usman, H. Barton, C. Wilhelm-Benartzi, and C.C. Lees, Ultrasound is better tolerated than vaginal examination in and before labour, Australian & New Zealand Journal of Obstetrics & Gynaecology 59 (2019), 362-366. |
|  |  | [195] | S. Usman, M. Wilkinson, H. Barton, and C.C. Lees, The feasibility and accuracy of ultrasound assessment in the labor room, Journal of Maternal-Fetal & Neonatal Medicine 32 (2019), 3442-3451. |
|  |  | [196] | L. van der Krogt, A.E. Ridout, P.T. Seed, and A.H. Shennan, Placental inflammation and its relationship to cervicovaginal fetal fibronectin in preterm birth, Eur J Obstet Gynecol Reprod Biol 214 (2017), 173-177. |
|  |  | [197] | B.I. Vandermolen, N.L. Hezelgrave, E.M. Smout, D.S. Abbott, P.T. Seed, and A.H. Shennan, Quantitative fetal fibronectin and cervical length to predict preterm birth in asymptomatic women with previous cervical surgery, Am J Obstet Gynecol 215 (2016), 480.e481-480.e410. |
|  |  | [198] | S.W. Verbruggen, M.L. Oyen, A.T. Phillips, and N.C. Nowlan, Function and failure of the fetal membrane: Modelling the mechanics of the chorion and amnion, PLoS One 12 (2017), e0171588. |
|  |  | [199] | N. Volpe, E. di Pasquo, A. Ferretti, A. Dall'Asta, S. Fieni, T. Frusca, and T. Ghi, Hyperechoic amniotic membranes in patients with preterm premature rupture of membranes (p-PROM) and pregnancy outcome, J Perinat Med 49 (2021), 311-318. |
|  |  | [200] | F. Wang, Y. Wang, R. Wang, H. Qiu, and L. Chen, Predictive value of maternal serum NF-κB p65 and sTREM-1 for subclinical chorioamnionitis in premature rupture of membranes, Am J Reprod Immunol 76 (2016), 217-223. |
|  |  | [201] | E. Weiner, J. Barrett, A. Zaltz, M. Ram, A. Aviram, M. Kibel, H. Lipworth, E. Asztalos, and N. Melamed, Amniotic fluid volume at presentation with early preterm prelabor rupture of membranes and association with severe neonatal respiratory morbidity, Ultrasound in Obstetrics & Gynecology 54 (2019), 767-773. |
|  |  | [202] | E. Weiner, Y. Mizrachi, E. Grinstein, O. Feldstein, N. Rymer-Haskel, E. Juravel, L. Schreiber, J. Bar, and M. Kovo, The role of placental histopathological lesions in predicting recurrence of preeclampsia, Prenat Diagn 36 (2016), 953-960. |
|  |  | [203] | Y. Yagur, O. Weitzner, E. Ravid, and T. Biron-Shental, Can we predict preterm delivery in patients with premature rupture of membranes?, Arch Gynecol Obstet 300 (2019), 615-621. |
|  |  | [204] | Y. Yamamoto, A. Hirose, V. Jain, and L.K. Hornberger, Branch pulmonary artery Doppler parameters predict early survival-non-survival in premature rupture of membranes, J Perinatol 40 (2020), 1821-1827. |
|  |  | [205] | F. Zhan, S. Zhu, H. Liu, Q. Wang, and G. Zhao, Blood routine test is a good indicator for predicting premature rupture of membranes, J Clin Lab Anal 33 (2019), e22673. |
|  |  | [206] | L.X. Zhang, Y. Sun, H. Zhao, N. Zhu, X.D. Sun, X. Jin, A.M. Zou, Y. Mi, and J.R. Xu, A Bayesian Stepwise Discriminant Model for Predicting Risk Factors of Preterm Premature Rupture of Membranes: A Case-control Study, Chin Med J (Engl) 130 (2017), 2416-2422. |
|  |  | [207] | L. Zhao, Y. Lin, T. Jiang, L. Wang, M. Li, Y. Wang, G. Sun, and M. Xiao, Prediction of the induction to delivery time interval in vaginal dinoprostone-induced labor: a retrospective study in a Chinese tertiary maternity hospital, J Int Med Res 47 (2019), 2647-2654. |
|  |  | [208] | J. Zhu, C. Ma, L. Zhu, J. Li, F. Peng, L. Huang, and X. Luan, A role for the NLRC4 inflammasome in premature rupture of membrane, PLoS One 15 (2020), e0237847. |
|  |  | [209] | Y. Zipori, C. Ben-David, R. Lauterbach, A. Weissman, R. Beloosesky, Y. Ginsberg, Z. Weiner, and N. Khatib, Vaginal birth after cesarean in women with pre-labor rupture of membranes at term, J Matern Fetal Neonatal Med (2020), 1-6. |
| 3. a. Eligible by title and abstract | 31 | [1] | Good clinical practice advice: Prediction of preterm labor and preterm premature rupture of membranes, Int J Gynaecol Obstet 144 (2019), 340-346. |
|  |  | [2] | K. Abdel-Malek, M.A. El-Halwagi, B.E. Hammad, O. Azmy, O. Helal, M. Eid, and M. Abdel-Rasheed, Role of maternal serum ferritin in prediction of preterm labour, J Obstet Gynaecol 38 (2018), 222-225. |
|  |  | [3] | M.T. Aung, Y. Yu, K.K. Ferguson, D.E. Cantonwine, L. Zeng, T.F. McElrath, S. Pennathur, B. Mukherjee, and J.D. Meeker, Prediction and associations of preterm birth and its subtypes with eicosanoid enzymatic pathways and inflammatory markers, Sci Rep 9 (2019), 17049. |
|  |  | [4] | U. Baboolall, Y. Zha, X. Gong, D.R. Deng, F. Qiao, and H. Liu, Variations of plasma D-dimer level at various points of normal pregnancy and its trends in complicated pregnancies: A retrospective observational cohort study, Medicine (Baltimore) 98 (2019), e15903. |
|  |  | [5] | D. Başbuğ, A. Başbuğ, and C. Gülerman, Is unexplained elevated maternal serum alpha-fetoprotein still important predictor for adverse pregnancy outcome?, Ginekol Pol 88 (2017), 325-330. |
|  |  | [6] | M.A. Belfort, Predicting premature preterm rupture of the membranes after fetal surgery, Bjog 125 (2018), 1293. |
|  |  | [7] | M.T. Canda, N. Demir, and O. Sezer, Fetal Nasal Bone Length as a Novel Marker for Prediction of Adverse Perinatal Outcomes in the First-Trimester of Pregnancy, Balkan Med J 34 (2017), 127-131. |
|  |  | [8] | H.H. Cha, J.M. Kim, H.M. Kim, M.J. Kim, G.O. Chong, and W.J. Seong, Association between gestational age at delivery and lymphocyte-monocyte ratio in the routine second trimester complete blood cell count, Yeungnam Univ J Med 38 (2021), 34-38. |
|  |  | [9] | V. El-Achi, B. de Vries, C. O'Brien, F. Park, J. Tooher, and J. Hyett, First-Trimester Prediction of Preterm Prelabour Rupture of Membranes, Fetal Diagn Ther 47 (2020), 624-629. |
|  |  | [10] | V. El-Achi, F. Park, C. O'Brien, J. Tooher, and J. Hyett, Does low dose aspirin prescribed for risk of early onset preeclampsia reduce the prevalence of preterm prelabor rupture of membranes?, J Matern Fetal Neonatal Med 34 (2021), 618-623. |
|  |  | [11] | S. Gupta, H.S. Gaikwad, B. Nath, and A. Batra, Can vitamin C and interleukin 6 levels predict preterm premature rupture of membranes: evaluating possibilities in North Indian population, Obstet Gynecol Sci 63 (2020), 432-439. |
|  |  | [12] | H. Kansu-Celik, Y. Tasci, B.K. Karakaya, M. Cinar, T. Candar, and G.S. Caglar, Maternal serum advanced glycation end products level as an early marker for predicting preterm labor/PPROM: a prospective preliminary study, J Matern Fetal Neonatal Med 32 (2019), 2758-2762. |
|  |  | [13] | S.Y. Kook, K.H. Park, J.A. Jang, Y.M. Kim, H. Park, and S.J. Jeon, Vitamin D-binding protein in cervicovaginal fluid as a non-invasive predictor of intra-amniotic infection and impending preterm delivery in women with preterm labor or preterm premature rupture of membranes, PLoS One 13 (2018), e0198842. |
|  |  | [14] | K. Kuhrt, N. Hezelgrave, C. Foster, P.T. Seed, and A.H. Shennan, Development and validation of a tool incorporating quantitative fetal fibronectin to predict spontaneous preterm birth in symptomatic women, Ultrasound in Obstetrics and Gynecology 47 (2016), 210-216. |
|  |  | [15] | K. Kuhrt, E. Smout, N. Hezelgrave, P.T. Seed, J. Carter, and A.H. Shennan, Development and validation of a tool incorporating cervical length and quantitative fetal fibronectin to predict spontaneous preterm birth in asymptomatic high-risk women, Ultrasound in Obstetrics and Gynecology 47 (2016), 104-109. |
|  |  | [16] | H. Liang, Z. Xie, B. Liu, X. Song, and G. Zhao, A routine urine test has partial predictive value in premature rupture of the membranes, J Int Med Res 47 (2019), 2361-2370. |
|  |  | [17] | K. Lyubomirskaya, Y. Krut, L. Sergeyeva, S. Khmil, O. Lototska, N. Petrenko, and A. Kamyshnyi, Preterm premature rupture of membranes: prediction of risks in women of Zaporizhzhia region of Ukraine, Pol Merkur Lekarski 48 (2020), 399-405. |
|  |  | [18] | P. Mach, A. Köninger, L. Wicherek, R. Kimmig, S. Kasimir-Bauer, C. Birdir, B. Schmidt, and A. Gellhaus, Serum concentrations of soluble B7-H4 in early pregnancy are elevated in women with preterm premature rupture of fetal membranes, Am J Reprod Immunol 76 (2016), 149-154. |
|  |  | [19] | A. Ozel, E. Alici Davutoglu, A. Yurtkal, and R. Madazli, How do platelet-to-lymphocyte ratio and neutrophil-to-lymphocyte ratio change in women with preterm premature rupture of membranes, and threaten preterm labour?, J Obstet Gynaecol 40 (2020), 195-199. |
|  |  | [20] | S. Park, Y.A. You, H. Yun, S.J. Choi, H.S. Hwang, S.K. Choi, S.M. Lee, and Y.J. Kim, Cervicovaginal fluid cytokines as predictive markers of preterm birth in symptomatic women, Obstet Gynecol Sci 63 (2020), 455-463. |
|  |  | [21] | M.S. Payne, J.P. Newnham, D.A. Doherty, L.L. Furfaro, N.L. Pendal, D.E. Loh, and J.A. Keelan, A specific bacterial DNA signature in the vagina of Australian women in midpregnancy predicts high risk of spontaneous preterm birth (the Predict1000 study), Am J Obstet Gynecol 224 (2021), 206.e201-206.e223. |
|  |  | [22] | G. Raba, M. Kacerovsky, and P. Laudański, Eotaxin-2 as a potential marker of preterm premature rupture of membranes: A prospective, cohort, multicenter study, Adv Clin Exp Med 30 (2021), 197-202. |
|  |  | [23] | J. Takeda, X. Fang, and D.M. Olson, Pregnant human peripheral leukocyte migration during several late pregnancy clinical conditions: a cross-sectional observational study, BMC Pregnancy Childbirth 17 (2017), 16. |
|  |  | [24] | P.J. Teoh, A. Ridout, P. Seed, R.M. Tribe, and A.H. Shennan, Gender and preterm birth: Is male fetal gender a clinically important risk factor for preterm birth in high-risk women?, Eur J Obstet Gynecol Reprod Biol 225 (2018), 155-159. |
|  |  | [25] | L.A. Underhill, N. Avalos, R. Tucker, Z. Zhang, G. Messerlian, and B. Lechner, Serum Decorin and Biglycan as Potential Biomarkers to Predict PPROM in Early Gestation, Reprod Sci (2019), 1933719119831790. |
|  |  | [26] | L.A. Underhill, N. Avalos, R. Tucker, Z. Zhang, G. Messerlian, and B. Lechner, Serum Decorin and Biglycan as Potential Biomarkers to Predict PPROM in Early Gestation, Reprod Sci 27 (2020), 1620-1626. |
|  |  | [27] | B.I. Vandermolen, N.L. Hezelgrave, E.M. Smout, D.S. Abbott, P.T. Seed, and A.H. Shennan, Quantitative fetal fibronectin and cervical length to predict preterm birth in asymptomatic women with previous cervical surgery, Am J Obstet Gynecol 215 (2016), 480.e481-480.e410. |
|  |  | [28] | S.W. Verbruggen, M.L. Oyen, A.T. Phillips, and N.C. Nowlan, Function and failure of the fetal membrane: Modelling the mechanics of the chorion and amnion, PLoS One 12 (2017), e0171588. |
|  |  | [29] | F. Zhan, S. Zhu, H. Liu, Q. Wang, and G. Zhao, Blood routine test is a good indicator for predicting premature rupture of membranes, J Clin Lab Anal 33 (2019), e22673. |
|  |  | [30] | L.X. Zhang, Y. Sun, H. Zhao, N. Zhu, X.D. Sun, X. Jin, A.M. Zou, Y. Mi, and J.R. Xu, A Bayesian Stepwise Discriminant Model for Predicting Risk Factors of Preterm Premature Rupture of Membranes: A Case-control Study, Chin Med J (Engl) 130 (2017), 2416-2422. |
|  |  | [31] | J. Zhu, C. Ma, L. Zhu, J. Li, F. Peng, L. Huang, and X. Luan, A role for the NLRC4 inflammasome in premature rupture of membrane, PLoS One 15 (2020), e0237847. |
| 3. b. PROM as population | 82 | [1] | S. Alla, A. Ramseyer, J.R. Whittington, S. Peeples, S.T. Ounpraseuth, and E.F. Magann, Maternal features at time of preterm prelabor rupture of membranes and short-term neonatal outcomes, J Matern Fetal Neonatal Med (2020), 1-7. |
|  |  | [2] | N. Aracic, I. Stipic, I. Jakus Alujevic, P. Poljak, and M. Stipic, The value of ultrasound measurement of cervical length and parity in prediction of cesarean section risk in term premature rupture of membranes and unfavorable cervix, J Perinat Med 45 (2017), 99-104. |
|  |  | [3] | N. Asadi, A. Faraji, A. Keshavarzi, M. Akbarzadeh-Jahromi, and S. Yoosefi, Predictive value of procalcitonin, C-reactive protein, and white blood cells for chorioamnionitis among women with preterm premature rupture of membranes, Int J Gynaecol Obstet 147 (2019), 83-88. |
|  |  | [4] | A. Aviram, P. Quaglietta, C. Warshafsky, A. Zaltz, E. Weiner, N. Melamed, E. Ng, J. Barrett, and S. Ronzoni, Utility of ultrasound assessment in management of pregnancies with preterm prelabor rupture of membranes, Ultrasound Obstet Gynecol 55 (2020), 806-814. |
|  |  | [5] | E. Baser, D. Aydogan Kirmizi, D. Ulubas Isik, S. Ozdemirci, T. Onat, E. Serdar Yalvac, N. Demirel, and O. Moraloglu Tekin, The effects of latency period in PPROM cases managed expectantly, J Matern Fetal Neonatal Med 33 (2020), 2274-2283. |
|  |  | [6] | J. Caloone, M. Rabilloud, F. Boutitie, A. Traverse-Glehen, F. Allias-Montmayeur, L. Denis, C. Boisson-Gaudin, I.J. Hot, P. Guerre, M. Cortet, and C. Huissoud, Accuracy of several maternal seric markers for predicting histological chorioamnionitis after preterm premature rupture of membranes: a prospective and multicentric study, Eur J Obstet Gynecol Reprod Biol 205 (2016), 133-140. |
|  |  | [7] | O. Cetin, Z.D. Aydın, F.F. Verit, A.G. Zebitay, E. Karaman, S. Elasan, M. Turfan, and O. Yucel, Is Maternal Blood Procalcitonin Level a Reliable Predictor for Early Onset Neonatal Sepsis in Preterm Premature Rupture of Membranes?, Gynecol Obstet Invest 82 (2017), 163-169. |
|  |  | [8] | I. Chandra and L. Sun, Third trimester preterm and term premature rupture of membranes: Is there any difference in maternal characteristics and pregnancy outcomes?, J Chin Med Assoc 80 (2017), 657-661. |
|  |  | [9] | H.Y. Cho, I. Jung, J.Y. Kwon, S.J. Kim, Y.W. Park, and Y.H. Kim, The Delta Neutrophil Index as a predictive marker of histological chorioamnionitis in patients with preterm premature rupture of membranes: A retrospective study, PLoS One 12 (2017), e0173382. |
|  |  | [10] | T. Cobo, V. Aldecoa, F. Figueras, A. Herranz, S. Ferrero, M. Izquierdo, C. Murillo, R. Amoedo, C. Rueda, J. Bosch, R.J. Martínez-Portilla, E. Gratacós, and M. Palacio, Development and validation of a multivariable prediction model of spontaneous preterm delivery and microbial invasion of the amniotic cavity in women with preterm labor, Am J Obstet Gynecol 223 (2020), 421.e421-421.e414. |
|  |  | [11] | T. Cobo, J. Munrós, J. Ríos, J. Ferreri, F. Migliorelli, N. Baños, E. Gratacós, and M. Palacio, Contribution of Amniotic Fluid along Gestation to the Prediction of Perinatal Mortality in Women with Early Preterm Premature Rupture of Membranes, Fetal Diagn Ther 43 (2018), 105-112. |
|  |  | [12] | G. Daskalakis, P. Fotinopoulos, V. Pergialiotis, M. Theodora, P. Antsaklis, M. Sindos, N. Papantoniou, and D. Loutradis, Delayed interval delivery of the second twin in a woman with altered markers of inflammation, BMC Pregnancy Childbirth 18 (2018), 206. |
|  |  | [13] | D.A. De Silva, S. Lisonkova, P. von Dadelszen, A.R. Synnes, and L.A. Magee, Timing of delivery in a high-risk obstetric population: a clinical prediction model, BMC Pregnancy Childbirth 17 (2017), 202. |
|  |  | [14] | N. Dorfeuille, V. Morin, A. Tétu, S. Demers, G. Laforest, K. Gouin, B. Piedboeuf, and E. Bujold, Vaginal Fluid Inflammatory Biomarkers and the Risk of Adverse Neonatal Outcomes in Women with PPROM, Am J Perinatol 33 (2016), 1003-1007. |
|  |  | [15] | J.R. Duncan, K.M. Dorsett, M.M. Aziz, Z. Bursac, M.A. Cleves, A.J. Talati, M.H. Schenone, N.L. Meyer, and G. Mari, Estimated fetal weight and severe neonatal outcomes in preterm prelabor rupture of membranes, J Perinat Med 48 (2020), 687-693. |
|  |  | [16] | J.R. Duncan, K.M. Dorsett, G. Vilchez, M.H. Schenone, and G. Mari, Uterine artery pulsatility index for the prediction of obstetrical complications in preterm prelabor rupture of membranes, J Matern Fetal Neonatal Med (2019), 1-4. |
|  |  | [17] | J.R. Duncan, K. Leavitt, K. Duncan, and G. Vilchez, Detection of small for gestational age in preterm prelabor rupture of membranes by Hadlock versus the Fetal Medicine Foundation growth charts, Obstet Gynecol Sci (2021). |
|  |  | [18] | J.R. Duncan, P. Sawangkum, E.A. Hoover, M.M. Aziz, and G. Vilchez, Birthweight and Apgar at 5 minutes of life for the prediction of severe neonatal outcomes in preterm prelabor rupture of membranes, J Matern Fetal Neonatal Med (2020), 1-5. |
|  |  | [19] | J.R. Duncan, C. Schenone, K.M. Dorset, P.J. Goedecke, A.M. Tobiasz, N.L. Meyer, and M.H. Schenone, Estimated fetal weight accuracy in pregnancies with preterm prelabor rupture of membranes by the Hadlock method, J Matern Fetal Neonatal Med (2020), 1-5. |
|  |  | [20] | J.R. Duncan, A.M. Tobiasz, K.M. Dorsett, M.M. Aziz, R.E. Thompson, Z. Bursac, A.J. Talati, G. Mari, and M.H. Schenone, Fetal pulmonary artery acceleration/ejection time prognostic accuracy for respiratory complications in preterm prelabor rupture of membranes, J Matern Fetal Neonatal Med 33 (2020), 2054-2058. |
|  |  | [21] | B. Dundar, B. Dincgez Cakmak, G. Ozgen, F.N. Tasgoz, T. Guclu, and G. Ocakoglu, Platelet indices in preterm premature rupture of membranes and their relation with adverse neonatal outcomes, J Obstet Gynaecol Res 44 (2018), 67-73. |
|  |  | [22] | G.U. Eleje, C.O. Ukah, I.V. Onyiaorah, E.C. Ezugwu, E.O. Ugwu, S.R. Ohayi, L.I. Eleje, R.O. Egeonu, I.U. Ezebialu, C.C. Obiora, J.T. Enebe, L.O. Ajah, C.G. Okafor, C.C. Okoro, A.O. Asogwa, D.K. Ogbuokiri, J.I. Ikechebelu, and A.C. Eke, Diagnostic value of Chorioquick for detecting chorioamnionitis in women with premature rupture of membranes, Int J Gynaecol Obstet 149 (2020), 98-105. |
|  |  | [23] | S. Esin, M. Hayran, Y.A. Tohma, M. Guden, I. Alay, D. Esinler, S. Yalvac, and O. Kandemir, Estimation of fetal weight by ultrasonography after preterm premature rupture of membranes: comparison of different formulas, J Perinat Med 45 (2017), 253-266. |
|  |  | [24] | F.B. Flosi, F.C. da Silva, G.R. de Jesús, L.G.C. Velarde, and R.A.M. de Sá, Assessment of Fetal Lung Maturity Using Quantitative Ultrasound Analysis in Patients with Prelabor Rupture of Membranes, Fetal Diagn Ther 47 (2020), 636-641. |
|  |  | [25] | R.B. Helmig and J.B. Gertsen, Diagnostic accuracy of polymerase chain reaction for intrapartum detection of group B streptococcus colonization, Acta Obstet Gynecol Scand 96 (2017), 1070-1074. |
|  |  | [26] | L. Hiersch, E. Krispin, A. Aviram, M. Mor-Shacham, R. Gabbay-Benziv, Y. Yogev, and E. Ashwal, Predictors for prolonged interval from premature rupture of membranes to spontaneous onset of labor at term, J Matern Fetal Neonatal Med 30 (2017), 1465-1470. |
|  |  | [27] | T. Horinouchi, T. Yoshizato, Y. Kozuma, T. Shinagawa, M. Muto, T. Yamasaki, D. Hori, and K. Ushijima, Prediction of histological chorioamnionitis and neonatal and infantile outcomes using procalcitonin in the umbilical cord blood and amniotic fluid at birth, J Obstet Gynaecol Res 44 (2018), 630-636. |
|  |  | [28] | P. Janku, M. Kacerovsky, B. Zednikova, C. Andrys, M. Kolackova, M. Drahosova, L. Pliskova, H. Zemlickova, R. Gerychova, O. Simetka, P. Matlak, B. Jacobsson, and I. Musilova, Pentraxin 3 in Noninvasively Obtained Cervical Fluid Samples from Pregnancies Complicated by Preterm Prelabor Rupture of Membranes, Fetal Diagn Ther 46 (2019), 402-410. |
|  |  | [29] | E.Y. Jung, K.H. Park, B.R. Han, S.H. Cho, and A. Ryu, Measurement of Interleukin 8 in Cervicovaginal Fluid in Women With Preterm Premature Rupture of Membranes: A Comparison of Amniotic Fluid Samples, Reproductive Sciences 24 (2017), 142-147. |
|  |  | [30] | M. Kacerovsky, I. Musilova, T. Bestvina, M. Stepan, T. Cobo, and B. Jacobsson, Preterm Prelabor Rupture of Membranes between 34 and 37 Weeks: A Point-of-Care Test of Vaginal Fluid Interleukin-6 Concentrations for a Noninvasive Detection of Intra-Amniotic Inflammation, Fetal Diagn Ther 43 (2018), 175-183. |
|  |  | [31] | V. Kathir, D. Maurya, and A. Keepanasseril, Transvaginal sonographic assessment of cervix in prediction of admission to delivery interval in preterm premature rupture of membranes, J Matern Fetal Neonatal Med 31 (2018), 2717-2720. |
|  |  | [32] | G. Kayem, F. Batteux, N. Girard, T. Schmitz, M. Willaime, F. Maillard, P.H. Jarreau, and F. Goffinet, Predictive value of vaginal IL-6 and TNFα bedside tests repeated until delivery for the prediction of maternal-fetal infection in cases of premature rupture of membranes, Eur J Obstet Gynecol Reprod Biol 211 (2017), 8-14. |
|  |  | [33] | H.J. Kim, K.H. Park, Y.M. Kim, E. Joo, K. Ahn, and S. Shin, A protein microarray analysis of amniotic fluid proteins for the prediction of spontaneous preterm delivery in women with preterm premature rupture of membranes at 23 to 30 weeks of gestation, PLoS One 15 (2020), e0244720. |
|  |  | [34] | V. Kiver, V. Boos, A. Thomas, W. Henrich, and A. Weichert, Perinatal outcomes after previable preterm premature rupture of membranes before 24 weeks of gestation, J Perinat Med 46 (2018), 555-565. |
|  |  | [35] | M. Kunze, M. Klar, C.A. Morfeld, B. Thorns, R.L. Schild, F. Markfeld-Erol, R. Rasenack, H. Proempeler, R. Hentschel, and W.R. Schaefer, Cytokines in noninvasively obtained amniotic fluid as predictors of fetal inflammatory response syndrome, Am J Obstet Gynecol 215 (2016), 96.e91-98. |
|  |  | [36] | M. Kurek Eken, A. Tüten, E. Özkaya, G. Karatekin, and A. Karateke, Major determinants of survival and length of stay in the neonatal intensive care unit of newborns from women with premature preterm rupture of membranes, J Matern Fetal Neonatal Med 30 (2017), 1972-1975. |
|  |  | [37] | S.M. Lee, K.H. Park, E.Y. Jung, J.A. Jang, and H.N. Yoo, Frequency and clinical significance of short cervix in patients with preterm premature rupture of membranes, PLoS One 12 (2017), e0174657. |
|  |  | [38] | S.M. Lee, K.H. Park, E.Y. Jung, S.Y. Kook, H. Park, and S.J. Jeon, Inflammatory proteins in maternal plasma, cervicovaginal and amniotic fluids as predictors of intra-amniotic infection in preterm premature rupture of membranes, PLoS One 13 (2018), e0200311. |
|  |  | [39] | Y.J. Lee, S.C. Kim, J.K. Joo, D.H. Lee, K.H. Kim, and K.S. Lee, Amniotic fluid index, single deepest pocket and transvaginal cervical length: Parameter of predictive delivery latency in preterm premature rupture of membranes, Taiwan J Obstet Gynecol 57 (2018), 374-378. |
|  |  | [40] | F. Lemoine, R.C. Moore, A. Chapple, F.A. Moore, and E. Sutton, Neonatal Survivability following Previable PPROM after Hospital Readmission for Intervention, AJP Reports 10 (2020), E395-E402. |
|  |  | [41] | J. Li, B. Yu, W. Wang, D. Luo, Q.L. Dai, and X.Q. Gan, Does intact umbilical cord milking increase infection rates in preterm infants with premature prolonged rupture of membranes?, Journal of Maternal-Fetal & Neonatal Medicine 33 (2020), 184-190. |
|  |  | [42] | Y. Ma, Y. Xu, L. Jiang, and X. Shao, Application of a Prediction Model Based on the Laboratory Index Score in Prelabor Rupture of Membranes with Histologic Chorioamnionitis During Late Pregnancy, Med Sci Monit 26 (2020), e924756. |
|  |  | [43] | Y. Maki, S. Furukawa, T. Nakayama, M. Oohashi, N. Shiiba, K. Furuta, S. Tokunaga, and H. Sameshima, Clinical chorioamnionitis criteria are not sufficient for predicting intra-amniotic infection, J Matern Fetal Neonatal Med (2020), 1-6. |
|  |  | [44] | R.J. Martinez-Portilla, A. Hawkins-Villarreal, P. Alvarez-Ponce, Z.L. Chinolla-Arellano, A.L. Moreno-Espinosa, A.L. Sandoval-Mejia, and N. Moreno-Uribe, Maternal Serum Interleukin-6: A Non-Invasive Predictor of Histological Chorioamnionitis in Women with Preterm-Prelabor Rupture of Membranes, Fetal Diagn Ther 45 (2019), 168-175. |
|  |  | [45] | M. Merello, L. Lotte, S. Gonfrier, S. Eleni Dit Trolli, F. Casagrande, R. Ruimy, and A. Bongain, Enterobacteria vaginal colonization among patients with preterm premature rupture of membranes from 24 to 34 weeks of gestation and neonatal infection risk, J Gynecol Obstet Hum Reprod 48 (2019), 187-191. |
|  |  | [46] | M. Mikołajczyk, P. Wirstlein, M. Adamczyk, J. Skrzypczak, and E. Wender-Ożegowska, Value of cervicovaginal fluid cytokines in prediction of fetal inflammatory response syndrome in pregnancies complicated with preterm premature rupture of membranes (pPROM), J Perinat Med 48 (2020), 249-255. |
|  |  | [47] | S. Mishra, S. Jaiswar, S. Saad, S. Tripathi, N. Singh, S. Deo, M. Agarwal, and N. Mishra, Platelet indices as a predictive marker in neonatal sepsis and respiratory distress in preterm prelabor rupture of membranes, Int J Hematol 113 (2021), 199-206. |
|  |  | [48] | A.S. Mousavi, N. Hashemi, M. Kashanian, N. Sheikhansari, A. Bordbar, and S. Parashi, Comparison between maternal and neonatal outcome of PPROM in the cases of amniotic fluid index (AFI) of more and less than 5 cm, J Obstet Gynaecol 38 (2018), 611-615. |
|  |  | [49] | I. Musilova, C. Andrys, M. Drahosova, O. Soucek, R. Kutova, L. Pliskova, R. Spacek, P. Laudanski, B. Jacobsson, and M. Kacerovsky, Amniotic fluid calreticulin in pregnancies complicated by the preterm prelabor rupture of membranes, Journal of Maternal-Fetal and Neonatal Medicine 29 (2016), 3921-3929. |
|  |  | [50] | I. Musilova, C. Andrys, M. Drahosova, O. Soucek, L. Pliskova, B. Jacobsson, and M. Kacerovsky, Cervical fluid interleukin 6 and intra-amniotic complications of preterm prelabor rupture of membranes, Journal of Maternal-Fetal & Neonatal Medicine 31 (2018), 827-836. |
|  |  | [51] | I. Musilova, C. Andrys, M. Drahosova, O. Soucek, L. Pliskova, M. Stepan, T. Bestvina, J. Maly, B. Jacobsson, and M. Kacerovsky, Amniotic fluid cathepsin-G in pregnancies complicated by the preterm prelabor rupture of membranes, Journal of Maternal-Fetal and Neonatal Medicine 30 (2017), 2097-2104. |
|  |  | [52] | I. Musilova, C. Andrys, M. Holeckova, V. Kolarova, L. Pliskova, M. Drahosova, R. Bolehovska, R. Pilka, K. Huml, T. Cobo, B. Jacobsson, and M. Kacerovsky, Interleukin-6 measured using the automated electrochemiluminescence immunoassay method for the identification of intra-amniotic inflammation in preterm prelabor rupture of membranes, J Matern Fetal Neonatal Med 33 (2020), 1919-1926. |
|  |  | [53] | I. Musilova, C. Andrys, H. Hornychova, L. Pliskova, M. Drahosova, B. Zednikova, R. Bolehovska, T. Faist, B. Jacobsson, and M. Kacerovsky, Gastric fluid used to assess changes during the latency period in preterm prelabor rupture of membranes, Pediatr Res 84 (2018), 240-247. |
|  |  | [54] | I. Musilova, T. Bestvina, M. Hudeckova, I. Michalec, T. Cobo, B. Jacobsson, and M. Kacerovsky, Vaginal fluid interleukin-6 concentrations as a point-of-care test is of value in women with preterm prelabor rupture of membranes, Am J Obstet Gynecol 215 (2016), 619.e611-619.e612. |
|  |  | [55] | I. Musilova, T. Bestvina, M. Hudeckova, I. Michalec, T. Cobo, B. Jacobsson, and M. Kacerovsky, Vaginal fluid IL-6 concentrations as a point-of-care test is of value in women with preterm PROM, Am J Obstet Gynecol (2016). |
|  |  | [56] | I. Musilova, M. Kacerovsky, M. Stepan, T. Bestvina, L. Pliskova, B. Zednikova, and B. Jacobsson, Maternal serum C-reactive protein concentration and intra-amniotic inflammation in women with preterm prelabor rupture of membranes, PLoS One 12 (2017), e0182731. |
|  |  | [57] | I. Musilova, L. Pliskova, R. Gerychova, P. Janku, O. Simetka, P. Matlak, B. Jacobsson, and M. Kacerovsky, Maternal white blood cell count cannot identify the presence of microbial invasion of the amniotic cavity or intra-amniotic inflammation in women with preterm prelabor rupture of membranes, PLoS One 12 (2017). |
|  |  | [58] | T. Myntti, L. Rahkonen, A. Pätäri-Sampo, M. Tikkanen, T. Sorsa, J. Juhila, O. Helve, S. Andersson, J. Paavonen, and V. Stefanovic, Comparison of amniotic fluid matrix metalloproteinase-8 and cathelicidin in the diagnosis of intra-amniotic infection, J Perinatol 36 (2016), 1049-1054. |
|  |  | [59] | M. Nakahara, S. Goto, E. Kato, S. Nojiri, A. Itakura, and S. Takeda, Maternal risk score for the prediction of fetal inflammatory response syndrome after preterm premature rupture of membranes, J Obstet Gynaecol Res 46 (2020), 2019-2026. |
|  |  | [60] | D. Odd, A. Heep, K. Luyt, and T. Draycott, Hypoxic-ischemic brain injury: Planned delivery before intrapartum events, J Neonatal Perinatal Med 10 (2017), 347-353. |
|  |  | [61] | K.J. Oh, J. Lee, R. Romero, H.S. Park, J.S. Hong, and B.H. Yoon, A new rapid bedside test to diagnose and monitor intraamniotic inflammation in preterm PROM using transcervically collected fluid, Am J Obstet Gynecol 223 (2020), 423.e421-423.e415. |
|  |  | [62] | T. Paramel Jayaprakash, E.C. Wagner, J. van Schalkwyk, A.Y. Albert, J.E. Hill, and D.M. Money, High Diversity and Variability in the Vaginal Microbiome in Women following Preterm Premature Rupture of Membranes (PPROM): A Prospective Cohort Study, PLoS One 11 (2016), e0166794. |
|  |  | [63] | J.W. Park, K.H. Park, J.E. Lee, Y.M. Kim, S.J. Lee, and D.H. Cheon, Antibody Microarray Analysis of Plasma Proteins for the Prediction of Histologic Chorioamnionitis in Women With Preterm Premature Rupture of Membranes, Reprod Sci 26 (2019), 1476-1484. |
|  |  | [64] | C. Petit, P. Deruelle, H. Behal, T. Rakza, S. Balagny, D. Subtil, E. Clouqueur, and C. Garabedian, Preterm premature rupture of membranes: Which criteria contraindicate home care management?, Acta Obstet Gynecol Scand 97 (2018), 1499-1507. |
|  |  | [65] | A.M. Ramseyer, J.R. Whittington, E.F. Magann, J.A. Pates, and S.T. Ounpraseuth, Can maternal characteristics on admission for preterm prelabor rupture of membranes predict pregnancy latency?, Am J Obstet Gynecol MFM 2 (2020), 100194. |
|  |  | [66] | R. Revello, M.J. Alcaide, D. Abehsera, M. Martin-Camean, E.F.G.M. Sousa, B. Alonso-Luque, and J.L. Bartha, Prediction of chorioamnionitis in cases of intraamniotic infection by ureaplasma urealyticum in women with very preterm premature rupture of membranes or preterm labour, J Matern Fetal Neonatal Med 31 (2018), 1839-1844. |
|  |  | [67] | M. Rewatkar, S. Jain, M. Jain, and K. Mohod, C-reactive protein and white blood cell count as predictors of maternal and neonatal infections in prelabour rupture of membranes between 34 and 41 weeks of gestation, J Obstet Gynaecol 38 (2018), 622-628. |
|  |  | [68] | S. Ronzoni, R. D'Souza, O. Shynlova, S. Lye, and K.E. Murphy, Maternal blood endotoxin activity in pregnancies complicated by preterm premature rupture of membranes, J Matern Fetal Neonatal Med 32 (2019), 3473-3479. |
|  |  | [69] | S. Ronzoni, V. Steckle, R. D'Souza, K.E. Murphy, S. Lye, and O. Shynlova, Cytokine Changes in Maternal Peripheral Blood Correlate With Time-to-Delivery in Pregnancies Complicated by Premature Prelabor Rupture of the Membranes, Reprod Sci 26 (2019), 1266-1276. |
|  |  | [70] | M. Rouzaire, M. Corvaisier, V. Roumeau, A. Mulliez, F. Sendy, A. Delabaere, and D. Gallot, Predictors of Short Latency Period Exceeding 48 h after Preterm Premature Rupture of Membranes, J Clin Med 10 (2021). |
|  |  | [71] | H.K. Ryu, J.H. Moon, H.J. Heo, J.W. Kim, and Y.H. Kim, Maternal c-reactive protein and oxidative stress markers as predictors of delivery latency in patients experiencing preterm premature rupture of membranes, Int J Gynaecol Obstet 136 (2017), 145-150. |
|  |  | [72] | W.A. Sayed Ahmed, M.R. Ahmed, M.L. Mohamed, M.A. Hamdy, Z. Kamel, and K.M. Elnahas, Maternal serum interleukin-6 in the management of patients with preterm premature rupture of membranes, J Matern Fetal Neonatal Med 29 (2016), 3162-3166. |
|  |  | [73] | G. Seliger, M. Bergner, R. Haase, H. Stepan, E. Schleußner, J. Zöllkau, S. Seeger, F.B. Kraus, G.G.R. Hiller, A. Wienke, and M. Tchirikov, Daily monitoring of vaginal interleukin 6 as a predictor of intraamniotic inflammation after preterm premature rupture of membranes - a new method of sampling studied in a prospective multicenter trial, J Perinat Med (2021). |
|  |  | [74] | W.H. Sim, H. Ng, and P. Sheehan, Maternal and neonatal outcomes following expectant management of preterm prelabor rupture of membranes before viability, J Matern Fetal Neonatal Med 33 (2020), 533-541. |
|  |  | [75] | J.H. Sung, J.Y. Kuk, H.H. Cha, S.J. Choi, S.Y. Oh, C.R. Roh, and J.H. Kim, Amniopatch treatment for preterm premature rupture of membranes before 23 weeks' gestation and factors associated with its success, Taiwan J Obstet Gynecol 56 (2017), 599-605. |
|  |  | [76] | M.E. Toukam, M. Luisin, J. Chevreau, S. Lanta-Delmas, J. Gondry, and P. Tourneux, A predictive neonatal mortality score for women with premature rupture of membranes after 22-27 weeks of gestation, J Matern Fetal Neonatal Med 32 (2019), 258-264. |
|  |  | [77] | N. Volpe, E. di Pasquo, A. Ferretti, A. Dall'Asta, S. Fieni, T. Frusca, and T. Ghi, Hyperechoic amniotic membranes in patients with preterm premature rupture of membranes (p-PROM) and pregnancy outcome, J Perinat Med 49 (2021), 311-318. |
|  |  | [78] | F. Wang, Y. Wang, R. Wang, H. Qiu, and L. Chen, Predictive value of maternal serum NF-κB p65 and sTREM-1 for subclinical chorioamnionitis in premature rupture of membranes, Am J Reprod Immunol 76 (2016), 217-223. |
|  |  | [79] | E. Weiner, J. Barrett, A. Zaltz, M. Ram, A. Aviram, M. Kibel, H. Lipworth, E. Asztalos, and N. Melamed, Amniotic fluid volume at presentation with early preterm prelabor rupture of membranes and association with severe neonatal respiratory morbidity, Ultrasound in Obstetrics & Gynecology 54 (2019), 767-773. |
|  |  | [80] | Y. Yagur, O. Weitzner, E. Ravid, and T. Biron-Shental, Can we predict preterm delivery in patients with premature rupture of membranes?, Arch Gynecol Obstet 300 (2019), 615-621. |
|  |  | [81] | Y. Yamamoto, A. Hirose, V. Jain, and L.K. Hornberger, Branch pulmonary artery Doppler parameters predict early survival-non-survival in premature rupture of membranes, J Perinatol 40 (2020), 1821-1827. |
|  |  | [82] | Y. Zipori, C. Ben-David, R. Lauterbach, A. Weissman, R. Beloosesky, Y. Ginsberg, Z. Weiner, and N. Khatib, Vaginal birth after cesarean in women with pre-labor rupture of membranes at term, J Matern Fetal Neonatal Med (2020), 1-6. |
| 3. c. Association study | 3 | [1] | I.M. Aris, S. Logan, C. Lim, M. Choolani, A. Biswas, and S. Bhattacharya, Preterm prelabour rupture of membranes: a retrospective cohort study of association with adverse outcome in subsequent pregnancy, Bjog-an International Journal of Obstetrics and Gynaecology 124 (2017), 1698-1707. |
|  |  | [2] | B. Kaya, U. Turhan, S. Sezer, S. Kaya, İ. Dağ, and A. Tayyar, Maternal serum galectin-1 and galectin-3 levels in pregnancies complicated with preterm prelabor rupture of membranes, J Matern Fetal Neonatal Med 33 (2020), 861-868. |
|  |  | [3] | B.P. Modi, H.I. Parikh, M.E. Teves, R. Kulkarni, J. Liyu, R. Romero, T.P. York, and J.F. Strauss, 3rd, Discovery of rare ancestry-specific variants in the fetal genome that confer risk of preterm premature rupture of membranes (PPROM) and preterm birth, BMC Med Genet 19 (2018), 181. |
| 3. d. Diagnostic prediction study | 11 | [1] | Z. Bouzari, R. Shahhosseini, M. Mohammadnetaj, S. Barat, S. Yazdani, and K. Hajian-Tilaki, Vaginal discharge concentrations of β-human chorionic gonadotropin, creatinine, and urea for the diagnosis of premature rupture of membranes, Int J Gynaecol Obstet 141 (2018), 97-101. |
|  |  | [2] | C.B. Dartibale, N.S. Uchimura, L. Nery, A.P. Schumeish, L.Y.T. Uchimura, R.G. Santana, and T.T. Uchimura, Qualitative Determination of Human Chorionic Gonadotropin in Vaginal Washings for the Early Diagnosis of Premature Rupture of Fetal Membranes, Rev Bras Ginecol Obstet 39 (2017), 317-321. |
|  |  | [3] | E. Elçi, G. Güneş Elçi, and S. Sayan, Comparison of the accuracy and reliability of the AmniSure, AMNIOQUICK, and AL-SENSE tests for early diagnosis of premature rupture of membranes, Int J Gynaecol Obstet 149 (2020), 93-97. |
|  |  | [4] | G.U. Eleje, E.C. Ezugwu, A.C. Eke, J.I. Ikechebelu, C.O. Ezeama, I.U. Ezebialu, N.O. Ojiegbe, C.C. Obiora, C.I. Okafor, G.O. Udigwe, B.O. Nwosu, and F.O. Ezugwu, Accuracy and response time of dual biomarker model of insulin-like growth factor binding protein-1/ alpha fetoprotein (Amnioquick duo+) in comparison to placental alpha-microglobulin-1 test in diagnosis of premature rupture of membranes, J Obstet Gynaecol Res 43 (2017), 825-833. |
|  |  | [5] | G.U. Eleje, E.C. Ezugwu, A.C. Eke, J.I. Ikechebelu, C.C. Obiora, N.O. Ojiegbe, I.U. Ezebialu, C.O. Ezeama, B.O. Nwosu, G.O. Udigwe, C.I. Okafor, and F.O. Ezugwu, Comparison of the duo of insulin-like growth factor binding protein-1/alpha fetoprotein (Amnioquick duo+®) and traditional clinical assessment for diagnosing premature rupture of fetal membranes, J Perinat Med 45 (2017), 105-112. |
|  |  | [6] | G.U. Eleje, E.C. Ezugwu, I.U. Ezebialu, N.O. Ojiegbe, R.O. Egeonu, C.C. Obiora, C.G. Okafor, J.I. Ikechebelu, and A.C. Eke, Performance indices of AmnioQuick Duo+ versus placental α-microglobulin-1 tests for women with prolonged premature rupture of membranes, Int J Gynaecol Obstet 144 (2019), 180-186. |
|  |  | [7] | C. Gezer, A. Ekin, C. Golbasi, C. Kocahakimoglu, U. Bozkurt, A. Dogan, U. Solmaz, H. Golbasi, and C.E. Taner, Use of urea and creatinine levels in vaginal fluid for the diagnosis of preterm premature rupture of membranes and delivery interval after membrane rupture, J Matern Fetal Neonatal Med 30 (2017), 772-778. |
|  |  | [8] | I. Igbinosa, F.A. Moore, 3rd, C. Johnson, and J.E. Block, Comparison of rapid immunoassays for rupture of fetal membranes, BMC Pregnancy Childbirth 17 (2017), 128. |
|  |  | [9] | M. Julien, L. N'Guema, A. Bouzerara, B. Toro, E. Lecarpentier, and J. Guibourdenche, Premature rupture of the membranes: analytical evaluation of diagnostic tests, Ann Biol Clin (Paris) 76 (2018), 300-306. |
|  |  | [10] | I.H. Polat, S. Marin, J. Ríos, M. Larroya, A.B. Sánchez-García, C. Murillo, C. Rueda, M. Cascante, E. Gratacós, and T. Cobo, Exploratory and confirmatory analysis to investigate the presence of vaginal metabolome expression of microbial invasion of the amniotic cavity in women with preterm labor using high-performance liquid chromatography, Am J Obstet Gynecol 224 (2021), 90.e91-90.e99. |
|  |  | [11] | L.C. Rogers, L. Scott, and J.E. Block, Accurate Point-of-Care Detection of Ruptured Fetal Membranes: Improved Diagnostic Performance Characteristics with a Monoclonal/Polyclonal Immunoassay, Clin Med Insights Reprod Health 10 (2016), 15-18. |
| 3. e. Special population | 12 | [1] | R.M. Alvarez, I. Biliatis, A. Rockall, E. Papadakou, S.A. Sohaib, N.M. deSouza, J. Butler, M. Nobbenhuis, D. Barton, J.H. Shepherd, and T. Ind, MRI measurement of residual cervical length after radical trachelectomy for cervical cancer and the risk of adverse pregnancy outcomes: a blinded imaging analysis, Bjog 125 (2018), 1726-1733. |
|  |  | [2] | R.D. Ducci, P.J. Lorenzoni, C.S. Kay, L.C. Werneck, and R.H. Scola, Clinical follow-up of pregnancy in myasthenia gravis patients, Neuromuscul Disord 27 (2017), 352-357. |
|  |  | [3] | N. Garry, O. Keenan, S.W. Lindow, and T. Darcy, Pregnancy outcomes following elective abdominal cerclage following cervical excision surgery for neoplastic disease, European Journal of Obstetrics & Gynecology and Reproductive Biology 256 (2021), 225-229. |
|  |  | [4] | K.M. Hughes, S.C. Kane, T.P. Haines, and P.M. Sheehan, Cervical length surveillance for predicting spontaneous preterm birth in women with uterine anomalies: A cohort study, Acta Obstet Gynecol Scand 99 (2020), 1519-1526. |
|  |  | [5] | C.L. Kothari, C. Romph, T. Bautista, and D. Lenz, Perinatal Periods of Risk Analysis: Disentangling Race and Socioeconomic Status to Inform a Black Infant Mortality Community Action Initiative, Matern Child Health J 21 (2017), 49-58. |
|  |  | [6] | W.F. Li, A.S. Chao, S.D. Chang, P.J. Cheng, L.Y. Yang, and Y.L. Chang, Effects and outcomes of septostomy in twin-to-twin transfusion syndrome after fetoscopic laser therapy, BMC Pregnancy Childbirth 19 (2019), 397. |
|  |  | [7] | L. Liu, H. Wang, Y. Zhang, J. Niu, Z. Li, and R. Tang, Effect of pregravid obesity on perinatal outcomes in singleton pregnancies following in vitro fertilization and the weight-loss goals to reduce the risks of poor pregnancy outcomes: A retrospective cohort study, PLoS One 15 (2020), e0227766. |
|  |  | [8] | Y. Mei and Y. Lin, Clinical significance of primary symptoms in women with placental abruption, J Matern Fetal Neonatal Med 31 (2018), 2446-2449. |
|  |  | [9] | A.F. Moron, A. Athanasiou, M. Barbosa, H.J. Milani, S.G. Sarmento, S. Cavalheiro, and S.S. Witkin, Amniotic fluid lactic acid and matrix metalloproteinase-8 levels at the time of fetal surgery for a spine defect: association with subsequent preterm prelabour rupture of membranes, Bjog 125 (2018), 1288-1292. |
|  |  | [10] | S. Shinar, S. Agrawal, D. El-Chaar, N. Abbasi, R. Beecroft, J. Kachura, J. Keunen, G. Seaward, T. Van Mieghem, R. Windrim, and G. Ryan, Selective fetal reduction in complicated monochorionic twin pregnancies: A comparison of techniques, Prenat Diagn 41 (2021), 52-60. |
|  |  | [11] | G. Sisti, S. Paccosi, A. Parenti, V. Seravalli, M. Di Tommaso, and S.S. Witkin, Insulin-like growth factor binding protein-1 predicts preterm premature rupture of membranes in twin pregnancies, Arch Gynecol Obstet 300 (2019), 583-587. |
|  |  | [12] | J.L.S. Tebet, G.M. Kirsztajn, T.A. Facca, S.K. Nishida, A.R. Pereira, S.R. Moreira, J.O.P. Medina, and N. Sass, Pregnancy in renal transplant patients: Renal function markers and maternal-fetal outcomes, Pregnancy Hypertens 15 (2019), 108-113. |
| 3. f. Not PROM | 68 | [1] | W.E.t. Ackerman, I.A. Buhimschi, D. Brubaker, S. Maxwell, K.M. Rood, M.R. Chance, H. Jing, S. Mesiano, and C.S. Buhimschi, Integrated microRNA and mRNA network analysis of the human myometrial transcriptome in the transition from quiescence to labor, Biol Reprod 98 (2018), 834-845. |
|  |  | [2] | F. Bahlmann and A. Al Naimi, Using the angiogenic factors sFlt-1 and PlGF with Doppler ultrasound of the uterine artery for confirming preeclampsia, Arch Gynecol Obstet 294 (2016), 1133-1139. |
|  |  | [3] | A.R. Ballard, L.H. Mallett, J.E. Pruszynski, and J.B. Cantey, Chorioamnionitis and subsequent bronchopulmonary dysplasia in very-low-birth weight infants: a 25-year cohort, J Perinatol 36 (2016), 1045-1048. |
|  |  | [4] | S.V. Barinov, G.C. Di Renzo, A.A. Belinina, O.V. Koliado, and O.V. Remneva, Clinical and biochemical markers of spontaneous preterm birth in singleton and multiple pregnancies, J Matern Fetal Neonatal Med (2021), 1-6. |
|  |  | [5] | J.B. Beavers, S. Bai, J. Perry, J. Simpson, and S. Peeples, Implementation and Evaluation of the Early-Onset Sepsis Risk Calculator in a High-Risk University Nursery, Clin Pediatr (Phila) 57 (2018), 1080-1085. |
|  |  | [6] | A.P. Burgess, J. Katz, J. Pessolano, J. Ponterio, M. Moretti, and N.A. Lakhi, Determination of antepartum and intrapartum risk factors associated with neonatal intensive care unit admission, J Perinat Med 44 (2016), 589-596. |
|  |  | [7] | W. Cao, C. Luo, M. Lei, M. Shen, W. Ding, M. Wang, M. Song, J. Ge, and Q. Zhang, Development and Validation of a Dynamic Nomogram to Predict the Risk of Neonatal White Matter Damage, Front Hum Neurosci 14 (2020), 584236. |
|  |  | [8] | D. Carola, M. Vasconcellos, A. Sloane, D. McElwee, C. Edwards, J. Greenspan, and Z.H. Aghai, Utility of Early-Onset Sepsis Risk Calculator for Neonates Born to Mothers with Chorioamnionitis, J Pediatr 195 (2018), 48-52.e41. |
|  |  | [9] | P. Chaemsaithong, R. Romero, N. Docheva, N. Chaiyasit, G. Bhatti, P. Pacora, S.S. Hassan, L. Yeo, and O. Erez, Comparison of rapid MMP-8 and interleukin-6 point-of-care tests to identify intra-amniotic inflammation/infection and impending preterm delivery in patients with preterm labor and intact membranes(), J Matern Fetal Neonatal Med 31 (2018), 228-244. |
|  |  | [10] | N. Chaiyasit, R. Romero, P. Chaemsaithong, N. Docheva, G. Bhatti, J.P. Kusanovic, Z. Dong, L. Yeo, P. Pacora, S.S. Hassan, and O. Erez, Clinical chorioamnionitis at term VIII: a rapid MMP-8 test for the identification of intra-amniotic inflammation, J Perinat Med 45 (2017), 539-550. |
|  |  | [11] | S. Chaudhury, J. Saqibuddin, R. Birkett, K. Falcon-Girard, M. Kraus, L.M. Ernst, W. Grobman, and K.K. Mestan, Variations in Umbilical Cord Hematopoietic and Mesenchymal Stem Cells With Bronchopulmonary Dysplasia, Front Pediatr 7 (2019), 475. |
|  |  | [12] | A.L. Chen, I.T. Goldfarb, A.O. Scourtas, and D.J. Roberts, The histologic evolution of revealed, acute abruptions, Hum Pathol 67 (2017), 187-197. |
|  |  | [13] | K.W. Cheung, L.N. Tan, M.T.Y. Seto, S. Moholkar, G. Masson, and M.D. Kilby, Prenatal Diagnosis, Management, and Outcome of Fetal Subdural Haematoma: A Case Report and Systematic Review, Fetal Diagn Ther 46 (2019), 285-295. |
|  |  | [14] | Y. Dabi, S. Nedellec, C. Bonneau, B. Trouchard, R. Rouzier, and A. Benachi, Clinical validation of a model predicting the risk of preterm delivery, PLoS One 12 (2017), e0171801. |
|  |  | [15] | C. Davies, G. Segre, A. Estradé, J. Radua, A. De Micheli, U. Provenzani, D. Oliver, G. Salazar de Pablo, V. Ramella-Cravaro, M. Besozzi, P. Dazzan, M. Miele, G. Caputo, C. Spallarossa, G. Crossland, A. Ilyas, G. Spada, P. Politi, R.M. Murray, P. McGuire, and P. Fusar-Poli, Prenatal and perinatal risk and protective factors for psychosis: a systematic review and meta-analysis, Lancet Psychiatry 7 (2020), 399-410. |
|  |  | [16] | H. Erfani, S. Haeri, S.A. Shainker, A.F. Saad, R. Ruano, T.N. Dunn, A. Rezaei, S. Aalipour, A.A. Nassr, A.A. Shamshirsaz, M. Vaughn, W. Lindsley, M.H. Spiel, S.A. Shazly, E.R. Ibirogba, S.L. Clark, G.R. Saade, M.A. Belfort, and A.A. Shamshirsaz, Vasa previa: a multicenter retrospective cohort study, Am J Obstet Gynecol 221 (2019), 644.e641-644.e645. |
|  |  | [17] | A. Frick, V. Kostiv, D. Vojtassakova, R. Akolekar, and K.H. Nicolaides, Comparison of different methods of measuring angle of progression in prediction of labor outcome, Ultrasound in Obstetrics & Gynecology 55 (2020), 391-400. |
|  |  | [18] | X.H. Fu, X.B. He, D.F. Sun, Q.H. Fan, B.L. Zhang, Q. Wang, Y.F. Wang, and H.L. Chen, Histological chorioamnionitis could be predicted with high accuracy in preterm labor with intact membranes before delivery, Clin Exp Obstet Gynecol 45 (2018), 720-724. |
|  |  | [19] | K. Fuwa, M. Seki, Y. Hirata, I. Yanagihara, Y. Nakura, C. Takano, K. Kuroda, and S. Hayakawa, Rapid and simple detection of Ureaplasma species from vaginal swab samples using a loop-mediated isothermal amplification method, Am J Reprod Immunol 79 (2018). |
|  |  | [20] | A. Gámez-Varela, M. Martínez-Rodríguez, H. López-Briones, J. Luna-García, E. Chávez-González, R. Villalobos-Gómez, E. Hernandez-Andrade, and R. Cruz-Martínez, Preoperative Cervical Length Predicts the Risk of Delivery within One Week after Pleuroamniotic Shunt in Fetuses with Severe Hydrothorax, Fetal Diagn Ther (2021), 1-7. |
|  |  | [21] | L.L. Gievers, J. Sedler, C.A. Phillipi, D. Dukhovny, J. Geddes, P. Graven, B. Chan, and S. Khaki, Implementation of the sepsis risk score for chorioamnionitis-exposed newborns, J Perinatol 38 (2018), 1581-1587. |
|  |  | [22] | M. Gioan, F. Fenollar, A. Loundou, J.P. Menard, J. Blanc, C. D'Ercole, and F. Bretelle, Development of a nomogram for individual preterm birth risk evaluation, J Gynecol Obstet Hum Reprod 47 (2018), 545-548. |
|  |  | [23] | P. Girón de Velasco-Sada, I. Falces-Romero, I. Quiles-Melero, A. García-Perea, and J. Mingorance, Evaluation of two real time PCR assays for the detection of bacterial DNA in amniotic fluid, J Microbiol Methods 144 (2018), 107-110. |
|  |  | [24] | A. Hesson and E. Langen, Outcomes in oligohydramnios: the role of etiology in predicting pulmonary morbidity/mortality, J Perinat Med 46 (2018), 948-950. |
|  |  | [25] | Y. Horikoshi, C. Yaguchi, N. Furuta-Isomura, T. Itoh, K. Kawai, T. Oda, M. Matsumoto, Y. Kohmura-Kobayashi, N. Tamura, T. Uchida, N. Kanayama, and H. Itoh, Gross appearance of the fetal membrane on the placental surface is associated with histological chorioamnionitis and neonatal respiratory disorders, PLoS One 15 (2020), e0242579. |
|  |  | [26] | K. Hughes, S. Sim, A. Roman, K. Michalak, S. Kane, and P. Sheehan, Outcomes and predictive tests from a dedicated specialist clinic for women at high risk of preterm labour: A ten year audit, Aust N Z J Obstet Gynaecol 57 (2017), 405-411. |
|  |  | [27] | A. Husain, A. Naseem, S. Anjum, S. Imran, M. Arifuzzaman, and S.O. Adil, Predictability of intrapartum cardiotocography with meconium stained liquor and its correlation with perinatal outcome, J Pak Med Assoc 68 (2018), 1014-1018. |
|  |  | [28] | J.K. Iversen, B.H. Kahrs, E.A. Torkildsen, and T.M. Eggebo, Fetal molding examined with transperineal ultrasound and associations with position and delivery mode, Am J Obstet Gynecol 223 (2020). |
|  |  | [29] | M. Jain, A. Bang, A. Tiwari, and S. Jain, Prediction of significant hyperbilirubinemia in term neonates by early non-invasive bilirubin measurement, World J Pediatr 13 (2017), 222-227. |
|  |  | [30] | F. Jochum, C. Le Ray, P. Blanc-Petitjean, B. Langer, N. Meyer, F. Severac, and N. Sananes, Externally Validated Score to Predict Cesarean Delivery After Labor Induction With Cervi Ripening, Obstet Gynecol 134 (2019), 502-510. |
|  |  | [31] | S. Jwala, T.L. Tran, C. Terenna, A. McGregor, J. Andrel, B.E. Leiby, J.K. Baxter, and V. Berghella, Evaluation of additive effect of quantitative fetal fibronectin to cervical length for prediction of spontaneous preterm birth among asymptomatic low-risk women, Acta Obstet Gynecol Scand 95 (2016), 948-955. |
|  |  | [32] | M. Kaneko, M. Sato, K. Ogasawara, T. Imamura, K. Hashimoto, N. Momoi, and M. Hosoya, Serum cytokine concentrations, chorioamnionitis and the onset of bronchopulmonary dysplasia in premature infants, J Neonatal Perinatal Med 10 (2017), 147-155. |
|  |  | [33] | T. Kawakita, U.M. Reddy, J.C. Huang, T.C. Auguste, D. Bauer, and R.T. Overcash, Externally Validated Prediction Model of Vaginal Delivery After Preterm Induction With Unfavorable Cervix, Obstet Gynecol 136 (2020), 716-724. |
|  |  | [34] | A. Kesrouani, E. Chalhoub, E. El Rassy, M. Germanos, A. Khazzaka, J. Rizkallah, E. Attieh, and N. Aouad, Prediction of preterm delivery by second trimester inflammatory biomarkers in the amniotic fluid, Cytokine 85 (2016), 67-70. |
|  |  | [35] | Y.M. Kim, J.Y. Park, J.H. Sung, S.J. Choi, S.Y. Oh, C.R. Roh, and J.H. Kim, Predicting factors for success of vaginal delivery in preterm induction with prostaglandin E(2), Obstet Gynecol Sci 60 (2017), 163-169. |
|  |  | [36] | P. Kosinski and M. Wielgos, Foetoscopic endotracheal occlusion (FETO) for severe isolated left-sided congenital diaphragmatic hernia: single center Polish experience, Journal of Maternal-Fetal & Neonatal Medicine 31 (2018), 2521-2526. |
|  |  | [37] | N.B. Kunzier, W.L. Kinzler, M.R. Chavez, T.M. Adams, D.A. Brand, and A.M. Vintzileos, The use of cervical sonography to differentiate true from false labor in term patients presenting for labor check, Am J Obstet Gynecol 215 (2016), 372.e371-375. |
|  |  | [38] | M. Kurakazu, F. Yotsumoto, H. Arima, D. Izuchi, D. Urushiyama, K. Miyata, C. Kiyoshima, S. Fukagawa, K. Yoshikawa, M. Kurakazu, T. Hirakawa, K. Shigekawa, R. Araki, A. Sanui, M. Murata, K. Nabeshima, and S. Miyamoto, The combination of maternal blood and amniotic fluid biomarkers improves the predictive accuracy of histologic chorioamnionitis, Placenta 80 (2019), 4-7. |
|  |  | [39] | C.A. Labarrere, H.L. DiCarlo, E. Bammerlin, J.W. Hardin, Y.M. Kim, P. Chaemsaithong, D.M. Haas, G.S. Kassab, and R. Romero, Failure of physiologic transformation of spiral arteries, endothelial and trophoblast cell activation, and acute atherosis in the basal plate of the placenta, Am J Obstet Gynecol 216 (2017), 287.e281-287.e216. |
|  |  | [40] | Y. Liu, Y. Liu, C. Du, R. Zhang, Z. Feng, and J. Zhang, Diagnostic value of amniotic fluid inflammatory biomarkers for subclinical chorioamnionitis, Int J Gynaecol Obstet 134 (2016), 160-164. |
|  |  | [41] | G.C. Ma, Y.C. Chen, W.J. Wu, S.P. Chang, T.Y. Chang, W.H. Lin, and M. Chen, Prenatal Diagnosis of Autosomal Recessive Renal Tubular Dysgenesis with Anhydramnios Caused by a Mutation in the AGT Gene, Diagnostics 9 (2019). |
|  |  | [42] | H. Miremberg, N. Elia, C. Marelly, O. Gluck, G. Barda, J. Bar, and E. Weiner, Should the interval between doses of antenatal corticosteroids be shortened in certain cases? Factors predicting preterm delivery < 48 h from presentation, Arch Gynecol Obstet (2021). |
|  |  | [43] | L.J. Moulton, J. Eric Jelovsek, M. Lachiewicz, K. Chagin, and O. Goje, A model to predict risk of postpartum infection after Caesarean delivery, J Matern Fetal Neonatal Med 31 (2018), 2409-2417. |
|  |  | [44] | G. Natarajan, S. Shankaran, S. Saha, A. Laptook, A. Das, R. Higgins, B.J. Stoll, E.F. Bell, W.A. Carlo, C. D'Angio, S.B. DeMauro, P. Sanchez, K. Van Meurs, B. Vohr, N. Newman, E. Hale, and M. Walsh, Antecedents and Outcomes of Abnormal Cranial Imaging in Moderately Preterm Infants, J Pediatr 195 (2018), 66-72.e63. |
|  |  | [45] | S. Ona, S.R. Easter, M. Prabhu, G. Wilkie, R.E. Tuomala, L.E. Riley, and K. Diouf, Diagnostic Validity of the Proposed Eunice Kennedy Shriver National Institute of Child Health and Human Development Criteria for Intrauterine Inflammation or Infection, Obstet Gynecol 133 (2019), 33-39. |
|  |  | [46] | P. Pacora, R. Romero, O. Erez, E. Maymon, B. Panaitescu, J.P. Kusanovic, A.L. Tarca, C.D. Hsu, and S.S. Hassan, The diagnostic performance of the beta-glucan assay in the detection of intra-amniotic infection with Candida species, J Matern Fetal Neonatal Med 32 (2019), 1703-1720. |
|  |  | [47] | H. Park, K.H. Park, Y.M. Kim, S.Y. Kook, S.J. Jeon, and H.N. Yoo, Plasma inflammatory and immune proteins as predictors of intra-amniotic infection and spontaneous preterm delivery in women with preterm labor: a retrospective study, BMC Pregnancy Childbirth 18 (2018), 146. |
|  |  | [48] | J.W. Park, K.H. Park, S.Y. Kook, Y.M. Jung, and Y.M. Kim, Immune biomarkers in maternal plasma to identify histologic chorioamnionitis in women with preterm labor, Arch Gynecol Obstet 299 (2019), 725-732. |
|  |  | [49] | A.S. Patil, N.W. Gaikwad, C.A. Grotegut, S.D. Dowden, and D.M. Haas, Alterations in endogenous progesterone metabolism associated with spontaneous very preterm delivery, Hum Reprod Open 2020 (2020), hoaa007. |
|  |  | [50] | A. Pinton, F. Severac, N. Meyer, C.Y. Akladios, A. Gaudineau, R. Favre, B. Langer, and N. Sananes, A comparison of vaginal ultrasound and digital examination in predicting preterm delivery in women with threatened preterm labor: a cohort study, Acta Obstet Gynecol Scand 96 (2017), 447-453. |
|  |  | [51] | N. Pruksanusak, P. Thongphanang, T. Suntharasaj, C. Suwanrath, and A. Geater, Combined maternal-associated risk factors with intrapartum fetal heart rate classification systems to predict peripartum asphyxia neonates, Eur J Obstet Gynecol Reprod Biol 218 (2017), 85-91. |
|  |  | [52] | S. Rasmussen, C. Ebbing, and L.M. Irgens, Predicting preeclampsia from a history of preterm birth, PLoS One 12 (2017), e0181016. |
|  |  | [53] | M. Roethlisberger, B. Strizek, I. Gottschalk, M.R. Mallmann, A. Geipel, U. Gembruch, and C. Berg, First-trimester intervention in twin reversed arterial perfusion sequence: does size matter?, Ultrasound Obstet Gynecol 50 (2017), 40-44. |
|  |  | [54] | M. Sahni, M.E. Franco-Fuenmayor, and K. Shattuck, Management of Late Preterm and Term Neonates exposed to maternal Chorioamnionitis, BMC Pediatr 19 (2019), 282. |
|  |  | [55] | T. Samejima, T. Nagamatsu, D.J. Schust, N. Itaoka, T. Iriyama, T. Nakayama, A. Komatsu, K. Kawana, Y. Osuga, and T. Fujii, Elevated concentration of secretory leukocyte protease inhibitor in the cervical mucus before delivery, Am J Obstet Gynecol 214 (2016), 741.e741-747. |
|  |  | [56] | T. Samejima and K. Takechi, Elevated C-reactive protein levels in histological chorioamnionitis at term: impact of funisitis on term neonates, J Matern Fetal Neonatal Med 30 (2017), 1428-1433. |
|  |  | [57] | G.H. Son, Y. Kim, J.J. Lee, K.Y. Lee, H. Ham, J.E. Song, S.T. Park, and Y.H. Kim, MicroRNA-548 regulates high mobility group box 1 expression in patients with preterm birth and chorioamnionitis, Sci Rep 9 (2019), 19746. |
|  |  | [58] | J. Stranik, M. Kacerovsky, O. Soucek, M. Kolackova, I. Musilova, L. Pliskova, R. Bolehovska, P. Bostik, J. Matulova, B. Jacobsson, and C. Andrys, IgGFc-binding protein in pregnancies complicated by spontaneous preterm delivery: a retrospective cohort study, Sci Rep 11 (2021), 6107. |
|  |  | [59] | J.K. Straughen, D.P. Misra, L.M. Ernst, A.K. Charles, S. VanHorn, S. Ghosh, I. Buhimschi, C. Buhimschi, G. Divine, and C.M. Salafia, Methods to decrease variability in histological scoring in placentas from a cohort of preterm infants, BMJ Open 7 (2017), e013877. |
|  |  | [60] | J.H. Sung, S.J. Choi, S.Y. Oh, C.R. Roh, and J.H. Kim, Revisiting the diagnostic criteria of clinical chorioamnionitis in preterm birth, Bjog 124 (2017), 775-783. |
|  |  | [61] | S. Suzuki, Association between clinical chorioamnionitis and histological funisitis at term, J Neonatal Perinatal Med 12 (2019), 37-40. |
|  |  | [62] | K. Uno, M. Mayama, M. Yoshihara, T. Takeda, S. Tano, T. Suzuki, Y. Kishigami, and H. Oguchi, Reasons for previous Cesarean deliveries impact a woman's independent decision of delivery mode and the success of trial of labor after Cesarean, BMC Pregnancy Childbirth 20 (2020). |
|  |  | [63] | D. Urushiyama, W. Suda, E. Ohnishi, R. Araki, C. Kiyoshima, M. Kurakazu, A. Sanui, F. Yotsumoto, M. Murata, K. Nabeshima, S. Yasunaga, S. Saito, M. Nomiyama, M. Hattori, S. Miyamoto, and K. Hata, Microbiome profile of the amniotic fluid as a predictive biomarker of perinatal outcome, Sci Rep 7 (2017), 12171. |
|  |  | [64] | S. Usman, H. Barton, C. Wilhelm-Benartzi, and C.C. Lees, Ultrasound is better tolerated than vaginal examination in and before labour, Australian & New Zealand Journal of Obstetrics & Gynaecology 59 (2019), 362-366. |
|  |  | [65] | S. Usman, M. Wilkinson, H. Barton, and C.C. Lees, The feasibility and accuracy of ultrasound assessment in the labor room, Journal of Maternal-Fetal & Neonatal Medicine 32 (2019), 3442-3451. |
|  |  | [66] | L. van der Krogt, A.E. Ridout, P.T. Seed, and A.H. Shennan, Placental inflammation and its relationship to cervicovaginal fetal fibronectin in preterm birth, Eur J Obstet Gynecol Reprod Biol 214 (2017), 173-177. |
|  |  | [67] | E. Weiner, Y. Mizrachi, E. Grinstein, O. Feldstein, N. Rymer-Haskel, E. Juravel, L. Schreiber, J. Bar, and M. Kovo, The role of placental histopathological lesions in predicting recurrence of preeclampsia, Prenat Diagn 36 (2016), 953-960. |
|  |  | [68] | L. Zhao, Y. Lin, T. Jiang, L. Wang, M. Li, Y. Wang, G. Sun, and M. Xiao, Prediction of the induction to delivery time interval in vaginal dinoprostone-induced labor: a retrospective study in a Chinese tertiary maternity hospital, J Int Med Res 47 (2019), 2647-2654. |
| 3. g. Impact study | 2 | [1] | L. Goodfellow, A. Care, A. Sharp, J. Ivandic, B. Poljak, D. Roberts, and Z. Alfirevic, Effect of QUiPP prediction algorithm on treatment decisions in women with a previous preterm birth: a prospective cohort study, Bjog 126 (2019), 1569-1575. |
|  |  | [2] | J.B. Sauberan, B. Choi, A.R. Paradyse, and J. Le, Quality and Clinical Outcomes Associated with a Gentamicin Use System Change for Managing Chorioamnionitis, J Med Syst 41 (2017), 202. |
| 4. a. Retrieved | 30 | [1] | Good clinical practice advice: Prediction of preterm labor and preterm premature rupture of membranes, Int J Gynaecol Obstet 144 (2019), 340-346. |
|  |  | [2] | K. Abdel-Malek, M.A. El-Halwagi, B.E. Hammad, O. Azmy, O. Helal, M. Eid, and M. Abdel-Rasheed, Role of maternal serum ferritin in prediction of preterm labour, J Obstet Gynaecol 38 (2018), 222-225. |
|  |  | [3] | M.T. Aung, Y. Yu, K.K. Ferguson, D.E. Cantonwine, L. Zeng, T.F. McElrath, S. Pennathur, B. Mukherjee, and J.D. Meeker, Prediction and associations of preterm birth and its subtypes with eicosanoid enzymatic pathways and inflammatory markers, Sci Rep 9 (2019), 17049. |
|  |  | [4] | U. Baboolall, Y. Zha, X. Gong, D.R. Deng, F. Qiao, and H. Liu, Variations of plasma D-dimer level at various points of normal pregnancy and its trends in complicated pregnancies: A retrospective observational cohort study, Medicine (Baltimore) 98 (2019), e15903. |
|  |  | [5] | D. Başbuğ, A. Başbuğ, and C. Gülerman, Is unexplained elevated maternal serum alpha-fetoprotein still important predictor for adverse pregnancy outcome?, Ginekol Pol 88 (2017), 325-330. |
|  |  | [6] | M.A. Belfort, Predicting premature preterm rupture of the membranes after fetal surgery, Bjog 125 (2018), 1293. |
|  |  | [7] | M.T. Canda, N. Demir, and O. Sezer, Fetal Nasal Bone Length as a Novel Marker for Prediction of Adverse Perinatal Outcomes in the First-Trimester of Pregnancy, Balkan Med J 34 (2017), 127-131. |
|  |  | [8] | H.H. Cha, J.M. Kim, H.M. Kim, M.J. Kim, G.O. Chong, and W.J. Seong, Association between gestational age at delivery and lymphocyte-monocyte ratio in the routine second trimester complete blood cell count, Yeungnam Univ J Med 38 (2021), 34-38. |
|  |  | [9] | V. El-Achi, B. de Vries, C. O'Brien, F. Park, J. Tooher, and J. Hyett, First-Trimester Prediction of Preterm Prelabour Rupture of Membranes, Fetal Diagn Ther 47 (2020), 624-629. |
|  |  | [10] | V. El-Achi, F. Park, C. O'Brien, J. Tooher, and J. Hyett, Does low dose aspirin prescribed for risk of early onset preeclampsia reduce the prevalence of preterm prelabor rupture of membranes?, J Matern Fetal Neonatal Med 34 (2021), 618-623. |
|  |  | [11] | S. Gupta, H.S. Gaikwad, B. Nath, and A. Batra, Can vitamin C and interleukin 6 levels predict preterm premature rupture of membranes: evaluating possibilities in North Indian population, Obstet Gynecol Sci 63 (2020), 432-439. |
|  |  | [12] | H. Kansu-Celik, Y. Tasci, B.K. Karakaya, M. Cinar, T. Candar, and G.S. Caglar, Maternal serum advanced glycation end products level as an early marker for predicting preterm labor/PPROM: a prospective preliminary study, J Matern Fetal Neonatal Med 32 (2019), 2758-2762. |
|  |  | [13] | S.Y. Kook, K.H. Park, J.A. Jang, Y.M. Kim, H. Park, and S.J. Jeon, Vitamin D-binding protein in cervicovaginal fluid as a non-invasive predictor of intra-amniotic infection and impending preterm delivery in women with preterm labor or preterm premature rupture of membranes, PLoS One 13 (2018), e0198842. |
|  |  | [14] | K. Kuhrt, N. Hezelgrave, C. Foster, P.T. Seed, and A.H. Shennan, Development and validation of a tool incorporating quantitative fetal fibronectin to predict spontaneous preterm birth in symptomatic women, Ultrasound in Obstetrics and Gynecology 47 (2016), 210-216. |
|  |  | [15] | K. Kuhrt, E. Smout, N. Hezelgrave, P.T. Seed, J. Carter, and A.H. Shennan, Development and validation of a tool incorporating cervical length and quantitative fetal fibronectin to predict spontaneous preterm birth in asymptomatic high-risk women, Ultrasound in Obstetrics and Gynecology 47 (2016), 104-109. |
|  |  | [16] | H. Liang, Z. Xie, B. Liu, X. Song, and G. Zhao, A routine urine test has partial predictive value in premature rupture of the membranes, J Int Med Res 47 (2019), 2361-2370. |
|  |  | [17] | K. Lyubomirskaya, Y. Krut, L. Sergeyeva, S. Khmil, O. Lototska, N. Petrenko, and A. Kamyshnyi, Preterm premature rupture of membranes: prediction of risks in women of Zaporizhzhia region of Ukraine, Pol Merkur Lekarski 48 (2020), 399-405. |
|  |  | [18] | P. Mach, A. Köninger, L. Wicherek, R. Kimmig, S. Kasimir-Bauer, C. Birdir, B. Schmidt, and A. Gellhaus, Serum concentrations of soluble B7-H4 in early pregnancy are elevated in women with preterm premature rupture of fetal membranes, Am J Reprod Immunol 76 (2016), 149-154. |
|  |  | [19] | A. Ozel, E. Alici Davutoglu, A. Yurtkal, and R. Madazli, How do platelet-to-lymphocyte ratio and neutrophil-to-lymphocyte ratio change in women with preterm premature rupture of membranes, and threaten preterm labour?, J Obstet Gynaecol 40 (2020), 195-199. |
|  |  | [20] | S. Park, Y.A. You, H. Yun, S.J. Choi, H.S. Hwang, S.K. Choi, S.M. Lee, and Y.J. Kim, Cervicovaginal fluid cytokines as predictive markers of preterm birth in symptomatic women, Obstet Gynecol Sci 63 (2020), 455-463. |
|  |  | [21] | M.S. Payne, J.P. Newnham, D.A. Doherty, L.L. Furfaro, N.L. Pendal, D.E. Loh, and J.A. Keelan, A specific bacterial DNA signature in the vagina of Australian women in midpregnancy predicts high risk of spontaneous preterm birth (the Predict1000 study), Am J Obstet Gynecol 224 (2021), 206.e201-206.e223. |
|  |  | [22] | G. Raba, M. Kacerovsky, and P. Laudański, Eotaxin-2 as a potential marker of preterm premature rupture of membranes: A prospective, cohort, multicenter study, Adv Clin Exp Med 30 (2021), 197-202. |
|  |  | [23] | J. Takeda, X. Fang, and D.M. Olson, Pregnant human peripheral leukocyte migration during several late pregnancy clinical conditions: a cross-sectional observational study, BMC Pregnancy Childbirth 17 (2017), 16. |
|  |  | [24] | P.J. Teoh, A. Ridout, P. Seed, R.M. Tribe, and A.H. Shennan, Gender and preterm birth: Is male fetal gender a clinically important risk factor for preterm birth in high-risk women?, Eur J Obstet Gynecol Reprod Biol 225 (2018), 155-159. |
|  |  | [25] | L.A. Underhill, N. Avalos, R. Tucker, Z. Zhang, G. Messerlian, and B. Lechner, Serum Decorin and Biglycan as Potential Biomarkers to Predict PPROM in Early Gestation, Reprod Sci (2019), 1933719119831790. |
|  |  | [26] | B.I. Vandermolen, N.L. Hezelgrave, E.M. Smout, D.S. Abbott, P.T. Seed, and A.H. Shennan, Quantitative fetal fibronectin and cervical length to predict preterm birth in asymptomatic women with previous cervical surgery, Am J Obstet Gynecol 215 (2016), 480.e481-480.e410. |
|  |  | [27] | S.W. Verbruggen, M.L. Oyen, A.T. Phillips, and N.C. Nowlan, Function and failure of the fetal membrane: Modelling the mechanics of the chorion and amnion, PLoS One 12 (2017), e0171588. |
|  |  | [28] | F. Zhan, S. Zhu, H. Liu, Q. Wang, and G. Zhao, Blood routine test is a good indicator for predicting premature rupture of membranes, J Clin Lab Anal 33 (2019), e22673. |
|  |  | [29] | L.X. Zhang, Y. Sun, H. Zhao, N. Zhu, X.D. Sun, X. Jin, A.M. Zou, Y. Mi, and J.R. Xu, A Bayesian Stepwise Discriminant Model for Predicting Risk Factors of Preterm Premature Rupture of Membranes: A Case-control Study, Chin Med J (Engl) 130 (2017), 2416-2422. |
|  |  | [30] | J. Zhu, C. Ma, L. Zhu, J. Li, F. Peng, L. Huang, and X. Luan, A role for the NLRC4 inflammasome in premature rupture of membrane, PLoS One 15 (2020), e0237847. |
| 4. b. Duplicated but different year | 1 | [1] | L.A. Underhill, N. Avalos, R. Tucker, Z. Zhang, G. Messerlian, and B. Lechner, Serum Decorin and Biglycan as Potential Biomarkers to Predict PPROM in Early Gestation, Reprod Sci 27 (2020), 1620-1626. |
| 5. a. Eligible by full-text | 2 | [1] | D. Başbuğ, A. Başbuğ, and C. Gülerman, Is unexplained elevated maternal serum alpha-fetoprotein still important predictor for adverse pregnancy outcome?, Ginekol Pol 88 (2017), 325-330. |
|  |  | [2] | V. El-Achi, B. de Vries, C. O'Brien, F. Park, J. Tooher, and J. Hyett, First-Trimester Prediction of Preterm Prelabour Rupture of Membranes, Fetal Diagn Ther 47 (2020), 624-629. |
| 5. b. EPV <20 | 2 | [1] | P. Mach, A. Köninger, L. Wicherek, R. Kimmig, S. Kasimir-Bauer, C. Birdir, B. Schmidt, and A. Gellhaus, Serum concentrations of soluble B7-H4 in early pregnancy are elevated in women with preterm premature rupture of fetal membranes, Am J Reprod Immunol 76 (2016), 149-154. |
|  |  | [2] | L.A. Underhill, N. Avalos, R. Tucker, Z. Zhang, G. Messerlian, and B. Lechner, Serum Decorin and Biglycan as Potential Biomarkers to Predict PPROM in Early Gestation, Reprod Sci 27 (2020), 1620-1626. |
| 5. c. PROM as population | 3 | [1] | S.Y. Kook, K.H. Park, J.A. Jang, Y.M. Kim, H. Park, and S.J. Jeon, Vitamin D-binding protein in cervicovaginal fluid as a non-invasive predictor of intra-amniotic infection and impending preterm delivery in women with preterm labor or preterm premature rupture of membranes, PLoS One 13 (2018), e0198842. |
|  |  | [2] | H. Liang, Z. Xie, B. Liu, X. Song, and G. Zhao, A routine urine test has partial predictive value in premature rupture of the membranes, J Int Med Res 47 (2019), 2361-2370. |
|  |  | [3] | K. Lyubomirskaya, Y. Krut, L. Sergeyeva, S. Khmil, O. Lototska, N. Petrenko, and A. Kamyshnyi, Preterm premature rupture of membranes: prediction of risks in women of Zaporizhzhia region of Ukraine, Pol Merkur Lekarski 48 (2020), 399-405. |
| 5. d. Association study | 6 | [1] | M.T. Canda, N. Demir, and O. Sezer, Fetal Nasal Bone Length as a Novel Marker for Prediction of Adverse Perinatal Outcomes in the First-Trimester of Pregnancy, Balkan Med J 34 (2017), 127-131. |
|  |  | [2] | H.H. Cha, J.M. Kim, H.M. Kim, M.J. Kim, G.O. Chong, and W.J. Seong, Association between gestational age at delivery and lymphocyte-monocyte ratio in the routine second trimester complete blood cell count, Yeungnam Univ J Med 38 (2021), 34-38. |
|  |  | [3] | V. El-Achi, F. Park, C. O'Brien, J. Tooher, and J. Hyett, Does low dose aspirin prescribed for risk of early onset preeclampsia reduce the prevalence of preterm prelabor rupture of membranes?, J Matern Fetal Neonatal Med 34 (2021), 618-623. |
|  |  | [4] | P.J. Teoh, A. Ridout, P. Seed, R.M. Tribe, and A.H. Shennan, Gender and preterm birth: Is male fetal gender a clinically important risk factor for preterm birth in high-risk women?, Eur J Obstet Gynecol Reprod Biol 225 (2018), 155-159. |
|  |  | [5] | S.W. Verbruggen, M.L. Oyen, A.T. Phillips, and N.C. Nowlan, Function and failure of the fetal membrane: Modelling the mechanics of the chorion and amnion, PLoS One 12 (2017), e0171588. |
|  |  | [6] | J. Zhu, C. Ma, L. Zhu, J. Li, F. Peng, L. Huang, and X. Luan, A role for the NLRC4 inflammasome in premature rupture of membrane, PLoS One 15 (2020), e0237847. |
| 5. e. Diagnostic prediction study | 8 | [1] | M.T. Aung, Y. Yu, K.K. Ferguson, D.E. Cantonwine, L. Zeng, T.F. McElrath, S. Pennathur, B. Mukherjee, and J.D. Meeker, Prediction and associations of preterm birth and its subtypes with eicosanoid enzymatic pathways and inflammatory markers, Sci Rep 9 (2019), 17049. |
|  |  | [2] | S. Gupta, H.S. Gaikwad, B. Nath, and A. Batra, Can vitamin C and interleukin 6 levels predict preterm premature rupture of membranes: evaluating possibilities in North Indian population, Obstet Gynecol Sci 63 (2020), 432-439. |
|  |  | [3] | A. Ozel, E. Alici Davutoglu, A. Yurtkal, and R. Madazli, How do platelet-to-lymphocyte ratio and neutrophil-to-lymphocyte ratio change in women with preterm premature rupture of membranes, and threaten preterm labour?, J Obstet Gynaecol 40 (2020), 195-199. |
|  |  | [4] | S. Park, Y.A. You, H. Yun, S.J. Choi, H.S. Hwang, S.K. Choi, S.M. Lee, and Y.J. Kim, Cervicovaginal fluid cytokines as predictive markers of preterm birth in symptomatic women, Obstet Gynecol Sci 63 (2020), 455-463. |
|  |  | [5] | G. Raba, M. Kacerovsky, and P. Laudański, Eotaxin-2 as a potential marker of preterm premature rupture of membranes: A prospective, cohort, multicenter study, Adv Clin Exp Med 30 (2021), 197-202. |
|  |  | [6] | J. Takeda, X. Fang, and D.M. Olson, Pregnant human peripheral leukocyte migration during several late pregnancy clinical conditions: a cross-sectional observational study, BMC Pregnancy Childbirth 17 (2017), 16. |
|  |  | [7] | F. Zhan, S. Zhu, H. Liu, Q. Wang, and G. Zhao, Blood routine test is a good indicator for predicting premature rupture of membranes, J Clin Lab Anal 33 (2019), e22673. |
|  |  | [8] | L.X. Zhang, Y. Sun, H. Zhao, N. Zhu, X.D. Sun, X. Jin, A.M. Zou, Y. Mi, and J.R. Xu, A Bayesian Stepwise Discriminant Model for Predicting Risk Factors of Preterm Premature Rupture of Membranes: A Case-control Study, Chin Med J (Engl) 130 (2017), 2416-2422. |
| 5. f. Special population | 3 | [1] | K. Kuhrt, N. Hezelgrave, C. Foster, P.T. Seed, and A.H. Shennan, Development and validation of a tool incorporating quantitative fetal fibronectin to predict spontaneous preterm birth in symptomatic women, Ultrasound in Obstetrics and Gynecology 47 (2016), 210-216. |
|  |  | [2] | K. Kuhrt, E. Smout, N. Hezelgrave, P.T. Seed, J. Carter, and A.H. Shennan, Development and validation of a tool incorporating cervical length and quantitative fetal fibronectin to predict spontaneous preterm birth in asymptomatic high-risk women, Ultrasound in Obstetrics and Gynecology 47 (2016), 104-109. |
|  |  | [3] | B.I. Vandermolen, N.L. Hezelgrave, E.M. Smout, D.S. Abbott, P.T. Seed, and A.H. Shennan, Quantitative fetal fibronectin and cervical length to predict preterm birth in asymptomatic women with previous cervical surgery, Am J Obstet Gynecol 215 (2016), 480.e481-480.e410. |
| 5. g. Not PROM | 4 | [1] | K. Abdel-Malek, M.A. El-Halwagi, B.E. Hammad, O. Azmy, O. Helal, M. Eid, and M. Abdel-Rasheed, Role of maternal serum ferritin in prediction of preterm labour, J Obstet Gynaecol 38 (2018), 222-225. |
|  |  | [2] | U. Baboolall, Y. Zha, X. Gong, D.R. Deng, F. Qiao, and H. Liu, Variations of plasma D-dimer level at various points of normal pregnancy and its trends in complicated pregnancies: A retrospective observational cohort study, Medicine (Baltimore) 98 (2019), e15903. |
|  |  | [3] | H. Kansu-Celik, Y. Tasci, B.K. Karakaya, M. Cinar, T. Candar, and G.S. Caglar, Maternal serum advanced glycation end products level as an early marker for predicting preterm labor/PPROM: a prospective preliminary study, J Matern Fetal Neonatal Med 32 (2019), 2758-2762. |
|  |  | [4] | M.S. Payne, J.P. Newnham, D.A. Doherty, L.L. Furfaro, N.L. Pendal, D.E. Loh, and J.A. Keelan, A specific bacterial DNA signature in the vagina of Australian women in midpregnancy predicts high risk of spontaneous preterm birth (the Predict1000 study), Am J Obstet Gynecol 224 (2021), 206.e201-206.e223. |
| 5. h. Not original article | 2 | [1] | Good clinical practice advice: Prediction of preterm labor and preterm premature rupture of membranes, Int J Gynaecol Obstet 144 (2019), 340-346. |
|  |  | [2] | M.A. Belfort, Predicting premature preterm rupture of the membranes after fetal surgery, Bjog 125 (2018), 1293. |

eTable 13. Association tests

| Latent candidate predictors | Outcome regression | Inverse probability weighting (IPW) |
| --- | --- | --- |
| Multiple pregnancy | 1.325 (95% CI 1.118 to 1.569)* | 1.062 (95% CI 1.055 to 1.068)* |
| Chorioamnionitis | 5.77 (95% CI 3.823 to 8.707)* | 1.351 (95% CI 1.33 to 1.372)* |
| IAI | 2.134 (95% CI 1 to 4.555) | 1.118 (95% CI 1.083 to 1.153)* |
| APH | 0.413 (95% CI 0.287 to 0.594)* | 0.929 (95% CI 0.924 to 0.933)* |
| GTI | 2.138 (95% CI 1.368 to 3.342)* | 1.116 (95% CI 1.101 to 1.132)* |
| Periodontal disease | 0.383 (95% CI 0.26 to 0.566)* | 0.967 (95% CI 0.96 to 0.973)* |
| Polyhydramnios | 1.238 (95% CI 0.851 to 1.801) | 0.998 (95% CI 0.989 to 1.006) |
| Pneumonia | 0.91 (95% CI 0.538 to 1.539) | 1.037 (95% CI 1.025 to 1.049)* |
| Asthma | 0.649 (95% CI 0.514 to 0.82)* | 0.971 (95% CI 0.966 to 0.977)* |
| Low SES | 0.837 (95% CI 0.808 to 0.867)* | 0.979 (95% CI 0.978 to 0.98)* |
| Maternal age | 0.761 (95% CI 0.73 to 0.793)* | 0.969 (95% CI 0.969 to 0.97)* |
| Influenza | 0.957 (95% CI 0.863 to 1.061) | 0.995 (95% CI 0.993 to 0.997)* |
| * Statistically significant by 95% CI |  |  |
